# The COVID-19 vaccination campaign in Switzerland and its impact on disease spread

**DOI:** 10.1101/2023.04.06.23288251

**Authors:** M. Bekker-Nielsen Dunbar, L. Held

## Abstract

We analyse infectious disease case surveillance data stratified by region and age group to estimate COVID-19 spread and gain an understanding of the impact of introducing vaccines to counter the disease in Switzerland. The data used in this work is extensive and detailed and includes information on weekly number of cases and vaccination rates by age and region. Our approach takes into account waning immunity. The statistical analysis allows us to determine the effects of choosing alternative vaccination strategies. Our results indicate greater uptake of vaccine would have led to fewer cases with a particularly large effect on undervaccinated regions while an alternative distribution scheme ignoring age would affect the vulnerable population at the time (the elderly) and is less ideal.

## 1 Introduction

Each year vaccines (pre-exposure pharmaceutical prophylaxis interventions) save lives by reducing preventable illness and death. Though vaccines are tested vigorously to ensure they are efficacious and monitored extensively to ensure they are effective and safe, they are underutilised in routine immunisation. This has lead to outbreaks of vaccine-preventable diseases that were previously eliminated in Europe, such as measles [1]. There is a concern that newly developed vaccines in response to pandemics will not be accepted by the population they seek to protect.

In December 2020, vaccines to prevent severe acute respiratory syndrome coronavirus 2 (SARS-CoV-2, the causative agent of COVID-19) infections were authorised for use in Switzerland. They were distributed in 2021 following an age-based distribution scheme going from oldest to youngest. A booster (additional dose of vaccine after immunity is achieved which is used to maintain immunity) was included in the immunisation schedule for COVID-19 late in 2021 and continued during 2022 following the same age-based distribution scheme. As those aged 65 years and older were first invited to be immunised, later followed by 16 to 64 year olds, and finally 12 to 15 year olds, changes in the age profile of cases (who is getting sick) have been observed. Additionally, in Switzerland, changes in the spatial distribution of cases (where people are getting sick) have also been observed, with a shift from urban centres to more rural, unvaccinated communities.

Vaccine hesitancy (defined as a delay in acceptance or refusal of vaccines despite availability) is found in Switzerland, where vaccination is not mandatory. This is a current issue of mounting concern that needs to be solved. The World Health Organization named vaccine hesitancy as one of the ten threats to global health already in 2019, noting its detrimental effects on populations who are less protected against life-threatening diseases even in non-pandemic settings. Vaccine hesitancy affects the uptake of vaccines and likely also the adherence to other public health interventions in certain population groups. An undervaccinated population has the potential to hamper disease control efforts for the entire country. Others have noted that in the Swiss context “vaccine hesitancy and underimmunisation seem to be specific to certain population subgroups” [2]. COVID-19 is currently the only human coronavirus for which there is a vaccine available but hesitancy persists [3]. Ongoing work seeks to understand the drivers of Swiss vaccine hesitancy [4].

We study COVID-19 incidence data from 2021 (from 1st January 2021 to 30th November 2021, both dates included) to examine the effect of vaccines on the spread of disease in Switzerland. We analyse weekly cases in two separate analyses, one is stratified by age group and one is stratified by region. This approach is motivated by cases being observed among younger ages in this time period (compared with earlier), with cases also exhibiting spatial heterogeneity in regions, as well as vaccination rates differing by unit (age group or region). Switzerland provides a unique opportunity to examine the effects of vaccination heterogeneity due to its federalised nature. Regions are less harmonised than in other European countries. We seek to determine the impact of the current vaccination strategy and the effect of regions with low vaccine uptake.

Using epidemic data sources from infectious disease surveillance systems at weekly resolution, we are able to capture the spread of disease across space and between age groups through an endemic-epidemic modelling approach. The vaccination coverage is time-dependent, age group-dependent, and region-dependent. Our approach builds upon a unique incorporation of time-varying vaccination coverage in an endemic-epidemic model with time-varying transmission weights. Time-varying transmission weights reflecting weekly levels of situational measures (amount of disease control) is a novel inclusion in the model since the advent of COVID-19, which has proven to be useful in examining policy questions [5–7]. Outlined here is a wider application which also includes vaccination effects. We invite interested readers to our study protocol [8] which contains even more detail.

Vaccination coverage has previously been included in endemic-epidemic modelling [9–12] for routine immunisation without taking into account changes over time (assuming stationarity). We include vaccination coverage in a similar manner in our models; to our knowledge the first endemic-epidemic model for COVID-19 which includes time and unit-specific vaccination rates. Calls for other vaccination distribution criteria than age to be considered when introducing novel vaccines to immunisation schedules in pandemic settings, such as socio-economic status [13], have been made. For this reason, we analyse alternative scenarios (an approach which has successfully been utilised in the analysis of non-pharmaceutical interventions to evaluate the impact of their timing [5–7]) to examine other rollout strategy schemes and uptake options (increased coverage).

## 2 Material

The data considered in this work includes temporal variables (week of reporting or recording/entry), biological variables (number of cases, age of case, dose-specific vaccination information), and spatial variables (region). Data is publicly available from the Swiss public health authority*’*s COVID-19-specific website (BAG) www.covid19.admin.ch and introduced in our study protocol [8]. Auxiliary data is provided by Mistry et al. [14] (contacts), Hale et al. [15] (policy), and Data for Good^1^ (mobility) and where merging of data is necessary, the groupings and temporal resolution used in the data from BAG are matched. The age bands provided are ten year age bands, however we have not included the age group 0 − 9 in this work as the focus is on the protective effect of vaccines and they received none during the study period leading to a vaccination coverage which is constantly zero, providing no information to the model.

### 2.1 Spatial dispersion

Spatial dispersion is included in the model to reflect how disease spreads. Switzerland consists of 26 regions (Figure 2). The adjacencies between the 26 Swiss regions are given in a matrix of neighbourhood order **o** = (*o*_*rr′*_) denoting the distance from one region *r* to another *r*^*′*^ (Figure 2). In this calculation, we decided that Neuchâtel (NE) and Jura (JU) as well as Schaffhausen (SH) and Thurgau (TG) do not share a border as when looking at a regular map they did not share most of their border in common.

To reflect additional changes in movement during the study period as a result of disease control policy implemented, we include mobility data *m*_*rt*_ (Figure 1) measuring the average change in mobility in region *r* in week *t*. The data is provided as a change from baseline (February 2020) which we normalise by transforming it 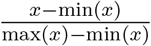 to ensure that we do not have negative transmission weights in our model (as the change from baseline is sometimes given as a negative value). Mobility data was not available for Appenzell Innerrhoden (AI) so we imputed them with values from the adjoining region Appenzell Ausserrhoden (AR).

**Figure 1:**
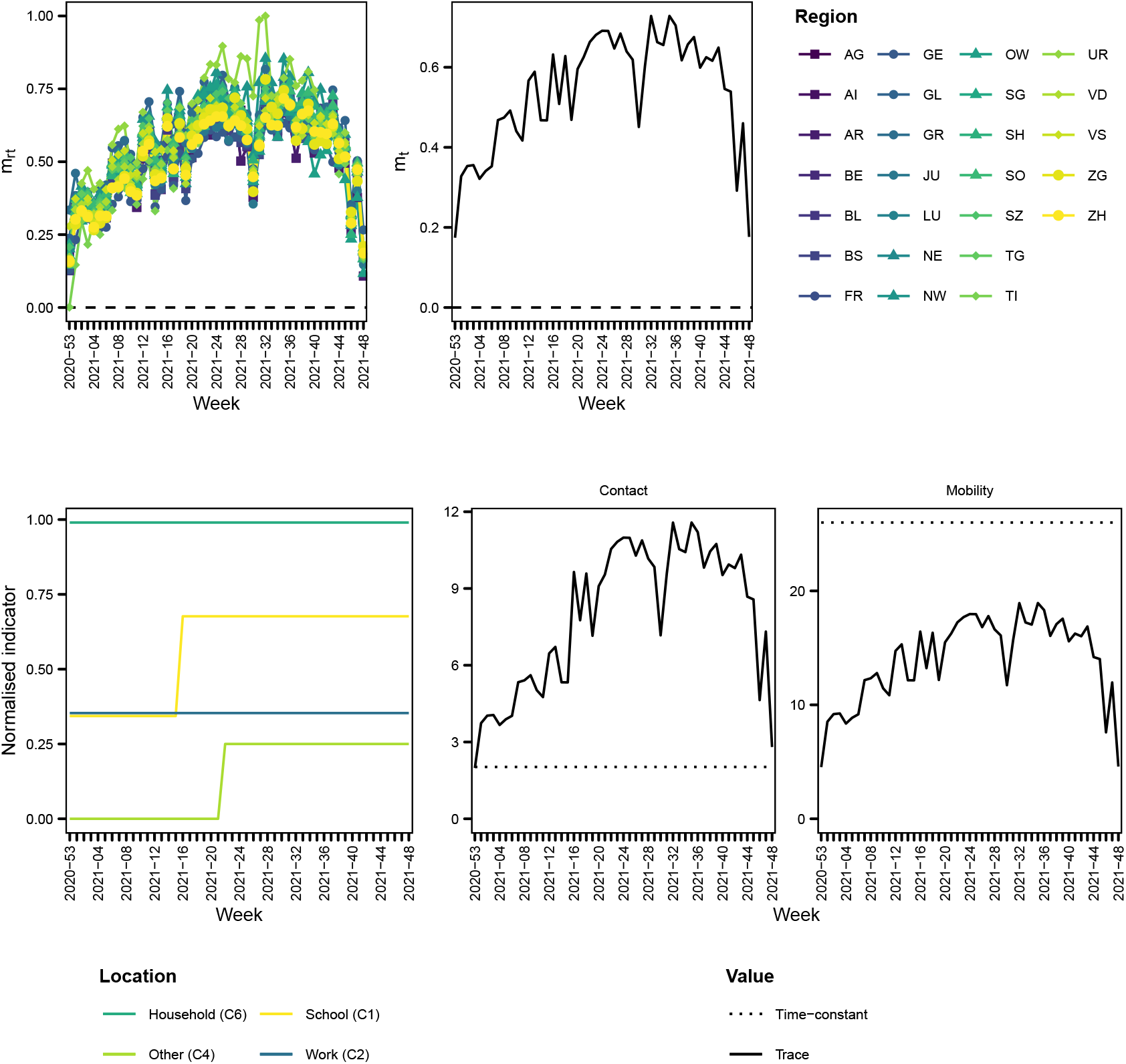
Mobility *m*_*rt*_ (above left) and *m*_*t*_ (above right), policy *q*_*t*_ (bottom left), and the trace (sum of the diagonal) of the contact matrix (bottom middle) and adjacency matrix (bottom right) at each time compared with the trace of the equivalent unadjusted hence time-constant transmission weights matrix (dashed lines). The abbreviations for regions are listed in the supporting information (file supp)

The adjacency matrix is adjusted by mobility data like in Grimée et al. [5] such that we obtain time-varying adjacency matrices **w** (short form for adjacency matrices at each time point *t*) with entries:

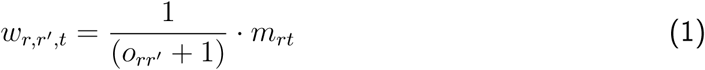

(Figure 3). We see an increase in mobility during the middle of the study period (Figure 1). This means we will expect to observe increased transmission opportunities in certain regions during that time.

**Figure 2:**
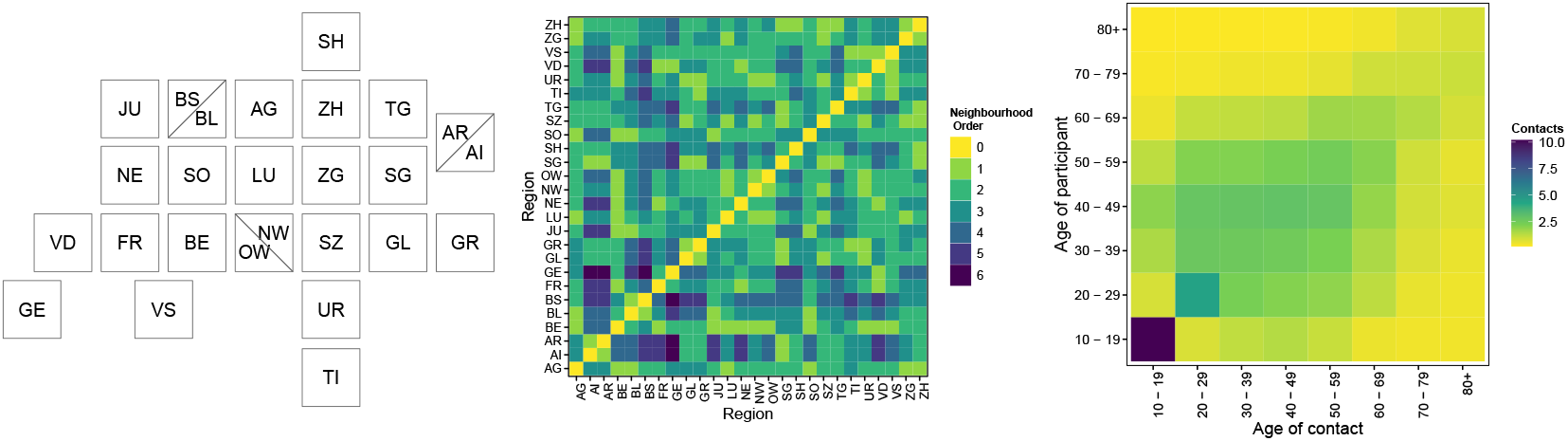
Swiss regions presented as map tiles (left; source: EBG [16]), adjacencies between the regions **o** = (*o*_*rr′*_) (middle), and contacts between age groups in Switzerland **c** = (*c*_*aa′*_) (right)

**Figure 3:**
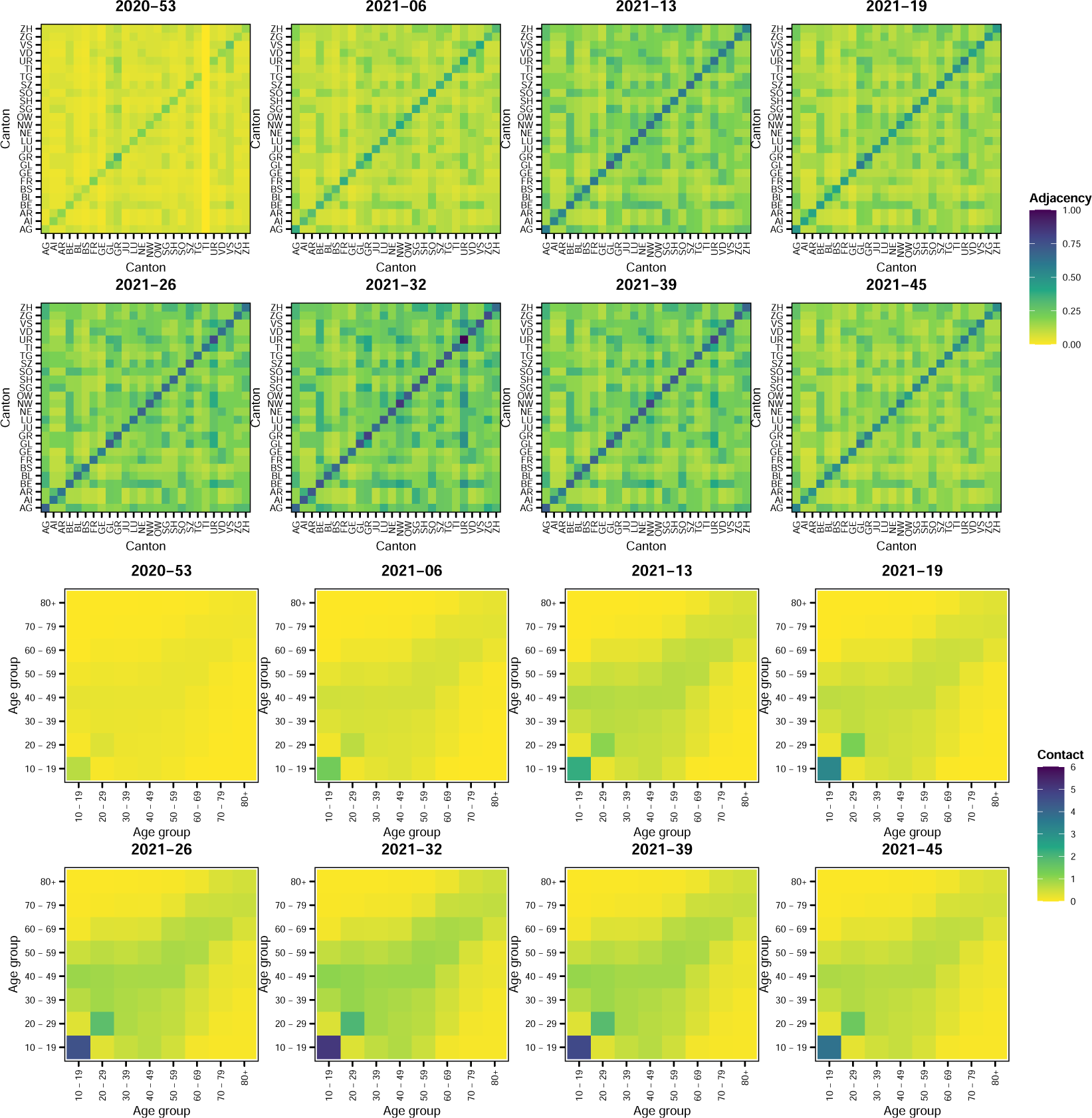
Snapshot of time-varying adjacency matrices **w** (above) and snapshot of time-varying contact matrices **w** (below), see the supporting information (file supp) for the full sets

### 2.2 Contacts

Respiratory diseases such as COVID-19 are transmitted through contact between age group *a* and *a*^*′*^. We incorporate a contact matrix **c** = (*c*_*aa′*_) in the model to reflect the pattern of spread of the disease across age groups. The Swiss contact matrices from Mistry et al. [14] (Figure 2) are used in this work and aggregated to the ten year age bands used in the case data. Contacts occur in four locations *l* (household, school, work, and other). The contact matrices are adjusted by the policy data like in Bekker-Nielsen Dunbar et al. [6, 7] such that we obtain time-varying contact matrices **w** for each week *t* across all locations *l* with entries given by:

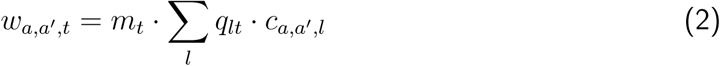

where the proportionality factor *q*_*lt*_ = *q*_*l*_ *· q*_*t*_ is informed by *q*_*l*_ which is a weight for contacts in location *l* (which is provided with the location-specific contact matrices) and *q*_*t*_ which denotes policy implemented in week *t* (Figure 3). Once we have calculated the total contacts using this factor, we additionally incorporate the average change in mobility across regions in week *t, m*_*t*_ (Figure 1) across all contacts because the policy indicators *q*_*t*_ do not capture more nuanced changes in transmission opportunities such as those caused by public holidays whereas the mobility data is expected to encapsulate such aspects. The mobility *m*_*rt*_ is averaged over regions to obtain *m*_*t*_. Earlier policy data was more granular. We divide the raw policy indicators by the maximum level it can take and then impose a lower bound of 0.001 to obtain *q*_*t*_ (the lower bound reflects that some contacts still occur when measures are in place). As in Bekker-Nielsen Dunbar et al. [6, 7] *q*_*t*_ *≡* 1 in the household setting.

### 2.3 Vaccines

The vaccination coverage is calculated based on the second (full immunity) and third doses (“booster”) of the vaccine as

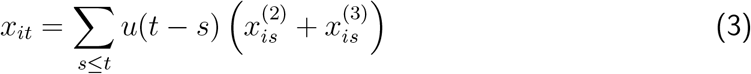

where 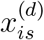 is the vaccination coverage of dose *d* (*d* = 2, 3) for unit *i* in week *s* (doses given at time *s* scaled by the population of unit *i*), week *s* occurs before week *t*, and *u*(.) is the waning function (Figure 4). We apply the waning function *u*(.) to account for waning immunity in our vaccination coverage calculation (see the supporting information (file sens) for a sensitivity analysis of *u*(.)). The unit *i* can either be age groups (*a*) or regions (*r*). This approach allows us to determine the vaccination coverage in each week *t* taking into account that the COVID-19 vaccines do not provide lifelong immunity. For each unit at each time point, the cumulative doses given are the sum of vaccines given in that week as well as the waned vaccines given in previous weeks *t*. The vaccination coverage is shown in Figure 5.

**Figure 4:**
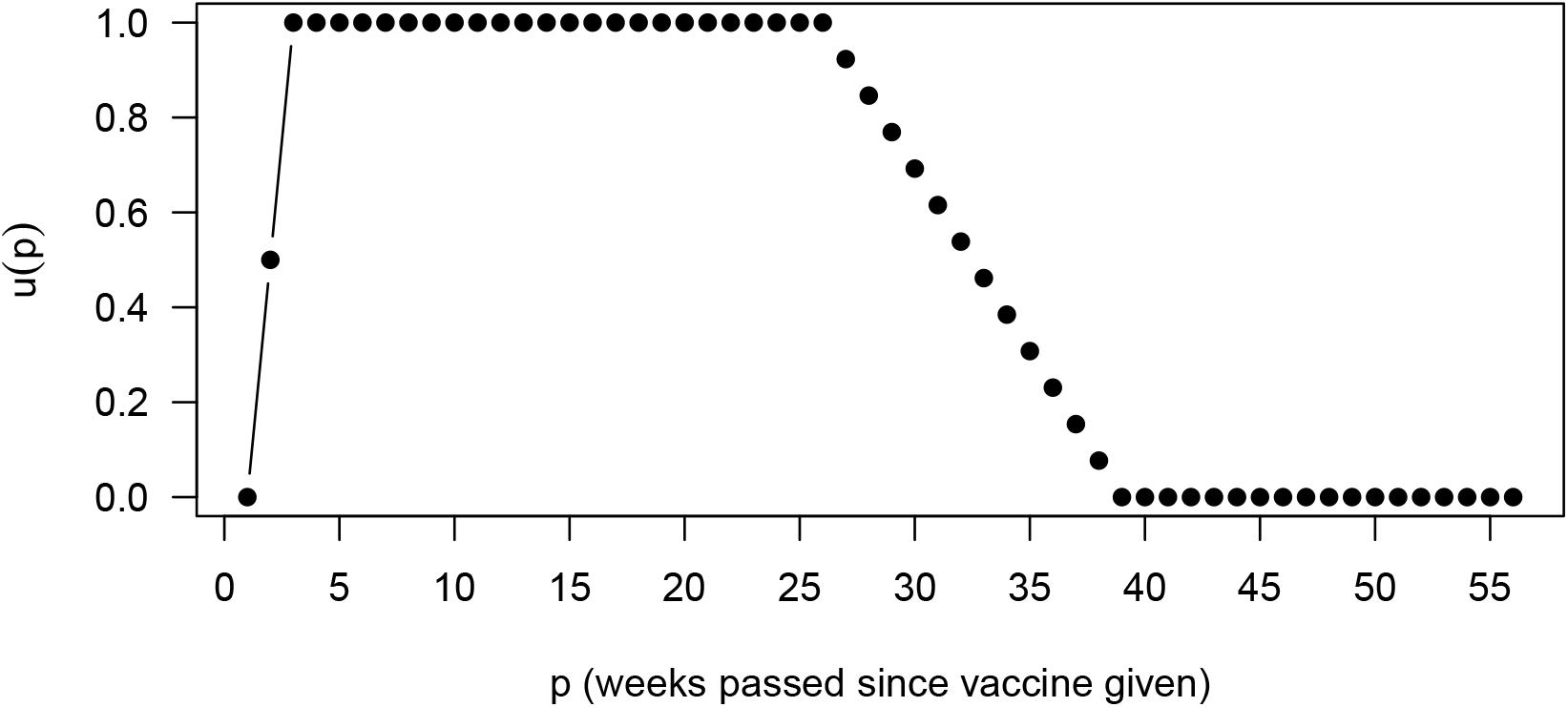
Waning function *u*(*p*) used in calculation of (3). Adapted from Bekker-Nielsen Dunbar and Held [8]

**Figure 5:**
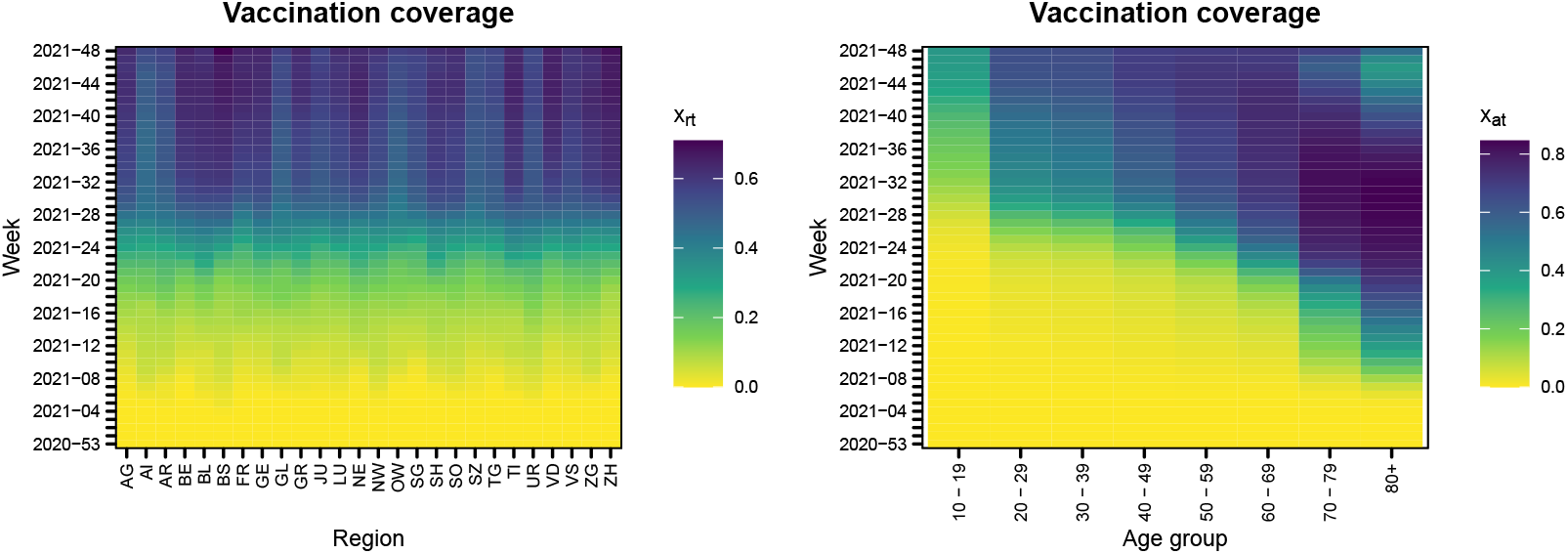
Calculated vaccination coverage taking into account waning immunity *x*_*it*_ by region (left) and age group (right). The time scale goes from bottom to top

## 3 Methods

We fit four types of endemic-epidemic models using the open source software package developed by Meyer et al. [17]. The endemic-epidemic model is a time series-based modelling approach for infectious disease surveillance first formulated in Held et al. [18]. Since its introduction it has been extended to include vaccination coverage [9], random intercepts [19], seasonal effects [20], time-constant transmission weights between units relaxing a homogenous mixing assumption [21, 22], prediction and forecasting [23], and most recently time-varying transmission weights [5–7].

We chose which effects to include in our model on the basis of scoping the literature for other endemic-epidemic models with vaccination coverage (Table 1). The inclusion of previous season*’*s incidence does not make sense for emerging infectious disease such as COVID-19 as the situation is ever-changing.

**Table 1:** Comparison with the literature (measles) and effects included in our COVID-19 models

| Effect | Measles |  |  |  | COVID-19 |  |
| --- | --- | --- | --- | --- | --- | --- |
|  | Herzog et al. [9] | Robert et al. [10] | Nguyen et al. [11] | Lu and Meyer [12] | Region | Age |
| Vaccination coverage | ✓<sup>1</sup> | ✓<sup>2</sup> | ✓<sup>1</sup> | ✓<sup>3</sup> | ✓ | ✓ |
| Sine-cosine wave | ✓ | ✓ | ✓ | ✓ | ✓ | ✓ |
| Population fraction offset in endemic component | ✓ | ✓<sup>4</sup> | ✓ | ✓ | ✓ | ✓ |
| Intercepts | ✓ | ✓ | ✓ | ✓ | ✓ | ✓ |
| Random effects of unit |  |  | ✓ |  | ✓ |  |
| Fixed effects of unit |  |  |  |  |  | ✓ |
| Centered time trend |  |  | ✓<sup>5</sup> |  | ✓ | ✓ |
| Previous season's incidence |  | ✓ |  |  | NA | NA |
| Geographical size (surface area) |  | ✓ |  |  |  |  |
| Gravity model in epidemic component |  | ✓ |  |  | ✓ |  |

1: constant, 2: averaged, 3: yearly, 4: is population count rather than fraction, 5: trend is not centered

### 3.1 Model

The endemic-epidemic model is given by

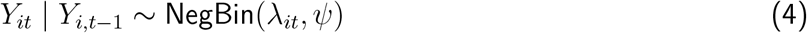

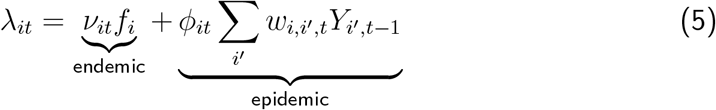

Cases *Y*_*it*_ observed in week *t* in unit *i* are conditionally negative binomially distributed with by mean *λ* and overdispersion *ψ* (4). The unit *i* can either be age groups (*a*) or regions (*r*). The mean of the negative binomial distribution *λ* is additively decomposed into an endemic component with log-linear predictor *ν* and an epidemic component with log-linear predictor *ϕ* (5). Previous cases *Y*_*·,t*−1_ in all units are weighted by the transmission weights *w*_*i,i′,t*_ and the population fraction *f*_*i*_ (given as the size of the population for unit *i* compared with the total population) enters as an offset in the endemic component.

We name our four types of endemic-epidemic models based on the components where the vaccination coverage is included. We consider the following four combinations of log-linear predictors *ν* and *ϕ*

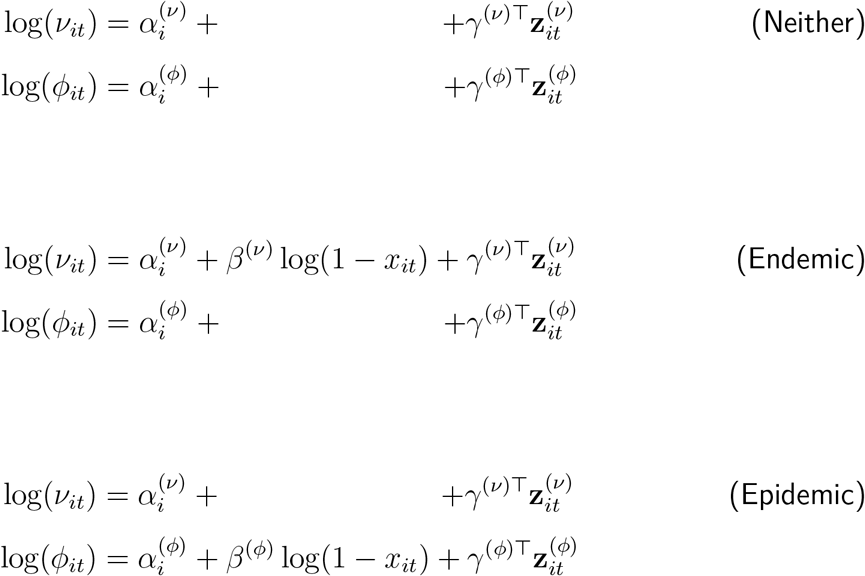

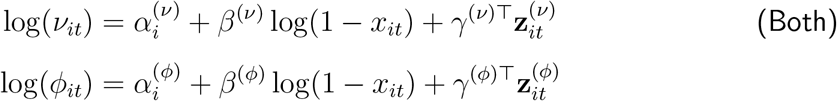

We use the log-proportion of the unvaccinated population as suggested by Herzog et al. [9] and used by other endemic-epidemic models in the literature (Table 1). Thus the log-transformed vaccination coverage log(1 − *x*_*it*_) enters the model as a coefficient in the log-linear predictors. Additional effects enter as either fixed or random effects [19] of unit *α* (intercepts) or additional covariates **z**. We consider

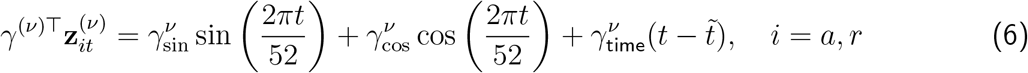

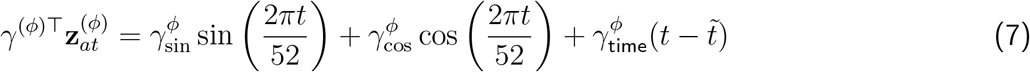

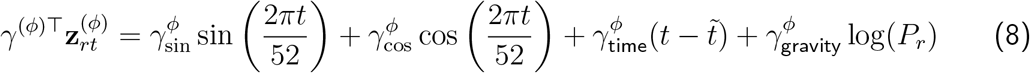

where we have incuded a sine-cosine wave to account for yearly (52 weeks) oscillation [20], 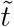 denotes the median of the study period (the time trend is centered to make it more stable and is intended to capture fluctuations not captured by the sine-cosine wave) and log(*P*_*r*_) is the log population count of region *r*. The latter is known as the gravity model [24] and reflects the fact that a greater attraction (higher mass) is to be expected from populous regions. We fit models to regional units *r* and age group units *a* separately.

While ours is the first endemic-epidemic model for COVID-19 with time-varying vaccination coverage at weekly granularity, previous models have been constructed for measles with vaccination coverage [9–12] (Table 1). While other research groups [9–12] considered a different disease (measles) and setting (non-pandemic), their estimated effects of vaccination coverage are included in Figure 8 as a comparison with our work. We estimate the effect of vaccination coverage for models with time constant transmission weights to allow for such that we can liken the results for novel vaccines with routing scheduled immunisation. We then focus on the results for models with time-varying transmission weights **w** = (*w*_*ii′t*_) as a constant transmission weight matrix **w** = (*w*_*ii′*_) is not informative for a situation with as many changes as expected in the setting considered; an emerging infectious disease with the introduction of a pharmaceutical countermeasure.

### 3.2 Scenario prediction

We predict the number of expected COVID-19 cases under the alternative scenarios of vaccination distribution hence coverage. Using the multivariate path forecasting method from Held et al. [23, Appendix A] we predict the mean (first moment) incidence by unit (age group or region). The single-step prediction approach is iteratively applied to obtain multivariate multi-step predictions. We use the estimated model coefficients in the prediction approach rather than refitting the model.

In one scenario we replace the observed vaccination coverage covariate (Figure 5) log(1 − *x*_*it*_) with an alternative option which for *x*_*rt*_ is the maximum vaccination coverage in any region *r* (Figure 6) and for *x*_*at*_ is the number of vaccines given to each age group *a* after redistributing the total vaccines given at any time to be given to all age groups by amounts proportional to the size of the age group (see the supporting information (file supp) for the redistribution). This scenario projection allows us to determine the expected impact of two alternative vaccination distribution schemes.

**Figure 6:**
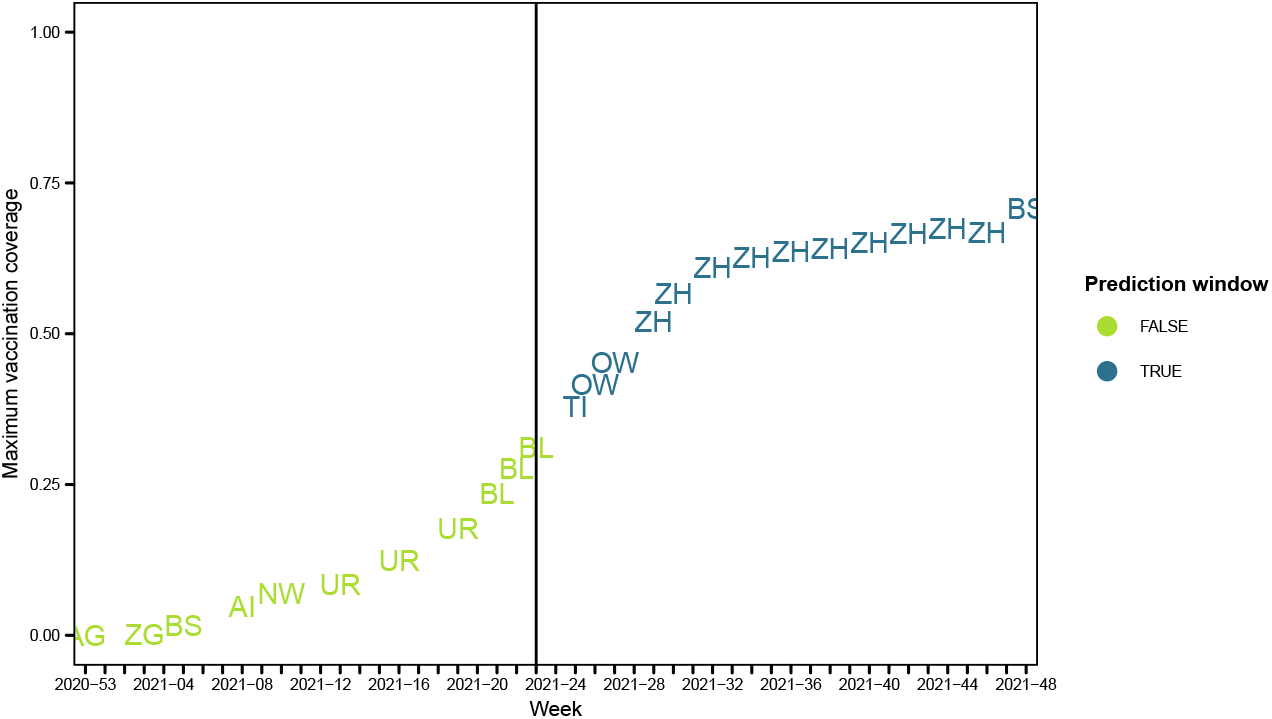
Overview of which region has the maximum vaccination coverage in a given week *t* provising the alternative uptake scenario we consider for regions. The vertical line denotes where our predictions begin

In line with recommendations by den Boon et al. [25] we present results with within-model uncertainty ranges. These are obtained by simulating the estimated coefficients (Table 4) from our fitted models assuming a multivariate normal distribution [like 5–7].

## 4 Results

We fit models to regional units *r* and age group units *a* separately. We provide here an overview of results and refer the interested reader to the supporting information (files models-age and models-regions) for full model results (there are results for in total 26 regions and eight age groups). The models contain the effects outlined in Table 1. Our estimated effect for vaccination coverage in a model with time constant transmission weights **w** = (*w*_*ii′*_) is in line with the results obtained by other research groups [9–12] (Table 2 and Figure 8). We now focus on the results for time-varying transmission weights **w** = (*w*_*ii′t*_).

**Table 2:** Vaccination coverage covariate estimates 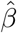 and standard errors in brackets

|  | Weight option | Neither | Endemic | Epidemic | Both |  |
| --- | --- | --- | --- | --- | --- | --- |
|  |  |  |  |  | Endemic | Epidemic |
| Region | Time-constant (1R) | – | 4.401 (1.343) | 3.251 (0.216) | 1.677 (1.038) | 3.206 (0.216) |
|  | Time-varying (2R) | – | 3.312 (1.035) | 2.688 (0.201) | 1.739 (0.846) | 2.642 (0.2) |
| Age | Time-constant (1A) | – | 2.042 (0.246) | 0.722 (0.062) | 2.118 (0.14) | 0.422 (0.055) |
|  | Time-varying (2A) | – | 2.234 (0.121) | 0.578 (0.07) | 2.033 (0.116) | 0.238 (0.059) |

As some of the endemic effects had large confidence intervals when including just a simple intercept (intercept that was not unit-specific *α*^(*·*)^), we included effects of unit (age group or region) *α*^(*ν*)^ in the endemic component. This is one of the strategies outlined in Meyer et al. [26]. As we have a large number of units in our analysis of regions we decided to include a random intercept rather than a fixed intercept for this analysis. However, when using random effects, Akaike*’*s information criterion can no longer be used for model selection and we elected instead to use the Dawid-Sebastani score (DSS) given in Held et al. [23]. We calculated the one-step-ahead score for the final observation date ISO week 2021-48 based on the predictions and observed data from the rest of the study period (ISO weeks 2020-53 to 2021-47). An overview of the different models is provided in Table 3. We see that the best fitting models with time-varying weights both have vaccination coverage covariate in the endemic component.

**Table 3:** Endemic-epidemic models and goodness of fit (the lowest Dawid-Sebastani score (DSS) values for the models with time-varying transmission weights are marked in bold)

| Transmission weights | Model | DSS |  |
| --- | --- | --- | --- |
|  |  | Region | Age |
| Time-constant | neither | 15.25 | 18.57 |
| Time-constant | endemic | 15.34 | 20.59 |
| Time-constant | epidemic | 15.24 | 16.08 |
| Time-constant | both | 15.24 | 18.6 |
| Time-varying | neither | 13.79 | 21.22 |
| Time-varying | endemic | **13.74** | **18.45** |
| Time-varying | epidemic | 17.08 | 23.9 |
| Time-varying | both | 16.99 | 19.61 |

We present results for the best fitting model (the model with vaccination coverage in the endemic component) for each unit (stratum defined by age group or region) type. We show the fitted values summed across units in Figure 7. while results for individual units are found in the supporting information (files models-age and models-regions). The models fit the data well and show similar patterns (Figure 7) with an increase in endemic cases in the summer and autumn (June to October; ISO weeks 2021-25 to 2021-40). We believe this to be a summer holiday effect as international travel increased after a year with many electing to have a “staycation” in 2020, so there is an increase in imported cases during this time.

**Figure 7:**
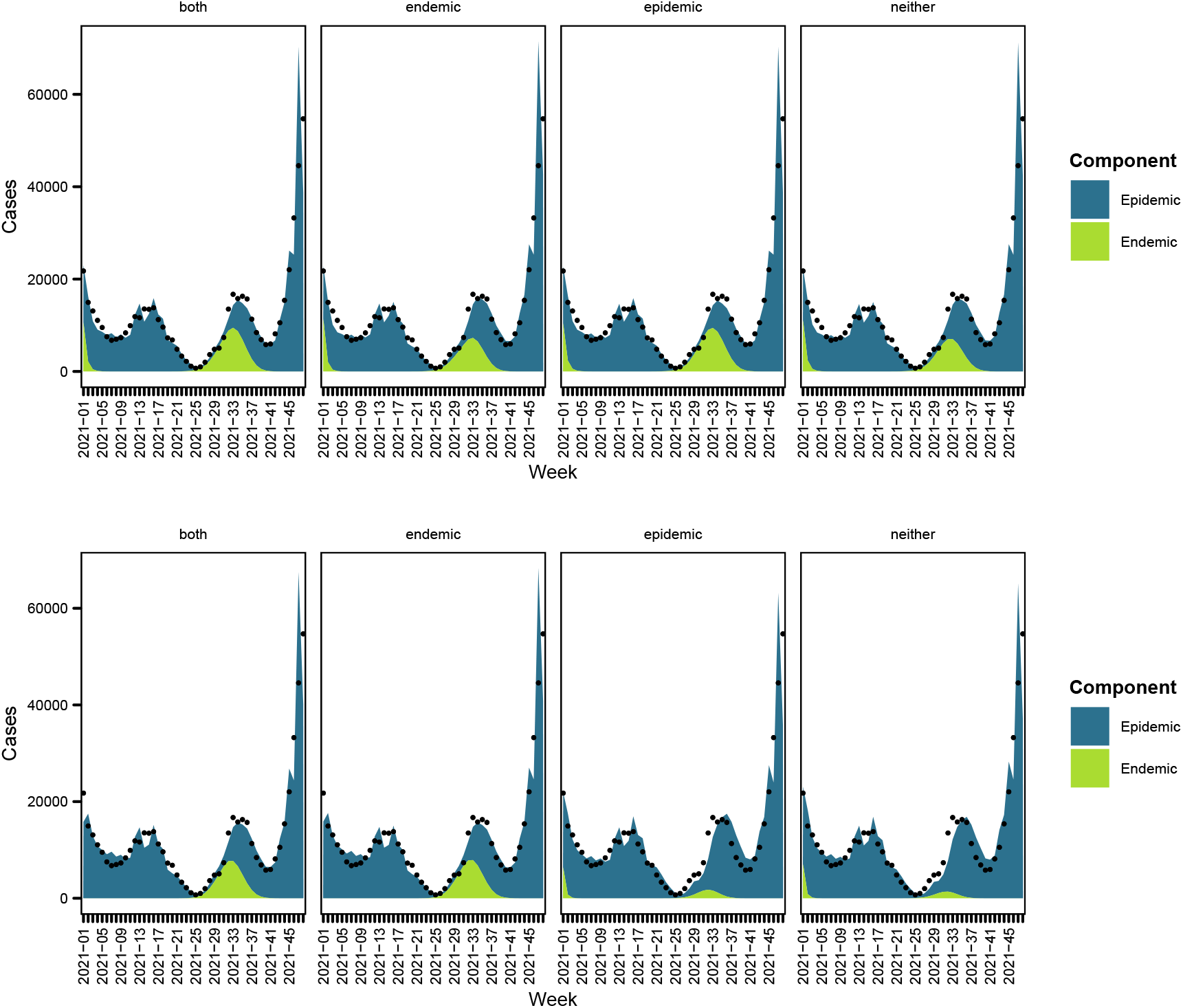
Model fits for models with time-varying transmission weights with region (above) and age groups (below)

### 4.1 Spatial dispersion

The model estimates for the models with time-varying transmission weights (Table 4) show a greater effect of vaccination coverage (*β*) in the epidemic component *β*^(*ν*)^ than the endemic component *β*^(*ϕ*)^ when both are included in the models for regions. The random effects for region are greater for the large regions (Geneva/GE, Basel/BS and BL, Zug/ZH, and Zurich/ZH) in the endemic component (Figure 9). Bordering regions Ticino (TI), Geneva (GE), Basel (BS and BL) and Schaffhausen (SH) have smaller effects in the epidemic component which may be due to cross-border medical seeking behaviour. The gravity model is included in the region-based model. The effect is positive indicating that there is more influx from larger populations (urban centres) and so we would expect to see more cases in such regions. It seems stable across models considered. The linear time trend does not seem to contribute much, indicating the yearly-duration sine-cosine waves may capture most of the fluctuation. The sine-cosine waves are most similar in the epidemic component (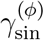 and 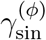), which is more pronounced for the model with regions. The overdispersion *ψ* is largest for the model without vaccination coverage but is similar for the model selected.

**Table 4:**
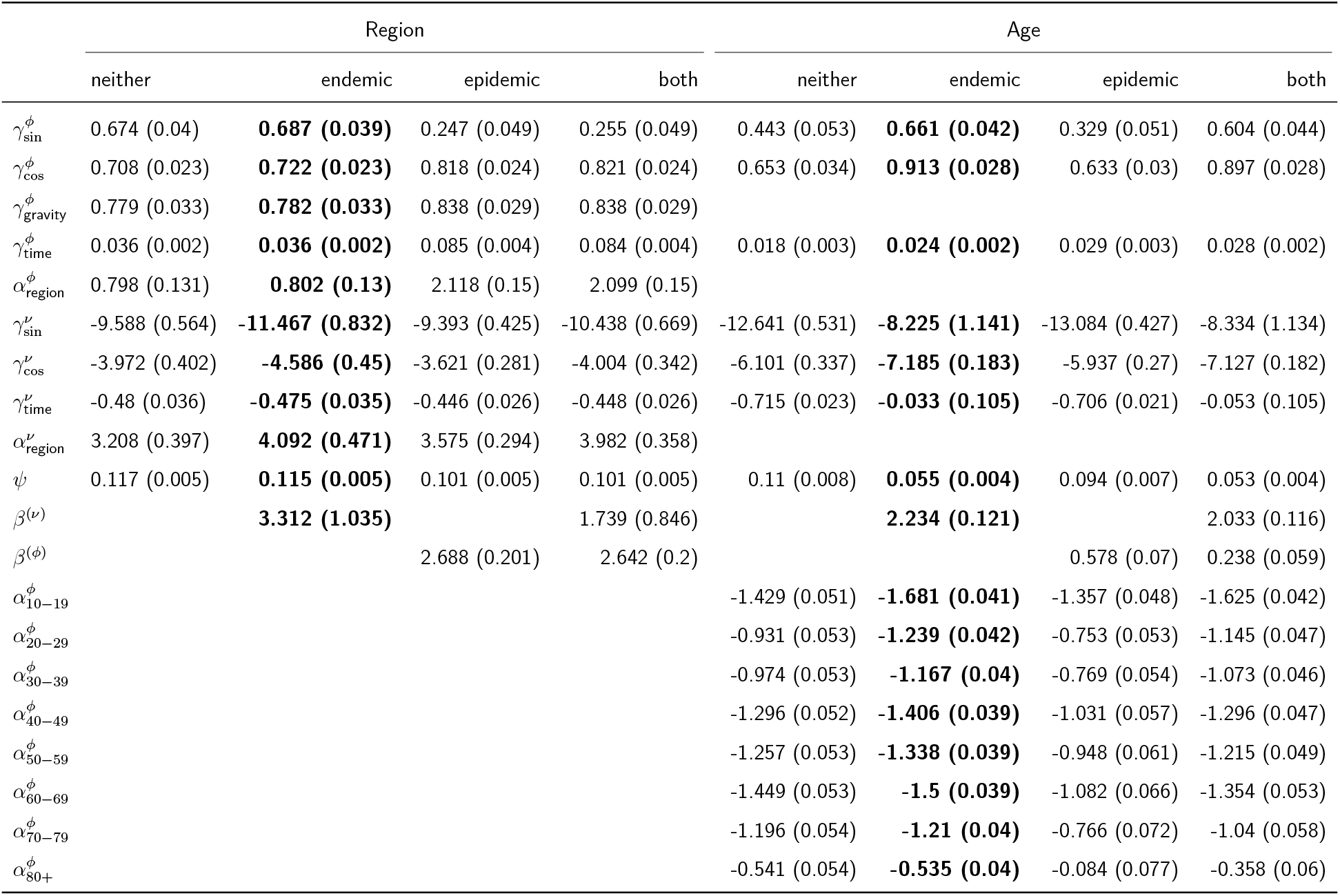
Model coefficient estimates for models with standard errors in brackets

**Figure 8:**
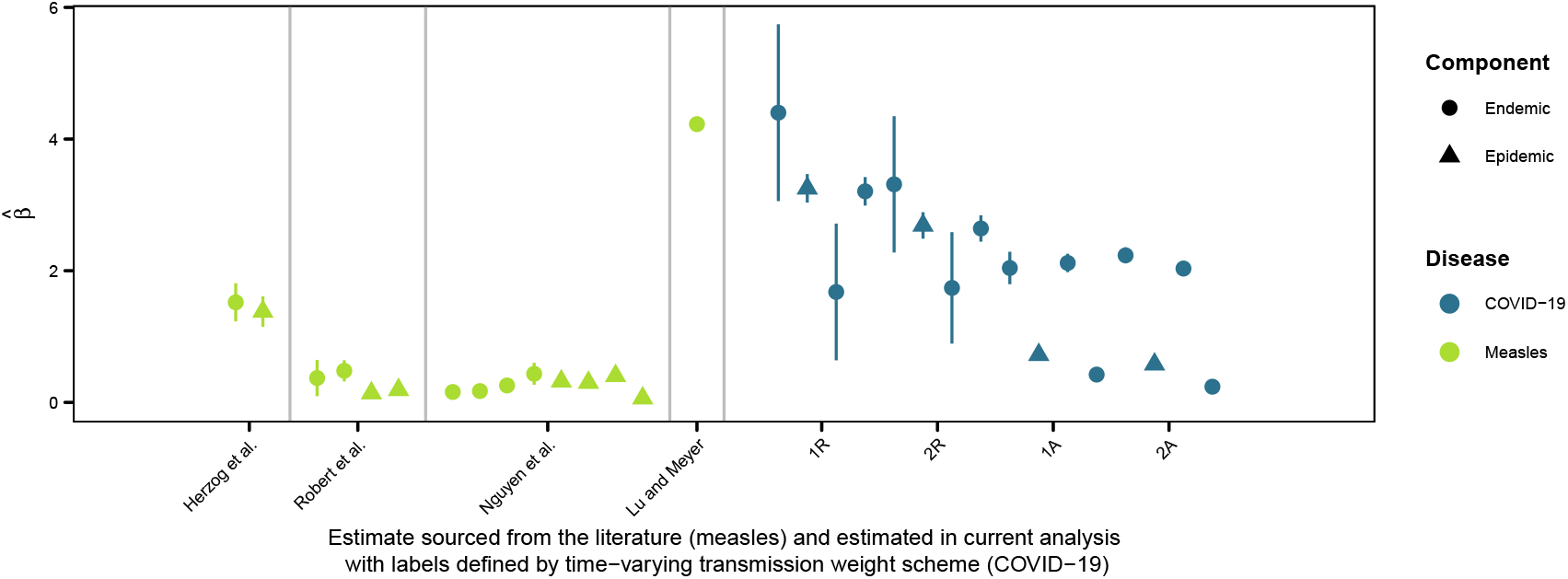
Overview of vaccination coverage estimates found in endemic-epidemic models. Labels for our models are provided in Table 2

**Figure 9:**
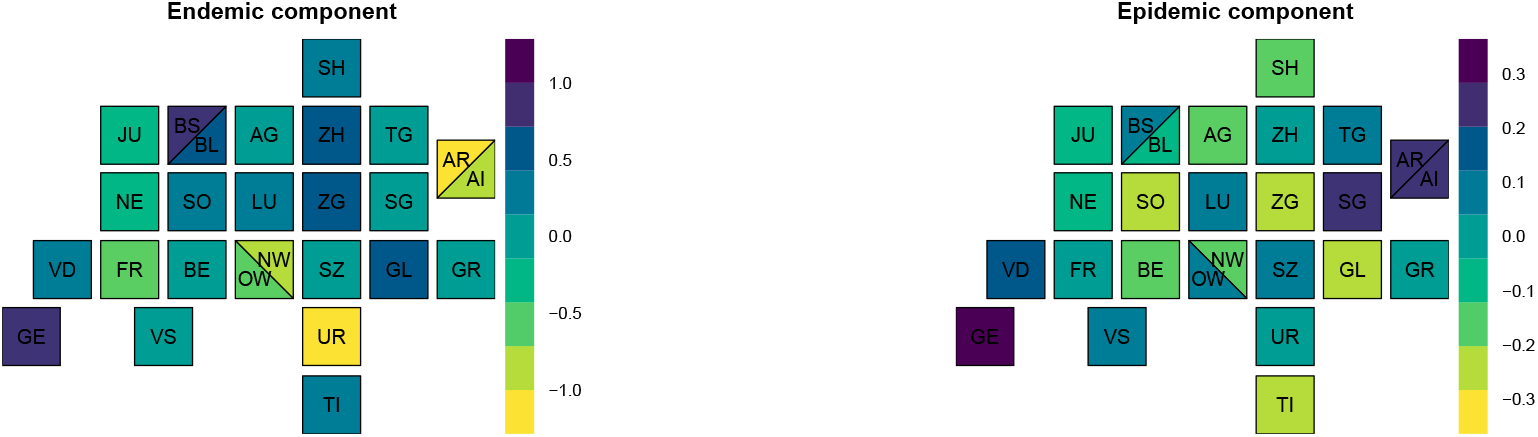
Estimated random effects of region in endemic-epidemic model with time-varying transmission weights and vaccination coverage in the endemic component

### 4.2 Contacts

There is a greater effect of vaccination coverage (*β*) in the epidemic component *β*^(*ν*)^ than the endemic component *β*^(*ϕ*)^ when both are included in the models for age group but effect in the endemic component is much smaller than seen in the regional model in the age group-based models with tighter confidence bands. There is no common pattern for the fixed effect of age group *α·*^(*ϕ*)^ indicating that this could be an important effect to include. Such an effect was not included in the endemic component *α·*^(*ν*)^ due to convergence issues. The sine-cosine waves are again rather stable in the epidemic component. The overdispersion *ψ* is largest for the model without vaccination coverage but experiences a greater decrease when this covariate is included for the models with age groups. The linear time trend again does not seem to contribute much to the models.

### 4.3 Scenario prediction

In the alternative scenario with more vaccines given throughout Switzerland (all regions get the maximum amount given for any region *r* at each time point), we would expect a lower mean incidence in Zurich (ZH), the most populous region which often had the greatest vaccination uptake (Figure 5). We find that regions with lower vaccination coverage such as Glarus (GL), Appenzell Innerrhoden (AI), and Sankt Gallen (SG) have a greater drop in cases under a scenario of increased uptake of vaccination (see the supporting information (file models-regions) for regional prediction plots) in ISO weeks 2021-30 and 2021-38. Overall less cases would be expected if more vaccines had been distributed (Figure 10).

**Figure 10:**
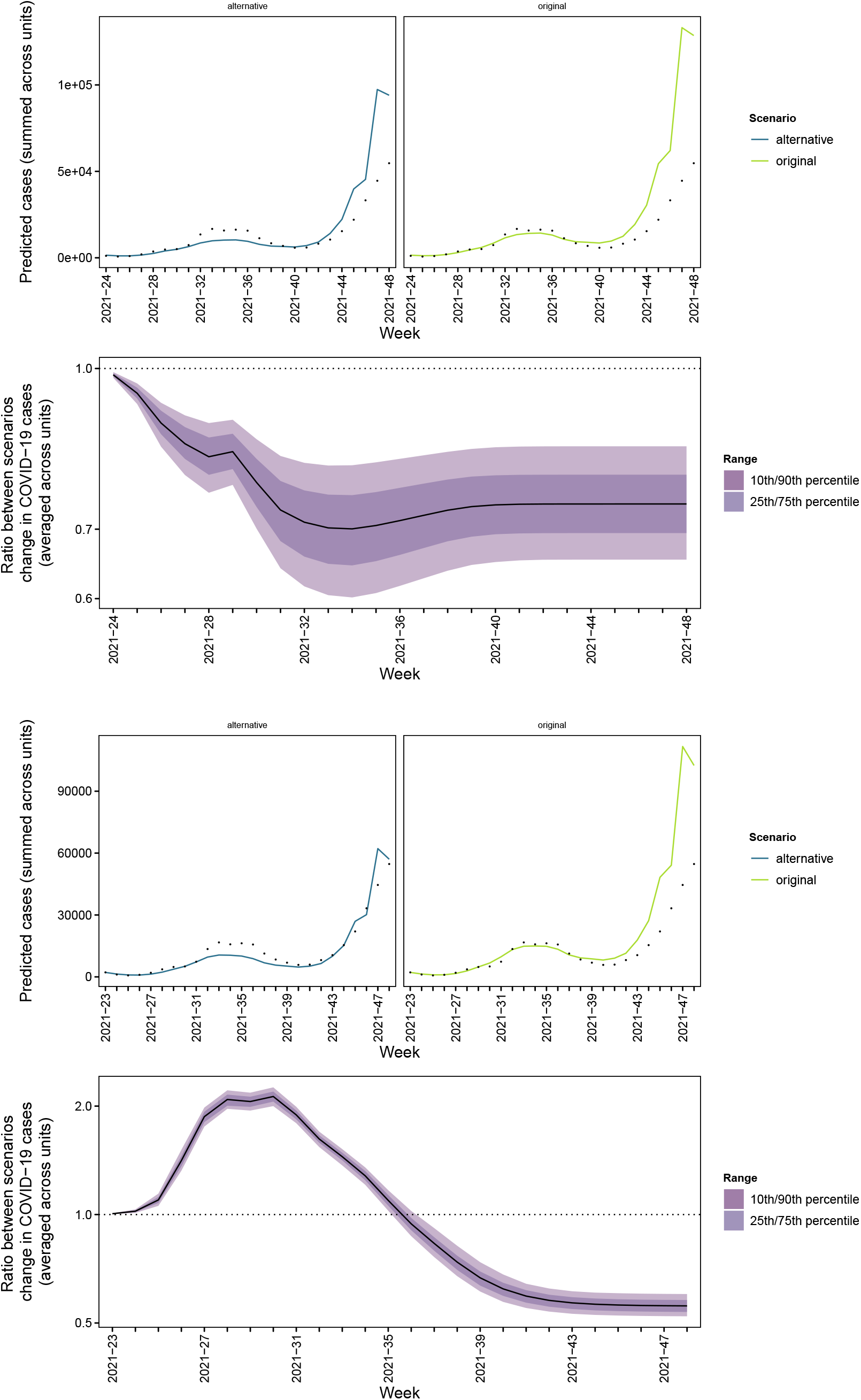
Predicted cases under the two alternative vaccination strategies for regions (above) and age groups (below)

We also see that the age-based distribution scheme is evident in the comparison with an alternative as the greatest expected increases initially are among those groups vaccinated first; over 65 year olds. We see an overall decrease in expected cases for younger age groups. The age group 50 − 59 is the youngest group to not only experience decreases but first see an increase then a decrease. This occurs at different times (ISO week 2021-34 for 50 − 59, 2021-37 for 60 − 69, and 2021-38 for 70 − 79 and 80+) but after week 2021-38 all age groups would be expected to have fewer cases (see the supporting information (file models-age) for prediction plots by age group). On average, an increase would be expected but at the end of the study period less cases would be expected. A distribution scheme which is more uniform across age groups leads to predicted proportions of cases being more equal (see supporting information (file supp) for plot), as would be expected.

## 5 Conclusion

This is the first use of endemic-epidemic modelling with time-varying transmission weights and time-varying vaccination coverage concurrently. The vaccination coverage is constructed in such a manner that is also takes into account waning immunity. The methods chosen for this work are relevant for case counts arising from a surveillance system for notifiable diseases and so are useful for researchers and public health agency staff who interact with such systems daily. The use of these models enable us to explore public health-related questions and concerns using statistically sound methodology.

This work complements our earlier work on time-varying weights in endemic-epidemic models [see 5–7]. The question for infectious disease models with transmission weight matrices has always been and remains how do you choose which matrix to use. Replacing who-acquires-infection-from-whom matrices with empirical or synthetic contact matrices provided freedom from making assumptions of specific mixing patterns. However, during a public health emergency such as COVID-19, assumptions of constant transmission opportunities may be violated due to disease control measures enacted. In such a setting the choice of a constant matrix becomes non-trivial as non-outbreak settings (where surveys of transmission opportunities are traditionally conducted) may no longer be representative at short or long term scale and is is not obvious what should be chosen in such a setting. For this reason, we adjusted the matrices to reflect situational changes using external information (policy and mobility). This does not fully answer the question but is an attempt at determining which transmission weight matrices to use. As the changes to different contact settings are not as obvious as in previous work [see 6, 7, and note the two flat lines in the bottom left panel in Figure 1], we effectively only have changes to contacts driven by specific locations. For other researchers wishing to do similar modelling, it should also be noted that the mobility data informing *m*_*rt*_ is only made available until May 2022 after which time it is no longer provided. This is an example of why private corporations should not be in charge of data gathering for emergency response–access or collection can and may be revoked at any time. In constructing such scenarios, we did not consider the situation of no vaccination as examined by Zwahlen and Staub [27] in their considerations of expected excess deaths in the absence of vaccines. As they note, an endemic situation with no vaccines and no disease control in place is unlikely. We would thus expect to see certain contrasting effects in the time-varying transmission weights and the time-varying vaccination coverage.

We showed that the endemic-epidemic modelling framework can be used to project the COVID-19 pandemic under different scenarios of vaccination coverage. This was possible as we worked with highly structured vaccine coverage data (which was stratified by week, age group, and region). The objective of this project was to determine the role of vaccines in slowing the spread of COVID-19 in Switzerland incorporating the specific demographics of the country to improve the understanding of the impact of vaccination on the ongoing pandemic. Statistical modelling was used with epidemiological data to determine the effect unvaccinated or under-vaccinated groups have on the spread of COVID-19 in other parts of Switzerland as well as the impact of the vaccination strategy used (age-based distribution).

The strength of our modelling work is the ability to project the epidemic under different scenarios for vaccination coverage taking into account waning. The highly structured case and vaccine data (given by weeks *t*, age group *a*, and region *r*) allows us to obtain multivariate predictions providing stratified mean incidence. This is uniquely informative and provides granular estimates of changes in numbers of cases. An alternative approach which would not allow for the evaluation of the vaccination coverage effect estimate (which was the focus of this work) but which could be used to examine similar research questions as considered here would be to inform the time-varying transmission weights by the level of vaccination coverage. We have not considered such an approach in this work as it is out of scope but mention it should other researchers wish to attempt it. Meyer et al. [26] note that when you assume a common intercept for the unit, you do not have to exclude units without reported cases. This should not be an issue here based on visual inspection of the outcome variable data. Later developments [12] have examined how to incorporate low case counts in endemic-epidemic models and may be another option to consider for researchers interested in similar questions.

We used the log-proportion of the unvaccinated population to represent the susceptible population. This is based in the law of mass action which relies on a homogeneous mixing assumption. We believe that our inclusion of time-varying heterogeneous contact and spatial dispersion matrices as well as allowing the vaccination coverage to vary over time relaxes some of the unrealistic aspects of such an assumption. Our work implicitly assumed that in the scenarios of increased uptake of vaccines the alternative amount of vaccine would be available which might not be the case in reality and so we note that our work provides optimistic estimates. Because the Swiss national identity is very divided (Switzerland itself has notable linguistic divisions), it is perhaps not so surprising that Ticino (TI) had early uptake of vaccines (Figure 3.2) as those residents would all other things equal be assumed to have read more of the Italian language reporting on COVID-19; Italy being the early European epicentre in 2022 could have had an impact. However, Rudolf Steiner-based initiatives are head-quartered in Switzerland (particularly in Solothun (SO) which never achieves the maximum vaccination coverage in the study period) and are known to harbour pseudo-scientific and anti-vaccination sentiments [1] which could serve as a hindrance to improvements in vaccination and health. Waldorf school-driven outbreaks of infectious diseases are a known epidemiologic issue [28]. With even more granular information, the effects of undervaccinated school districts could be explored.

Much has been discussed about herd immunity during the COVID-19 pandemic after the introduction of vaccines. Herd immunity is the proportion of population that needs to be vaccinated in order to curb disease spread. The modelling approach used here can in future also be used to examine the effects of achieving herd immunity, once this threshold has been fully determined for this disease.

## Supporting information

Supporting information (models-age)

Supporting information (models-regions)

Supporting information (sens)

## Data Availability

Our analysis is fully reproducible using the code available at github.com/mariabnd/ee-vax. This includes a script which downloads the input data used in the analysis as well as scripts we used for pre-processing this data. Our derived data is also released to safeguard for the future; the derived data will remain available even when the input data may not longer be accessible. The code used in the documents (manuscript and supporting information) is also provided such that users can reproduce figures should they wish to.

## Reproducibility

Our analysis is fully reproducible using the code available at github.com/mariabnd/ ee-vax. This includes a script which downloads the input data used in the analysis as well as scripts we used for pre-processing this data. Our derived data is also released to safeguard for the future; the derived data will remain available even when the input data may not longer be accessible. The code used in the documents (manuscript and supporting information) is also provided such that users can reproduce figures should they wish to.

## Footnotes

1 Use of this data is not an endorsement of Facebook/Meta

