## Supporting information (models-age) for "The COVID-19 vaccination campaign in Switzerland and its impact on disease spread"

### MODEL – age only $Y_{at}$, transmission weights $\mathbf{w} = (w_{a,a',t})$

M. Bekker-Nielsen Dunbar and L. Held
Epidemiology, Biostatistics and Prevention Institute, University of Zurich

This document contains additional models to those outlined in the study protocol [1]. Knowledge of the analyses from the main manuscript is assumed. The models introduced here have only age group effects. This means we are comparing the models:

$$\begin{aligned} \log(\nu_{at}) &= \alpha_a^{(\nu)} + \qquad\qquad\qquad\qquad + \gamma^{(\nu)\top}\mathbf{z}_{at}^{(\nu)} \\ \log(\phi_{at}) &= \alpha_a^{(\phi)} + \qquad\qquad\qquad\qquad + \gamma^{(\phi)\top}\mathbf{z}_{at}^{(\phi)} \end{aligned} \qquad \text{(neither)}$$

$$\begin{aligned} \log(\nu_{at}) &= \alpha_a^{(\nu)} + \beta_a^{(\nu)}\log(1 - x_{at}) + \gamma^{(\nu)\top}\mathbf{z}_{at}^{(\nu)} \\ \log(\phi_{at}) &= \alpha_a^{(\phi)} + \qquad\qquad\qquad\qquad + \gamma^{(\phi)\top}\mathbf{z}_{at}^{(\phi)} \end{aligned} \qquad \text{(endemic)}$$

$$\begin{aligned} \log(\nu_{at}) &= \alpha_a^{(\nu)} + \qquad\qquad\qquad\qquad + \gamma^{(\nu)\top}\mathbf{z}_{at}^{(\nu)} \\ \log(\phi_{at}) &= \alpha_a^{(\phi)} + \beta_a^{(\phi)}\log(1 - x_{at}) + \gamma^{(\phi)\top}\mathbf{z}_{at}^{(\phi)} \end{aligned} \qquad \text{(epidemic)}$$

$$\begin{aligned} \log(\nu_{at}) &= \alpha_a^{(\nu)} + \beta_a^{(\nu)}\log(1 - x_{at}) + \gamma^{(\nu)\top}\mathbf{z}_{at}^{(\nu)} \\ \log(\phi_{at}) &= \alpha_a^{(\phi)} + \beta_a^{(\phi)}\log(1 - x_{at}) + \gamma^{(\phi)\top}\mathbf{z}_{at}^{(\phi)} \end{aligned} \qquad \text{(both)}$$

$\mathbf{z}_{rt}$ is as given in the main manuscript.

### 1 Setup

The study period is 1st January 2021 to 30th November 2021 given by ISO weeks 2020-53 to 2021-48, both weeks included. This is a deviation from the protocol. The new study period is chosen on the basis of the epidemic calendar. This is similar to influenza research in the northern hemisphere; we want our years to start and end such that the entire expected wave is captured in a single year. Furthermore, the latter cut-off was chosen due to the presence of Omicron variant COVID-19 in the original study period. The reason for this is that the vaccines considered in this work were not created with this variant in mind and may provide less protection against this disease. The study period covers 49 observations. An additional deviation from the study protocol is that we do not include the youngest age group, as they do not receive any vaccines in the study period considered.

#### 1.1 Outcome

The outcome variable is cases. Here we consider all cases (i.e. not limited to hospitalisations). The cases are not equally distributed across age groups:

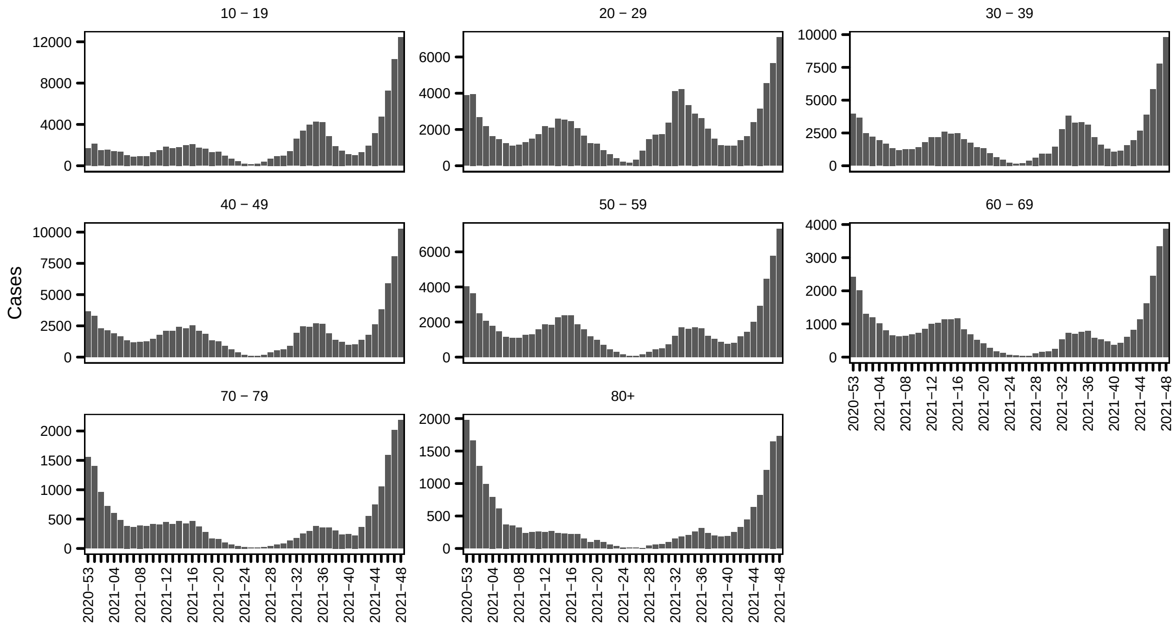

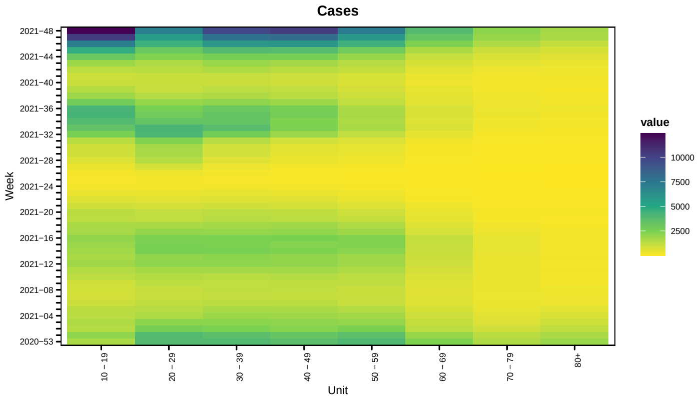


#### 1.2 Covariates

The vaccination coverage looks as follows (the patterns are not so surprising considering Switzerland followed an age-based distribution scheme).

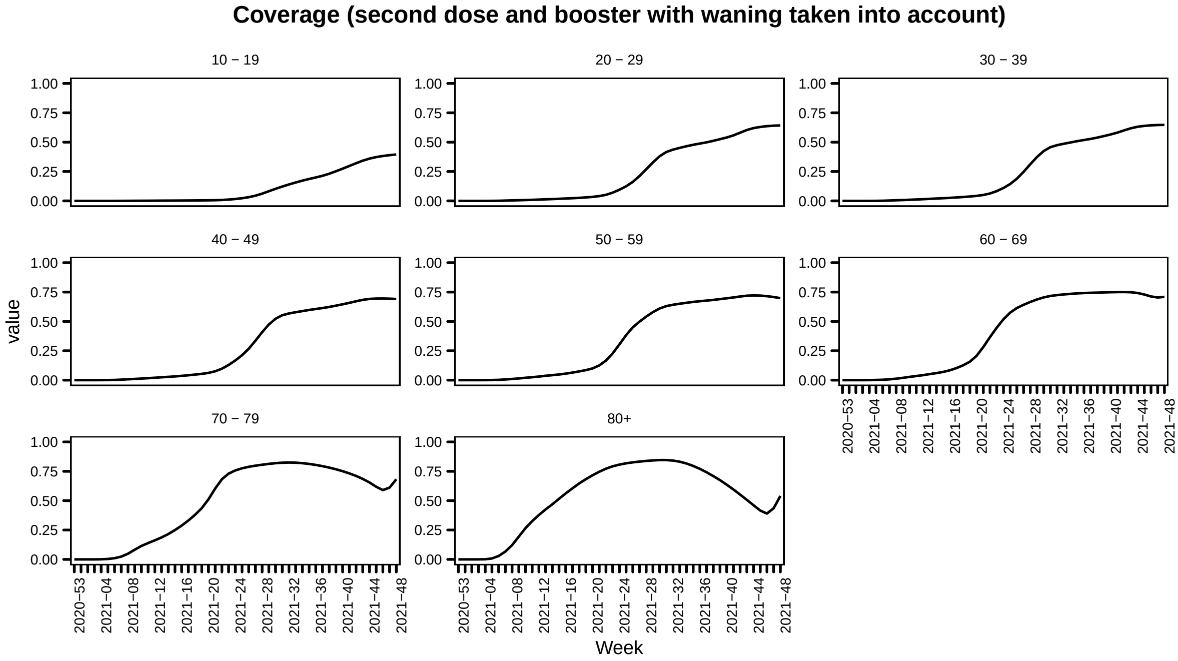

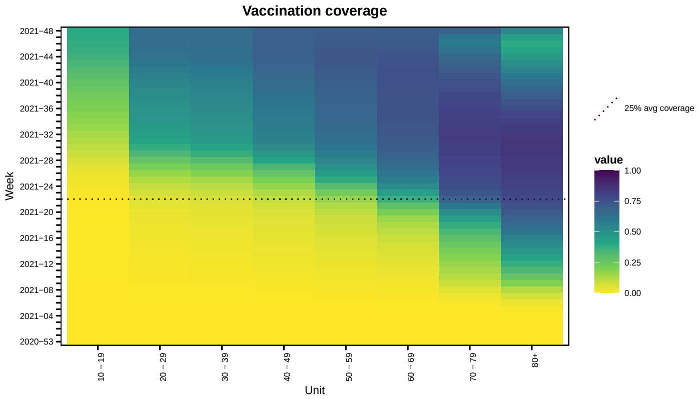


Transformed by $f(x) = \log(1 - x)$ this looks like

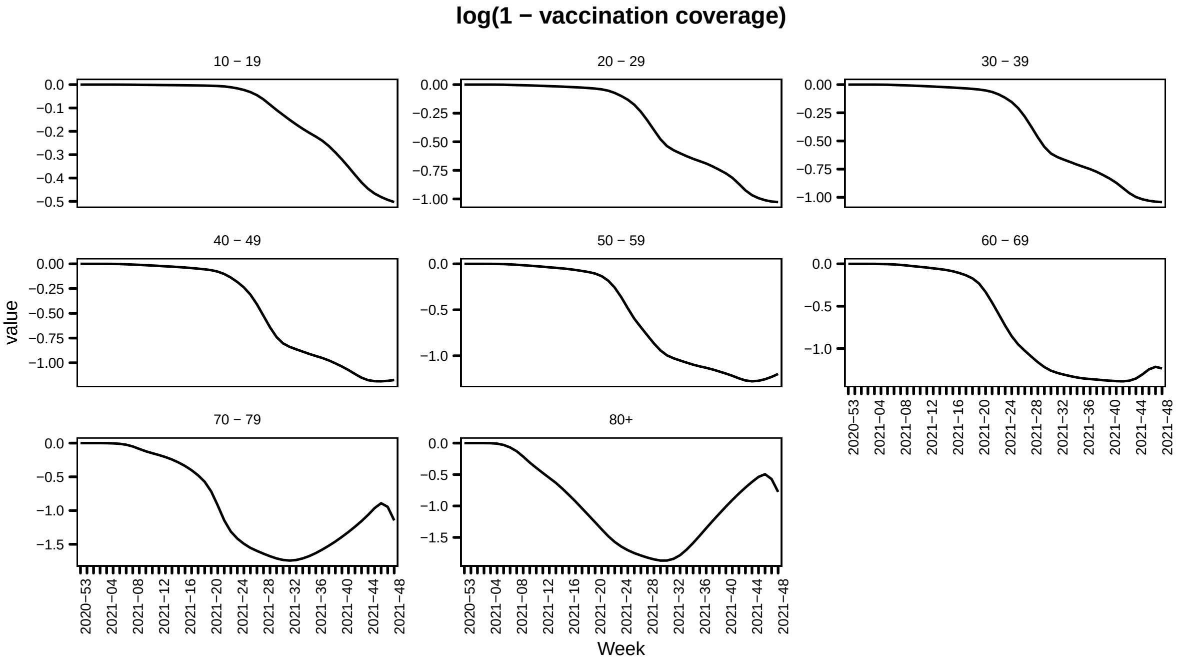


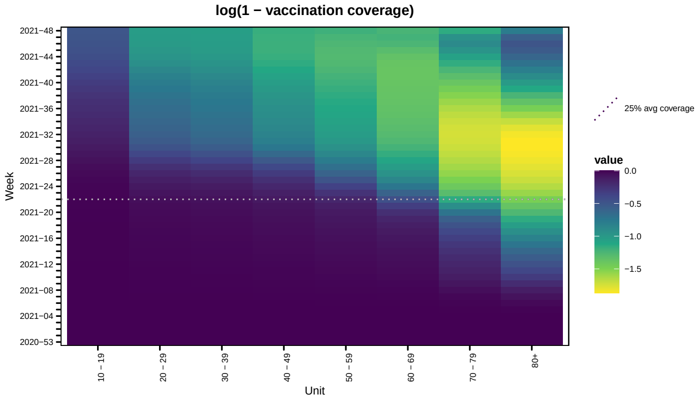

#### 2 Results overview

The following table provides the estimated effects of vaccination coverage for models with and without time-varying transmission weights.

| Weights | Model | Endemic | Epidemic | $\ell$ | DSS |
|---|---|---|---|---|---|
| Constant | Neither | | | -2639.142 | 18.572 |
| Constant | Endemic | 2.042 (SE 0.246) | | -2598.267 | 20.585 |
| Constant | Epidemic | | 0.722 (SE 0.062) | -2618.674 | 16.085 |
| Constant | Both | 2.118 (SE 0.14) | 0.422 (SE 0.055) | -2529.263 | 18.605 |
| Varying | Neither | | | -2703.612 | 21.222 |
| Varying | Endemic | 2.234 (SE 0.121) | | -2569.63 | 18.454 |
| Varying | Epidemic | | 0.578 (SE 0.07) | -2670.896 | 23.896 |
| Varying | Both | 2.033 (SE 0.116) | 0.238 (SE 0.059) | -2561.504 | 19.611 |

##### 2.1 Time-constant age group transmission weights

First we present the results of the modelling with the time-constant transmission weights. Due to diverging effects when including fixed effects of age in the endemic component, we remove those effects.

| | neither | endemic | epidemic | both |
|---|---|---|---|---|
| $\alpha^{\phi}_{10-19}$ | -0.35 (SE 0.253) | -1.536 (SE 0.251) | 0.146 (SE 0.041) | -0.082 (SE 0.038) |
| $\alpha^{\phi}_{20-29}$ | -0.128 (SE 0.253) | -1.305 (SE 0.25) | 0.496 (SE 0.045) | 0.162 (SE 0.043) |
| $\alpha^{\phi}_{30-39}$ | -0.283 (SE 0.255) | -1.47 (SE 0.251) | 0.414 (SE 0.046) | 0.149 (SE 0.042) |
| $\alpha^{\phi}_{40-49}$ | -0.648 (SE 0.26) | -1.854 (SE 0.252) | 0.19 (SE 0.049) | -0.044 (SE 0.043) |
| $\alpha^{\phi}_{50-59}$ | -0.679 (SE 0.265) | -1.892 (SE 0.253) | 0.283 (SE 0.053) | 0.041 (SE 0.045) |
| $\alpha^{\phi}_{60-69}$ | -0.97 (SE 0.265) | -2.154 (SE 0.25) | 0.101 (SE 0.057) | -0.148 (SE 0.049) |
| $\alpha^{\phi}_{70-79}$ | -0.812 (SE 0.258) | -1.938 (SE 0.247) | 0.326 (SE 0.063) | 0.078 (SE 0.053) |
| $\alpha^{\phi}_{80+}$ | -0.418 (SE 0.255) | -1.49 (SE 0.246) | 0.757 (SE 0.066) | 0.514 (SE 0.055) |
| $\gamma^{\phi}_{\text{time}}$ | 0.058 (SE 0.023) | 0.158 (SE 0.021) | 0.027 (SE 0.003) | 0.029 (SE 0.002) |
| $\gamma^{\phi}_{\sin(2\pi t/52)}$ | 0.265 (SE 0.135) | 0.659 (SE 0.109) | -0.016 (SE 0.044) | 0.256 (SE 0.042) |
| $\gamma^{\phi}_{\cos(2\pi t/52)}$ | -0.315 (SE 0.22) | -1.28 (SE 0.201) | 0.178 (SE 0.026) | 0.398 (SE 0.027) |
| $\alpha^{\nu}_{10-19}$ | -7.352 (SE 2.288) | -3.214 (SE 0.837) | | |
| $\alpha^{\nu}_{20-29}$ | -7.294 (SE 2.29) | -3.162 (SE 0.836) | | |
| $\alpha^{\nu}_{30-39}$ | -7.373 (SE 2.249) | -3.306 (SE 0.835) | | |
| $\alpha^{\nu}_{40-49}$ | -7.256 (SE 2.212) | -3.255 (SE 0.833) | | |
| $\alpha^{\nu}_{50-59}$ | -7.311 (SE 2.181) | -3.372 (SE 0.831) | | |
| $\alpha^{\nu}_{60-69}$ | -7.627 (SE 2.162) | -3.71 (SE 0.83) | | |
| $\alpha^{\nu}_{70-79}$ | -7.975 (SE 2.148) | -3.893 (SE 0.83) | | |
| $\alpha^{\nu}_{80+}$ | -7.611 (SE 2.137) | -3.296 (SE 0.832) | | |
| $\gamma^{\nu}_{\text{time}}$ | -1.117 (SE 0.129) | -0.93 (SE 0.057) | -0.623 (SE 0.059) | -0.059 (SE 0.118) |
| $\gamma^{\nu}_{\sin(2\pi t/52)}$ | 3.878 (SE 0.66) | 2.57 (SE 0.24) | -12.054 (SE 0.773) | -8.338 (SE 1.267) |
| $\gamma^{\nu}_{\cos(2\pi t/52)}$ | -8.768 (SE 0.959) | -8.242 (SE 0.5) | -5.444 (SE 0.308) | -7.03 (SE 0.204) |
| $\psi$ | 0.079 (SE 0.006) | 0.063 (SE 0.005) | 0.071 (SE 0.005) | 0.044 (SE 0.003) |
| $\beta^{(\nu)}$ | | 2.042 (SE 0.246) | | 2.118 (SE 0.14) |
| $\beta^{(\phi)}$ | | | 0.722 (SE 0.062) | 0.422 (SE 0.055) |

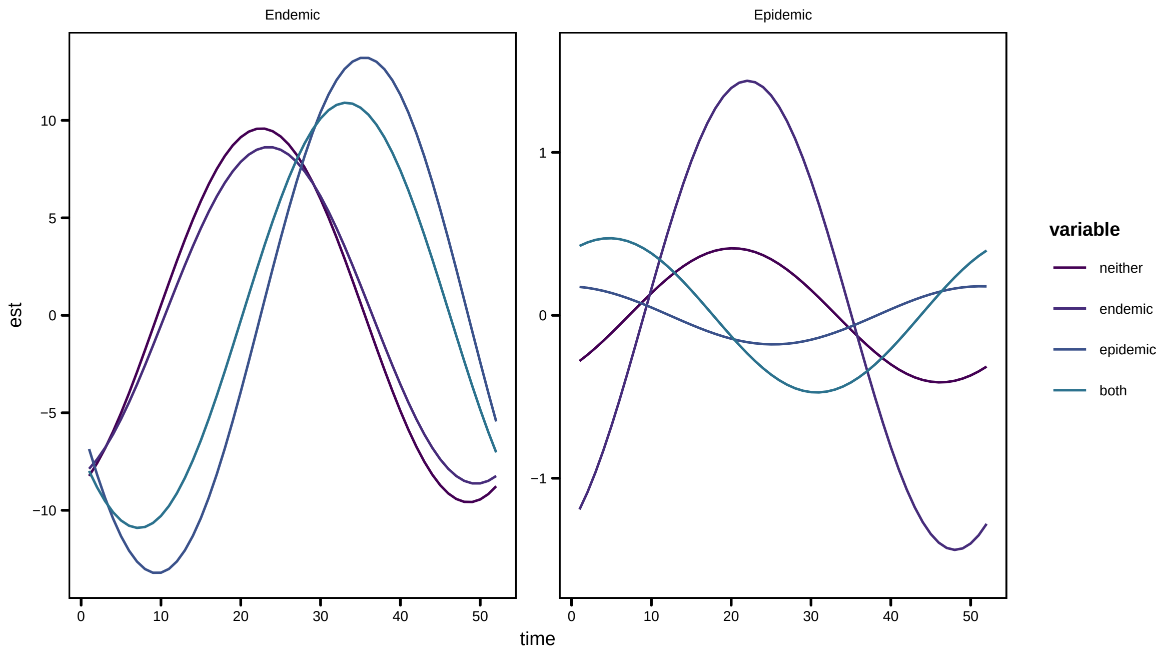
Endemic
Epidemic
est
time
variable
neither
endemic
epidemic
both

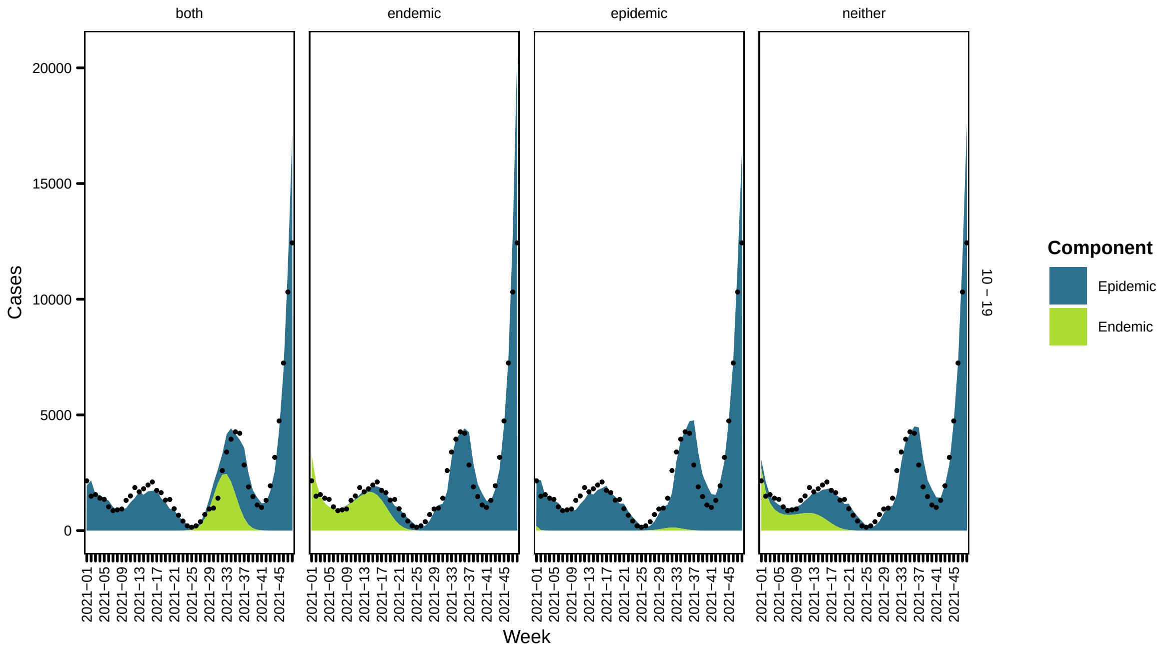
both
endemic
epidemic
neither
Cases
Week
10 – 19
Component
Epidemic
Endemic

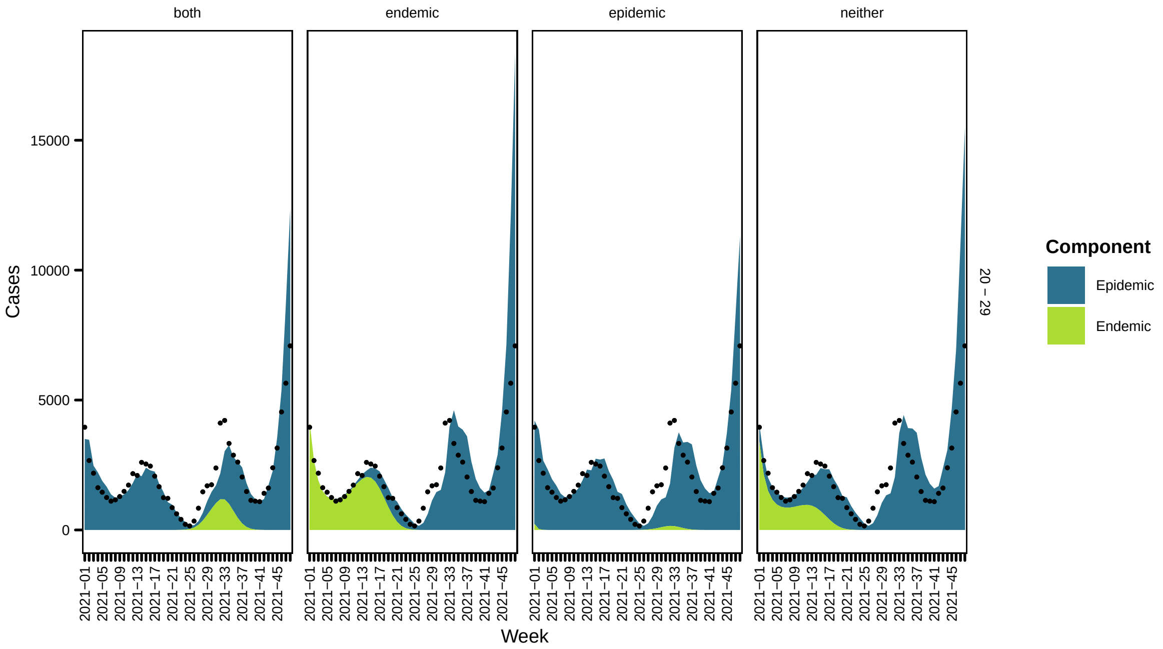
both
endemic
epidemic
neither
Cases
15000
10000
5000
0
Week
2021−01
2021−05
2021−09
2021−13
2021−17
2021−21
2021−25
2021−29
2021−33
2021−37
2021−41
2021−45
20 − 29
Component
Epidemic
Endemic

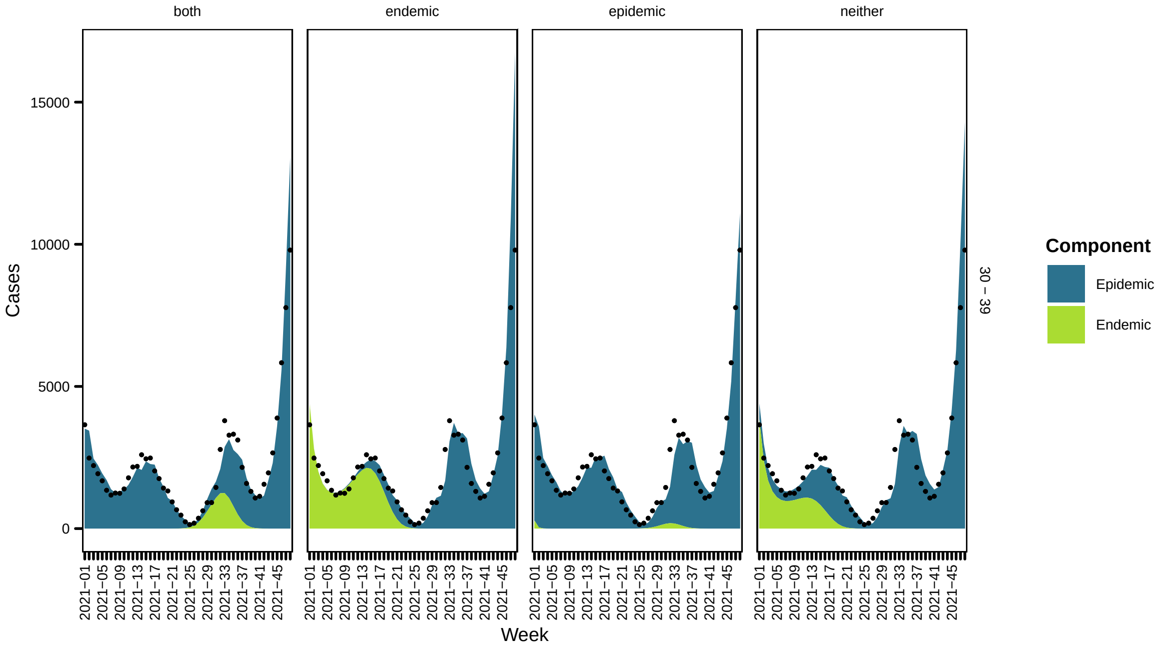
both
endemic
epidemic
neither
Cases
15000
10000
5000
0
Week
2021−01
2021−05
2021−09
2021−13
2021−17
2021−21
2021−25
2021−29
2021−33
2021−37
2021−41
2021−45
30 − 39
Component
Epidemic
Endemic

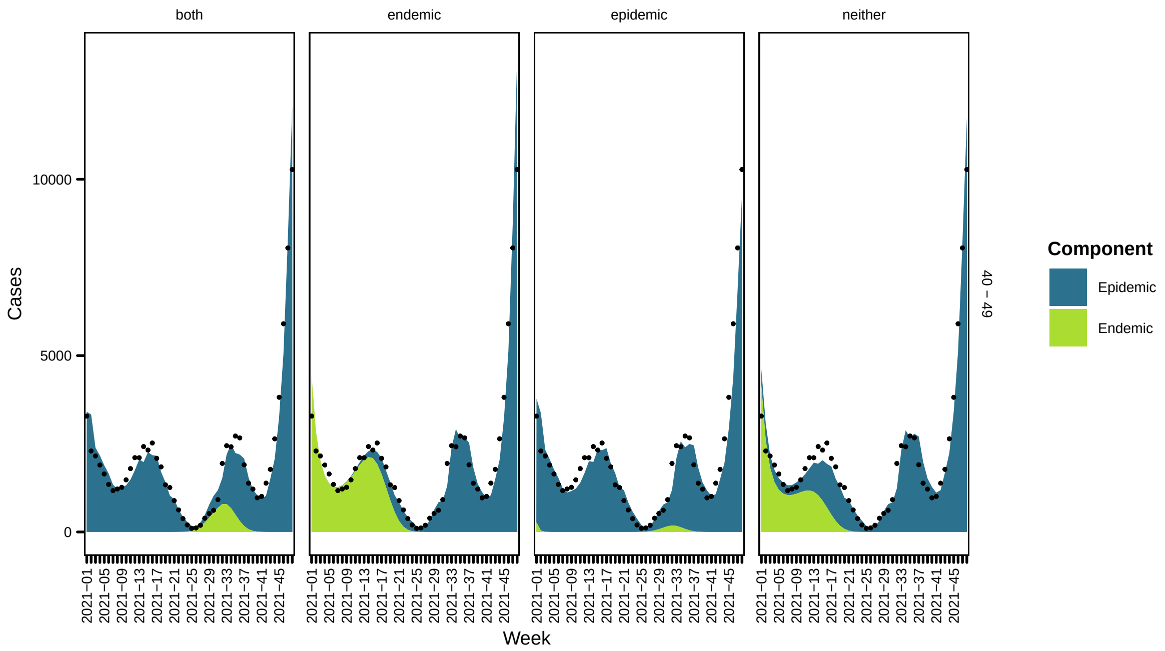
both
endemic
epidemic
neither
Cases
10000
5000
0
Week
40 – 49
Component
Epidemic
Endemic

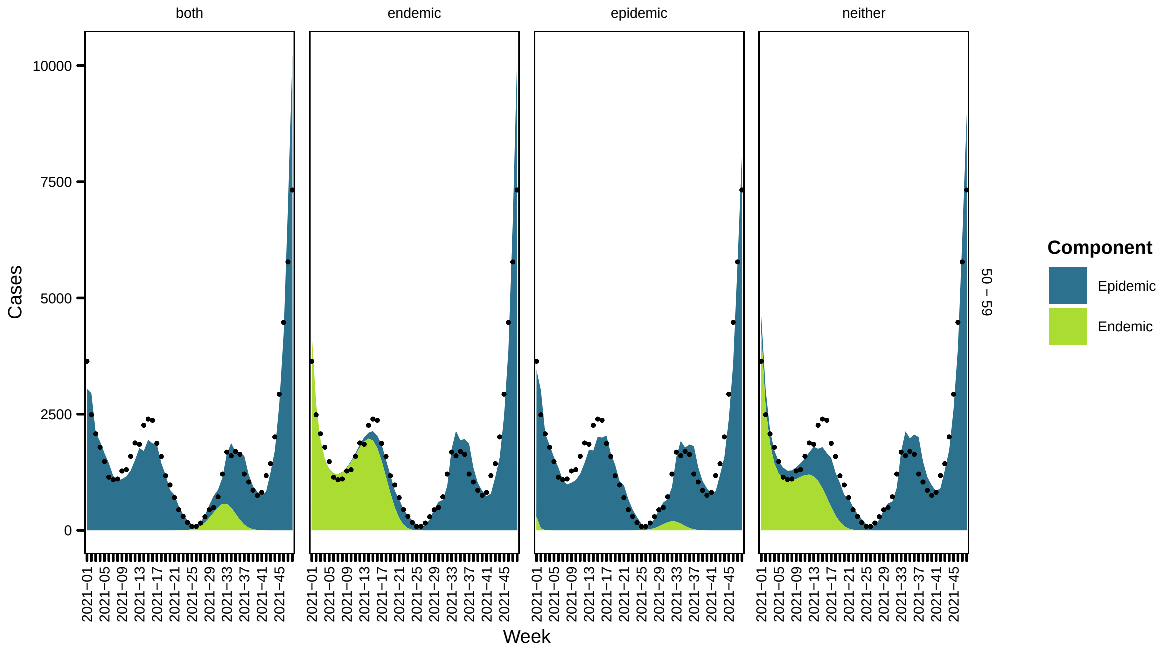
both
endemic
epidemic
neither
Cases
10000
7500
5000
2500
0
Week
50 – 59
Component
Epidemic
Endemic

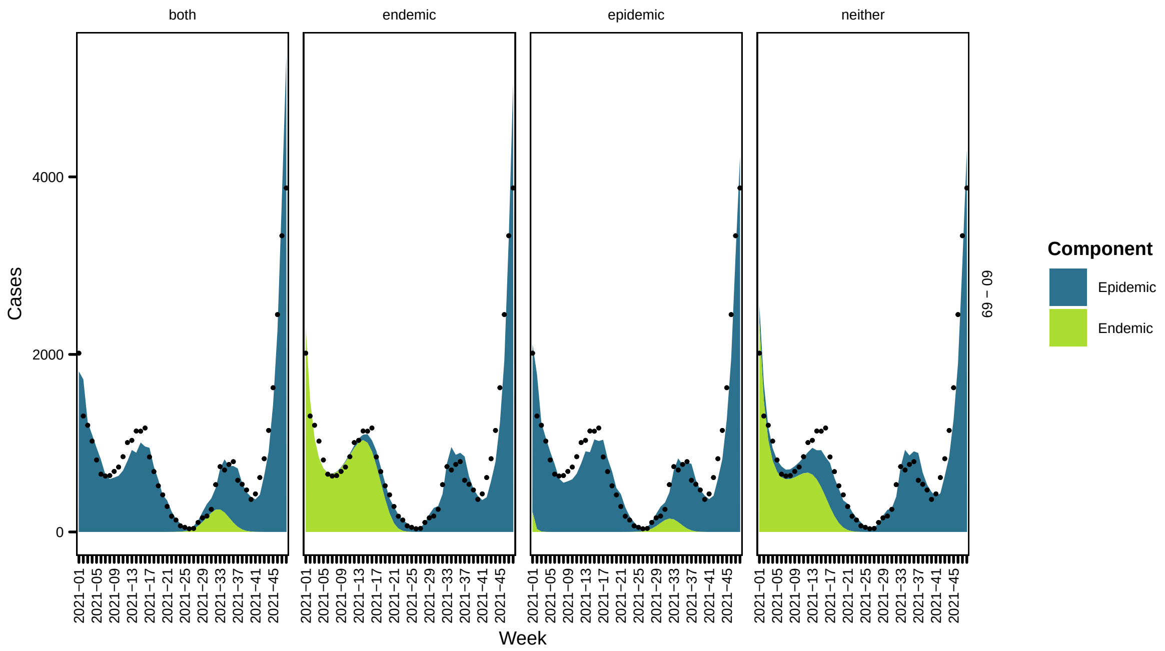
both
endemic
epidemic
neither
Cases
0
2000
4000
2021−01
2021−05
2021−09
2021−13
2021−17
2021−21
2021−25
2021−29
2021−33
2021−37
2021−41
2021−45
Week
60 − 69
Component
Epidemic
Endemic

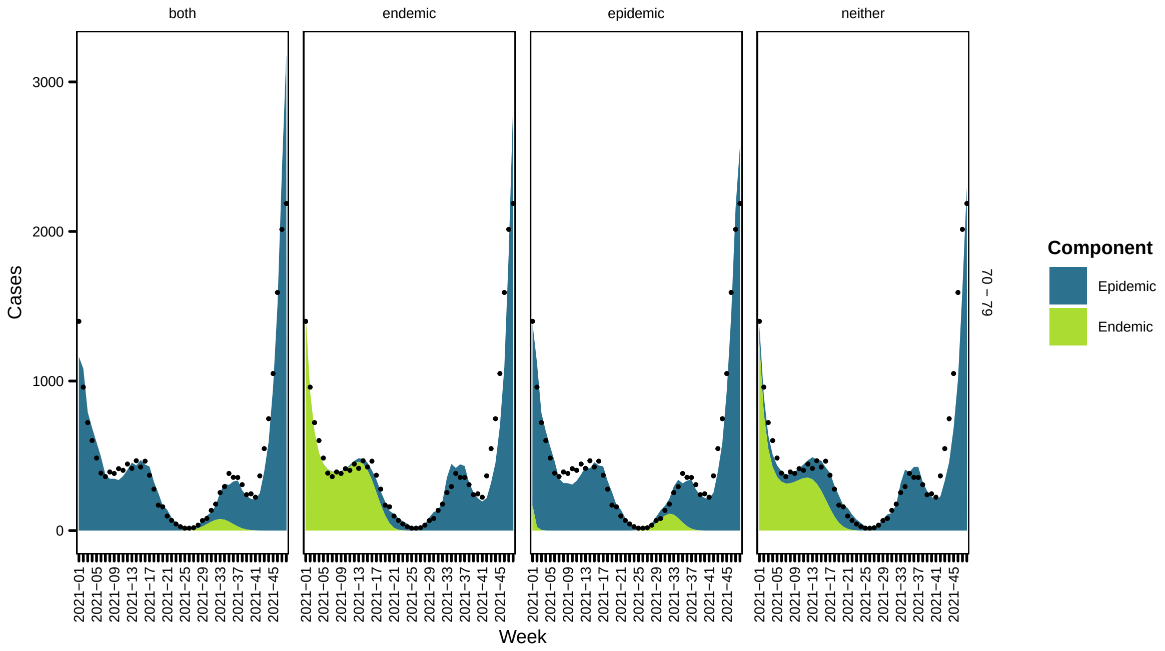
both
endemic
epidemic
neither
Cases
0
1000
2000
3000
2021−01
2021−05
2021−09
2021−13
2021−17
2021−21
2021−25
2021−29
2021−33
2021−37
2021−41
2021−45
Week
70 − 79
Component
Epidemic
Endemic

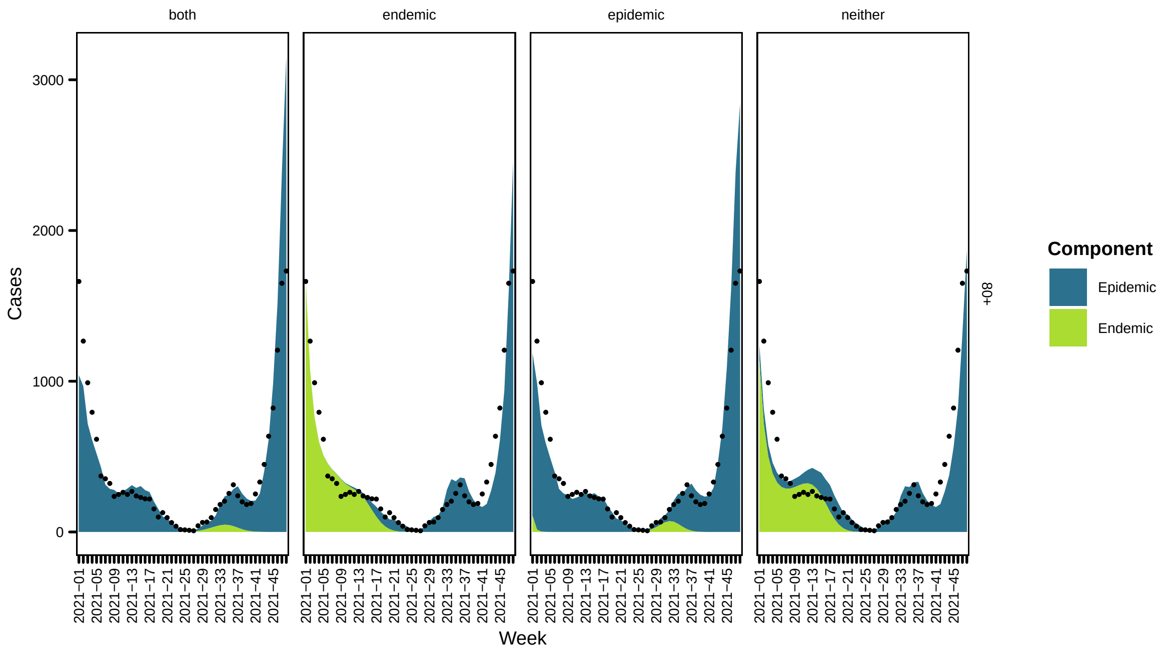


#### 2.2 Time-varying age group transmission weights

Now the model contains the time-varying transmission weights. A snapshot is shown below

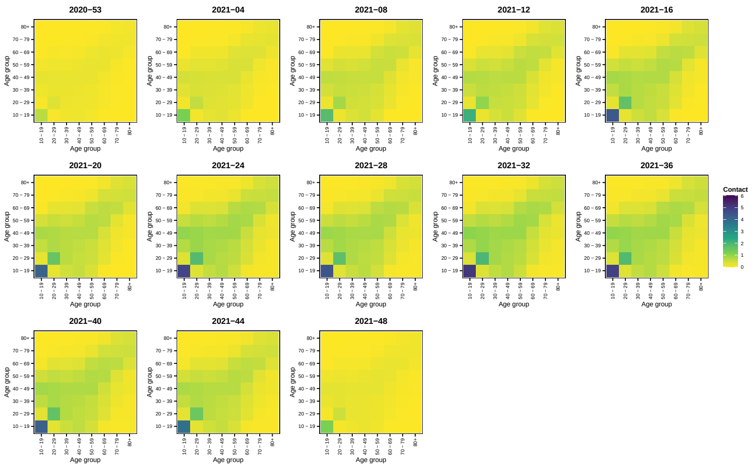

51

| | neither | endemic | epidemic | both |
|---|---|---|---|---|
| $\alpha^{\phi}_{10-19}$ | -1.429 (SE 0.051) | -1.681 (SE 0.041) | -1.357 (SE 0.048) | -1.625 (SE 0.042) |
| $\alpha^{\phi}_{20-29}$ | -0.931 (SE 0.053) | -1.239 (SE 0.042) | -0.753 (SE 0.053) | -1.145 (SE 0.047) |
| $\alpha^{\phi}_{30-39}$ | -0.974 (SE 0.053) | -1.167 (SE 0.04) | -0.769 (SE 0.054) | -1.073 (SE 0.046) |
| $\alpha^{\phi}_{40-49}$ | -1.296 (SE 0.052) | -1.406 (SE 0.039) | -1.031 (SE 0.057) | -1.296 (SE 0.047) |
| $\alpha^{\phi}_{50-59}$ | -1.257 (SE 0.053) | -1.338 (SE 0.039) | -0.948 (SE 0.061) | -1.215 (SE 0.049) |
| $\alpha^{\phi}_{60-69}$ | -1.449 (SE 0.053) | -1.5 (SE 0.039) | -1.082 (SE 0.066) | -1.354 (SE 0.053) |
| $\alpha^{\phi}_{70-79}$ | -1.196 (SE 0.054) | -1.21 (SE 0.04) | -0.766 (SE 0.072) | -1.04 (SE 0.058) |
| $\alpha^{\phi}_{80+}$ | -0.541 (SE 0.054) | -0.535 (SE 0.04) | -0.084 (SE 0.077) | -0.358 (SE 0.06) |
| $\gamma^{\phi}_{\text{time}}$ | 0.018 (SE 0.003) | 0.024 (SE 0.002) | 0.029 (SE 0.003) | 0.028 (SE 0.002) |
| $\gamma^{\phi}_{\sin(2\pi t/52)}$ | 0.443 (SE 0.053) | 0.661 (SE 0.042) | 0.329 (SE 0.051) | 0.604 (SE 0.044) |
| $\gamma^{\phi}_{\cos(2\pi t/52)}$ | 0.653 (SE 0.034) | 0.913 (SE 0.028) | 0.633 (SE 0.03) | 0.897 (SE 0.028) |
| $\gamma^{\nu}_{\text{time}}$ | -0.715 (SE 0.023) | -0.033 (SE 0.105) | -0.706 (SE 0.021) | -0.053 (SE 0.105) |
| $\gamma^{\nu}_{\sin(2\pi t/52)}$ | -12.641 (SE 0.531) | -8.225 (SE 1.141) | -13.084 (SE 0.427) | -8.334 (SE 1.134) |
| $\gamma^{\nu}_{\cos(2\pi t/52)}$ | -6.101 (SE 0.337) | -7.185 (SE 0.183) | -5.937 (SE 0.27) | -7.127 (SE 0.182) |
| $\psi$ | 0.11 (SE 0.008) | 0.055 (SE 0.004) | 0.094 (SE 0.007) | 0.053 (SE 0.004) |
| $\beta^{(\nu)}$ | | 2.234 (SE 0.121) | | 2.033 (SE 0.116) |
| $\beta^{(\phi)}$ | | | 0.578 (SE 0.07) | 0.238 (SE 0.059) |

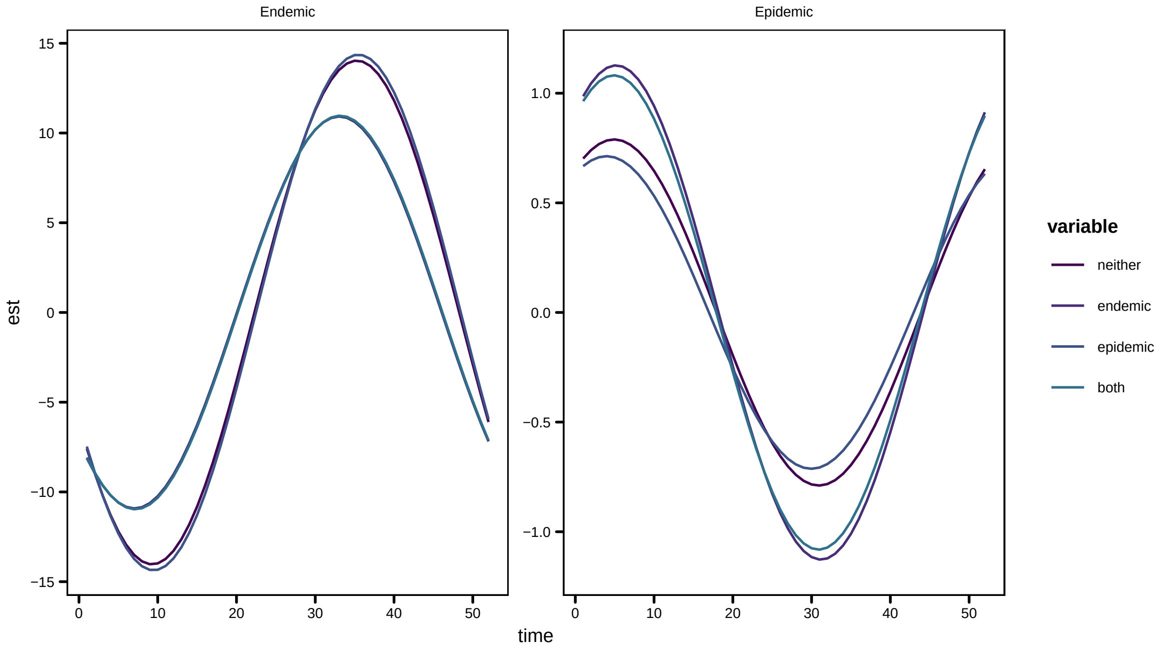


52

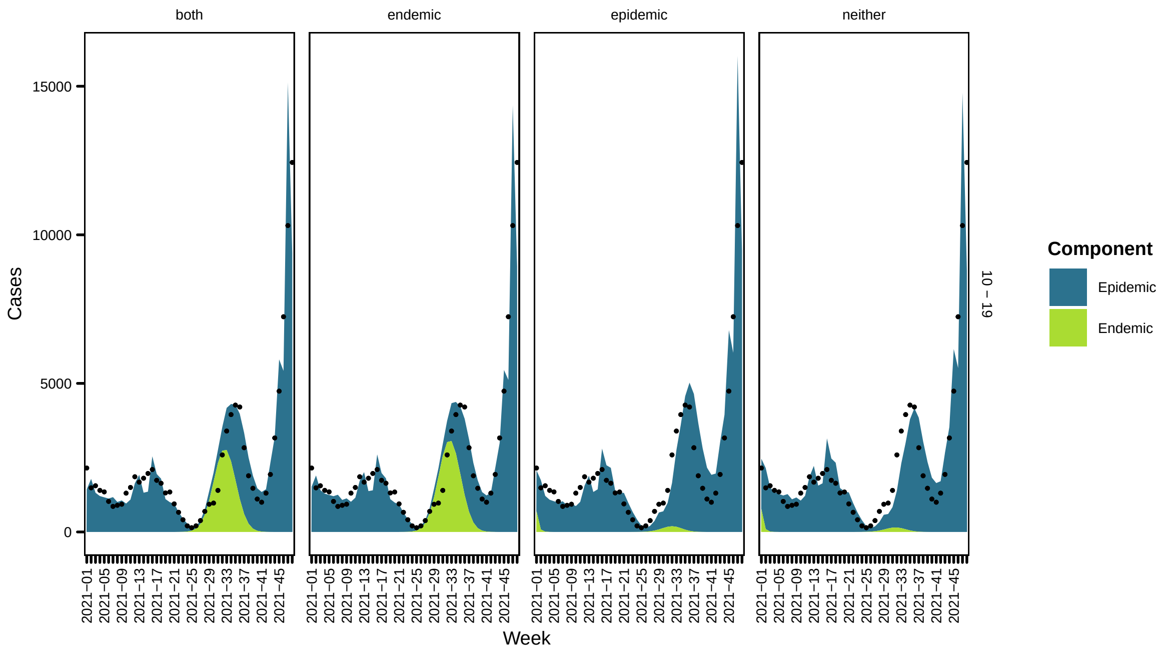
both
endemic
epidemic
neither
Cases
Week
10 – 19
Component
Epidemic
Endemic

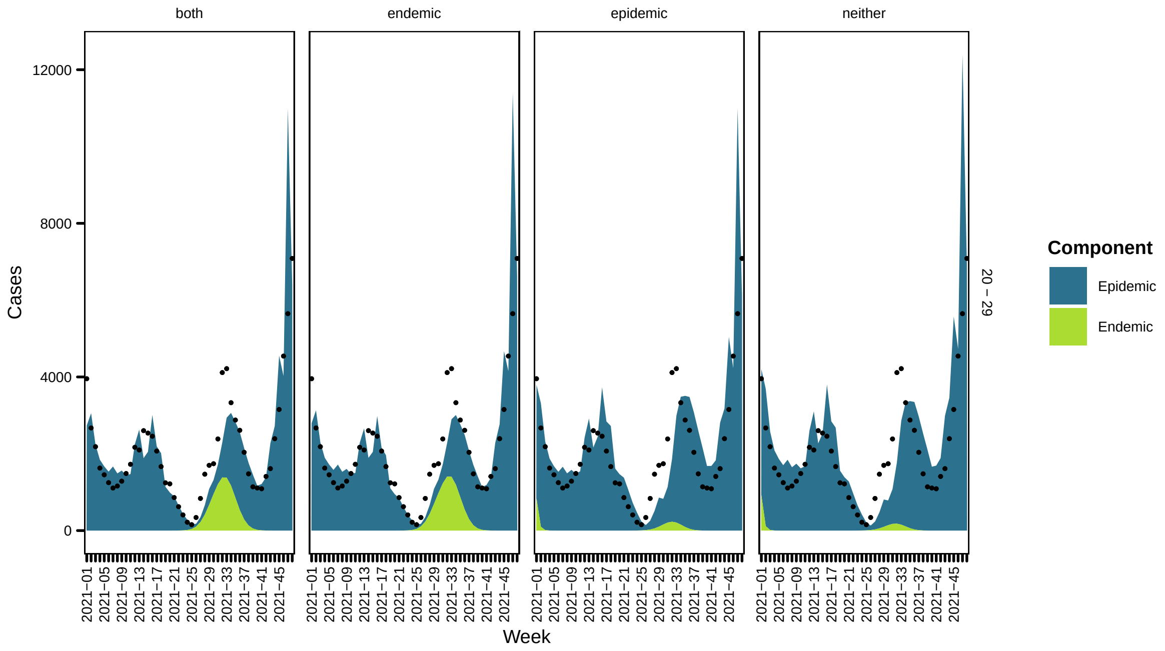
both
endemic
epidemic
neither
Cases
Week
20 – 29
Component
Epidemic
Endemic

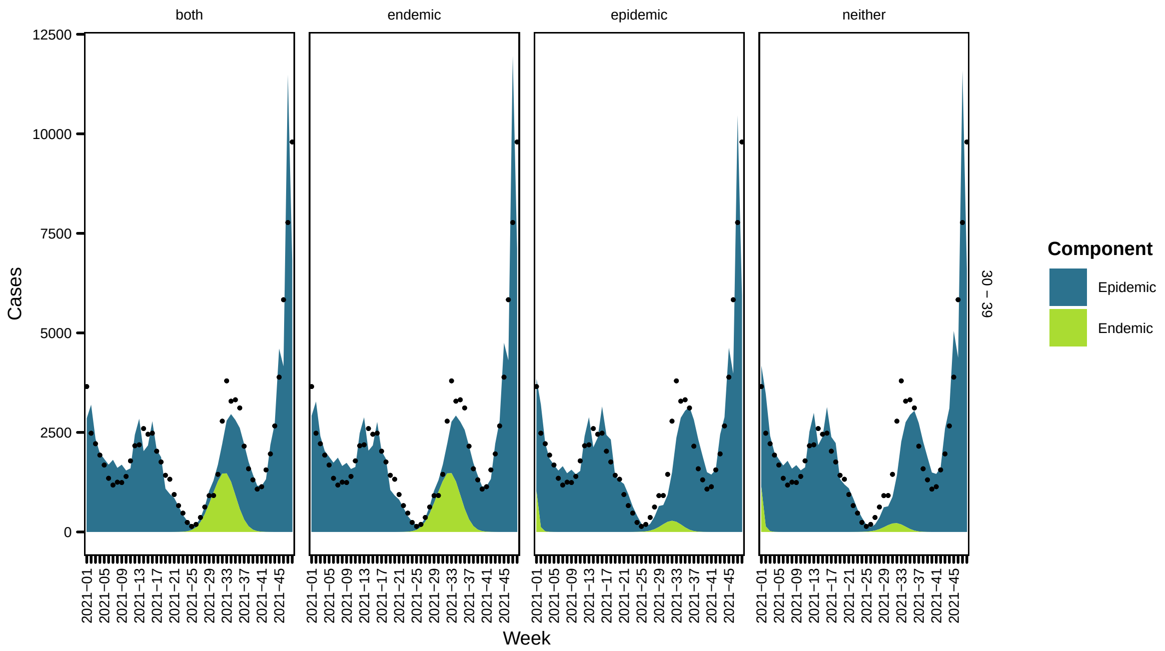
both
endemic
epidemic
neither
Cases
Week
0
2500
5000
7500
10000
12500
2021−01
2021−05
2021−09
2021−13
2021−17
2021−21
2021−25
2021−29
2021−33
2021−37
2021−41
2021−45
30 − 39
Component
Epidemic
Endemic

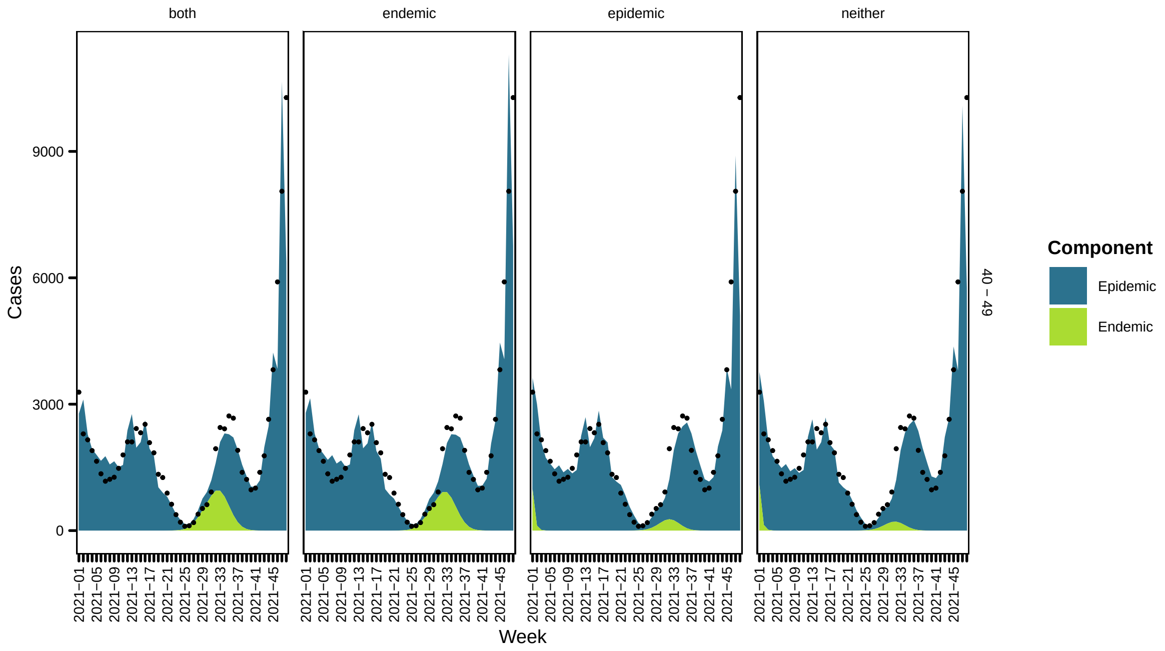
both
endemic
epidemic
neither
Cases
Week
0
3000
6000
9000
2021−01
2021−05
2021−09
2021−13
2021−17
2021−21
2021−25
2021−29
2021−33
2021−37
2021−41
2021−45
40 − 49
Component
Epidemic
Endemic

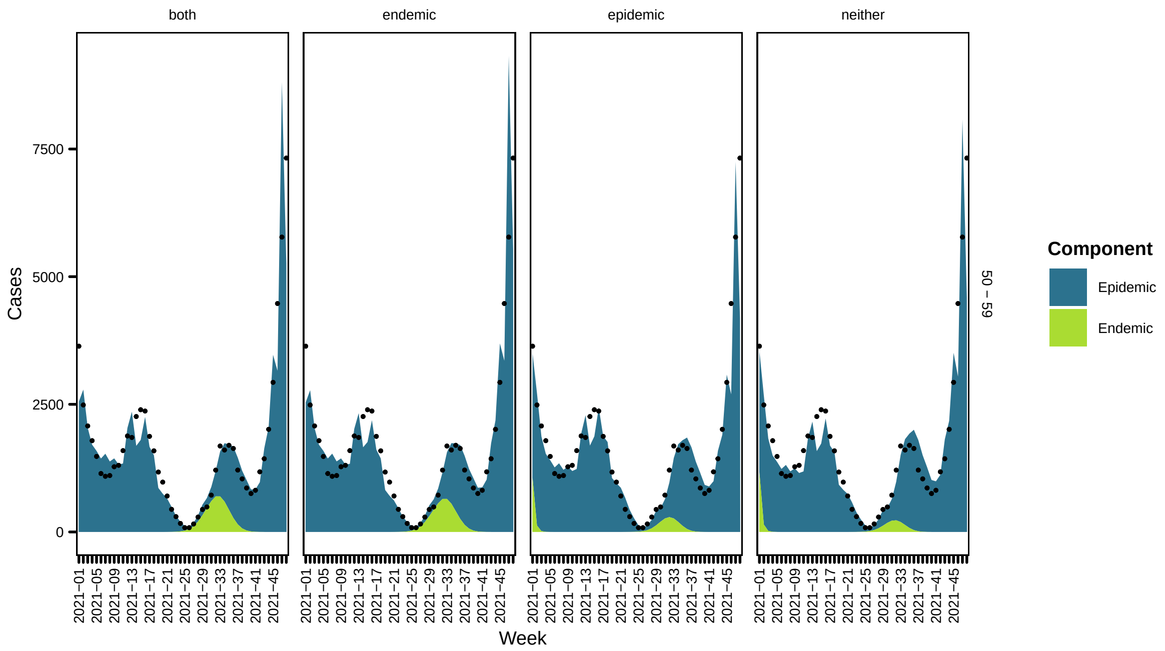

both
endemic
epidemic
neither
Cases
0
2500
5000
7500
Week
2021−01
2021−05
2021−09
2021−13
2021−17
2021−21
2021−25
2021−29
2021−33
2021−37
2021−41
2021−45
50 − 59
Component
Epidemic
Endemic


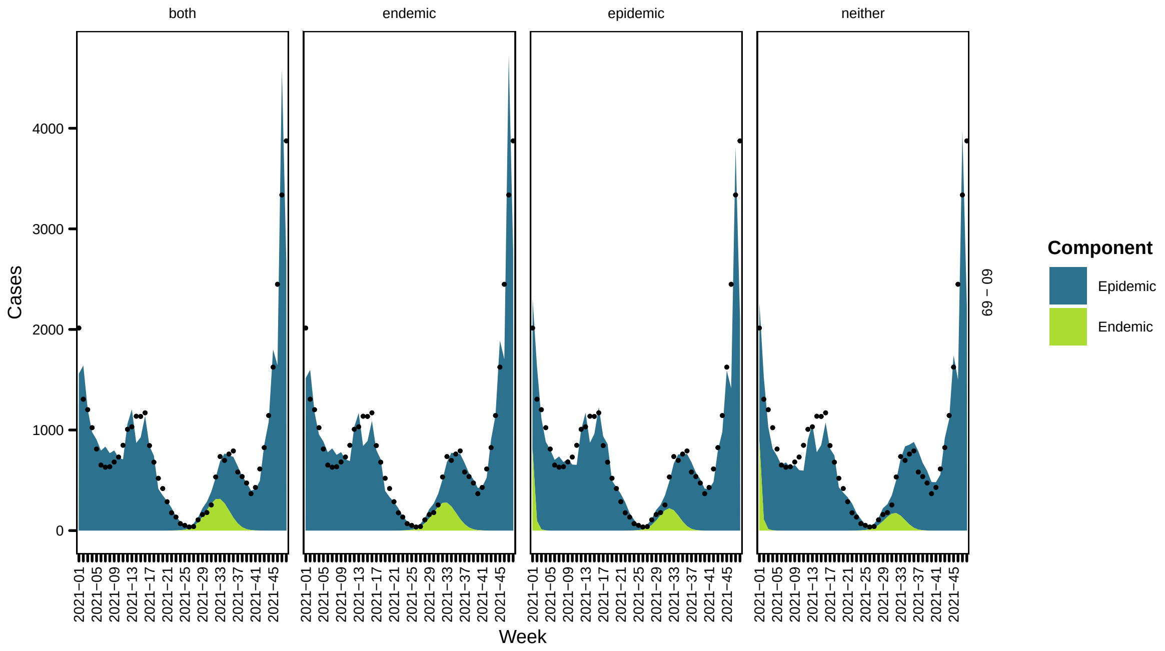

both
endemic
epidemic
neither
Cases
0
1000
2000
3000
4000
Week
2021−01
2021−05
2021−09
2021−13
2021−17
2021−21
2021−25
2021−29
2021−33
2021−37
2021−41
2021−45
60 − 69
Component
Epidemic
Endemic

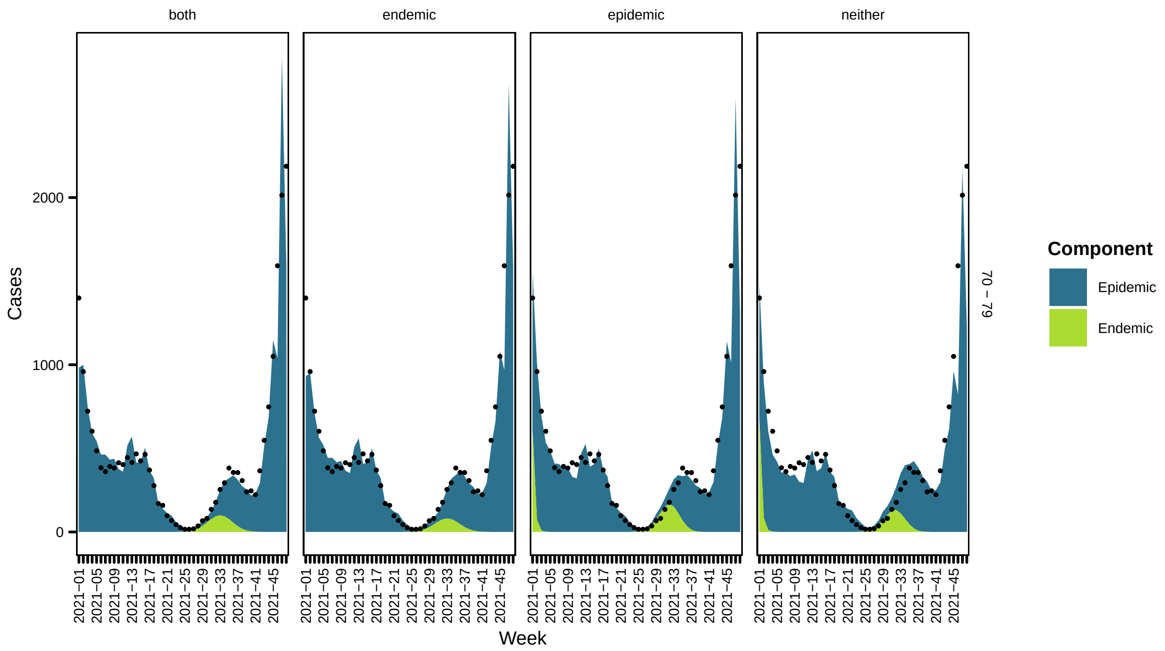


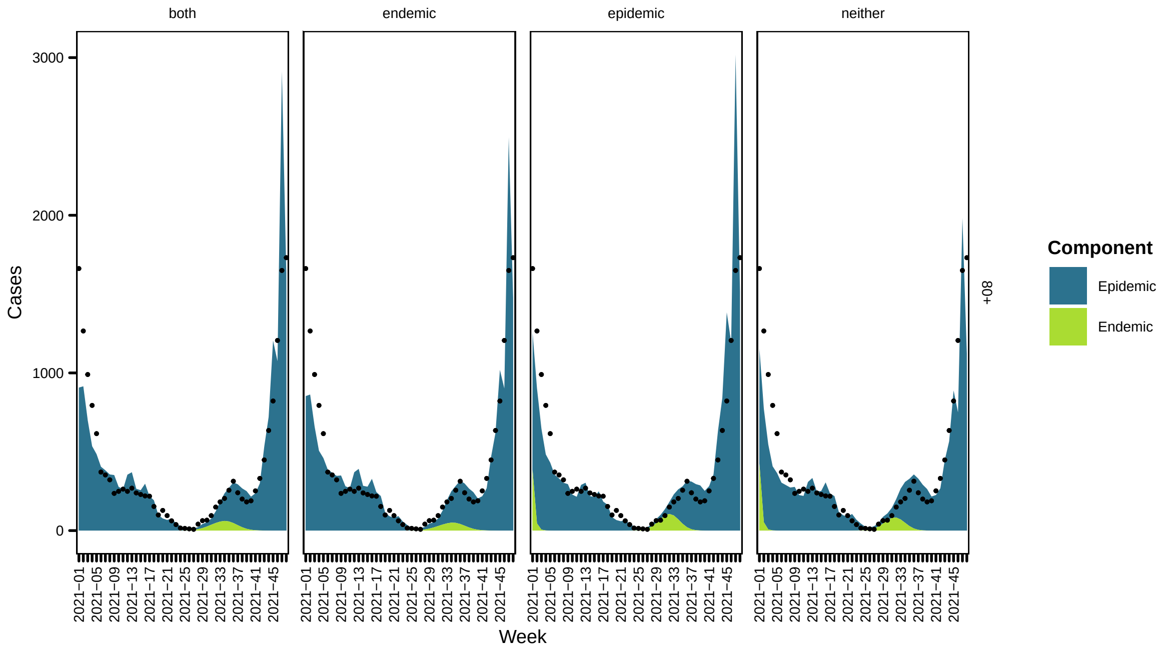


#### 3 Scenario analysis

Now we consider the effects of an alternative distribution scheme of vaccination. At each time point we count the total number of vaccines given and to evaluate the effects of not following an age-based distribution scheme, we distribute them across age groups proportional to the population size of the age group.

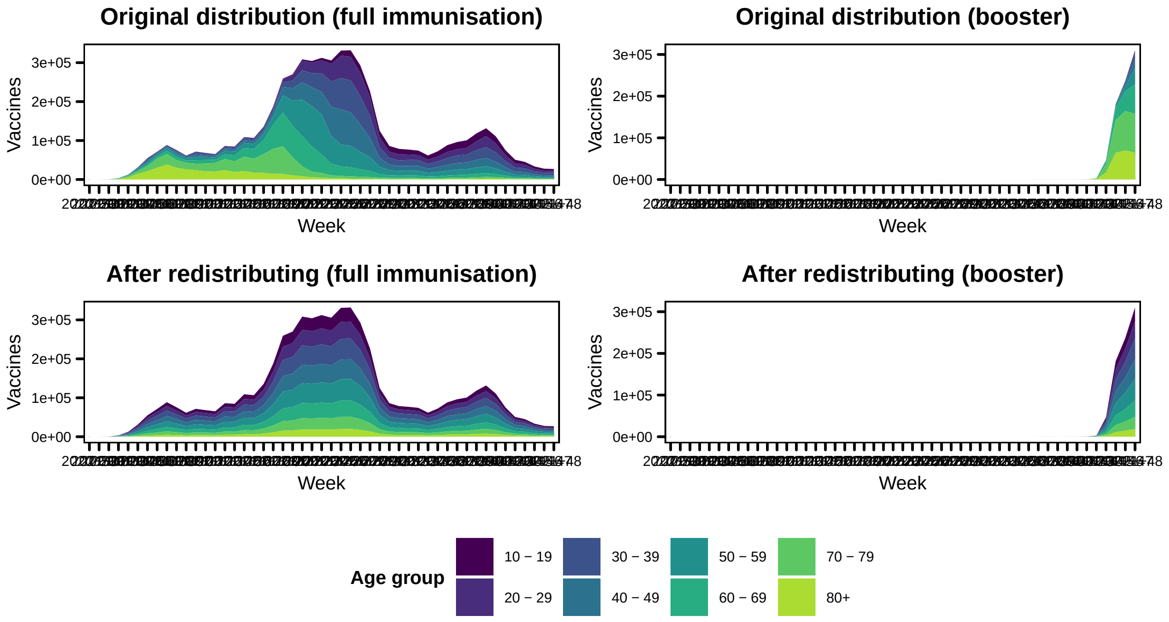


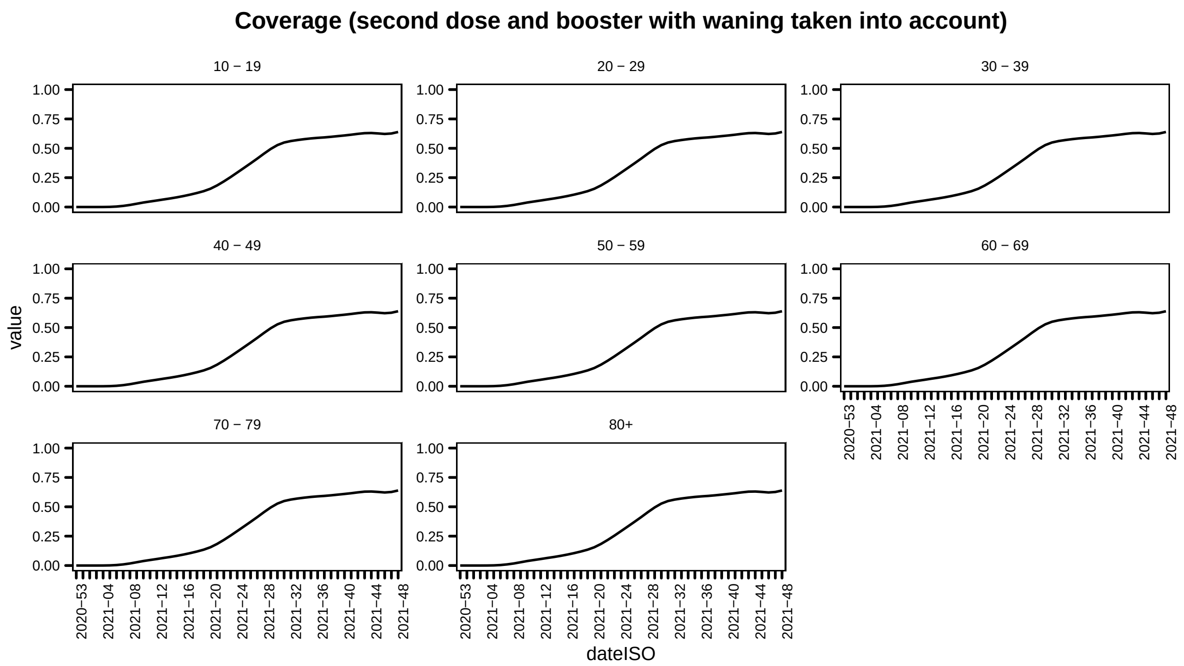

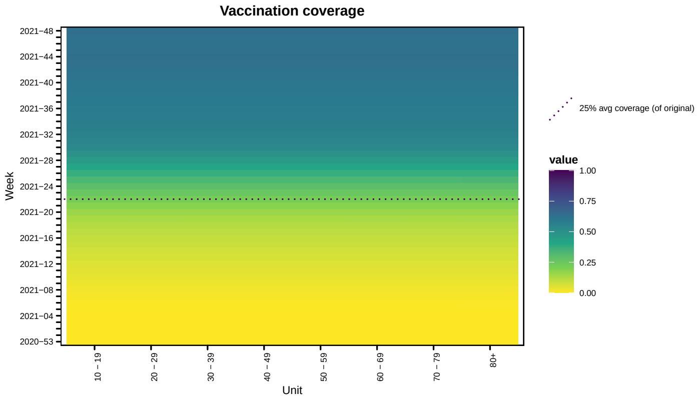


#### 3.1 Predicted cases

Under the assumption that the study period covers the outbreak, we calculate the predicted number of COVID-19 cases under the alternative vaccination coverage scenario scenario. This is using the same methods we have considered in previous work [see 2–4]. The final size of the outbreak is given (Table 1). The weekly predicted cases for the model with vaccination coverage in the endemic component with time-varying weights are presented as this was the best fitting model as measured by DSS. We compare the predictions under the true vaccination coverage (as observed by BAG) and the alternative scenario of redistributed coverage. The ratio is the ratio of predicted cases under the alternative vaccination coverage scenario against predicted cases under the observed vaccination coverage scenario.

Below we show predicted cases under the two vaccination coverages (as observed and maximum coverage across regions) and ratio between the two

**Table 1:** Final size estimates based on path forecasts

| | | Original coverage | | | Alternative coverage | | |
|---|---|---|---|---|---|---|---|
| | True | Endemic | Epidemic | Both | Endemic | Epidemic | Both |
| 10 - 19 | 73195 | 115341 | 497769 | 151064 | 54394 | 54878 | 44820 |
| 20 - 29 | 59099 | 94228 | 282376 | 105667 | 54644 | 74290 | 49647 |
| 30 - 39 | 62337 | 93760 | 265990 | 106912 | 54640 | 63686 | 49436 |
| 40 - 49 | 55949 | 84499 | 226076 | 95705 | 51097 | 57481 | 46829 |
| 50 - 59 | 40373 | 68097 | 171177 | 75906 | 44217 | 49229 | 41602 |
| 60 - 69 | 20807 | 33111 | 78320 | 36738 | 24494 | 26597 | 24109 |
| 70 - 79 | 11779 | 17250 | 49350 | 20853 | 14677 | 15245 | 15032 |
| 80+ | 9342 | 15400 | 59054 | 21075 | 12319 | 13778 | 12827 |

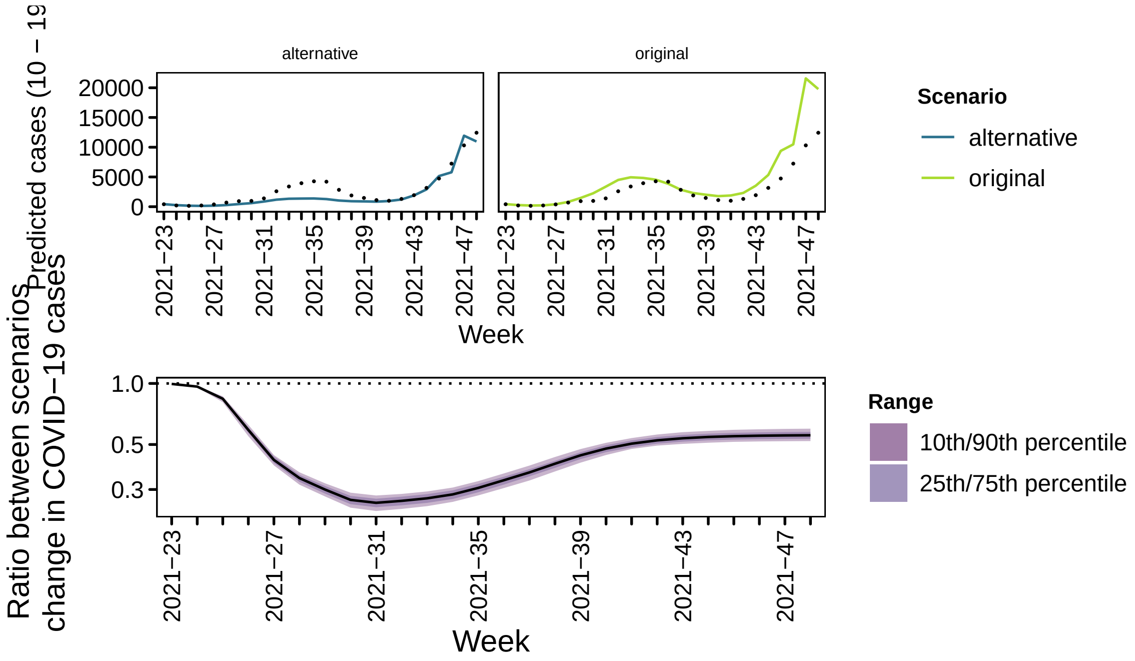


82

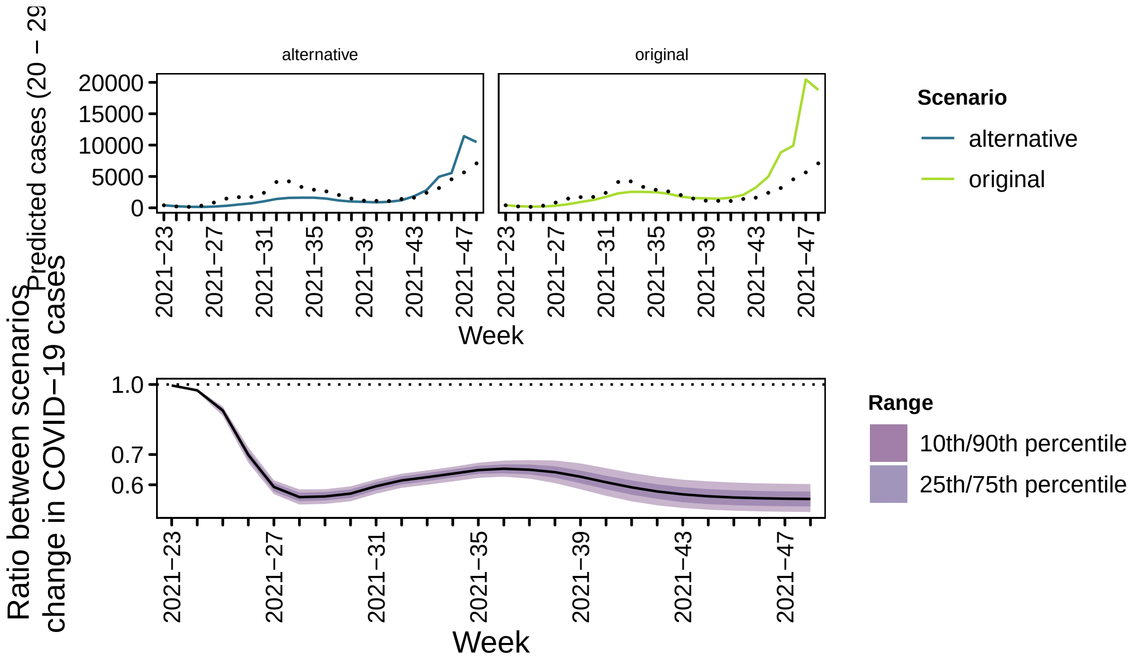
Predicted cases (20 – 29
alternative
original
20000
15000
10000
5000
0
2021−23
2021−27
2021−31
2021−35
2021−39
2021−43
2021−47
Week
Scenario
alternative
original
Ratio between scenarios change in COVID−19 cases
1.0
0.7
0.6
Range
10th/90th percentile
25th/75th percentile

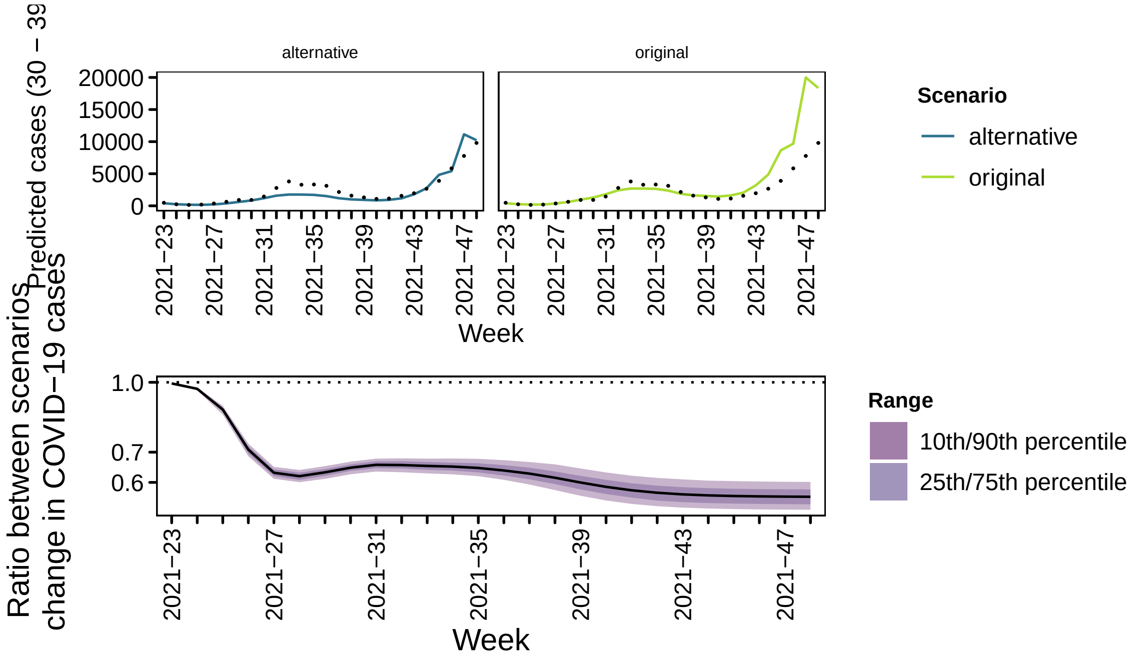
Predicted cases (30 – 39
alternative
original
20000
15000
10000
5000
0
2021−23
2021−27
2021−31
2021−35
2021−39
2021−43
2021−47
Week
Scenario
alternative
original
Ratio between scenarios change in COVID−19 cases
1.0
0.7
0.6
Range
10th/90th percentile
25th/75th percentile

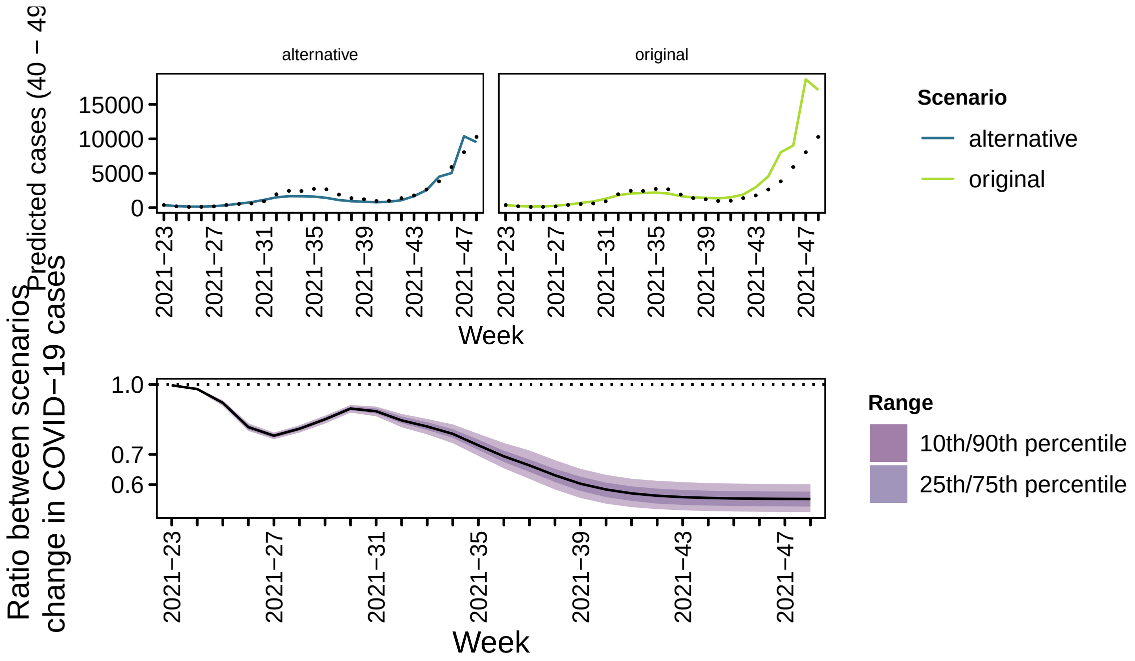
alternative
original
Predicted cases (40 – 49
Ratio between scenarios
change in COVID−19 cases
Week
Scenario
alternative
original
Range
10th/90th percentile
25th/75th percentile

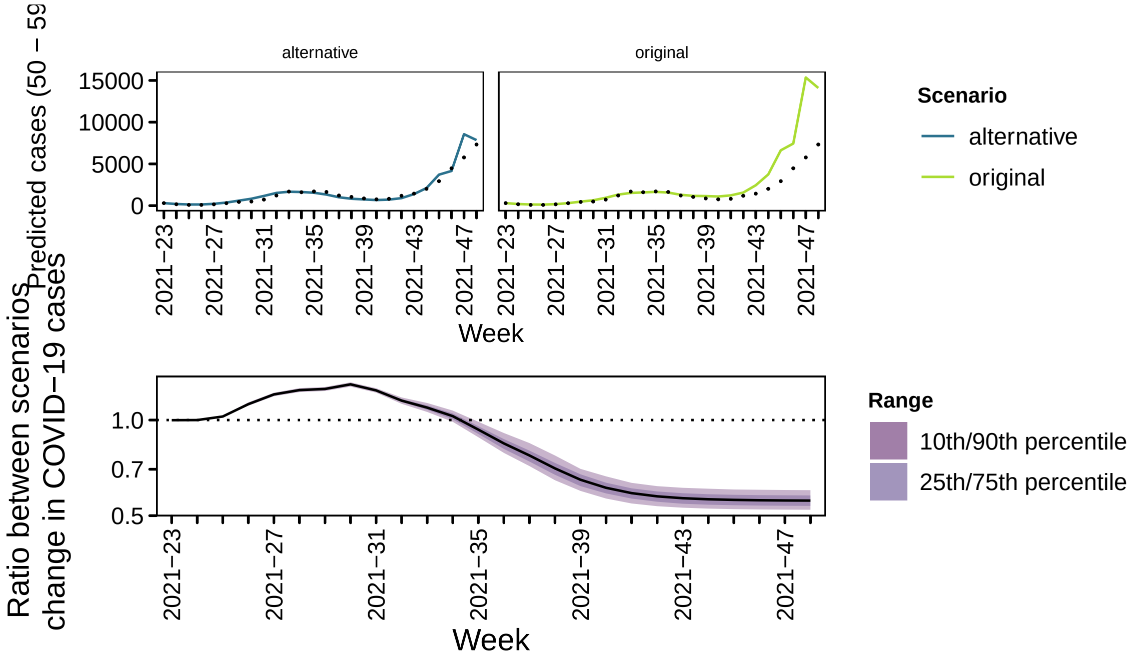
alternative
original
Predicted cases (50 – 59
Ratio between scenarios
change in COVID−19 cases
Week
Scenario
alternative
original
Range
10th/90th percentile
25th/75th percentile

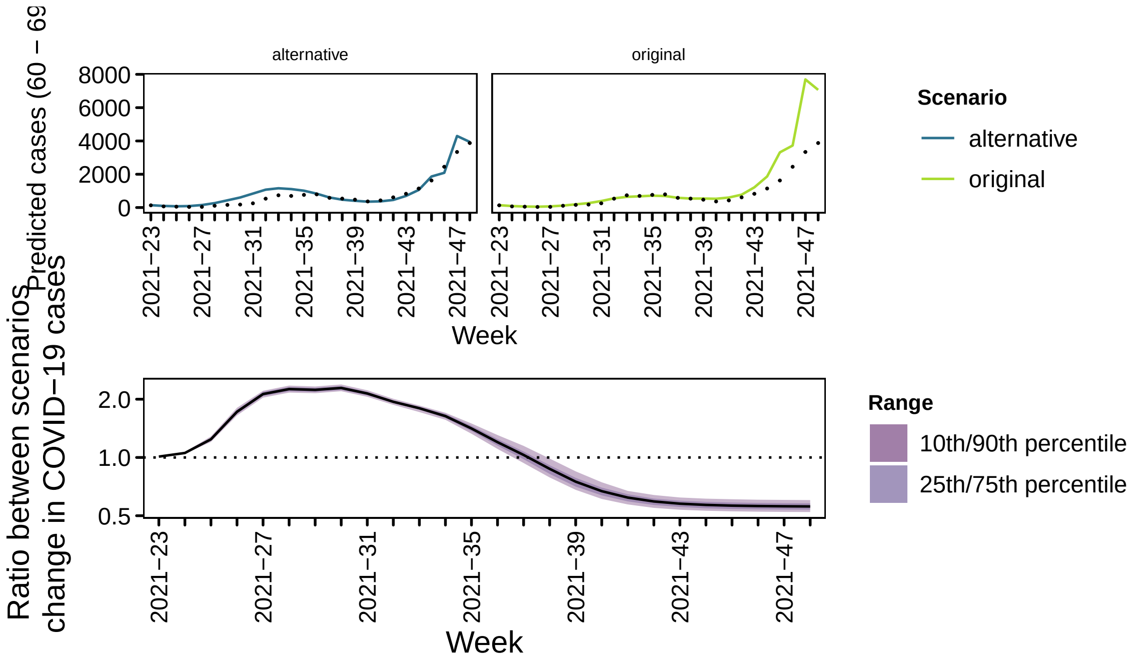
alternative
original
Predicted cases (60 − 69
Week
Scenario
alternative
original
Ratio between scenarios
change in COVID−19 cases
Week
Range
10th/90th percentile
25th/75th percentile

87

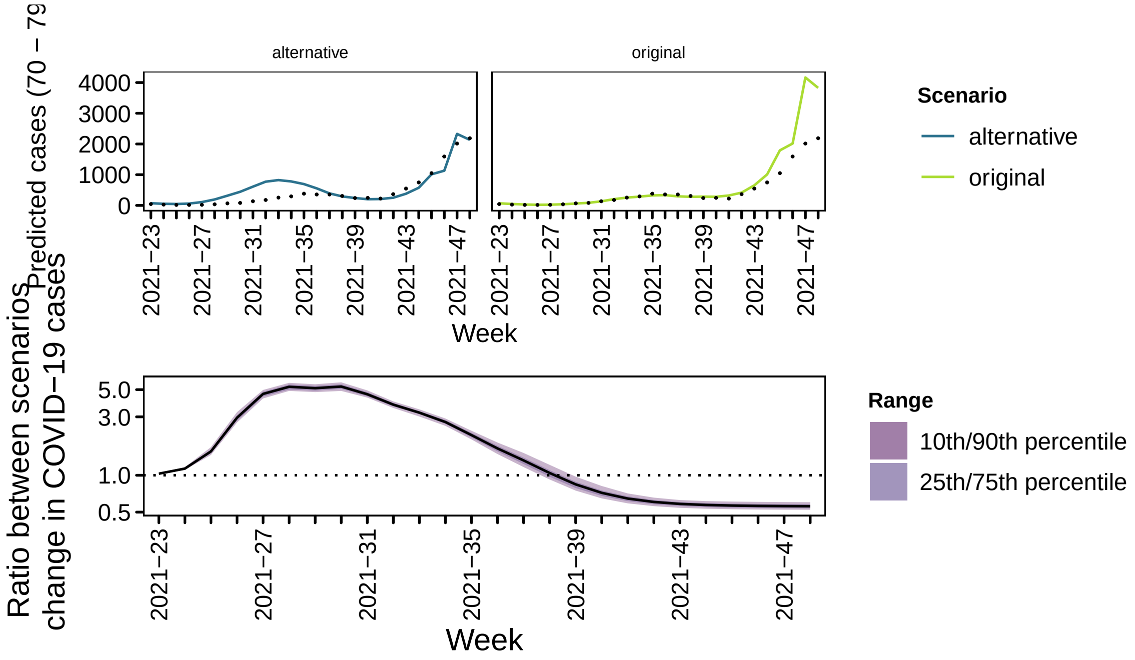
alternative
original
Predicted cases (70 − 79
Week
Scenario
alternative
original
Ratio between scenarios
change in COVID−19 cases
Week
Range
10th/90th percentile
25th/75th percentile

88

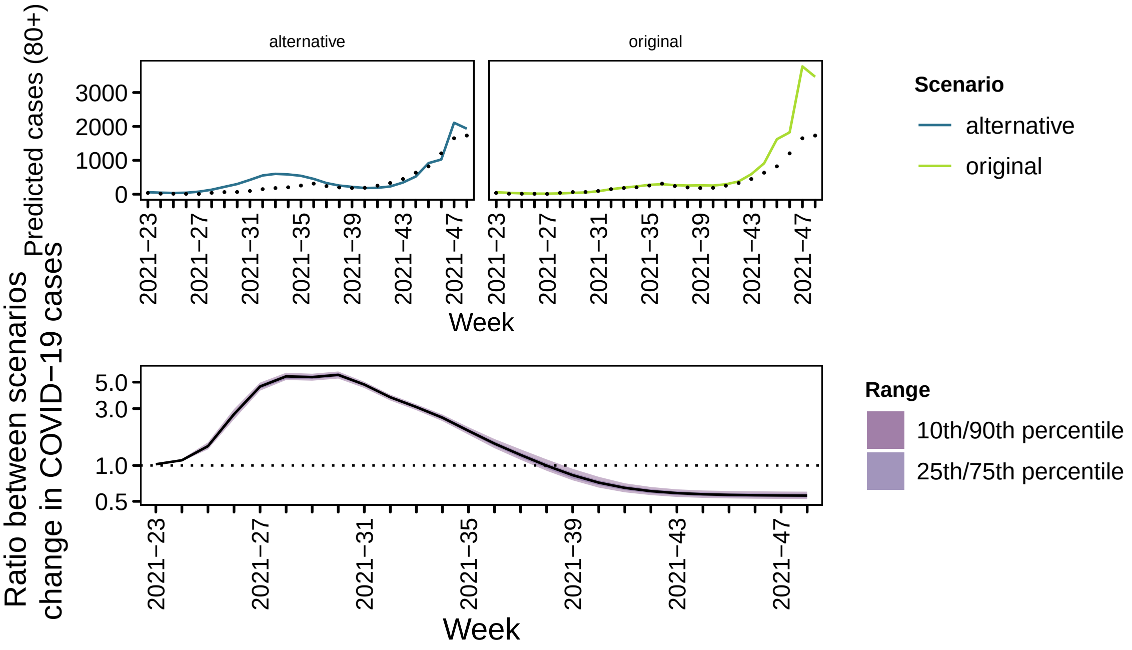


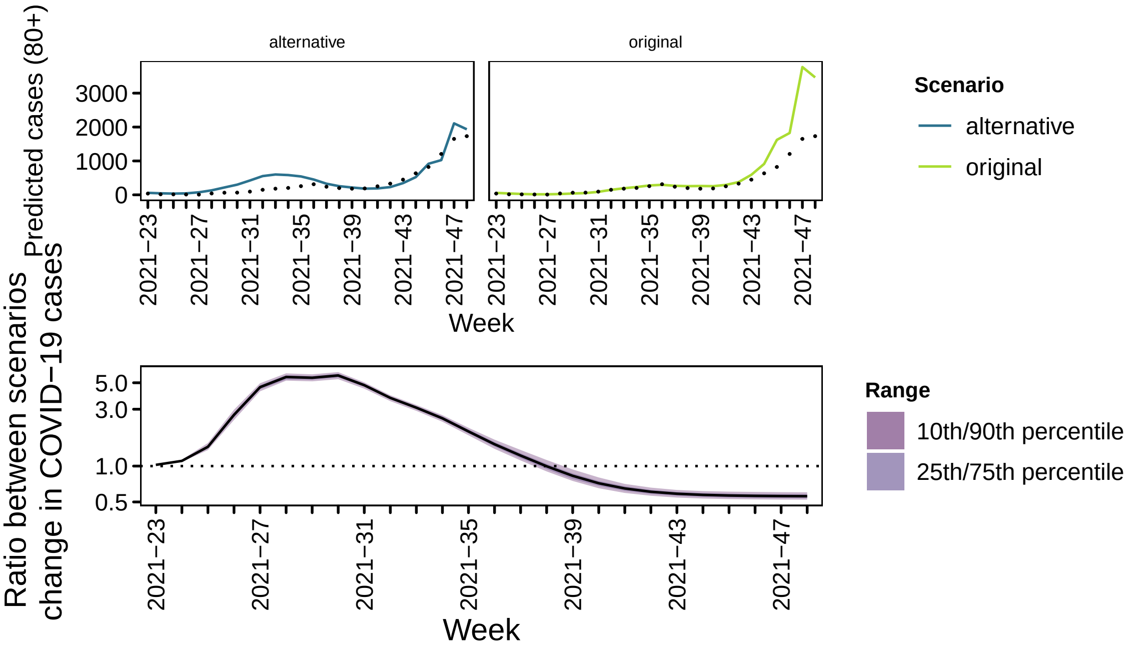
