## Supporting information (models-regions) for "The COVID-19 vaccination campaign in Switzerland and its impact on disease spread"

### MODEL – regions only $Y_{rt}$, transmission weights $\mathrm{w} = (w_{r,r',t})$

M. Bekker-Nielsen Dunbar and L. Held
Epidemiology, Biostatistics and Prevention Institute, University of Zurich

This document contains additional models to those outlined in the study protocol [1]. Knowledge of the analyses from the main manuscript is assumed. The models introduced here have only regional effects. This means we are comparing the models:

$$\begin{aligned}\log(\nu_{rt}) &= \alpha_r^{(\nu)} + \qquad\qquad + \gamma^{(\nu)\top}\mathbf{z}_{rt}^{(\nu)} \\ \log(\phi_{rt}) &= \alpha_r^{(\phi)} + \qquad\qquad + \gamma^{(\phi)\top}\mathbf{z}_{rt}^{(\phi)}\end{aligned} \qquad \text{(neither)}$$

$$\begin{aligned}\log(\nu_{rt}) &= \alpha_r^{(\nu)} + \beta_r^{(\nu)}\log(1 - x_{rt}) + \gamma^{(\nu)\top}\mathbf{z}_{rt}^{(\nu)} \\ \log(\phi_{rt}) &= \alpha_r^{(\phi)} + \qquad\qquad + \gamma^{(\phi)\top}\mathbf{z}_{rt}^{(\phi)}\end{aligned} \qquad \text{(endemic)}$$

$$\begin{aligned}\log(\nu_{rt}) &= \alpha_r^{(\nu)} + \qquad\qquad + \gamma^{(\nu)\top}\mathbf{z}_{rt}^{(\nu)} \\ \log(\phi_{rt}) &= \alpha_r^{(\phi)} + \beta_r^{(\phi)}\log(1 - x_{rt}) + \gamma^{(\phi)\top}\mathbf{z}_{rt}^{(\phi)}\end{aligned} \qquad \text{(epidemic)}$$

$$\begin{aligned}\log(\nu_{rt}) &= \alpha_r^{(\nu)} + \beta_r^{(\nu)}\log(1 - x_{rt}) + \gamma^{(\nu)\top}\mathbf{z}_{rt}^{(\nu)} \\ \log(\phi_{rt}) &= \alpha_r^{(\phi)} + \beta_r^{(\phi)}\log(1 - x_{rt}) + \gamma^{(\phi)\top}\mathbf{z}_{rt}^{(\phi)}\end{aligned} \qquad \text{(both)}$$

$\mathbf{z}_{rt}$ is as given in the main manuscript.

### 1 Setup

The study period is 1st January 2021 to 30th November 2021 given by ISO weeks 2020-53 to 2021-48, both weeks included. This is a deviation from the protocol. The new study period is chosen on the basis of the epidemic calendar. This is similar to influenza research in the northern hemisphere; we want our years to start and end such that the entire expected wave is captured in a single year. Furthermore, the latter cut-off was chosen due to the presence of Omicron variant COVID-19 in the original study period. The reason for this is that the vaccines considered in this work were not created with this variant in mind and may provide less protection against this disease. The study period covers 49 observations.

#### 1.1 Outcome

The outcome variable is cases. Here we consider all cases (i.e. not limited to hospitalisations). The cases are not equally distributed across cantons:

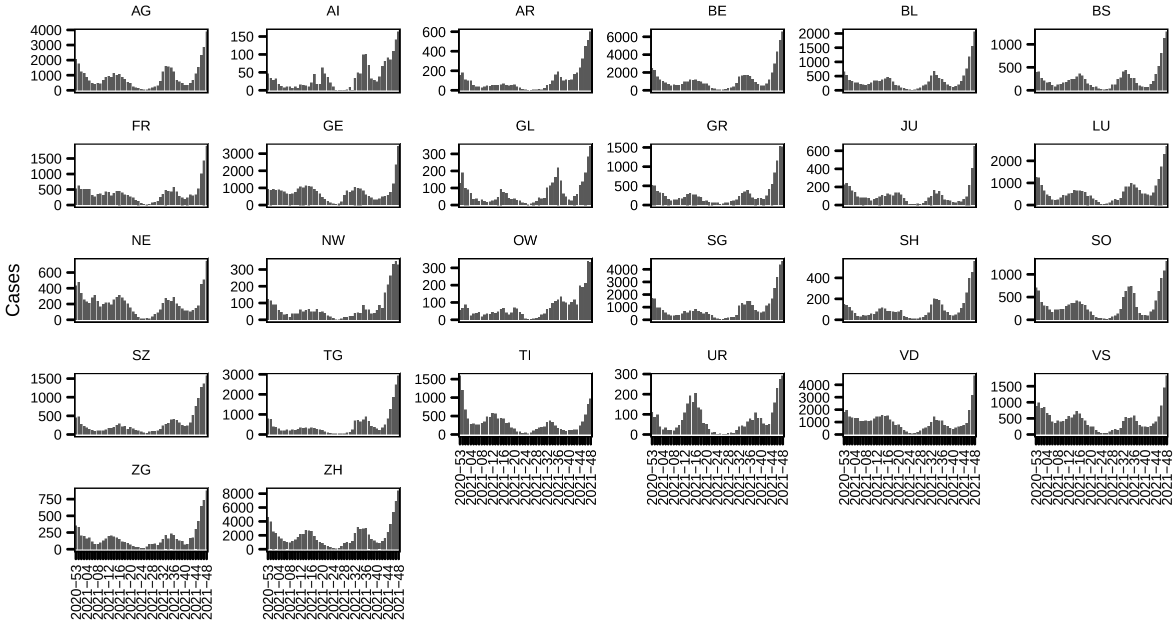

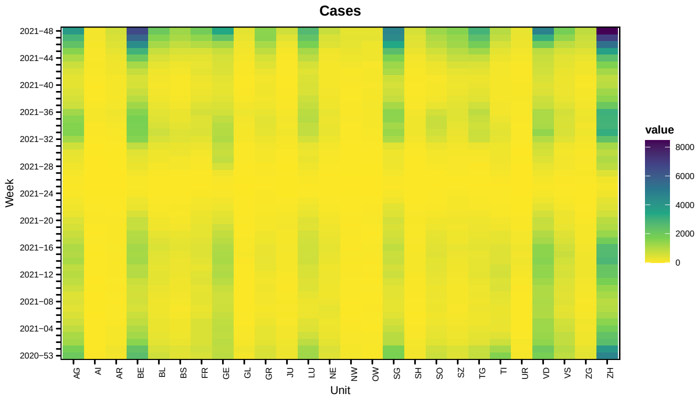


Taking into account that different cantons have different populations, differences in burden of disease are still evident

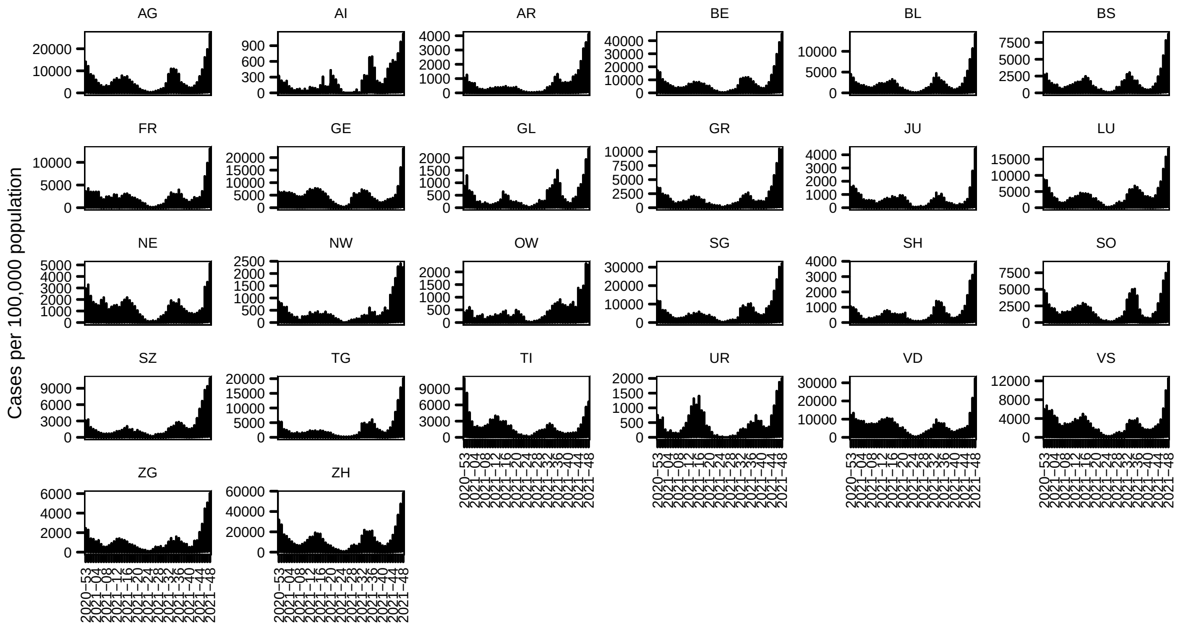


#### 1.2 Covariates

The vaccination coverage looks as follows

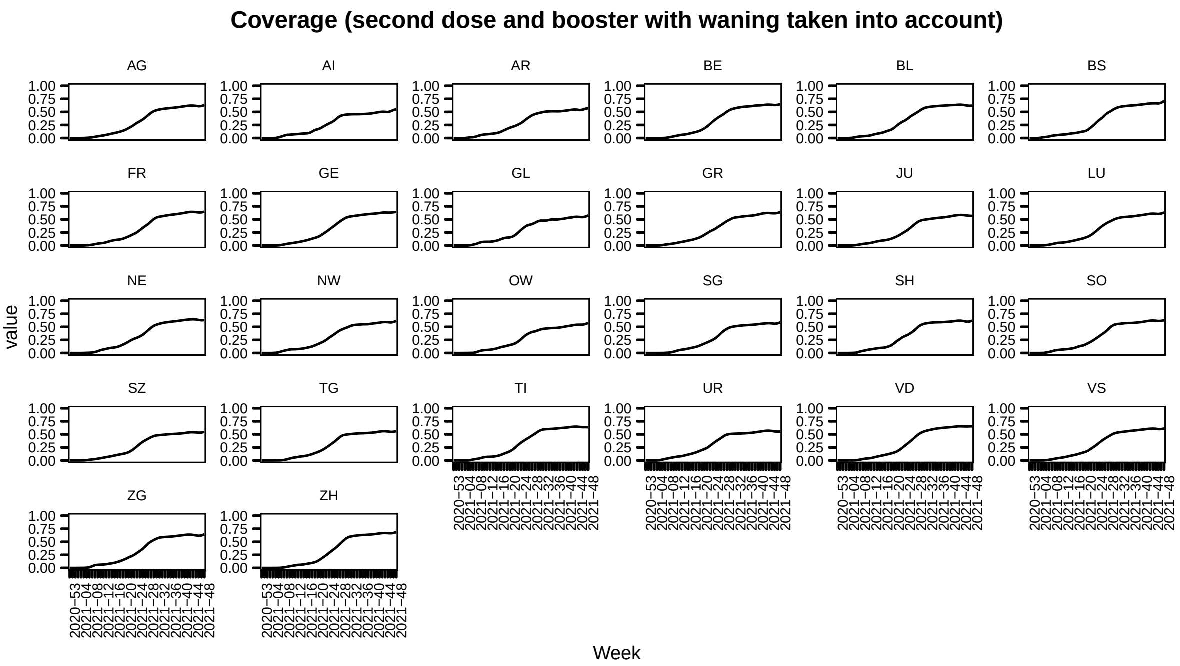
Coverage (second dose and booster with waning taken into account)
AG
AI
AR
BE
BL
BS
FR
GE
GL
GR
JU
LU
NE
NW
OW
SG
SH
SO
SZ
TG
TI
UR
VD
VS
ZG
ZH
value
1.00
0.75
0.50
0.25
0.00
2020−53
2021−04
2021−08
2021−12
2021−16
2021−20
2021−24
2021−28
2021−32
2021−36
2021−40
2021−44
2021−48
Week

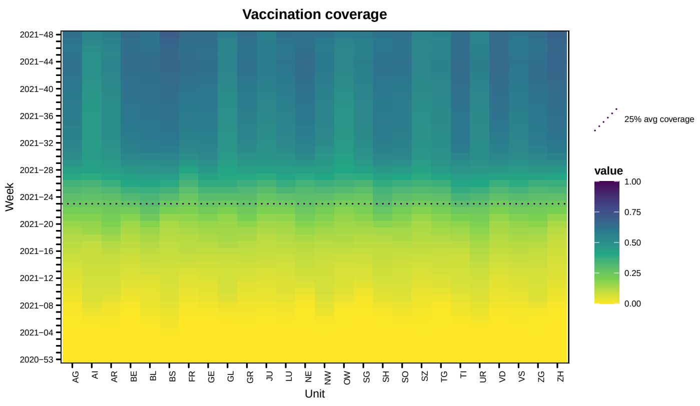
Vaccination coverage
Week
2021−48
2021−44
2021−40
2021−36
2021−32
2021−28
2021−24
2021−20
2021−16
2021−12
2021−08
2021−04
2020−53
AG
AI
AR
BE
BL
BS
FR
GE
GL
GR
JU
LU
NE
NW
OW
SG
SH
SO
SZ
TG
TI
UR
VD
VS
ZG
ZH
Unit
25% avg coverage
value
1.00
0.75
0.50
0.25
0.00

Transformed by $f(x) = \log(1 - x)$ this looks like

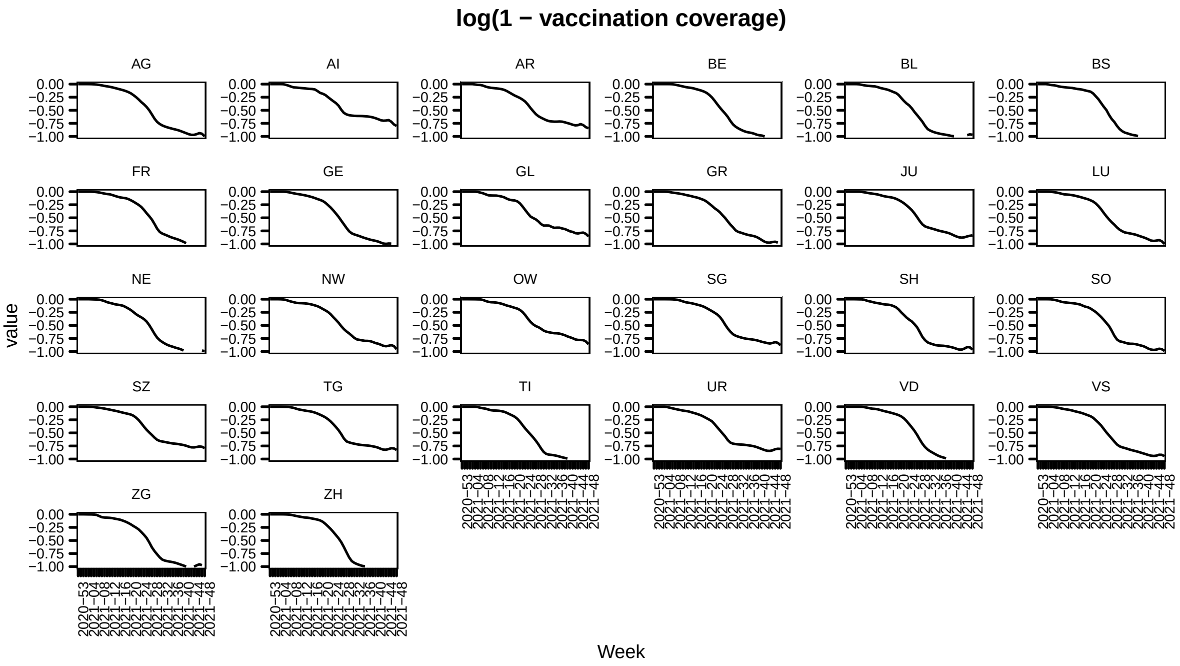


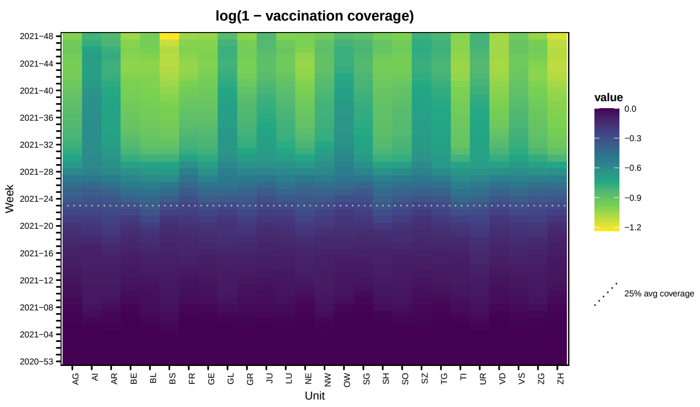


#### 2 Results overview

The following table provides the estimated effects of vaccination coverage for models with and without time-varying transmission weights.

```
## ! Non-convergence message from optimizer: iteration limit reached without converge
## Update of regression coefficients in iteration  1  unreliable
## ! Non-convergence message from optimizer: iteration limit reached without converge
## Update of regression coefficients in iteration  1  unreliable
```

| Weights | Model | Endemic | Epidemic | $\ell$ | DSS |
|---|---|---|---|---|---|
| Constant | Neither | | | -7198.294 | 15.253 |
| Constant | Endemic | 4.401 (SE 1.343) | | -7190.34 | 15.337 |
| Constant | Epidemic | | 3.251 (SE 0.216) | -7096.772 | 15.236 |
| Constant | Both | 1.677 (SE 1.038) | 3.206 (SE 0.216) | -7093.356 | 15.241 |
| Varying | Neither | | | -7091.831 | 13.787 |
| Varying | Endemic | 3.312 (SE 1.035) | | -7084.242 | 13.742 |
| Varying | Epidemic | | 2.688 (SE 0.201) | -7010.947 | 17.084 |
| Varying | Both | 1.739 (SE 0.846) | 2.642 (SE 0.2) | -7006.704 | 16.988 |

#### 2.1 Time-constant regional transmission weights

First we present the results of the modelling with the time-constant transmission weights.

| | neither | endemic | epidemic | both |
|---|---|---|---|---|
| $\gamma^{\phi}_{\sin(2\pi t/52)}$ | 0.405 (SE 0.043) | 0.43 (SE 0.043) | -0.132 (SE 0.053) | -0.123 (SE 0.053) |
| $\gamma^{\phi}_{\cos(2\pi t/52)}$ | 0.335 (SE 0.025) | 0.352 (SE 0.025) | 0.45 (SE 0.026) | 0.451 (SE 0.026) |
| $\gamma^{\phi}_{\text{gravity}}$ | 0.915 (SE 0.043) | 0.918 (SE 0.043) | 0.984 (SE 0.036) | 0.984 (SE 0.036) |
| $\gamma^{\phi}_{\text{time}}$ | 0.029 (SE 0.002) | 0.03 (SE 0.002) | 0.087 (SE 0.004) | 0.086 (SE 0.004) |
| $\alpha^{\phi}_{\text{region}}$ | -1.568 (SE 0.17) | -1.57 (SE 0.171) | 0.03 (SE 0.176) | 0.01 (SE 0.177) |
| $\gamma^{\nu}_{\sin(2\pi t/52)}$ | -9.468 (SE 0.69) | -11.773 (SE 1.015) | -9.282 (SE 0.487) | -10.29 (SE 0.795) |
| $\gamma^{\nu}_{\cos(2\pi t/52)}$ | -3.993 (SE 0.499) | -4.646 (SE 0.534) | -3.623 (SE 0.317) | -3.991 (SE 0.393) |
| $\gamma^{\nu}_{\text{time}}$ | -0.467 (SE 0.045) | -0.446 (SE 0.042) | -0.424 (SE 0.03) | -0.426 (SE 0.03) |
| $\alpha^{\nu}_{\text{region}}$ | 2.932 (SE 0.492) | 4.259 (SE 0.593) | 3.36 (SE 0.339) | 3.751 (SE 0.423) |
| $\psi$ | 0.141 (SE 0.006) | 0.138 (SE 0.006) | 0.119 (SE 0.005) | 0.118 (SE 0.005) |
| $\beta^{(\nu)}$ | | 4.401 (SE 1.343) | | 1.677 (SE 1.038) |
| $\beta^{(\phi)}$ | | | 3.251 (SE 0.216) | 3.206 (SE 0.216) |

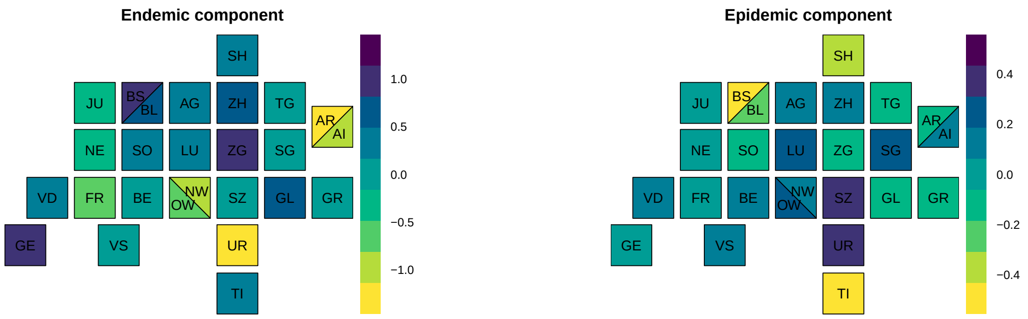
Endemic component
SH
JU
BS
BL
AG
ZH
TG
AR
AI
NE
SO
LU
ZG
SG
VD
FR
BE
NW
OW
SZ
GL
GR
GE
VS
UR
TI
1.0
0.5
0.0
−0.5
−1.0
Epidemic component
0.4
0.2
0.0
−0.2
−0.4

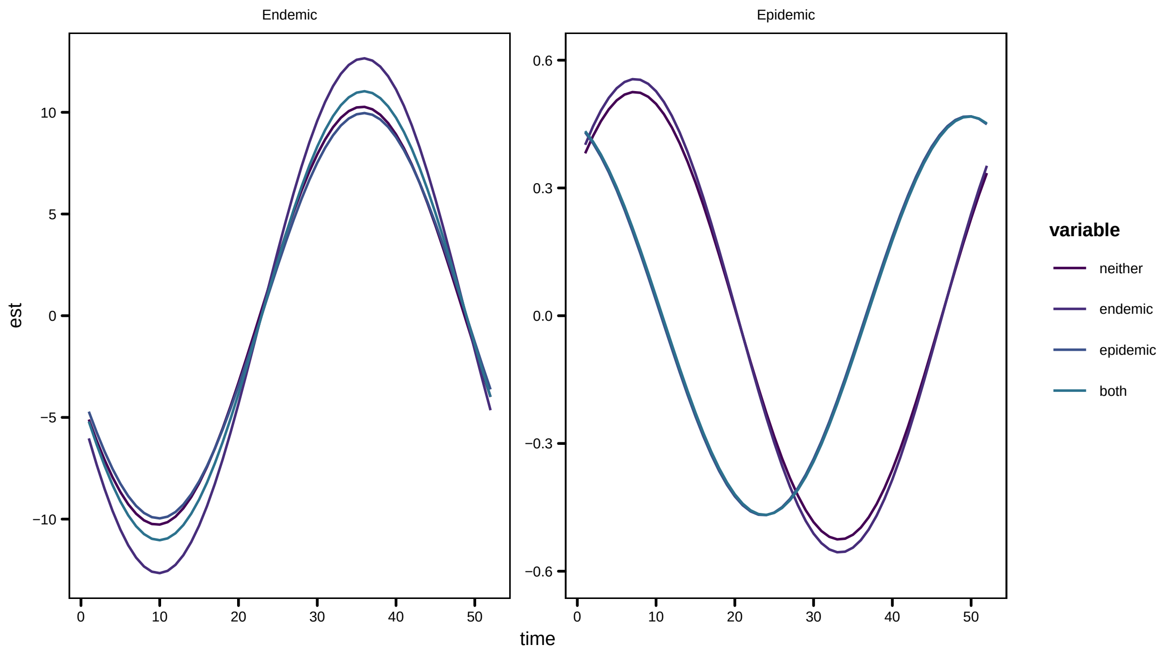
Endemic
Epidemic
est
time
variable
neither
endemic
epidemic
both
10
5
0
−5
−10
0.6
0.3
0.0
−0.3
−0.6
0
10
20
30
40
50

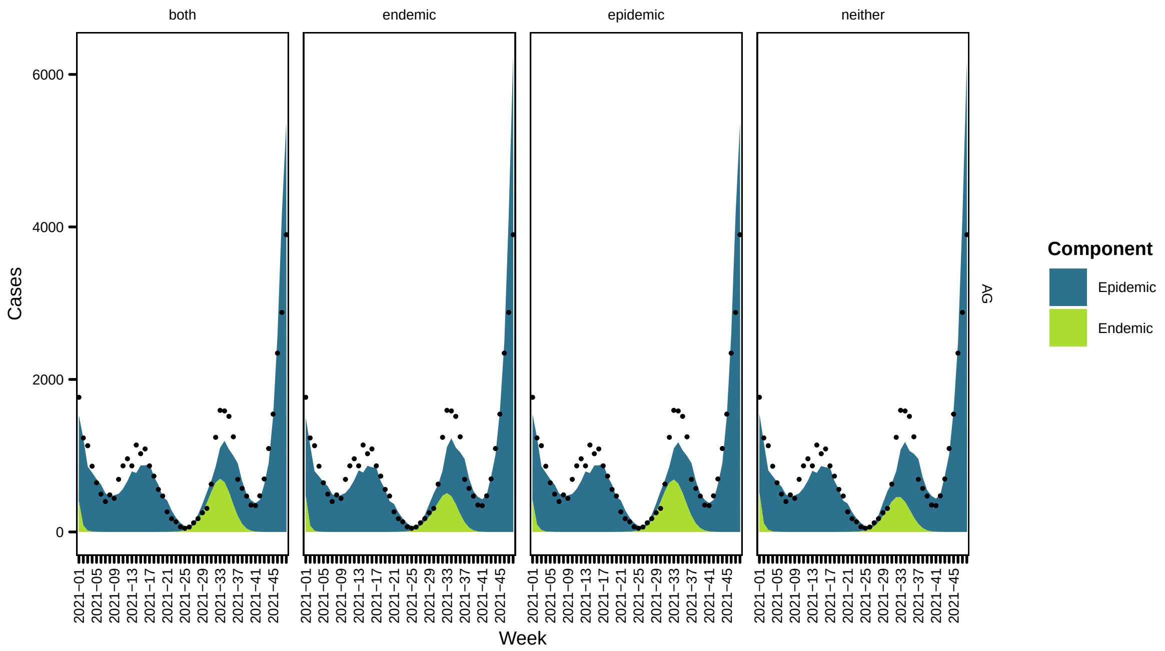
both
endemic
epidemic
neither
Cases
6000
4000
2000
0
2021−01
2021−05
2021−09
2021−13
2021−17
2021−21
2021−25
2021−29
2021−33
2021−37
2021−41
2021−45
Week
AG
Component
Epidemic
Endemic

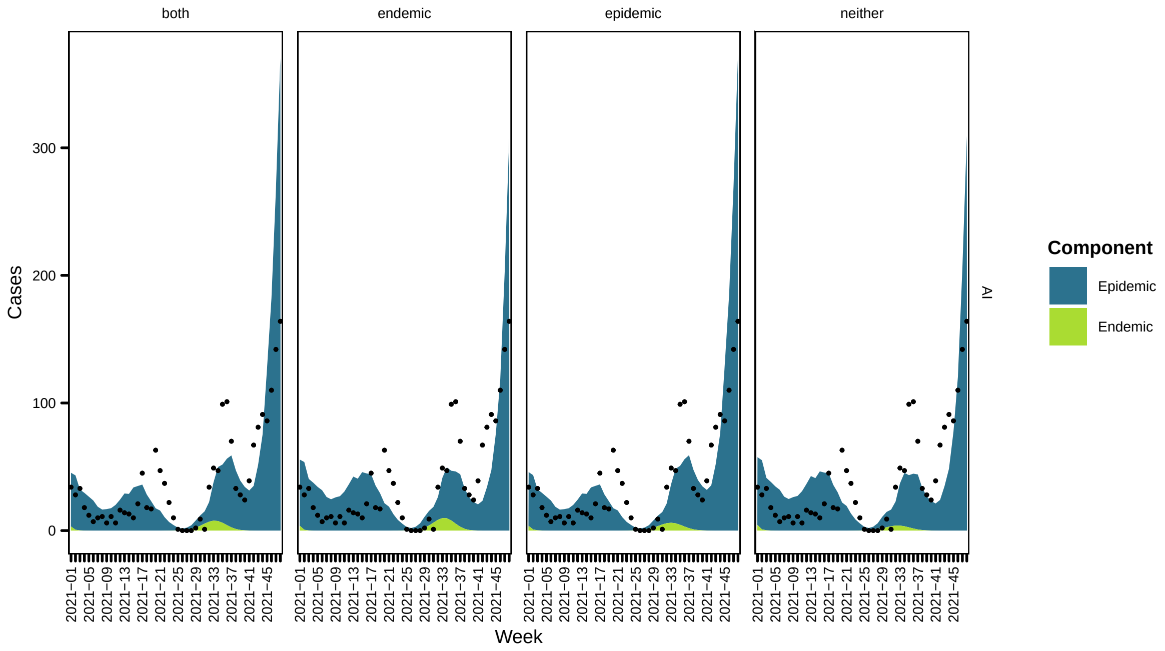
both
endemic
epidemic
neither
Cases
300
200
100
0
2021−01
2021−05
2021−09
2021−13
2021−17
2021−21
2021−25
2021−29
2021−33
2021−37
2021−41
2021−45
Week
AI
Component
Epidemic
Endemic

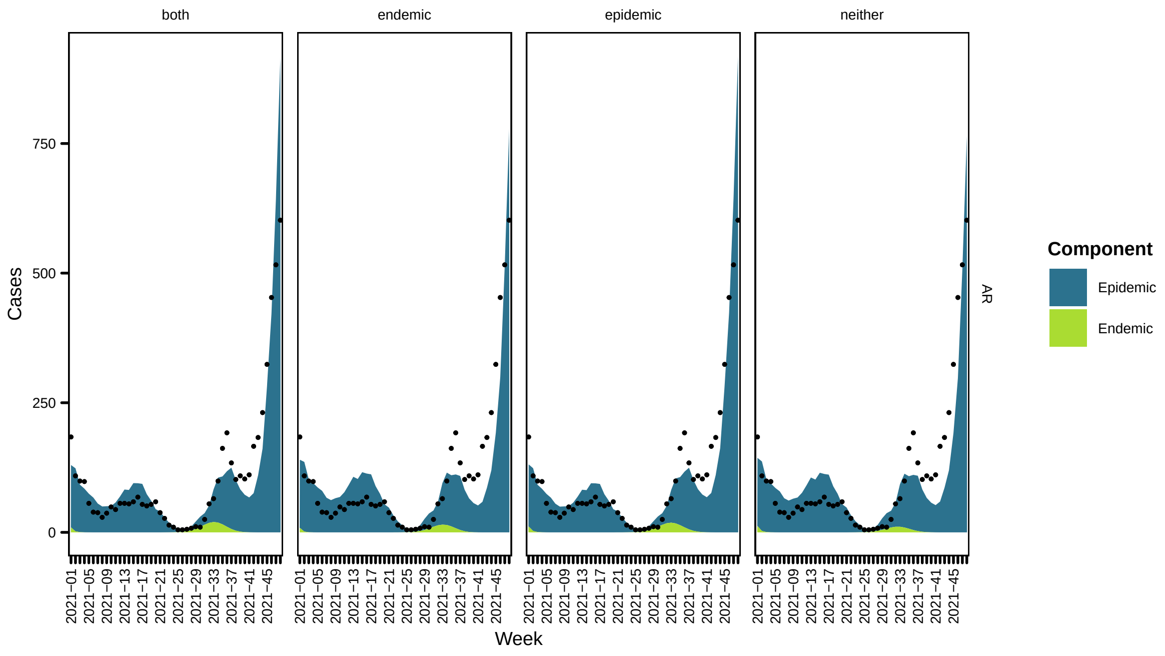
both
endemic
epidemic
neither
Cases
0
250
500
750
2021−01
2021−05
2021−09
2021−13
2021−17
2021−21
2021−25
2021−29
2021−33
2021−37
2021−41
2021−45
Week
AR
Component
Epidemic
Endemic

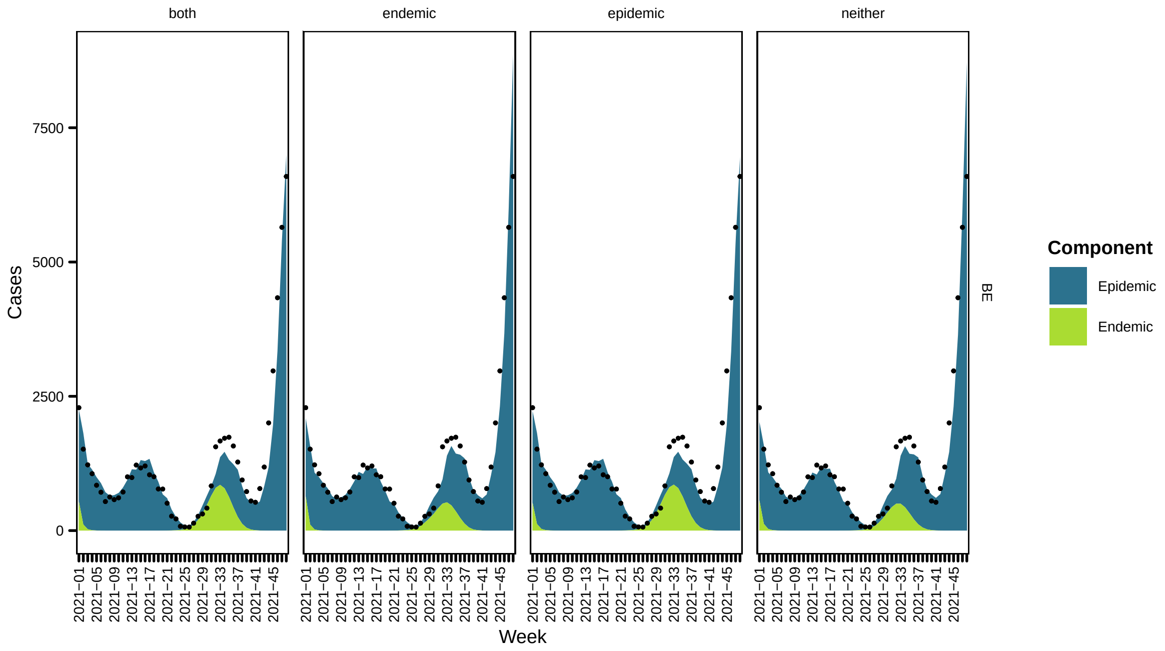
both
endemic
epidemic
neither
Cases
0
2500
5000
7500
2021−01
2021−05
2021−09
2021−13
2021−17
2021−21
2021−25
2021−29
2021−33
2021−37
2021−41
2021−45
Week
BE
Component
Epidemic
Endemic

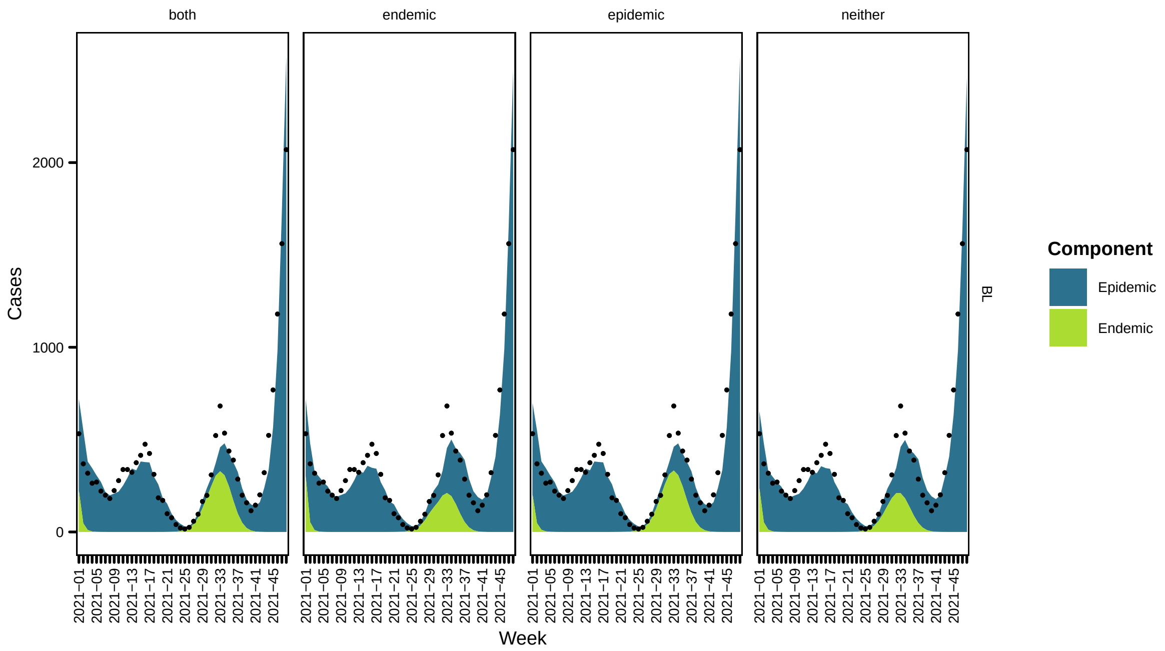
both
endemic
epidemic
neither
Cases
Week
BL
Component
Epidemic
Endemic

43

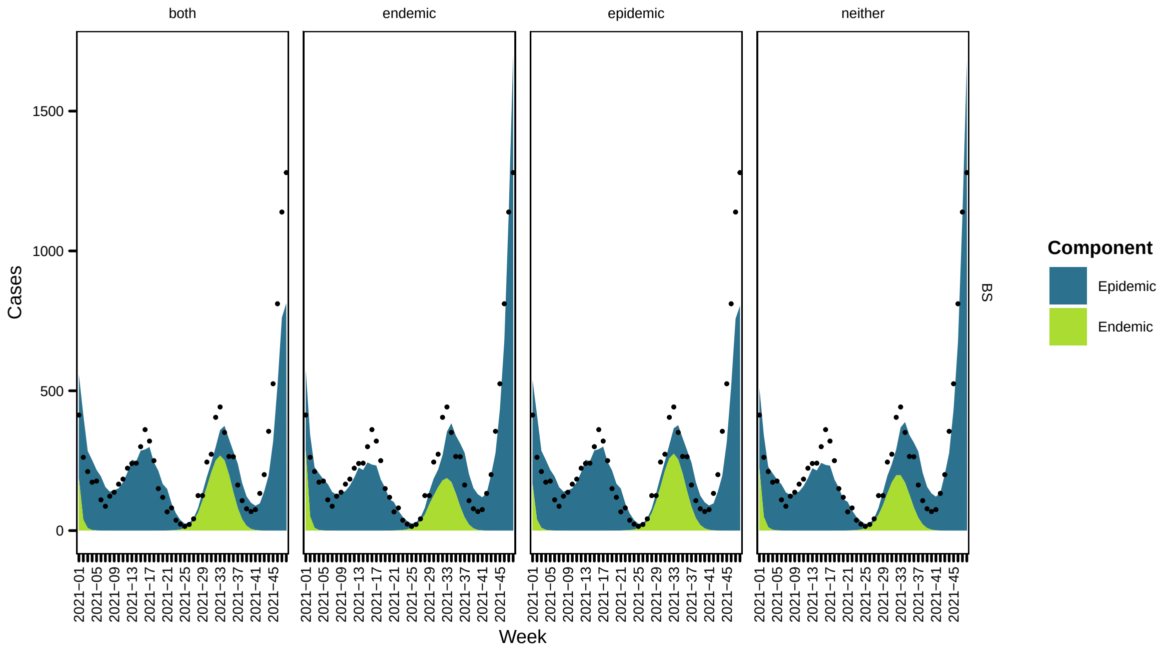
both
endemic
epidemic
neither
Cases
Week
BS
Component
Epidemic
Endemic

44

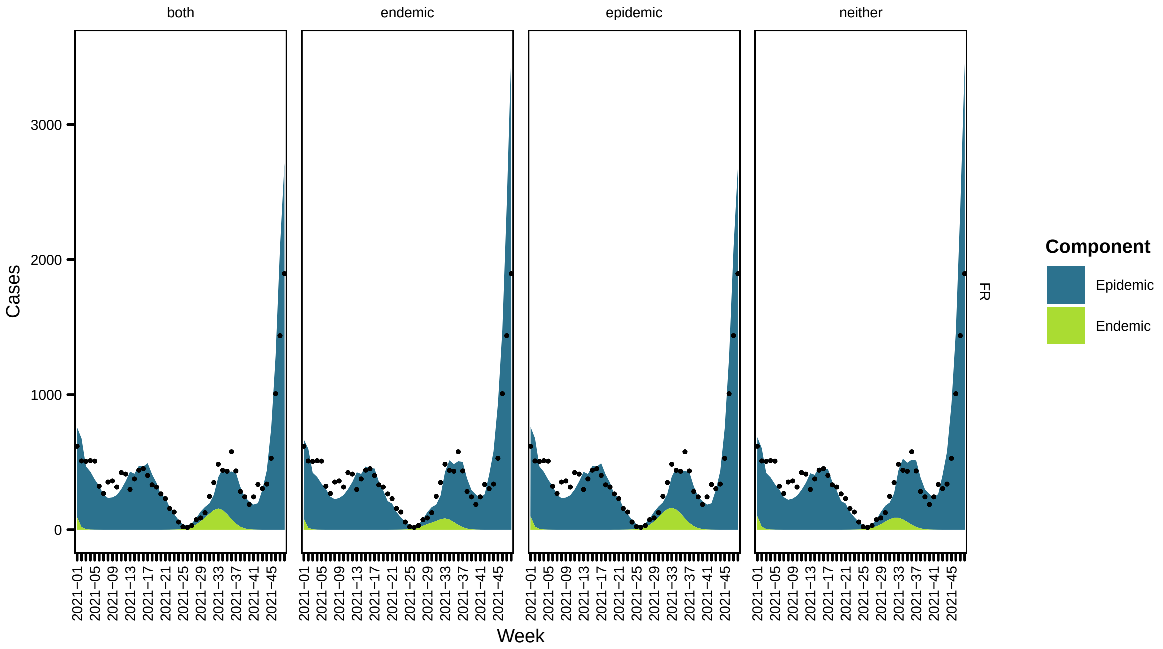
both
endemic
epidemic
neither
Cases
3000
2000
1000
0
Week
FR
Component
Epidemic
Endemic

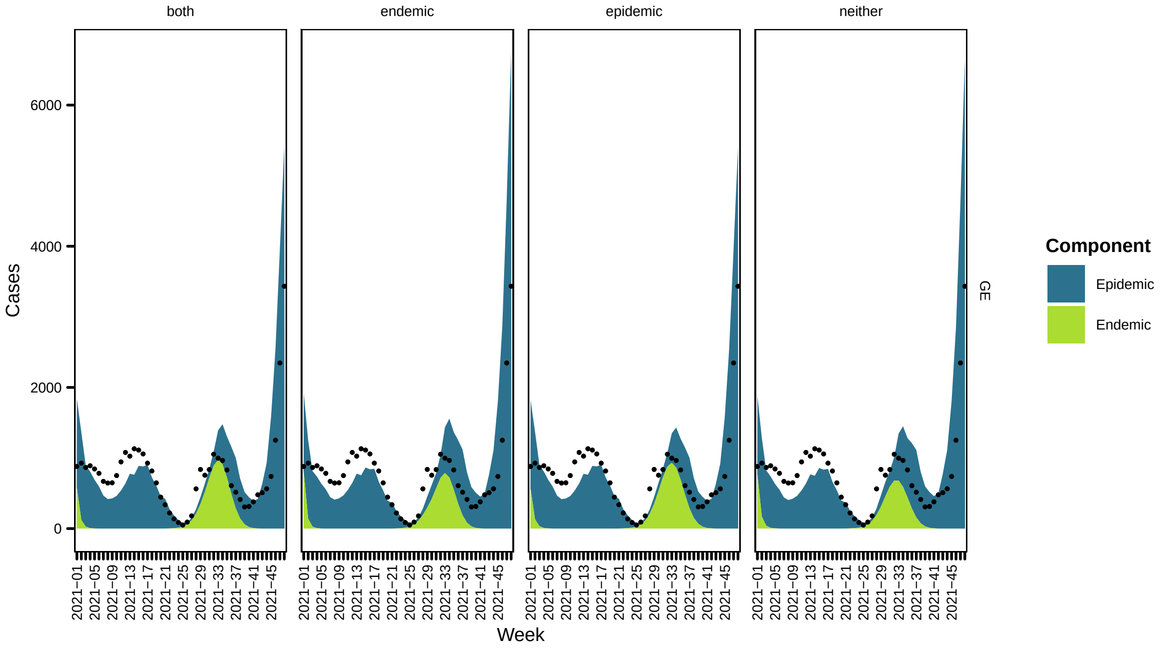
both
endemic
epidemic
neither
Cases
6000
4000
2000
0
Week
GE
Component
Epidemic
Endemic

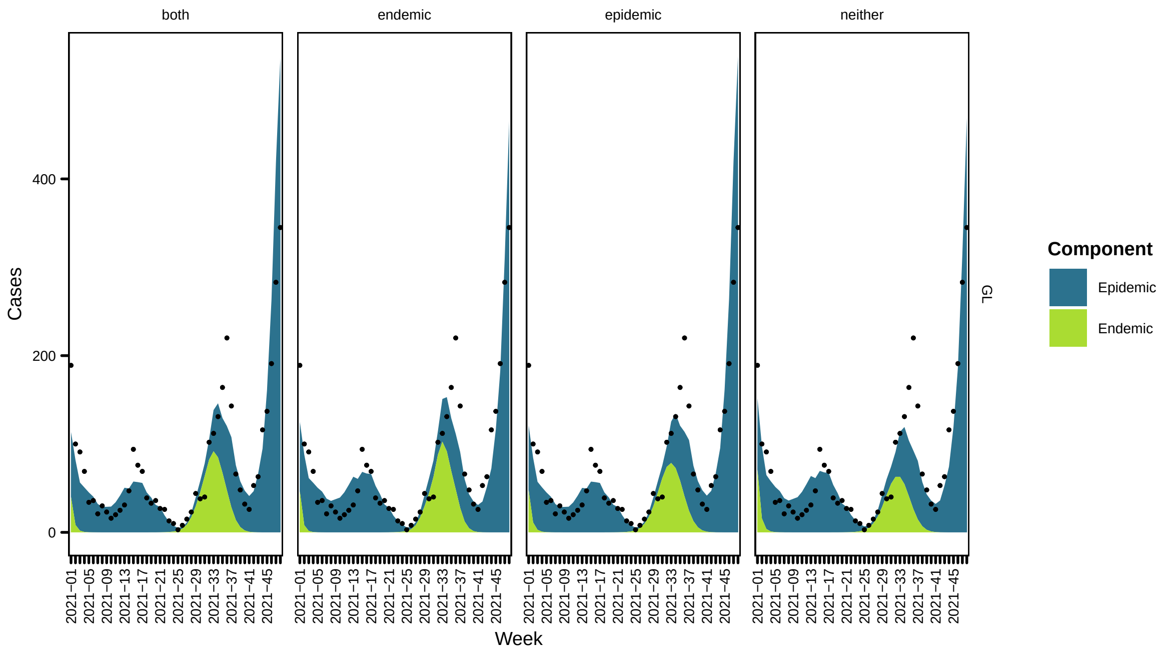
both
endemic
epidemic
neither
Cases
0
200
400
Week
2021−01
2021−05
2021−09
2021−13
2021−17
2021−21
2021−25
2021−29
2021−33
2021−37
2021−41
2021−45
GL
Component
Epidemic
Endemic

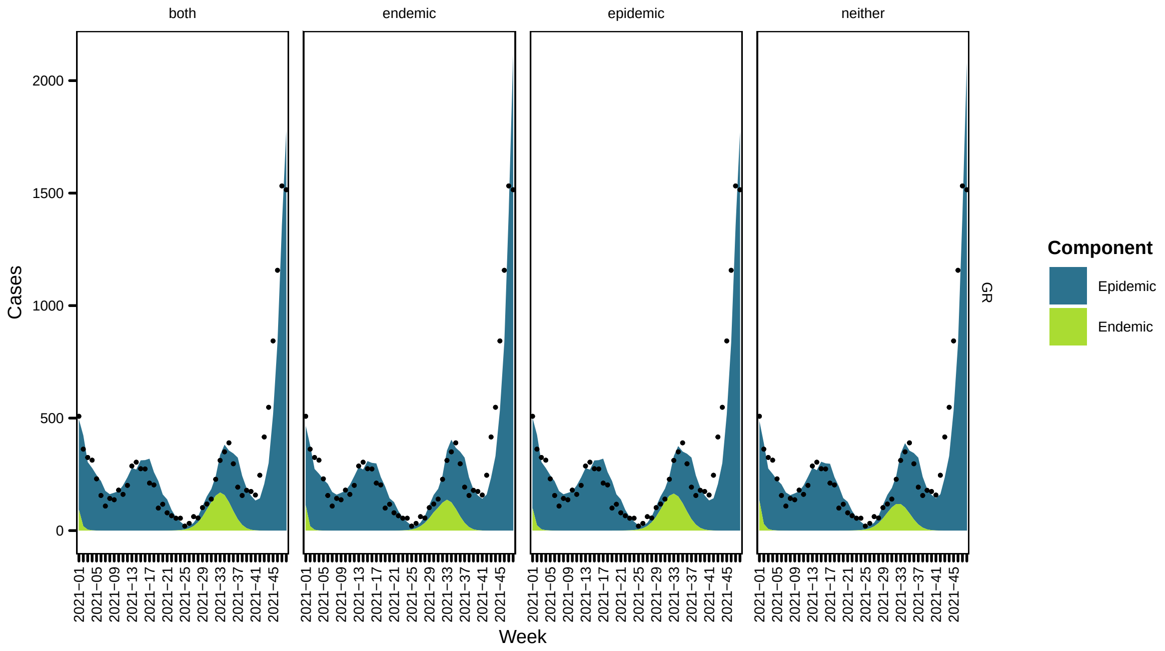
both
endemic
epidemic
neither
Cases
0
500
1000
1500
2000
Week
2021−01
2021−05
2021−09
2021−13
2021−17
2021−21
2021−25
2021−29
2021−33
2021−37
2021−41
2021−45
GR
Component
Epidemic
Endemic

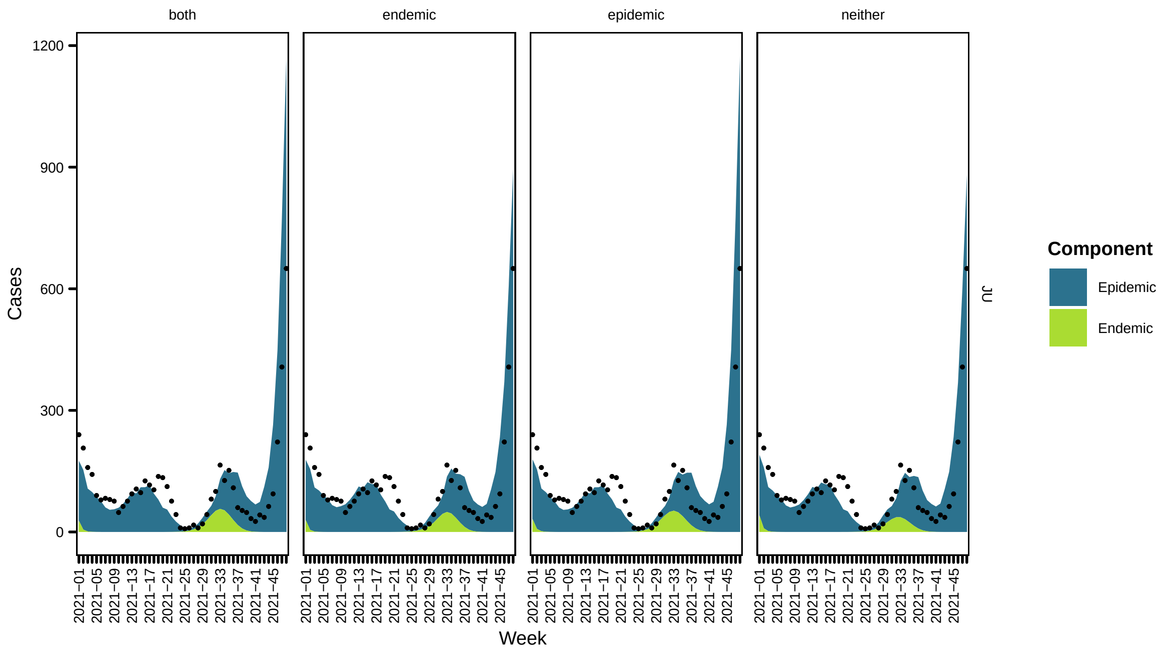
both
endemic
epidemic
neither
Cases
1200
900
600
300
0
2021−01
2021−05
2021−09
2021−13
2021−17
2021−21
2021−25
2021−29
2021−33
2021−37
2021−41
2021−45
Week
JU
Component
Epidemic
Endemic

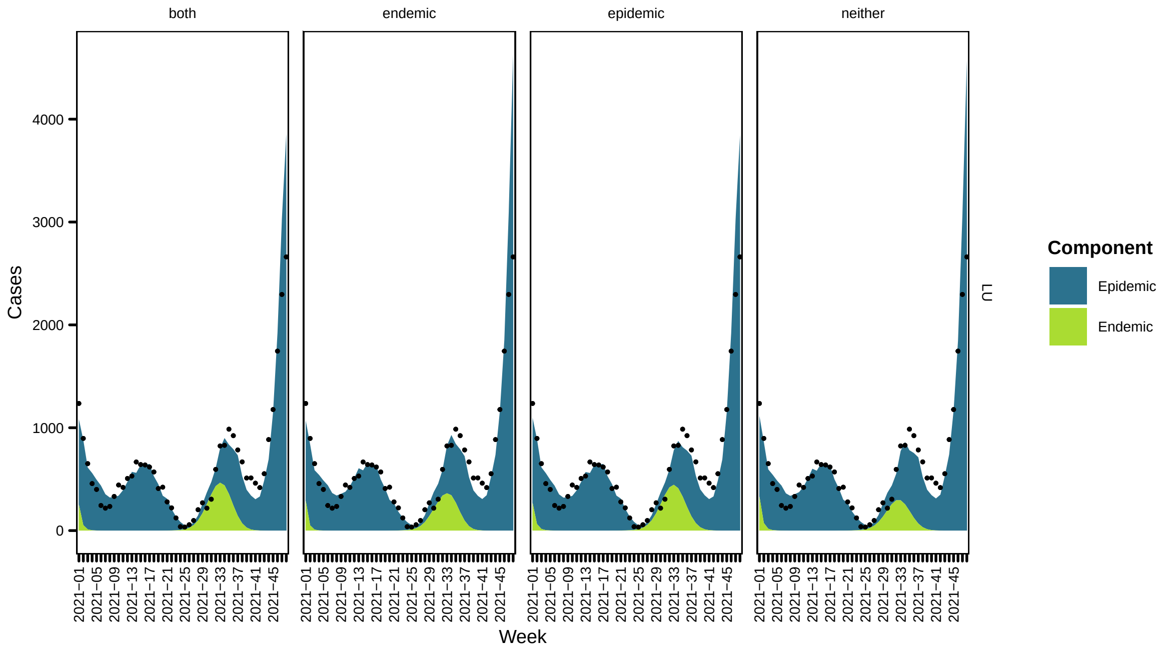
both
endemic
epidemic
neither
Cases
4000
3000
2000
1000
0
2021−01
2021−05
2021−09
2021−13
2021−17
2021−21
2021−25
2021−29
2021−33
2021−37
2021−41
2021−45
Week
LU
Component
Epidemic
Endemic

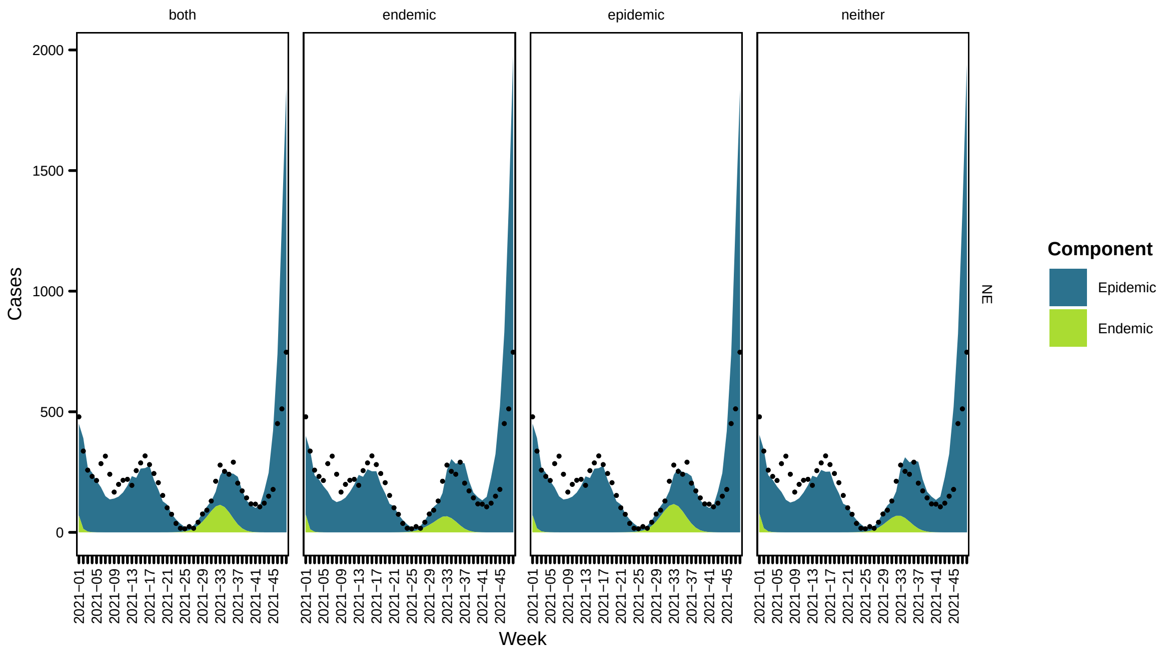
both
endemic
epidemic
neither
Cases
Week
Component
Epidemic
Endemic
NE

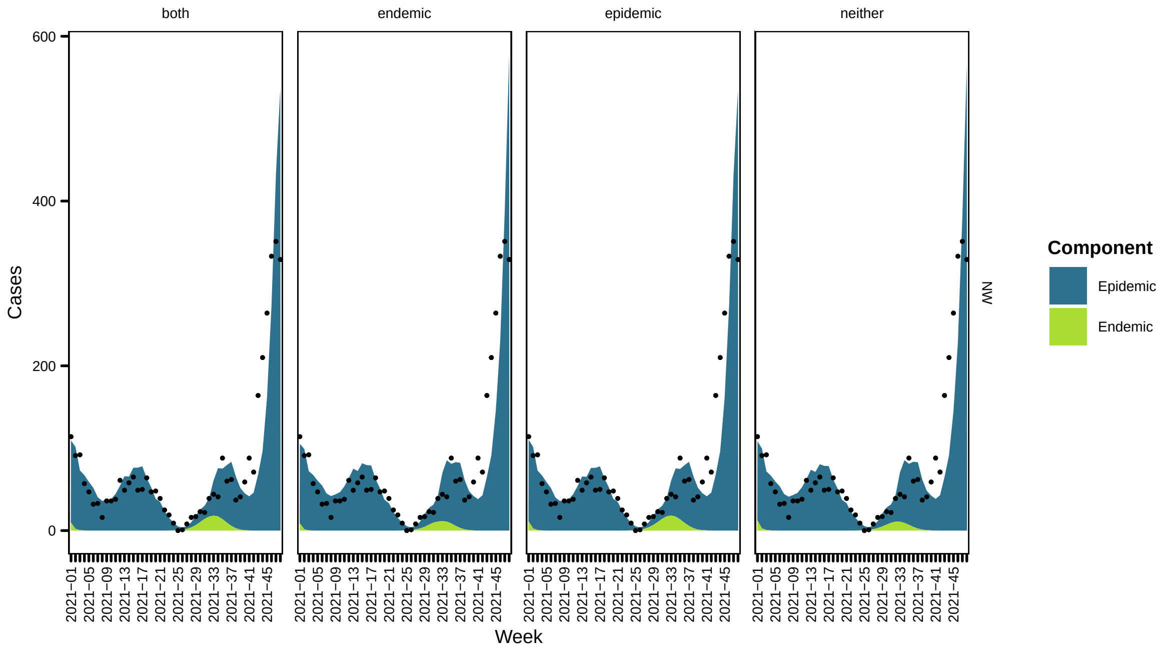
both
endemic
epidemic
neither
Cases
Week
Component
Epidemic
Endemic
NW

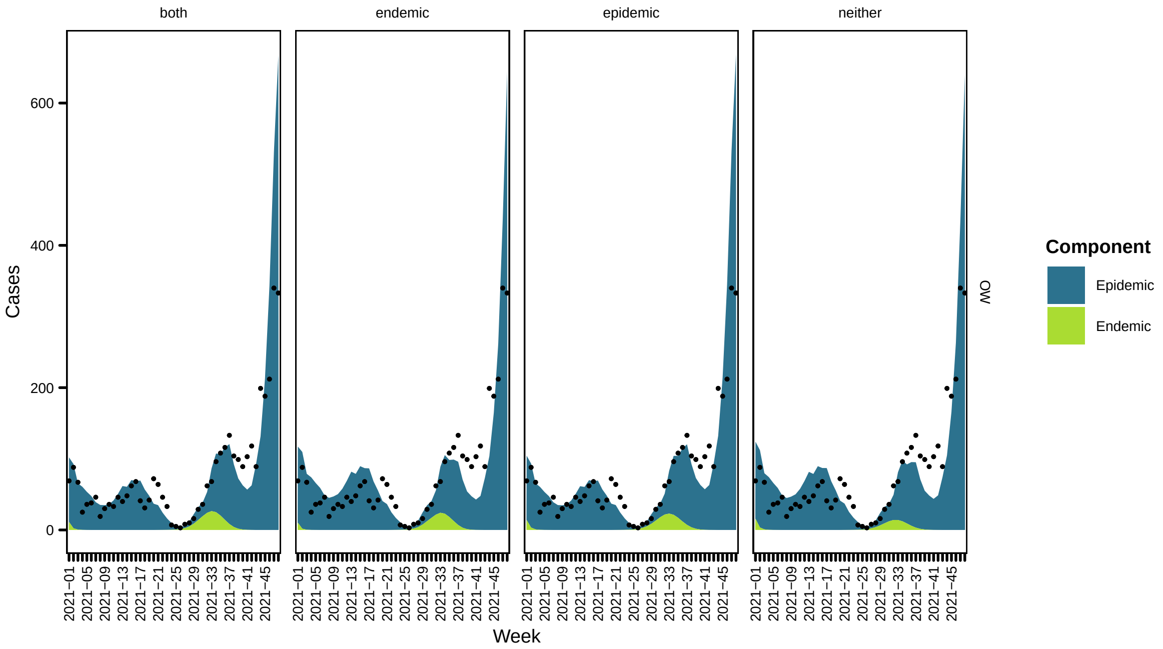
both
endemic
epidemic
neither
Cases
600
400
200
0
2021−01
2021−05
2021−09
2021−13
2021−17
2021−21
2021−25
2021−29
2021−33
2021−37
2021−41
2021−45
Week
OW
Component
Epidemic
Endemic

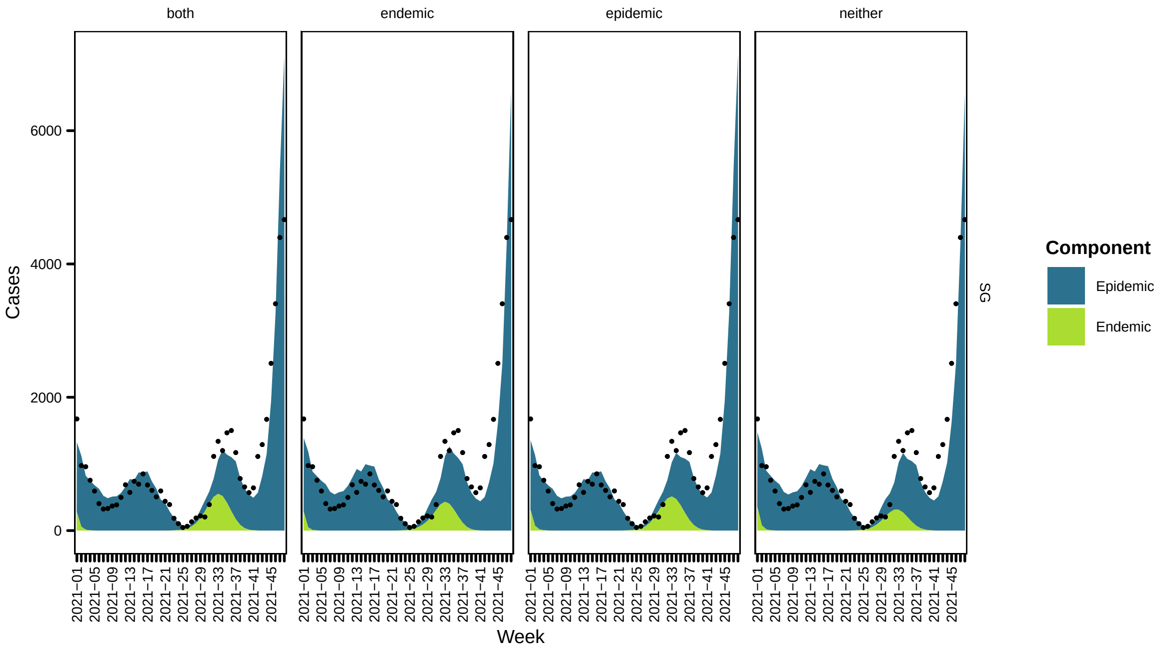
both
endemic
epidemic
neither
Cases
6000
4000
2000
0
2021−01
2021−05
2021−09
2021−13
2021−17
2021−21
2021−25
2021−29
2021−33
2021−37
2021−41
2021−45
Week
SG
Component
Epidemic
Endemic

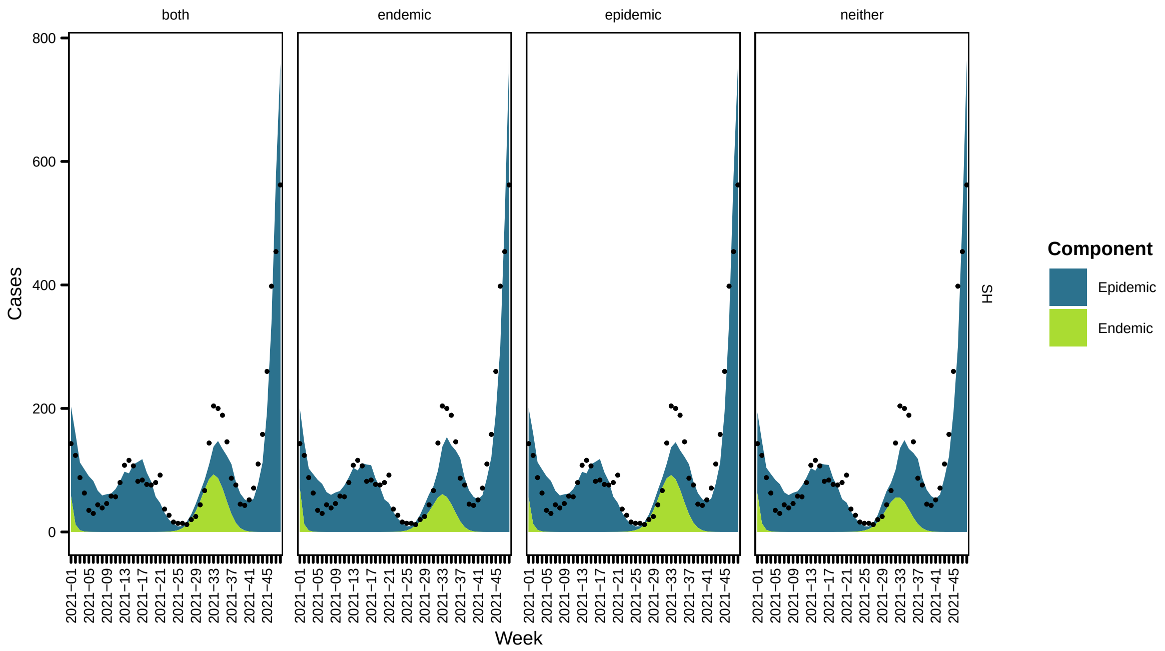
both
endemic
epidemic
neither
Cases
Week
Component
Epidemic
Endemic
SH

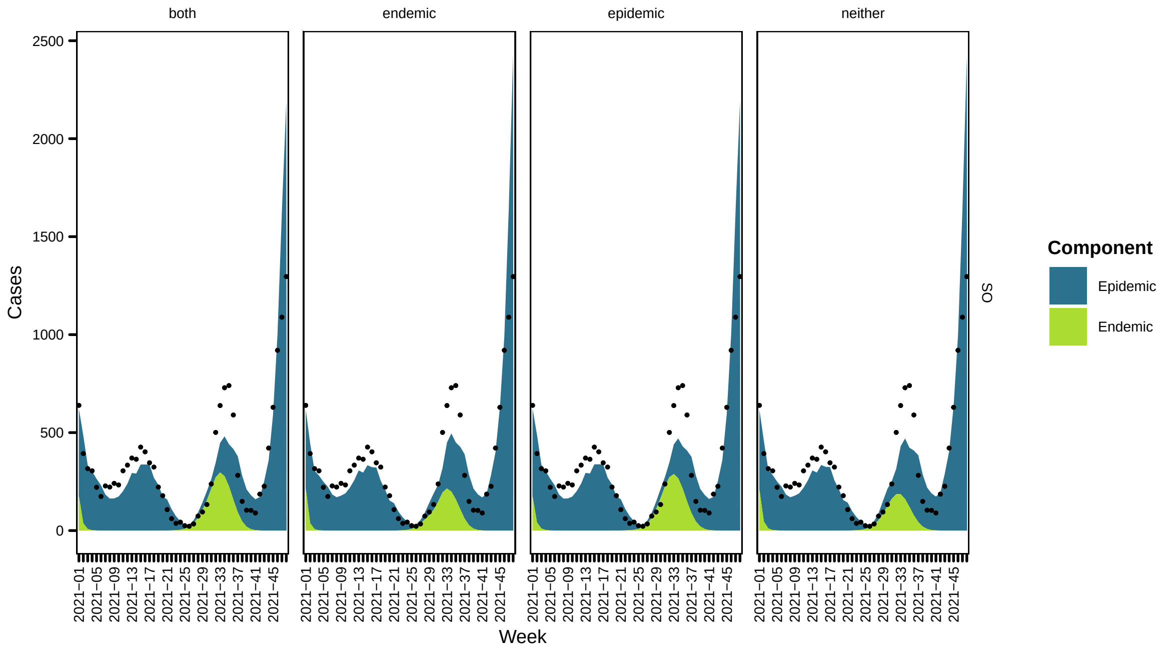
both
endemic
epidemic
neither
Cases
Week
Component
Epidemic
Endemic
SO

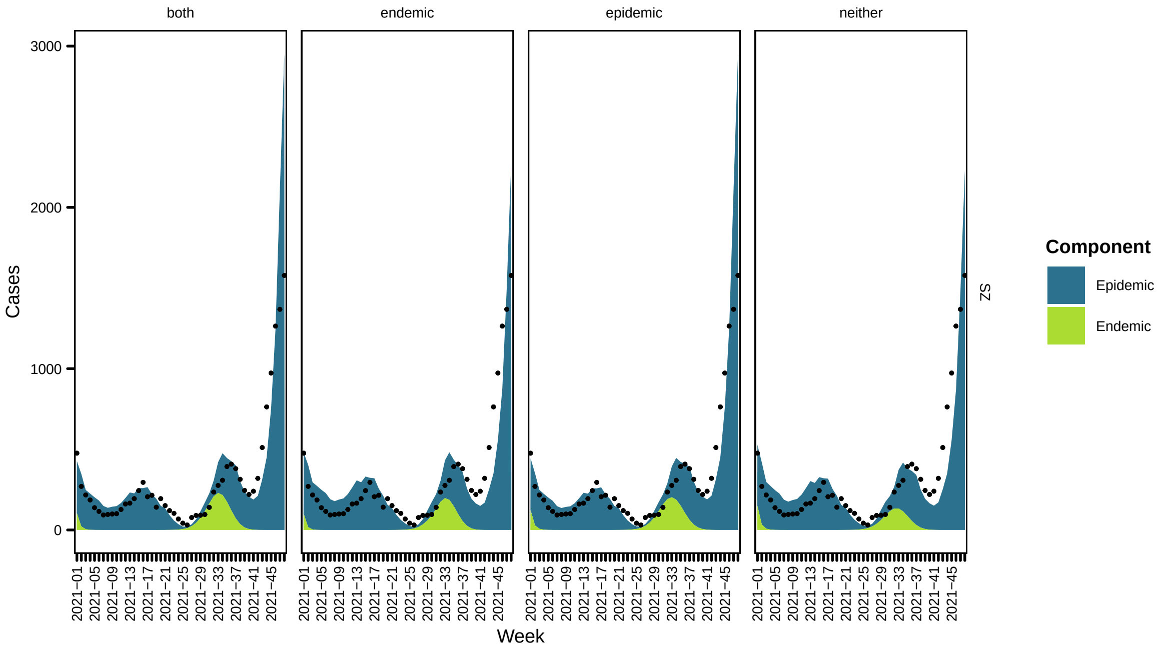

both
endemic
epidemic
neither
Cases
0
1000
2000
3000
2021−01
2021−05
2021−09
2021−13
2021−17
2021−21
2021−25
2021−29
2021−33
2021−37
2021−41
2021−45
Week
SZ
Component
Epidemic
Endemic


57

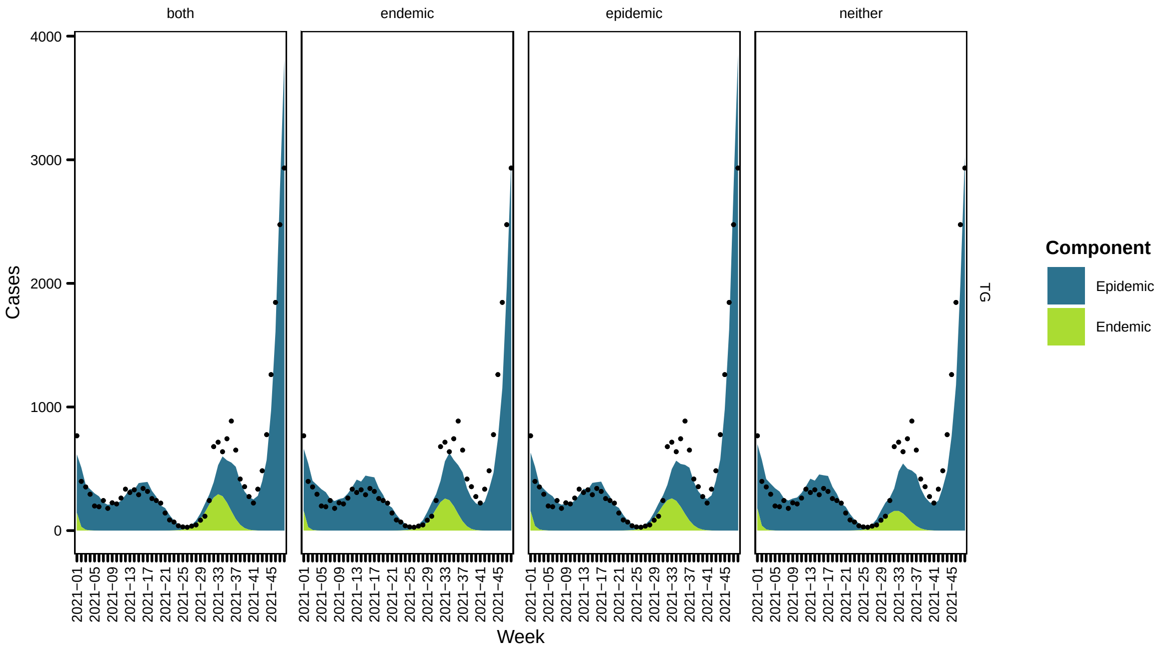

both
endemic
epidemic
neither
Cases
0
1000
2000
3000
4000
2021−01
2021−05
2021−09
2021−13
2021−17
2021−21
2021−25
2021−29
2021−33
2021−37
2021−41
2021−45
Week
TG
Component
Epidemic
Endemic


58

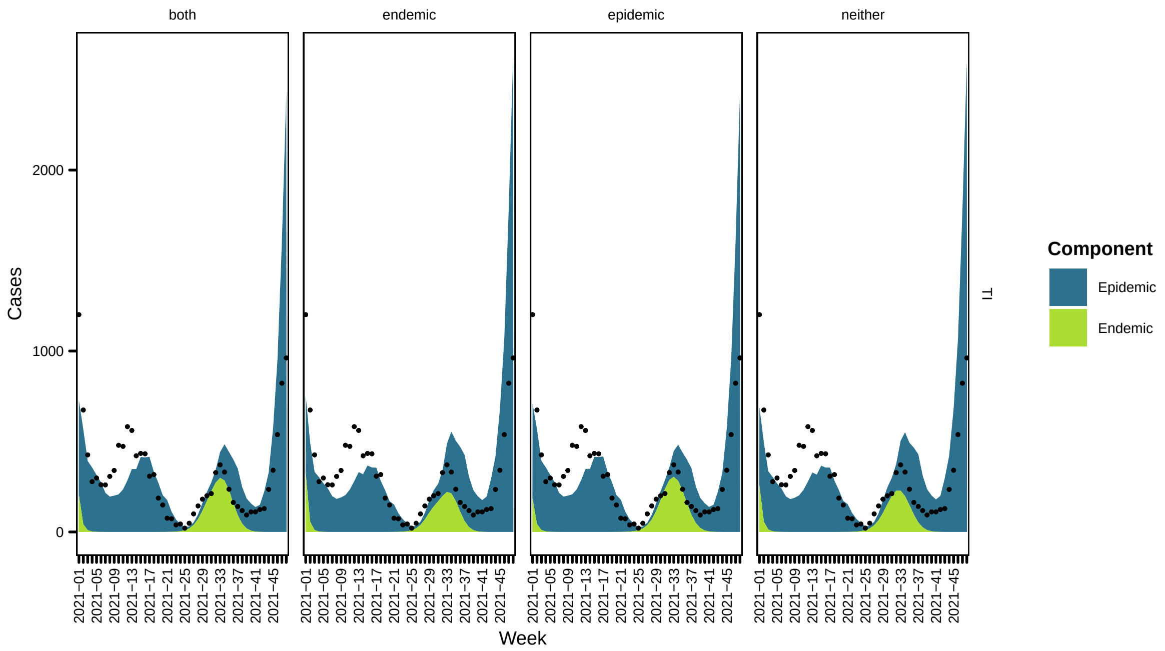
both
endemic
epidemic
neither
Cases
2000
1000
0
TI
Week
Component
Epidemic
Endemic

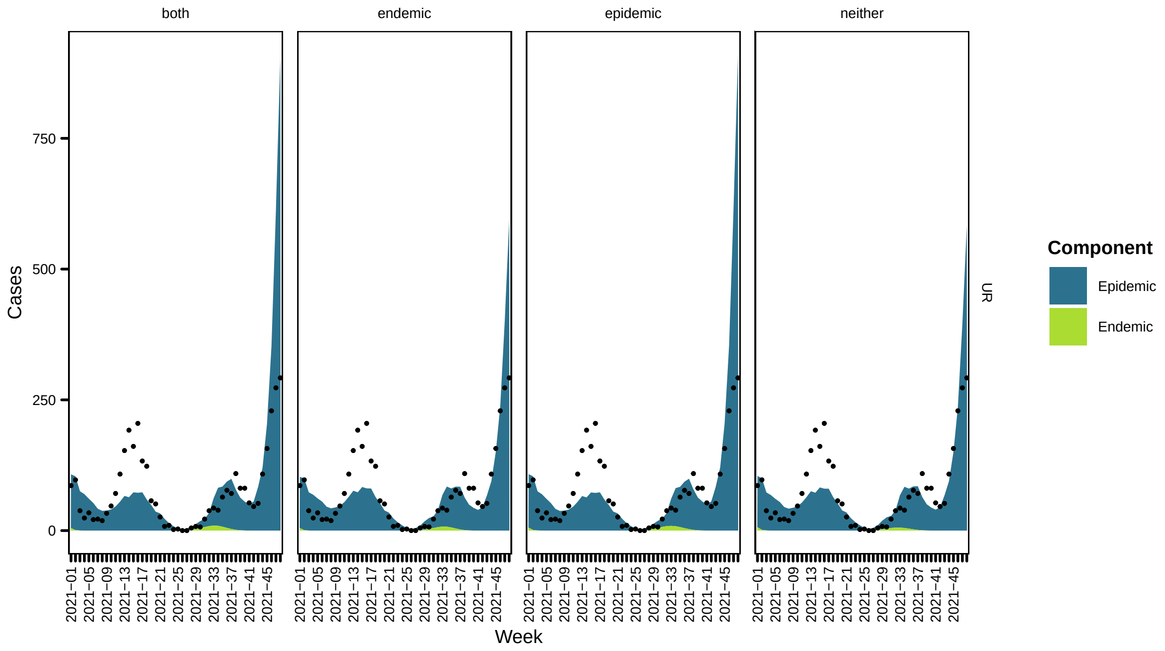
both
endemic
epidemic
neither
Cases
750
500
250
0
UR
Week
Component
Epidemic
Endemic

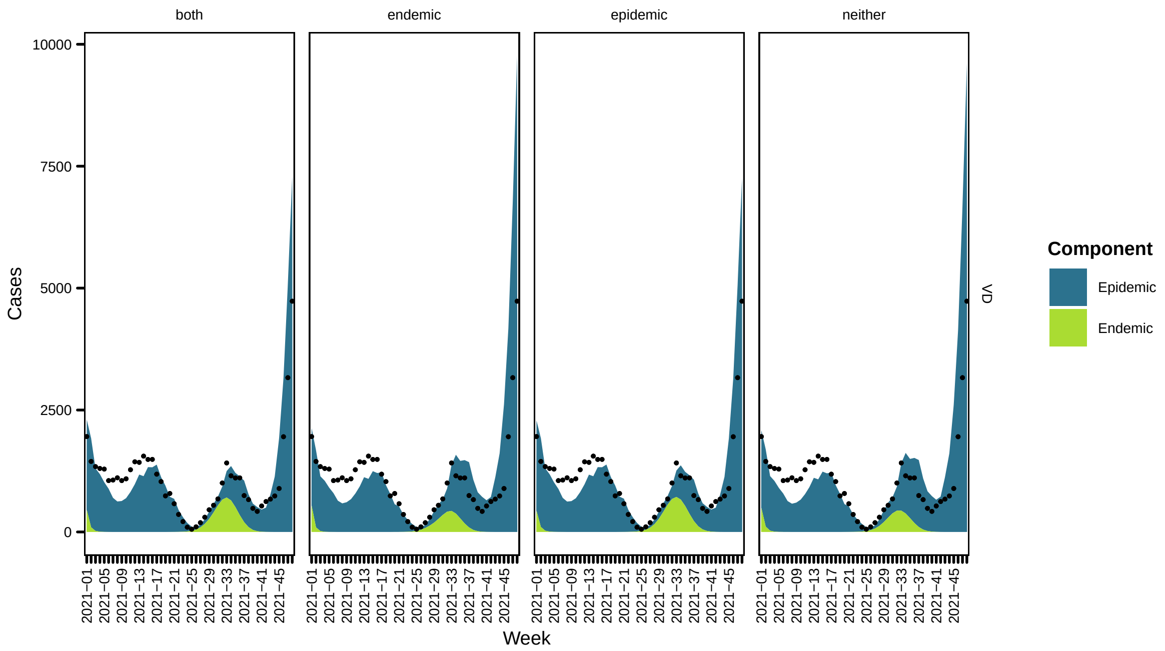
both
endemic
epidemic
neither
Cases
10000
7500
5000
2500
0
Week
VD
Component
Epidemic
Endemic

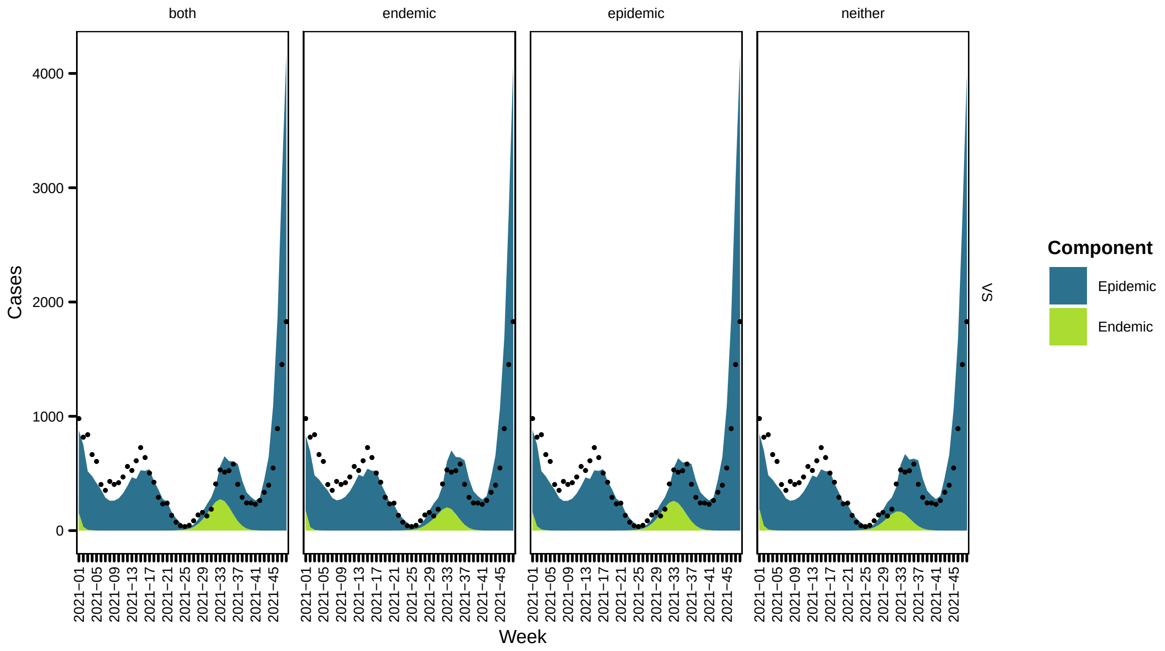
both
endemic
epidemic
neither
Cases
4000
3000
2000
1000
0
Week
VS
Component
Epidemic
Endemic

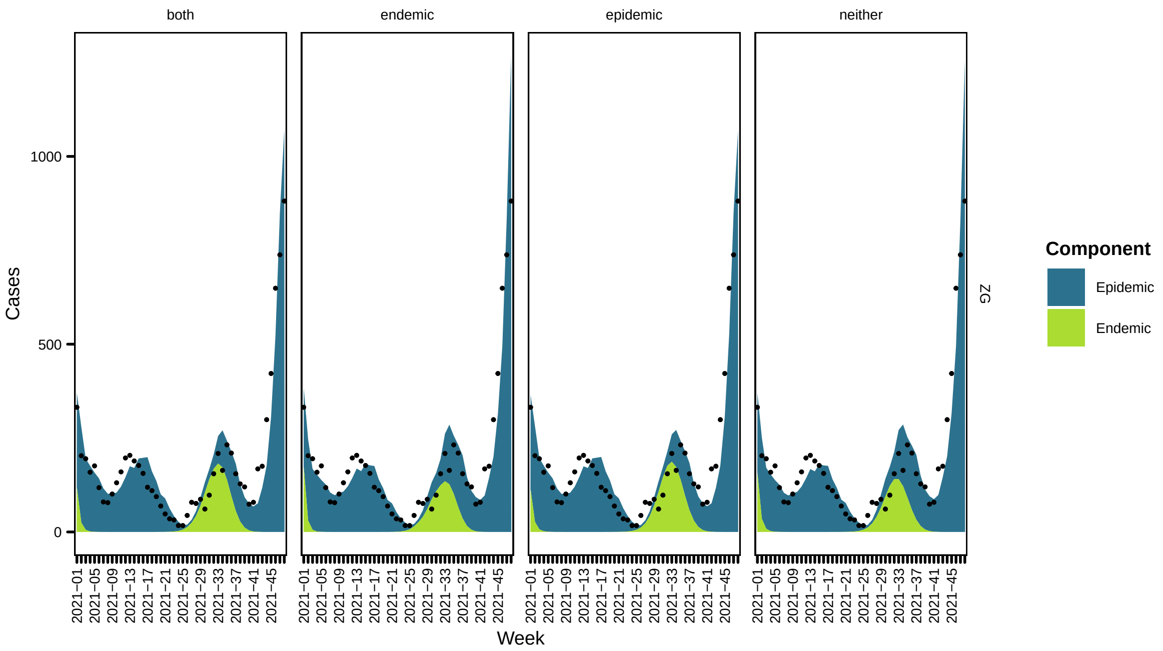


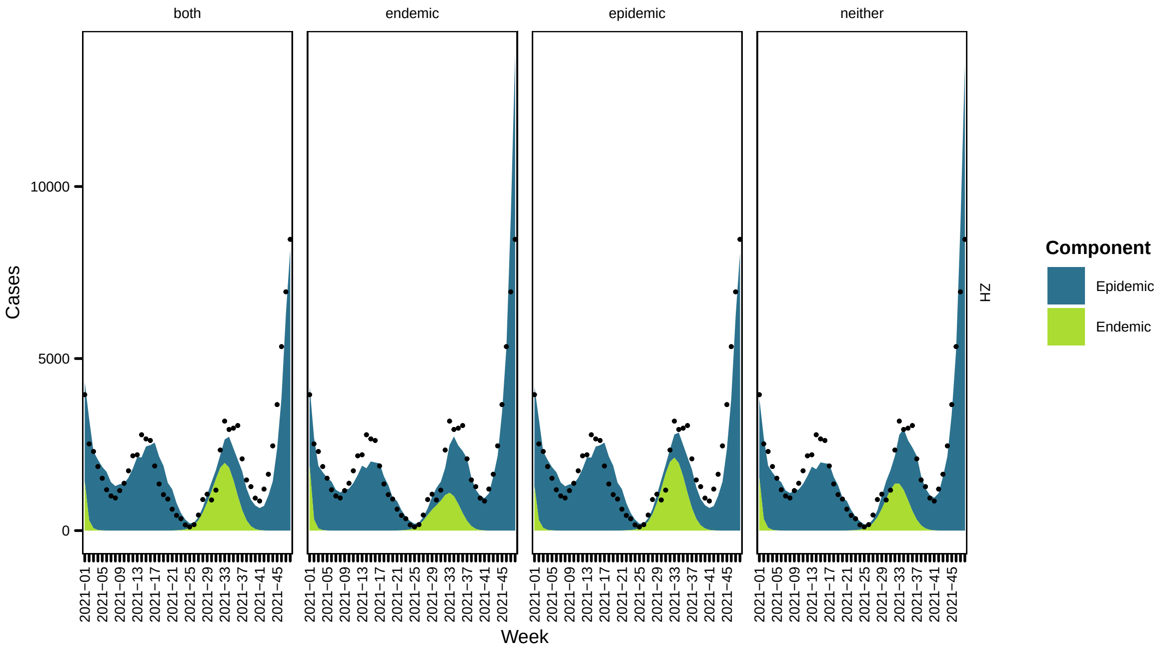


#### 2.2 Time-varying regional transmission weights

Now the model contains the time-varying transmission weights. A snapshot is shown below

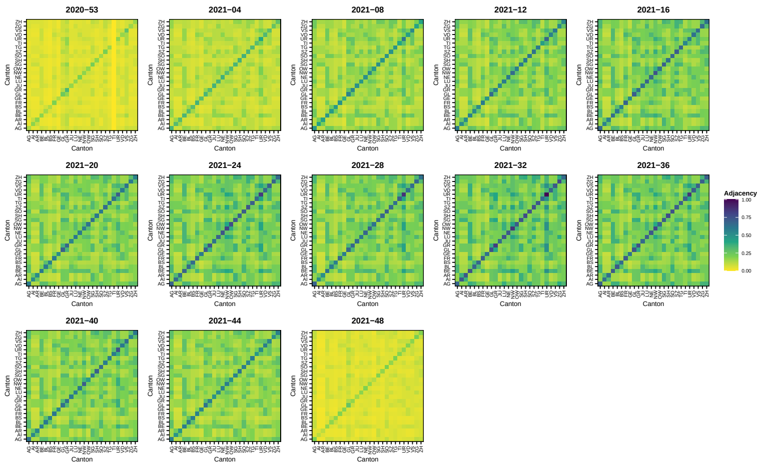


| | neither | endemic | epidemic | both |
|---|---|---|---|---|
| $\gamma^{\phi}_{\sin(2\pi t/52)}$ | 0.674 (SE 0.04) | 0.687 (SE 0.039) | 0.247 (SE 0.049) | 0.255 (SE 0.049) |
| $\gamma^{\phi}_{\cos(2\pi t/52)}$ | 0.708 (SE 0.023) | 0.722 (SE 0.023) | 0.818 (SE 0.024) | 0.821 (SE 0.024) |
| $\gamma^{\phi}_{\text{gravity}}$ | 0.779 (SE 0.033) | 0.782 (SE 0.033) | 0.838 (SE 0.029) | 0.838 (SE 0.029) |
| $\gamma^{\phi}_{\text{time}}$ | 0.036 (SE 0.002) | 0.036 (SE 0.002) | 0.085 (SE 0.004) | 0.084 (SE 0.004) |
| $\alpha^{\phi}_{\text{region}}$ | 0.798 (SE 0.131) | 0.802 (SE 0.13) | 2.118 (SE 0.15) | 2.099 (SE 0.15) |
| $\gamma^{\nu}_{\sin(2\pi t/52)}$ | -9.588 (SE 0.564) | -11.467 (SE 0.832) | -9.393 (SE 0.425) | -10.438 (SE 0.669) |
| $\gamma^{\nu}_{\cos(2\pi t/52)}$ | -3.972 (SE 0.402) | -4.586 (SE 0.45) | -3.621 (SE 0.281) | -4.004 (SE 0.342) |
| $\gamma^{\nu}_{\text{time}}$ | -0.48 (SE 0.036) | -0.475 (SE 0.035) | -0.446 (SE 0.026) | -0.448 (SE 0.026) |
| $\alpha^{\nu}_{\text{region}}$ | 3.208 (SE 0.397) | 4.092 (SE 0.471) | 3.575 (SE 0.294) | 3.982 (SE 0.358) |
| $\psi$ | 0.117 (SE 0.005) | 0.115 (SE 0.005) | 0.101 (SE 0.005) | 0.101 (SE 0.005) |
| $\beta^{(\nu)}$ | | 3.312 (SE 1.035) | | 1.739 (SE 0.846) |
| $\beta^{(\phi)}$ | | | 2.688 (SE 0.201) | 2.642 (SE 0.2) |

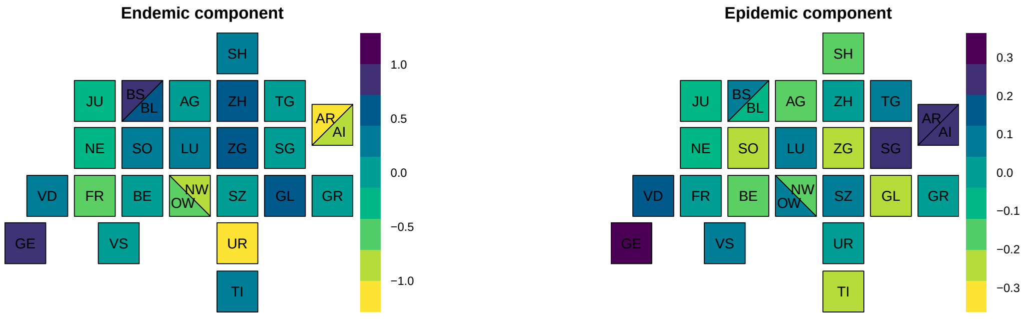

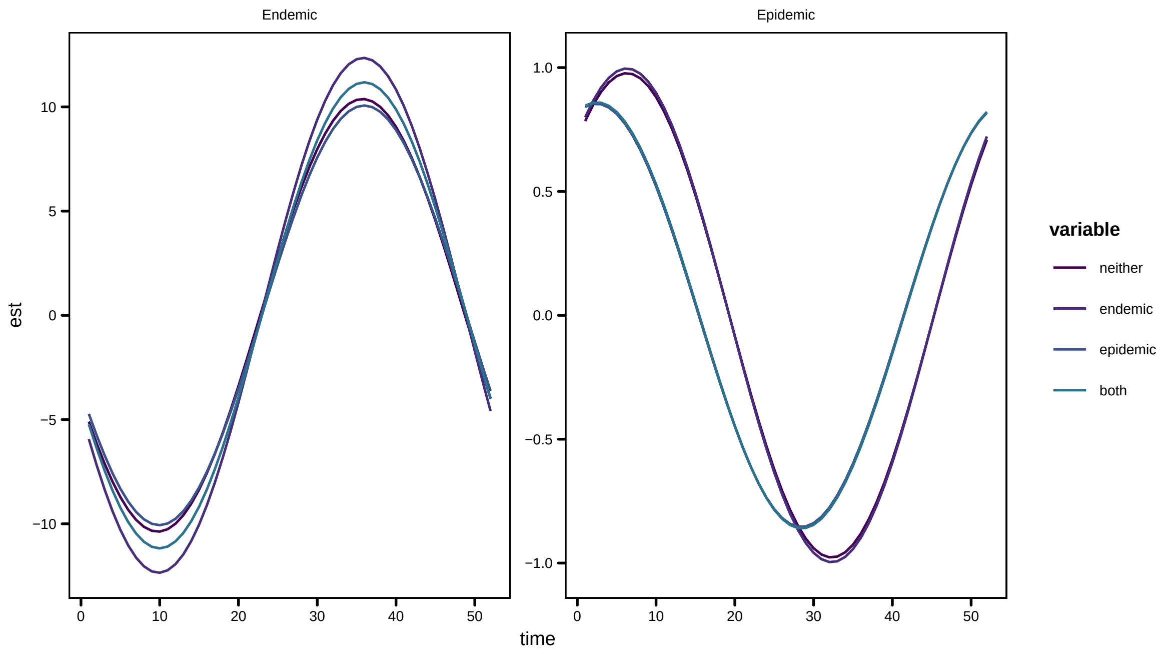
Endemic
Epidemic
est
time
variable
neither
endemic
epidemic
both

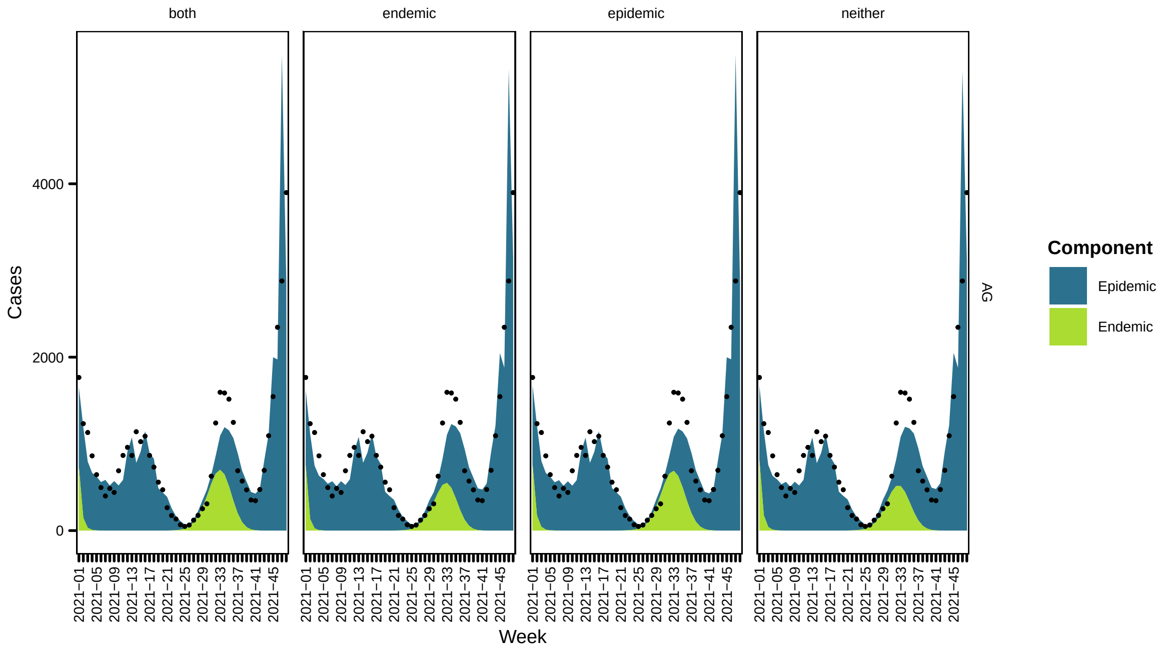
both
endemic
epidemic
neither
Cases
Week
AG
Component
Epidemic
Endemic

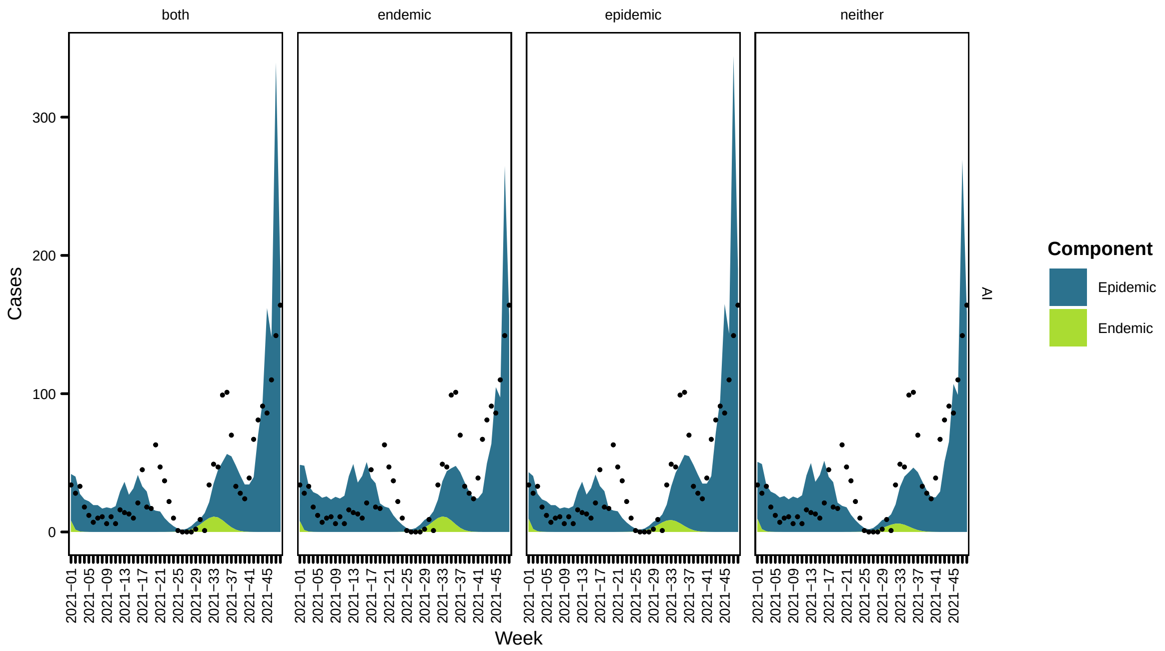

both
endemic
epidemic
neither
Cases
Week
Component
Epidemic
Endemic
AI


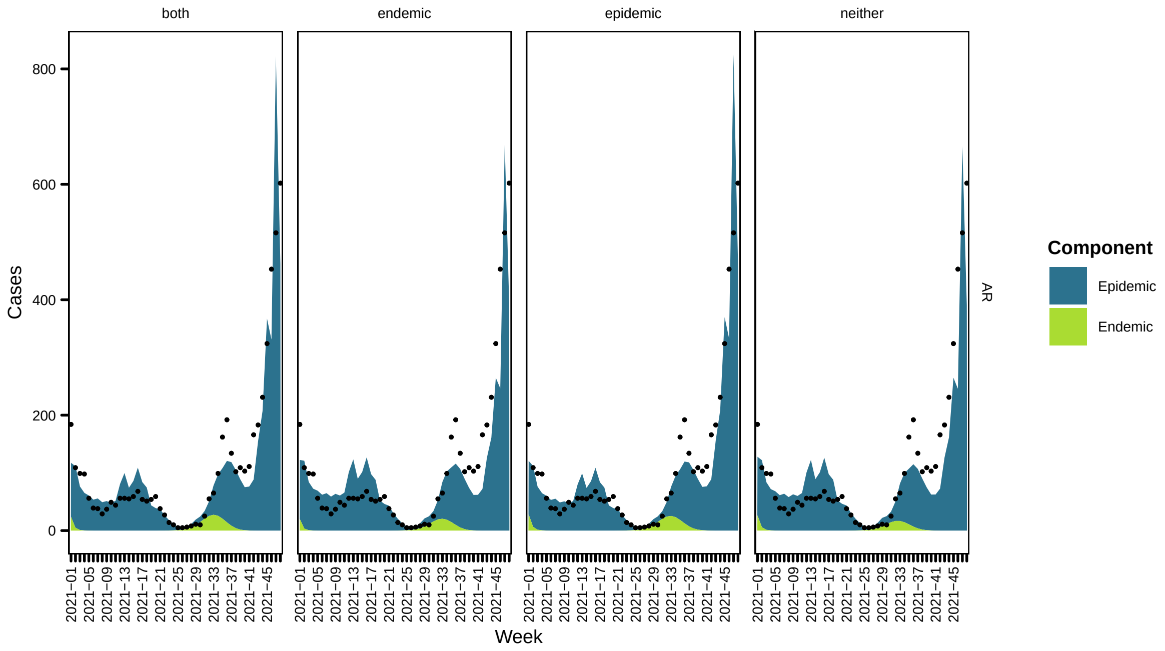

both
endemic
epidemic
neither
Cases
Week
Component
Epidemic
Endemic
AR

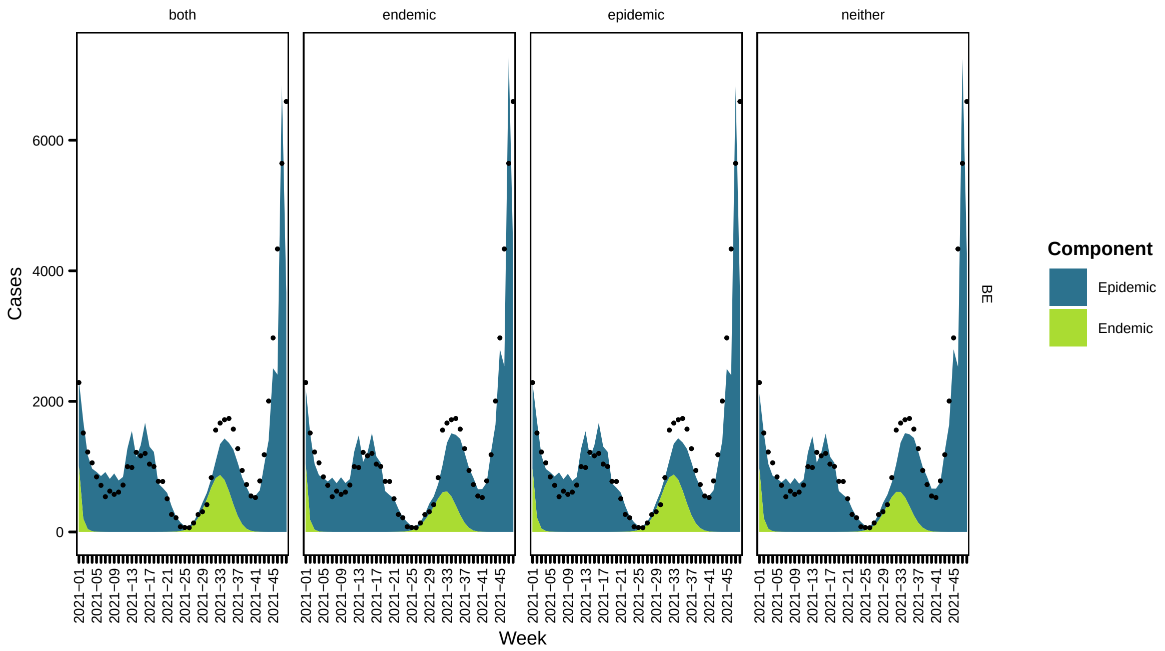
both
endemic
epidemic
neither
Cases
0
2000
4000
6000
Week
2021−01
2021−05
2021−09
2021−13
2021−17
2021−21
2021−25
2021−29
2021−33
2021−37
2021−41
2021−45
BE
Component
Epidemic
Endemic

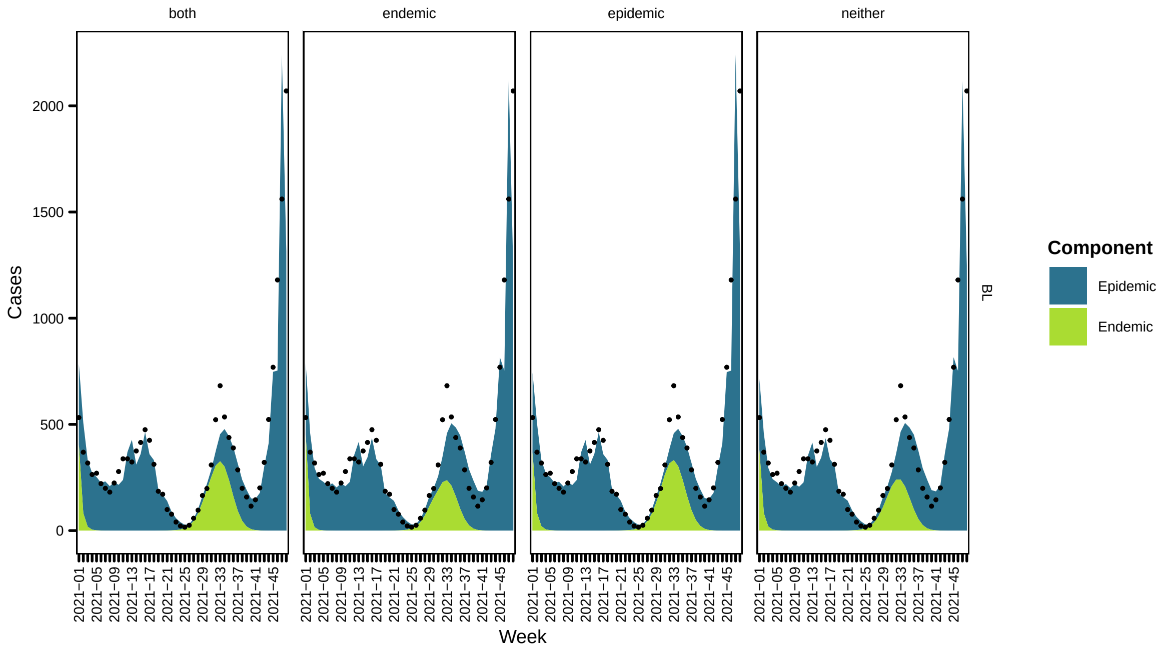
both
endemic
epidemic
neither
Cases
0
500
1000
1500
2000
Week
2021−01
2021−05
2021−09
2021−13
2021−17
2021−21
2021−25
2021−29
2021−33
2021−37
2021−41
2021−45
BL
Component
Epidemic
Endemic

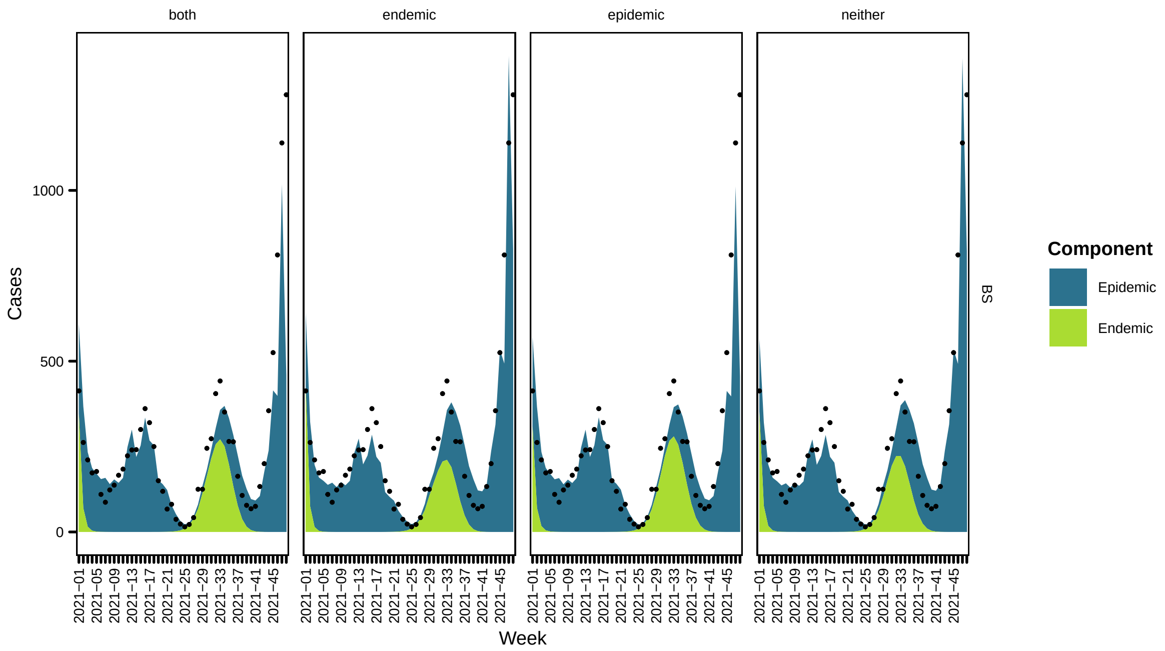

both
endemic
epidemic
neither
Cases
0
500
1000
Week
2021−01
2021−05
2021−09
2021−13
2021−17
2021−21
2021−25
2021−29
2021−33
2021−37
2021−41
2021−45
BS
Component
Epidemic
Endemic


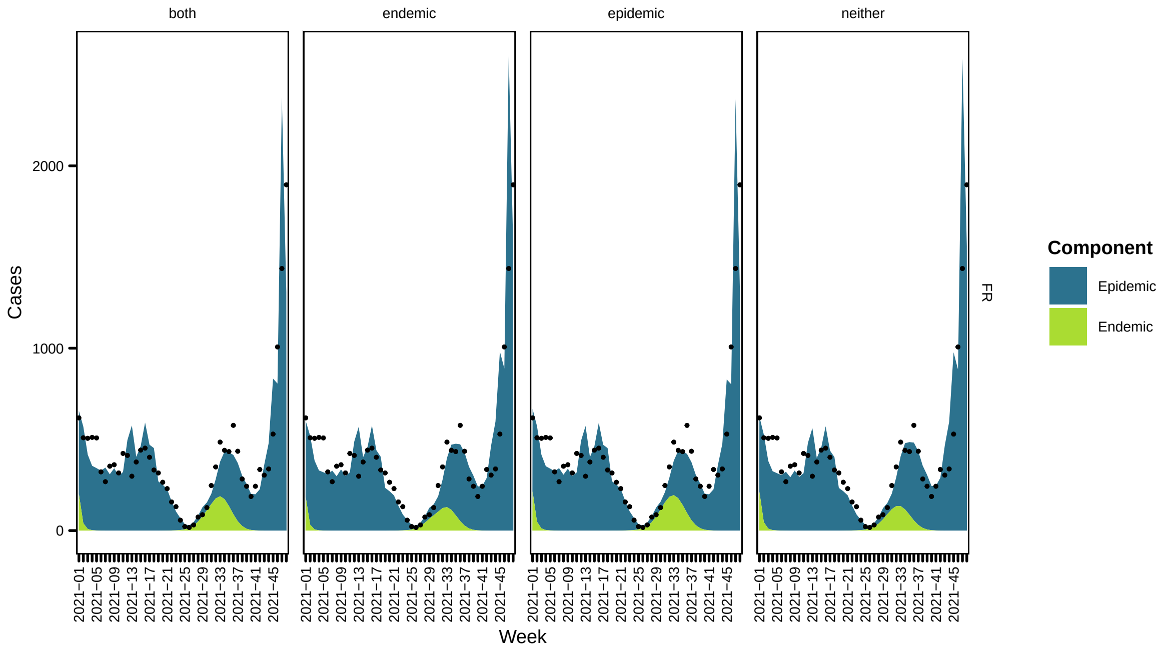

both
endemic
epidemic
neither
Cases
0
1000
2000
Week
2021−01
2021−05
2021−09
2021−13
2021−17
2021−21
2021−25
2021−29
2021−33
2021−37
2021−41
2021−45
FR
Component
Epidemic
Endemic

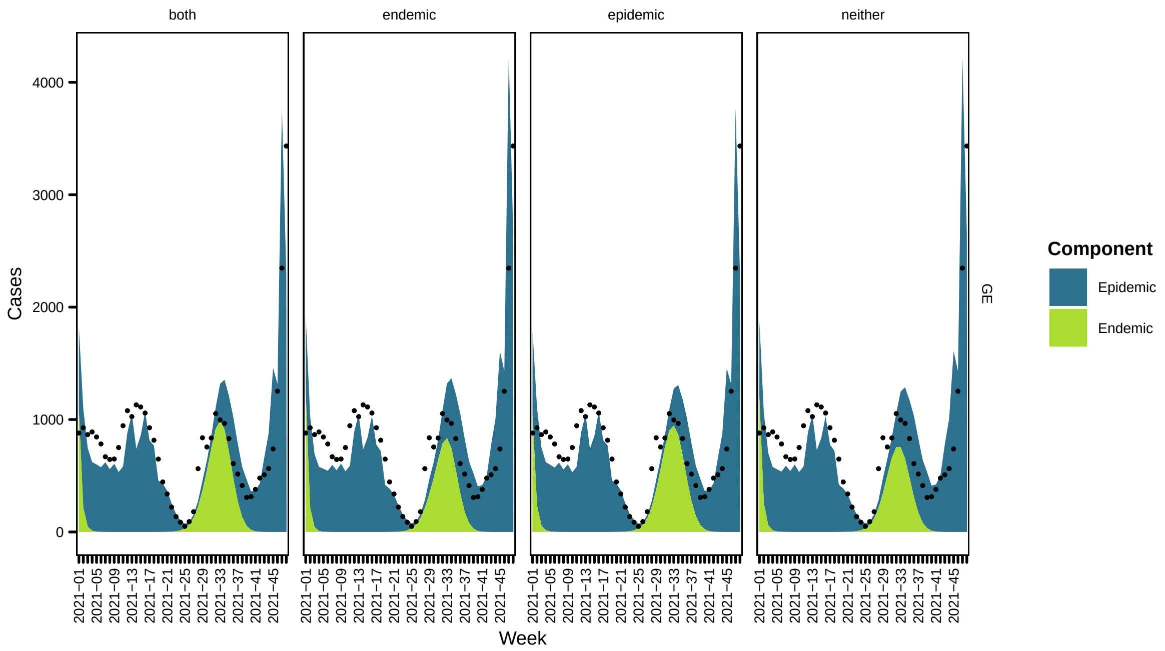
both
endemic
epidemic
neither
Cases
0
1000
2000
3000
4000
Week
GE
Component
Epidemic
Endemic
2021–01
2021–05
2021–09
2021–13
2021–17
2021–21
2021–25
2021–29
2021–33
2021–37
2021–41
2021–45

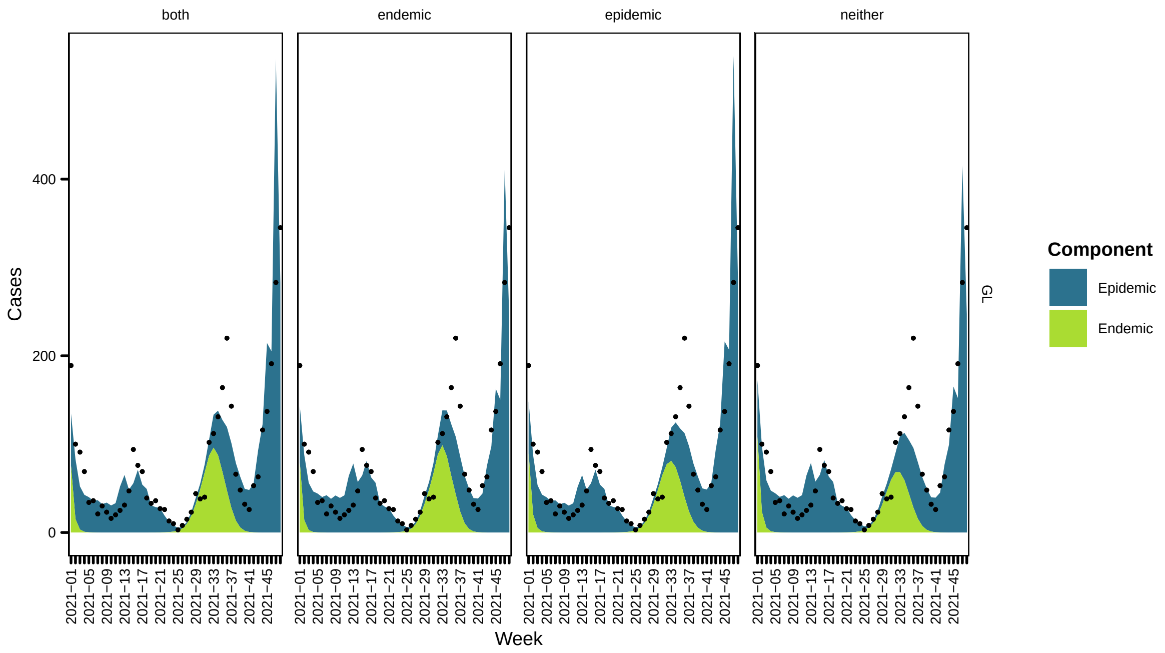
both
endemic
epidemic
neither
Cases
0
200
400
Week
GL
Component
Epidemic
Endemic
2021–01
2021–05
2021–09
2021–13
2021–17
2021–21
2021–25
2021–29
2021–33
2021–37
2021–41
2021–45

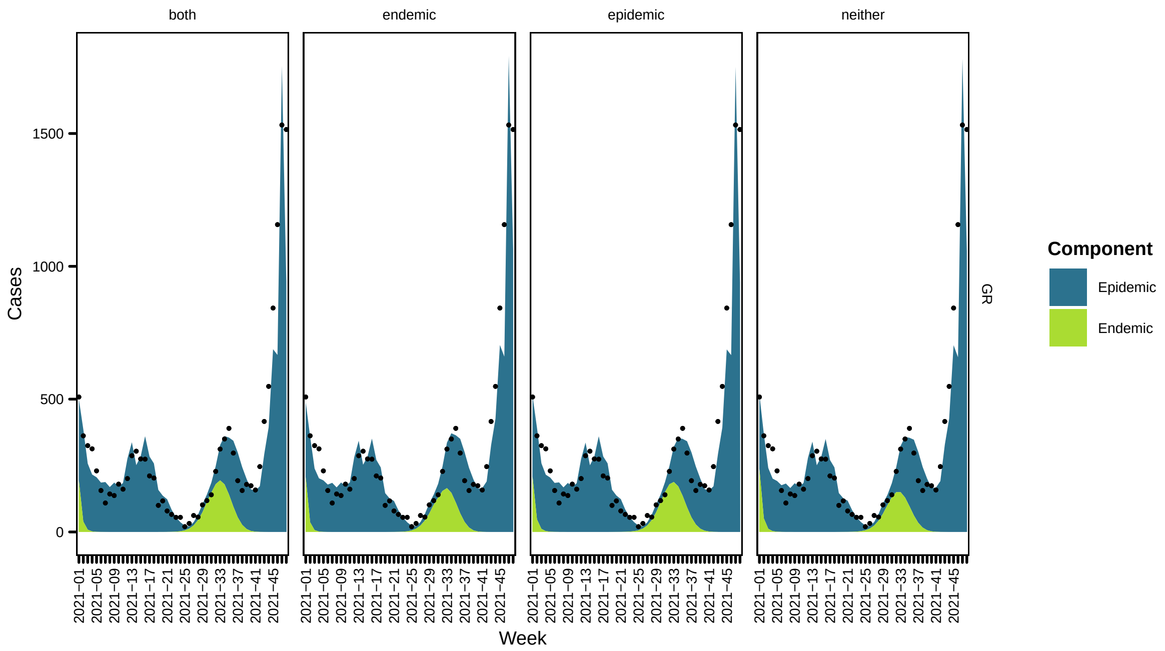
both
endemic
epidemic
neither
Cases
1500
1000
500
0
GR
Week
Component
Epidemic
Endemic

81

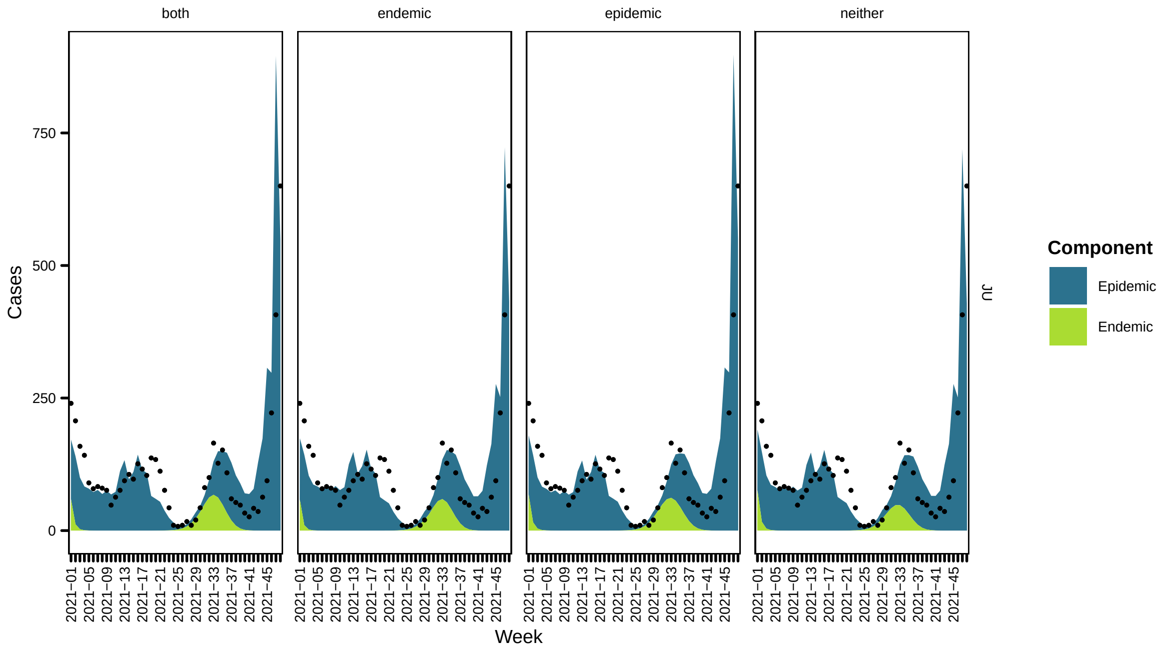
both
endemic
epidemic
neither
Cases
750
500
250
0
JU
Week
Component
Epidemic
Endemic

82

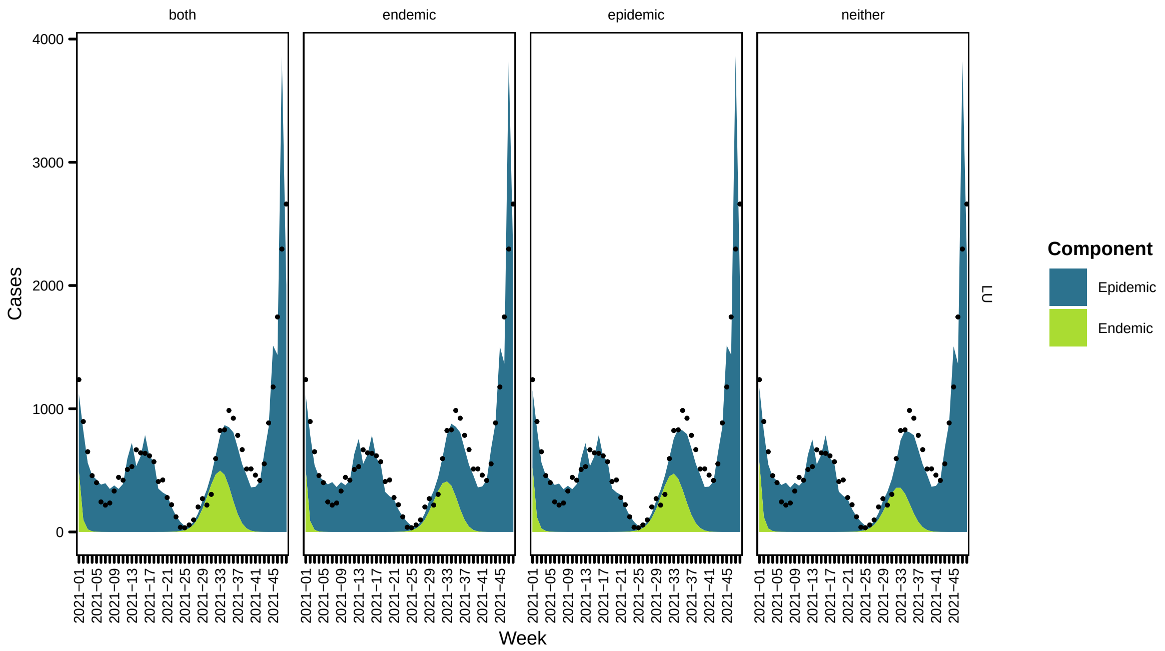
both
endemic
epidemic
neither
Cases
Week
Component
Epidemic
Endemic
LU

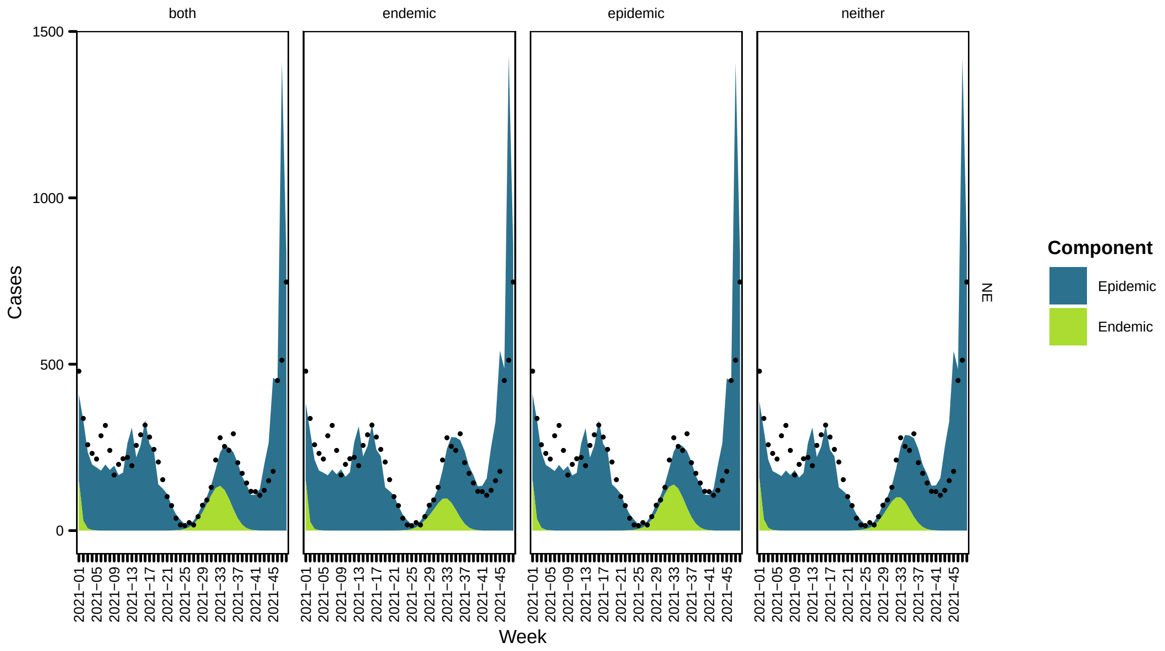
both
endemic
epidemic
neither
Cases
Week
Component
Epidemic
Endemic
NE

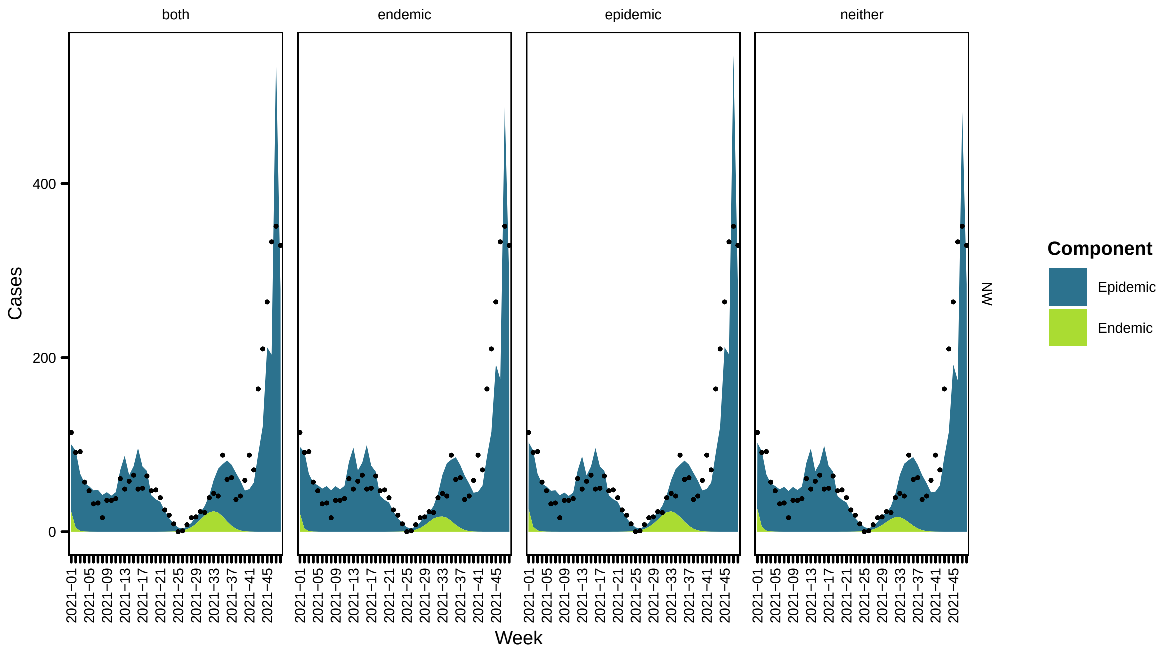
both
endemic
epidemic
neither
Cases
0
200
400
Week
2021–01
2021–05
2021–09
2021–13
2021–17
2021–21
2021–25
2021–29
2021–33
2021–37
2021–41
2021–45
NW
Component
Epidemic
Endemic

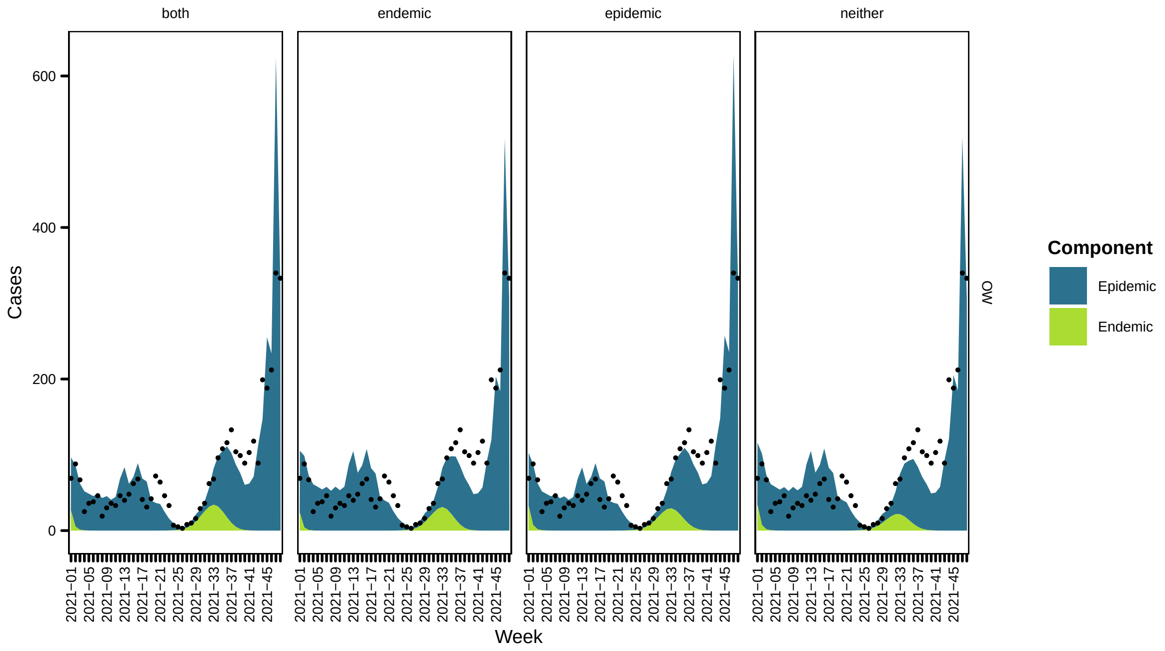
both
endemic
epidemic
neither
Cases
0
200
400
600
Week
2021–01
2021–05
2021–09
2021–13
2021–17
2021–21
2021–25
2021–29
2021–33
2021–37
2021–41
2021–45
OW
Component
Epidemic
Endemic

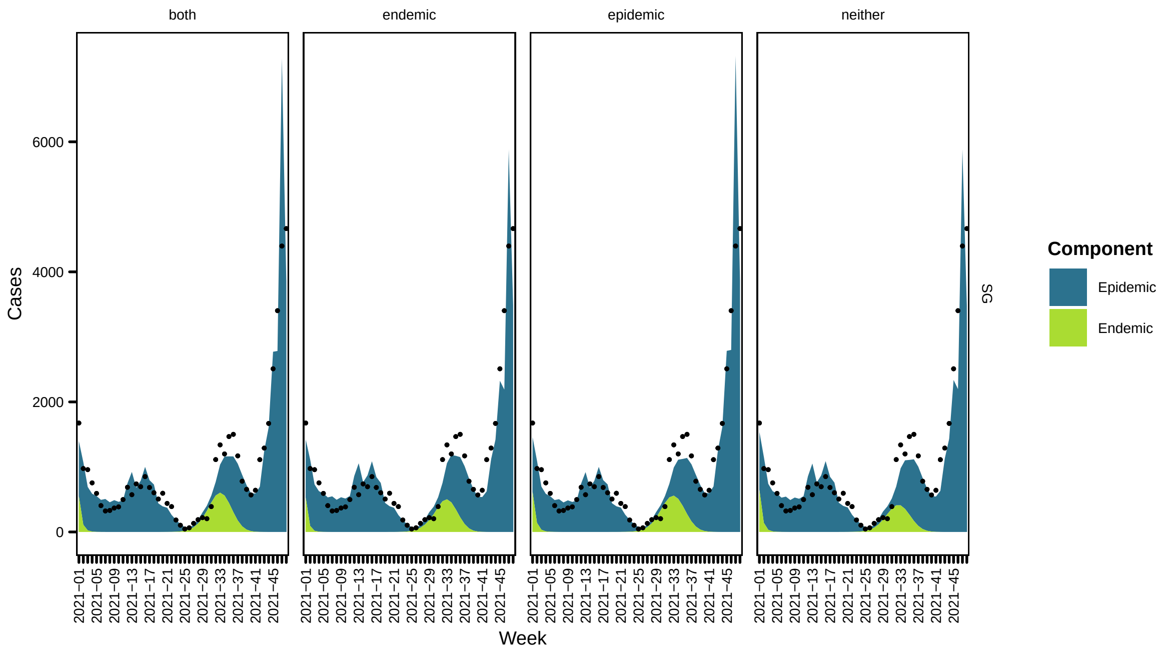
both
endemic
epidemic
neither
Cases
Week
SG
Component
Epidemic
Endemic

87

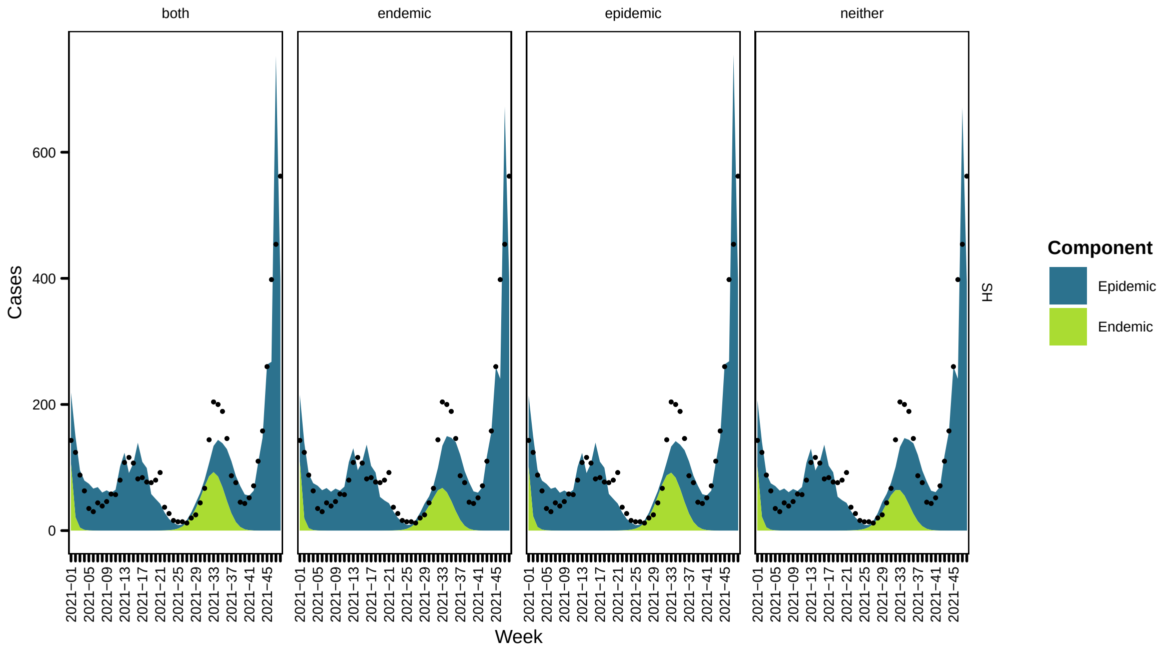
both
endemic
epidemic
neither
Cases
Week
SH
Component
Epidemic
Endemic

88

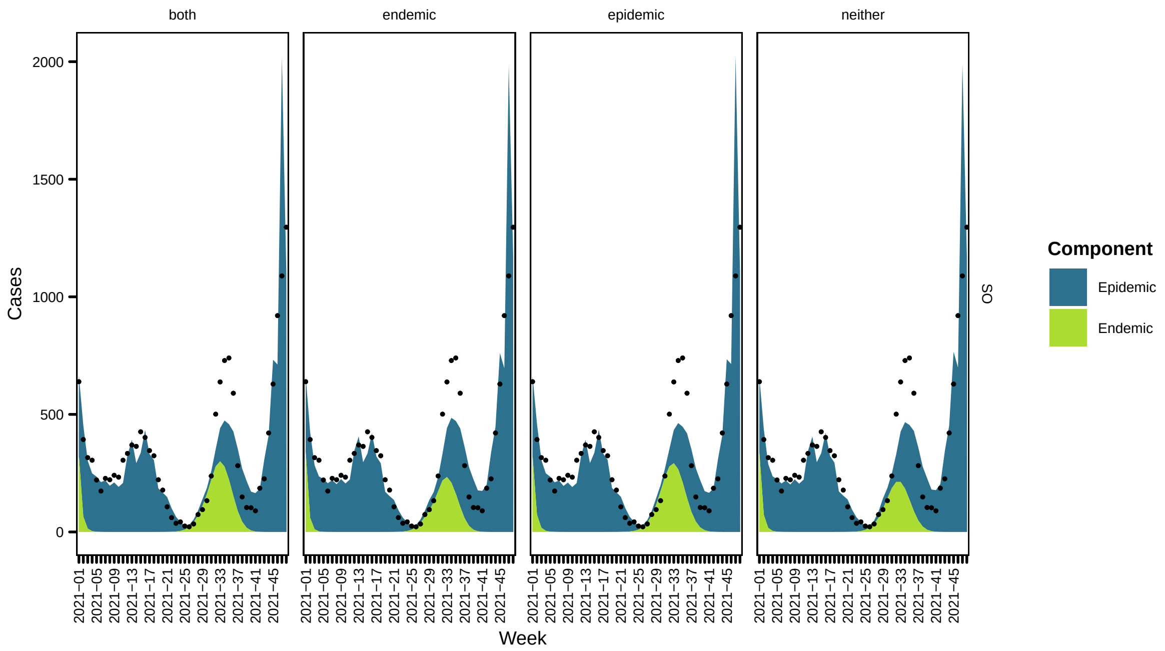
both
endemic
epidemic
neither
Cases
2000
1500
1000
500
0
2021−01
2021−05
2021−09
2021−13
2021−17
2021−21
2021−25
2021−29
2021−33
2021−37
2021−41
2021−45
Week
SO
Component
Epidemic
Endemic

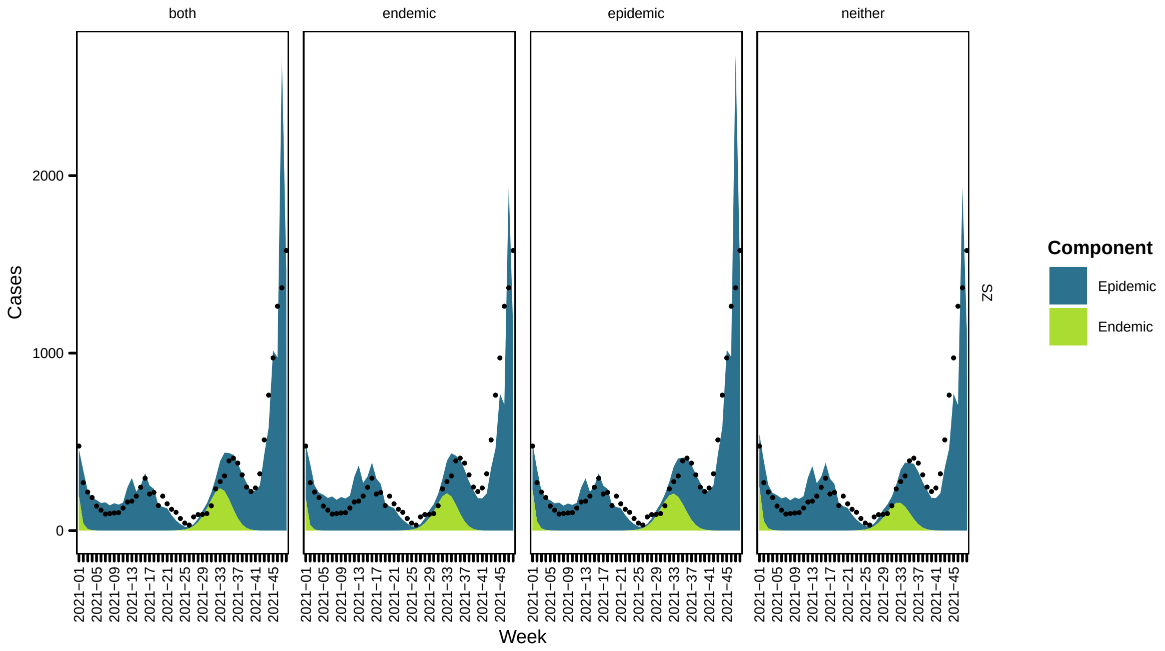
both
endemic
epidemic
neither
Cases
2000
1000
0
2021−01
2021−05
2021−09
2021−13
2021−17
2021−21
2021−25
2021−29
2021−33
2021−37
2021−41
2021−45
Week
SZ
Component
Epidemic
Endemic

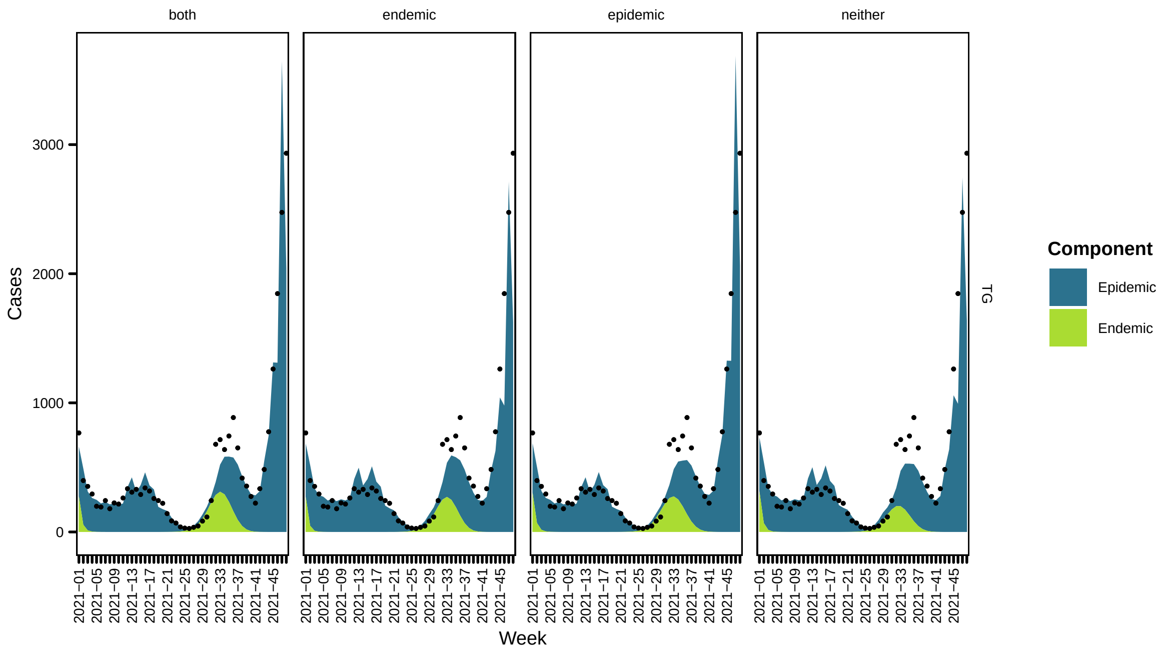
both
endemic
epidemic
neither
Cases
3000
2000
1000
0
Week
TG
Component
Epidemic
Endemic

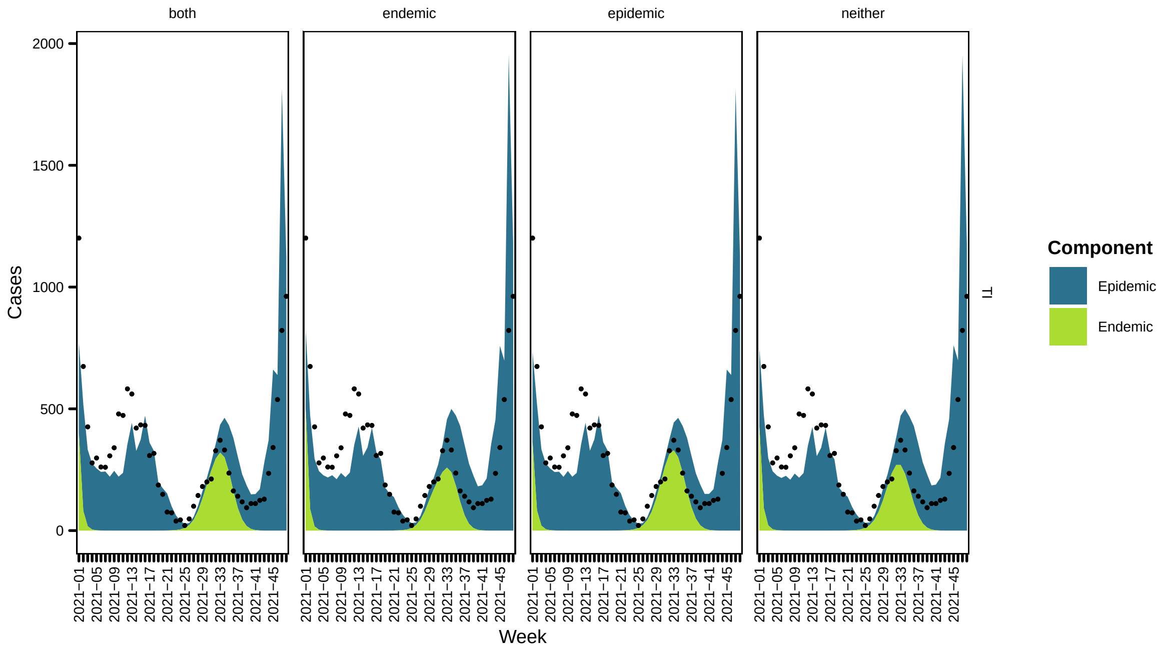
both
endemic
epidemic
neither
Cases
2000
1500
1000
500
0
Week
TI
Component
Epidemic
Endemic

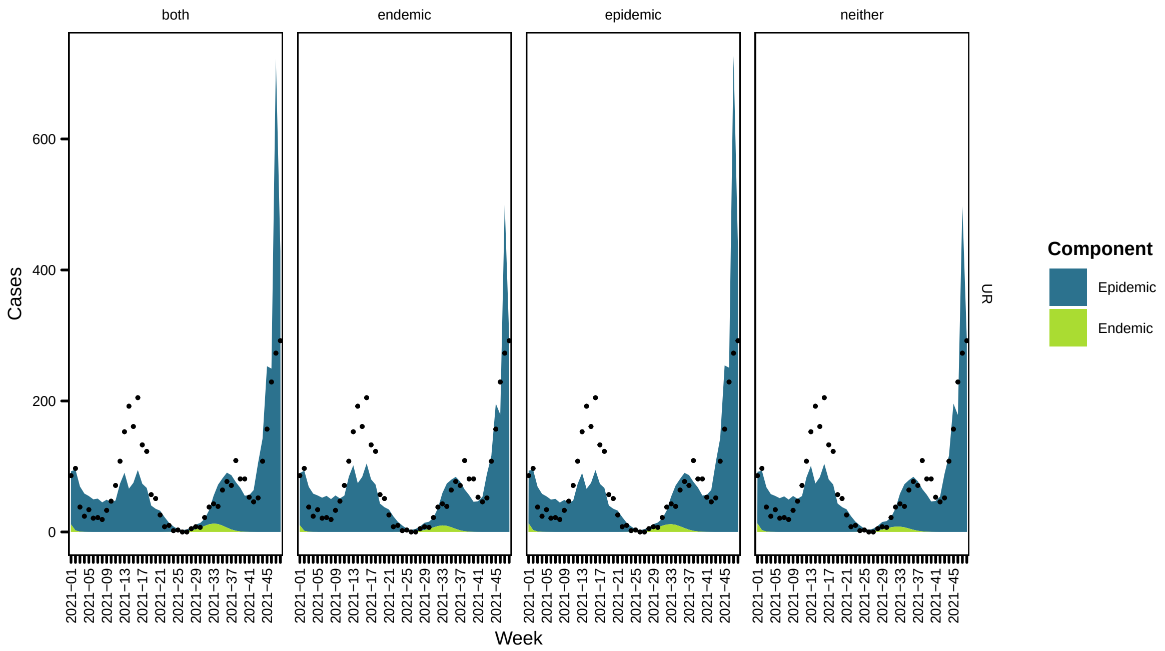
both
endemic
epidemic
neither
Cases
0
200
400
600
Week
UR
Component
Epidemic
Endemic

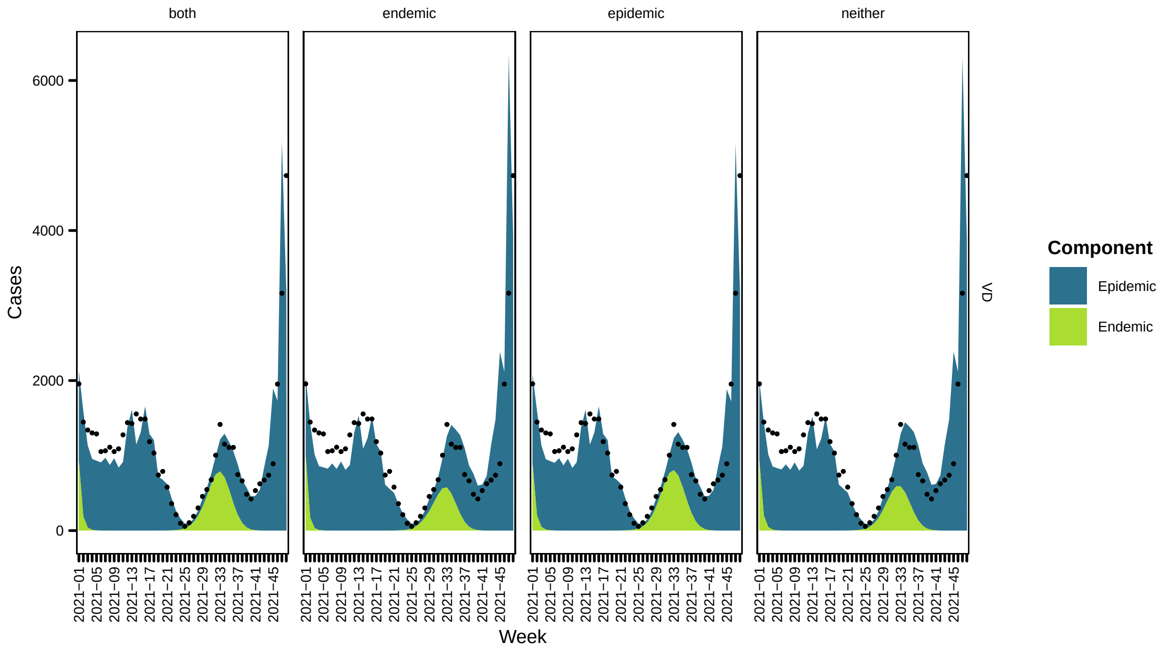
both
endemic
epidemic
neither
Cases
0
2000
4000
6000
Week
VD
Component
Epidemic
Endemic

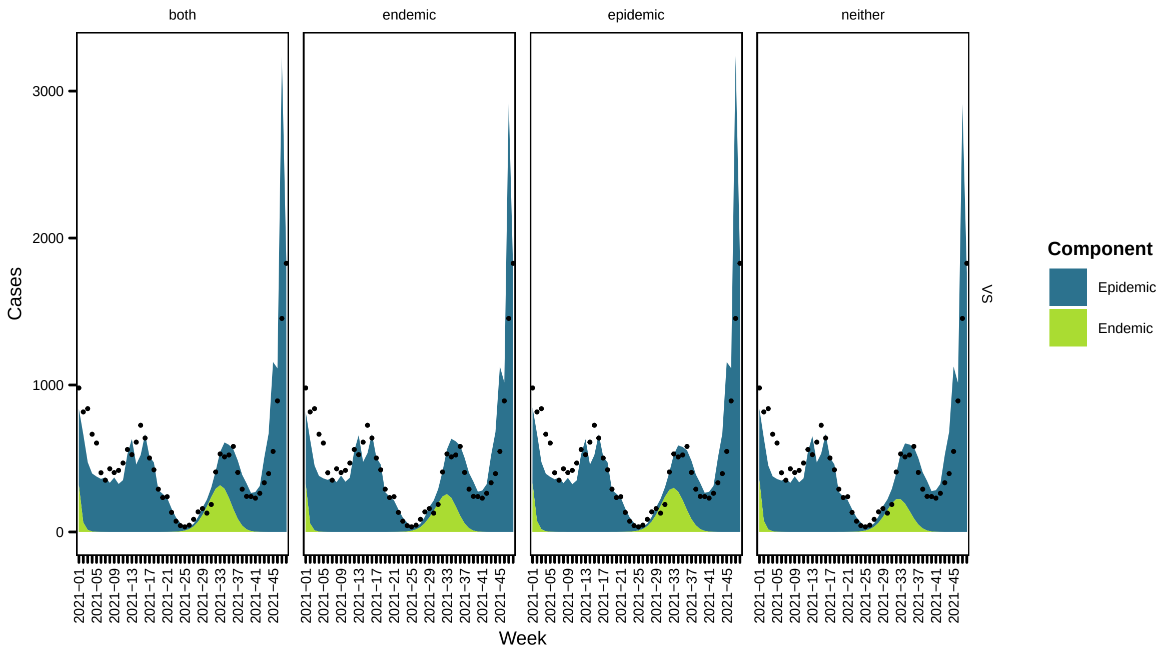
both
endemic
epidemic
neither
Cases
Week
VS
Component
Epidemic
Endemic

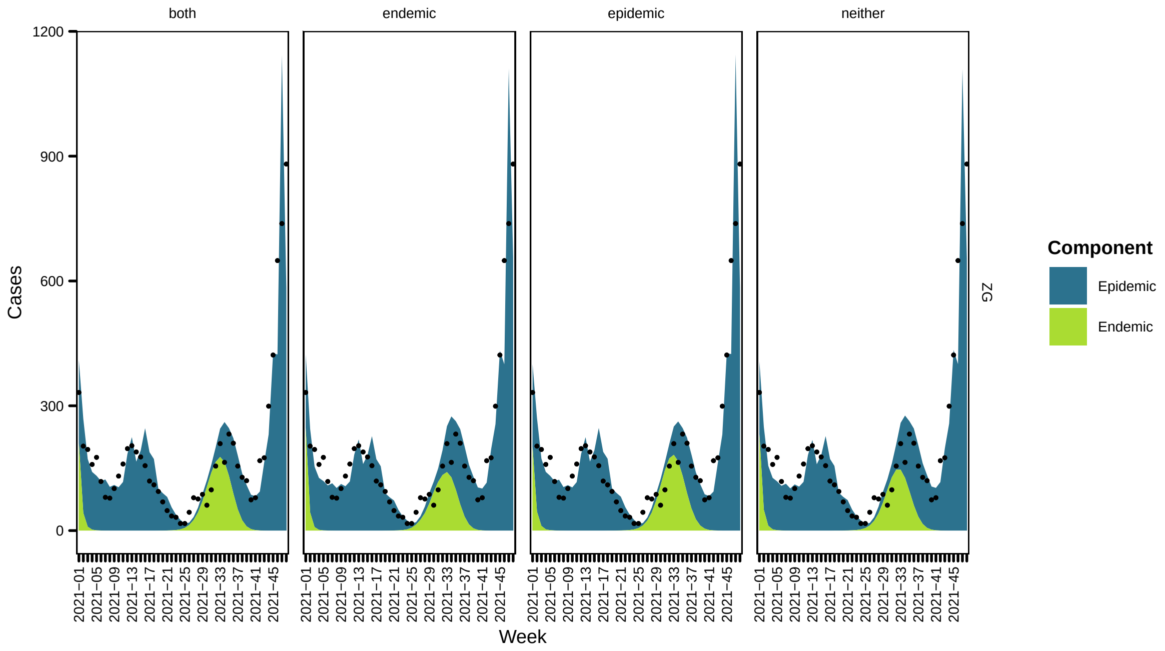
both
endemic
epidemic
neither
Cases
Week
ZG
Component
Epidemic
Endemic

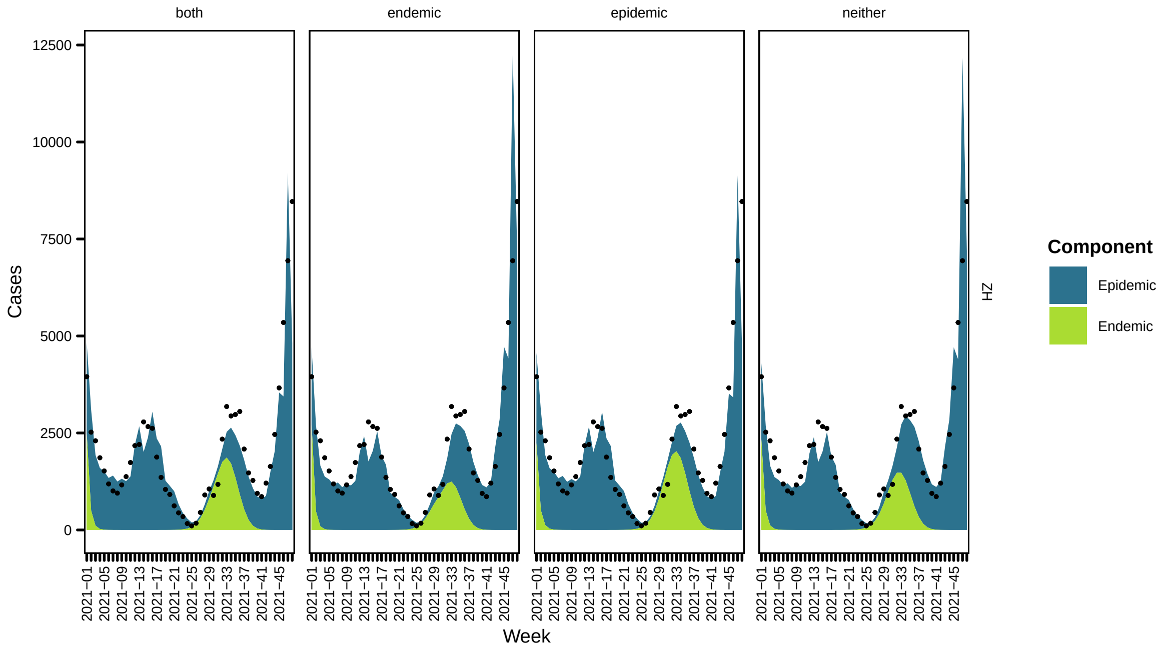


### 3 Scenario analysis

Now we consider the effects of vaccination coverage which is the maximum across regions, $\max_r x_{rt}$

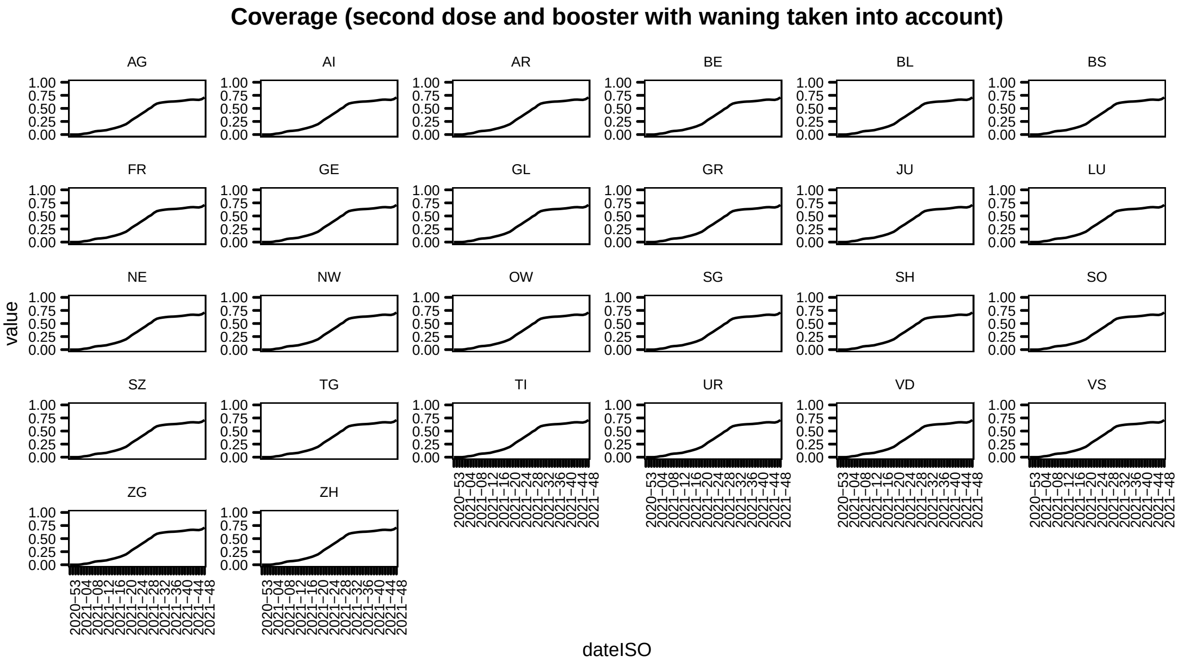

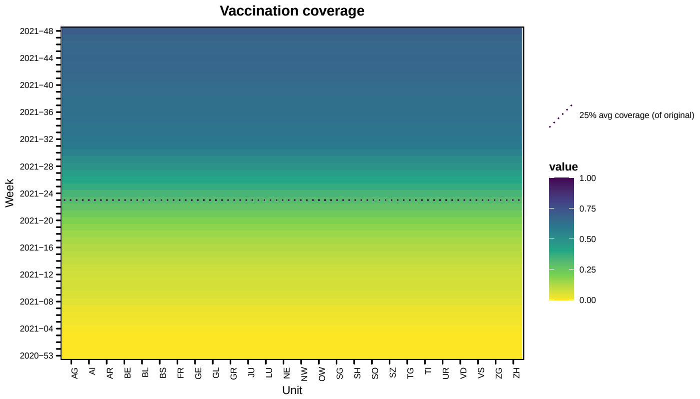


#### 3.1 Predicted cases

Under the assumption that the study period covers the outbreak, we calculate the predicted number of COVID-19 cases under the alternative vaccination coverage scenario scenario. This is using the same methods we have considered in previous work [see 2–4]. The final size of the outbreak is given (Table 1). The weekly predicted cases for the model with vaccination coverage in the endemic component with time-varying weights are presented as this was the best fitting model as measured by DSS. We compare the predictions under the true vaccination coverage (as observed by BAG) and the alternative scenario of maximum coverage. The ratio is the ratio of predicted cases under the alternative vaccination coverage scenario against predicted cases under the observed vaccination coverage scenario.

Below we show predicted cases under the two vaccination coverages (as observed and maximum coverage across regions) and ratio between the two

**Table 1:** Final size estimates based on path forecasts

| | | Original coverage | | | Alternative coverage | | |
|---|---|---|---|---|---|---|---|
| | True | Endemic | Epidemic | Both | Endemic | Epidemic | Both |
| AG | 24208 | 42919 | 23073 | 22502 | 31254 | 8402 | 6924 |
| AI | 1288 | 1925 | 1339 | 1292 | 1385 | 192 | 154 |
| AR | 3687 | 4795 | 3086 | 2988 | 3481 | 537 | 432 |
| BE | 37976 | 58133 | 28781 | 28026 | 42970 | 11296 | 9738 |
| BL | 10983 | 16992 | 9262 | 8999 | 12751 | 3810 | 3410 |
| BS | 7531 | 11523 | 5482 | 5341 | 8757 | 3007 | 2724 |
| FR | 10182 | 21343 | 9765 | 9495 | 15612 | 3234 | 2659 |
| GE | 19107 | 41614 | 21638 | 21308 | 30515 | 9968 | 8516 |
| GL | 2413 | 3541 | 2433 | 2423 | 2407 | 790 | 589 |
| GR | 9279 | 13867 | 7458 | 7290 | 10029 | 2480 | 2012 |
| JU | 2586 | 5798 | 3493 | 3410 | 4168 | 879 | 695 |
| LU | 18050 | 30844 | 16897 | 16602 | 22215 | 5761 | 4676 |
| NE | 4708 | 11888 | 5779 | 5611 | 8727 | 2022 | 1693 |
| NW | 2377 | 3684 | 2027 | 1969 | 2681 | 469 | 381 |
| OW | 2571 | 4049 | 2491 | 2430 | 2898 | 486 | 376 |
| SG | 30828 | 43552 | 27724 | 27038 | 31215 | 7094 | 5608 |
| SH | 3411 | 5272 | 2966 | 2884 | 3897 | 1087 | 938 |
| SO | 9357 | 16315 | 8919 | 8702 | 11941 | 3390 | 2856 |
| SZ | 10431 | 15106 | 10235 | 10068 | 10690 | 2467 | 1902 |
| TG | 16314 | 20355 | 13565 | 13226 | 14548 | 3399 | 2709 |
| TI | 6105 | 16835 | 8746 | 8485 | 12644 | 3754 | 3341 |
| UR | 1860 | 3762 | 2497 | 2410 | 2747 | 404 | 333 |
| VD | 23841 | 56845 | 25277 | 24551 | 42101 | 10648 | 9175 |
| VS | 10502 | 24863 | 13690 | 13375 | 18077 | 4157 | 3441 |
| ZG | 5337 | 9020 | 4908 | 4759 | 6702 | 2055 | 1775 |
| ZH | 55763 | 97121 | 44679 | 42971 | 73768 | 23259 | 20912 |

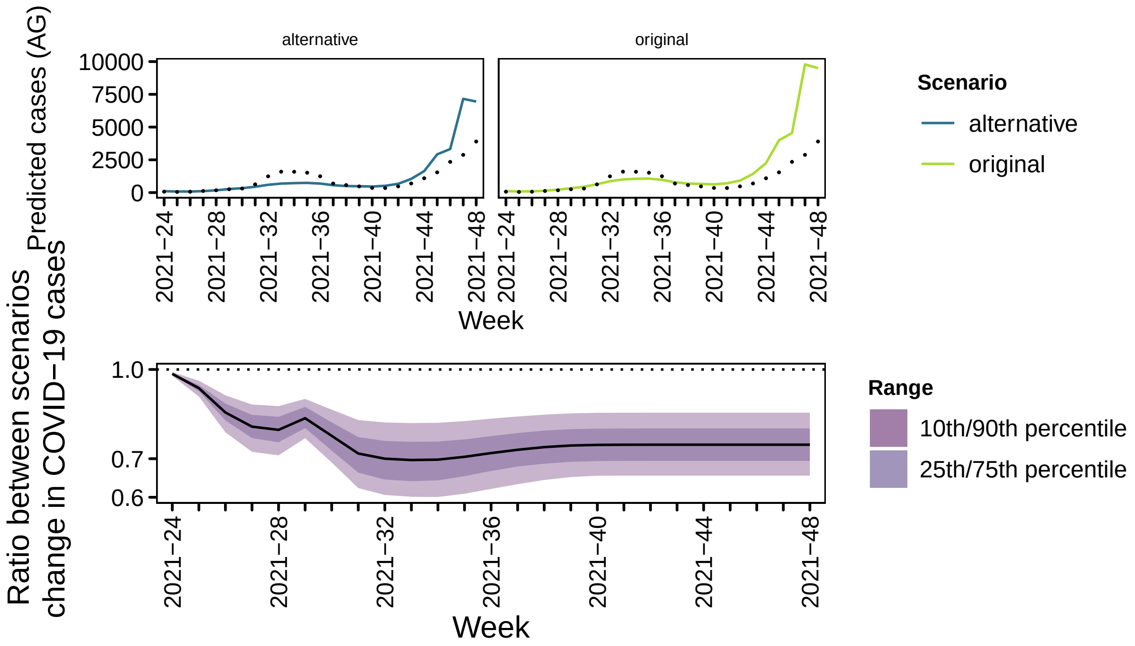
alternative
original
Predicted cases (AG)
10000
7500
5000
2500
0
2021−24
2021−28
2021−32
2021−36
2021−40
2021−44
2021−48
Week
Scenario
alternative
original
Ratio between scenarios
change in COVID−19 cases
1.0
0.7
0.6
Range
10th/90th percentile
25th/75th percentile

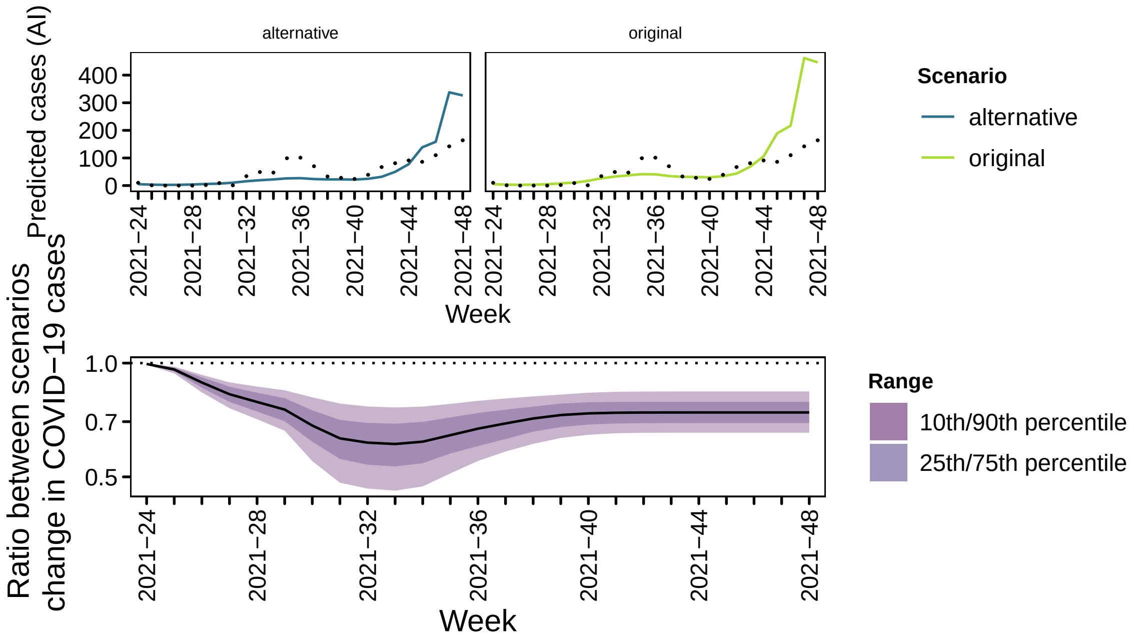
alternative
original
Predicted cases (AI)
400
300
200
100
0
2021−24
2021−28
2021−32
2021−36
2021−40
2021−44
2021−48
Week
Scenario
alternative
original
Ratio between scenarios
change in COVID−19 cases
1.0
0.7
0.5
Range
10th/90th percentile
25th/75th percentile

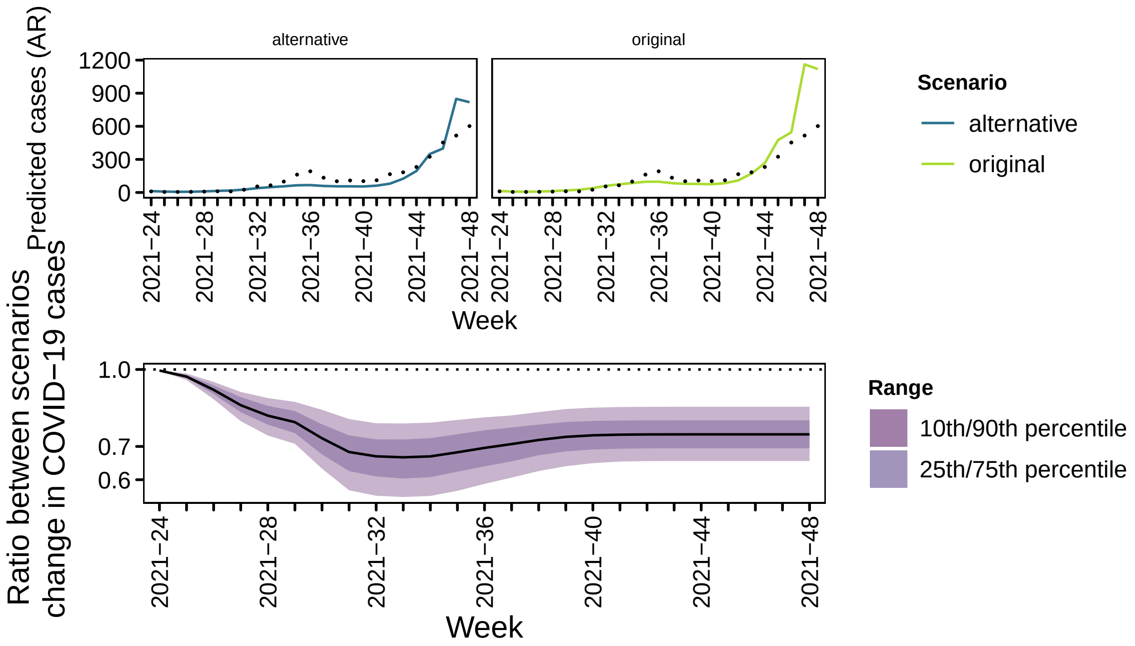
alternative
original
Predicted cases (AR)
1200
900
600
300
0
2021−24
2021−28
2021−32
2021−36
2021−40
2021−44
2021−48
Week
Scenario
alternative
original
Ratio between scenarios change in COVID−19 cases
1.0
0.7
0.6
Range
10th/90th percentile
25th/75th percentile

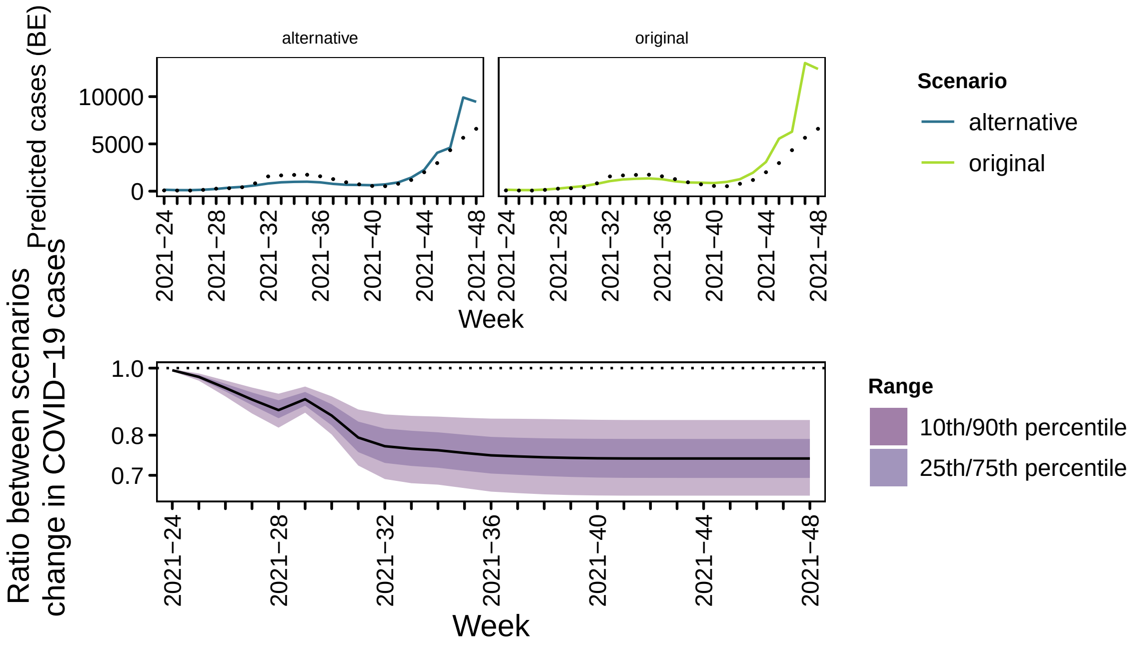
alternative
original
Predicted cases (BE)
10000
5000
0
2021−24
2021−28
2021−32
2021−36
2021−40
2021−44
2021−48
Week
Scenario
alternative
original
Ratio between scenarios change in COVID−19 cases
1.0
0.8
0.7
Range
10th/90th percentile
25th/75th percentile

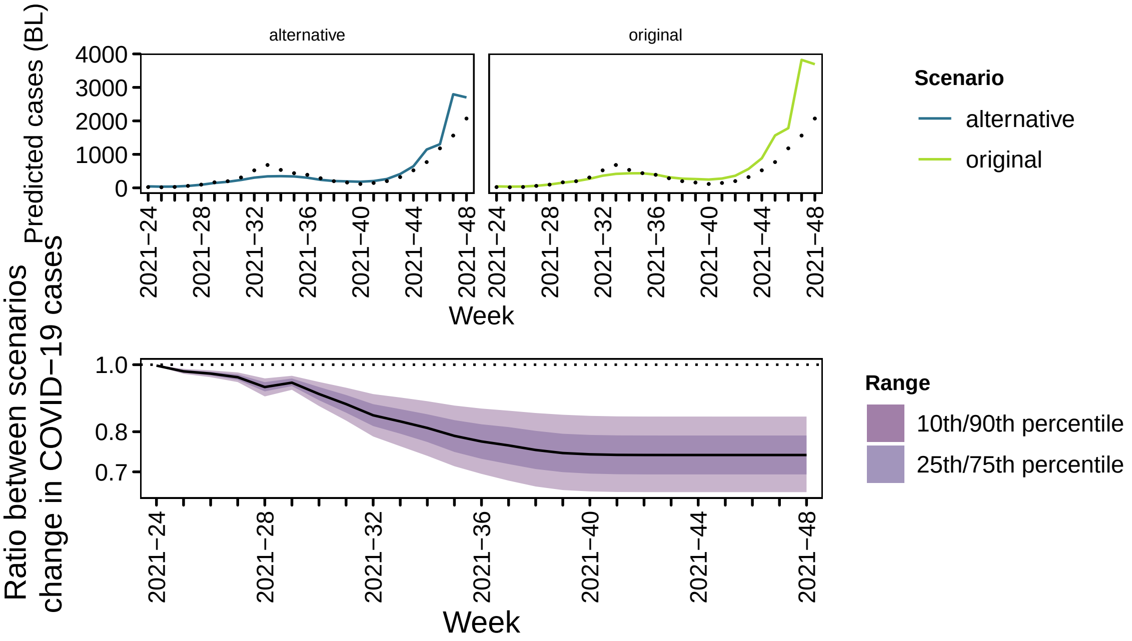
alternative
original
Predicted cases (BL)
4000
3000
2000
1000
0
2021−24
2021−28
2021−32
2021−36
2021−40
2021−44
2021−48
Week
Scenario
alternative
original
Ratio between scenarios
change in COVID−19 cases
1.0
0.8
0.7
Range
10th/90th percentile
25th/75th percentile

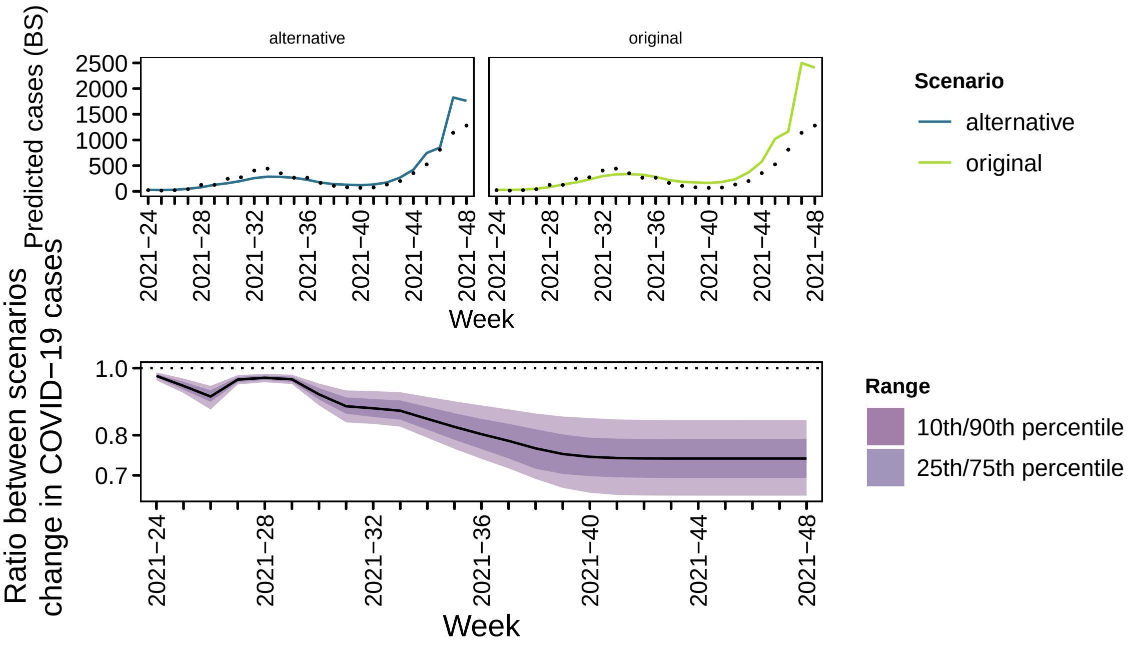
alternative
original
Predicted cases (BS)
2500
2000
1500
1000
500
0
2021−24
2021−28
2021−32
2021−36
2021−40
2021−44
2021−48
Week
Scenario
alternative
original
Ratio between scenarios
change in COVID−19 cases
1.0
0.8
0.7
Range
10th/90th percentile
25th/75th percentile

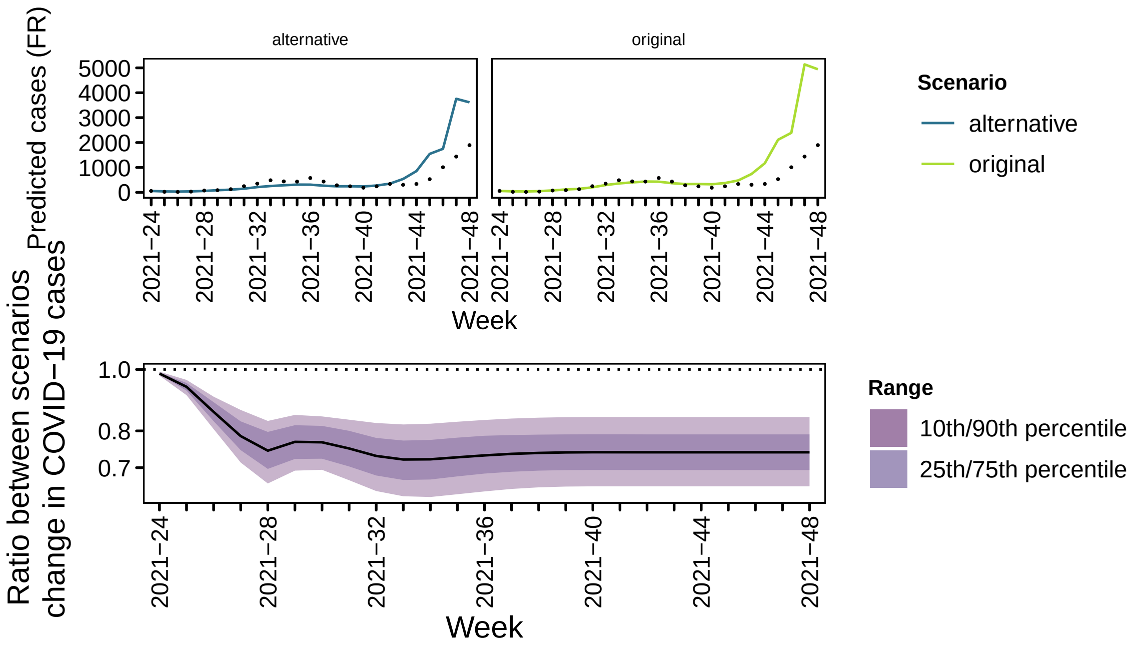

alternative
original
Predicted cases (FR)
5000
4000
3000
2000
1000
0
2021−24
2021−28
2021−32
2021−36
2021−40
2021−44
2021−48
Week
Scenario
alternative
original
Ratio between scenarios
change in COVID−19 cases
1.0
0.8
0.7
Week
Range
10th/90th percentile
25th/75th percentile


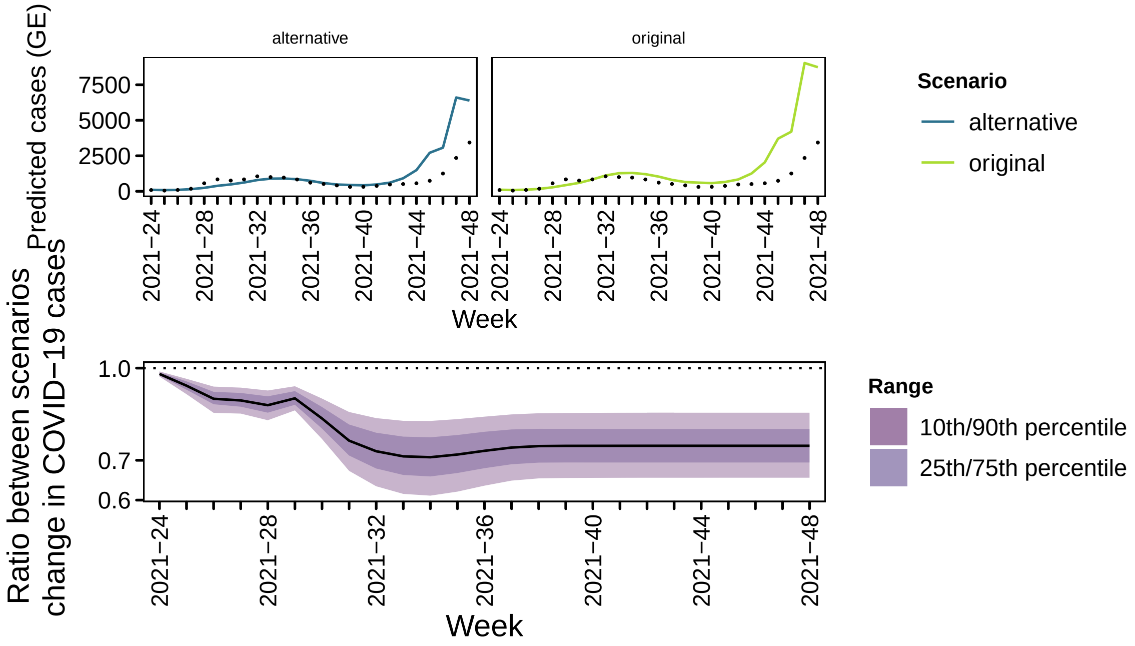

alternative
original
Predicted cases (GE)
7500
5000
2500
0
2021−24
2021−28
2021−32
2021−36
2021−40
2021−44
2021−48
Week
Scenario
alternative
original
Ratio between scenarios
change in COVID−19 cases
1.0
0.7
0.6
Week
Range
10th/90th percentile
25th/75th percentile

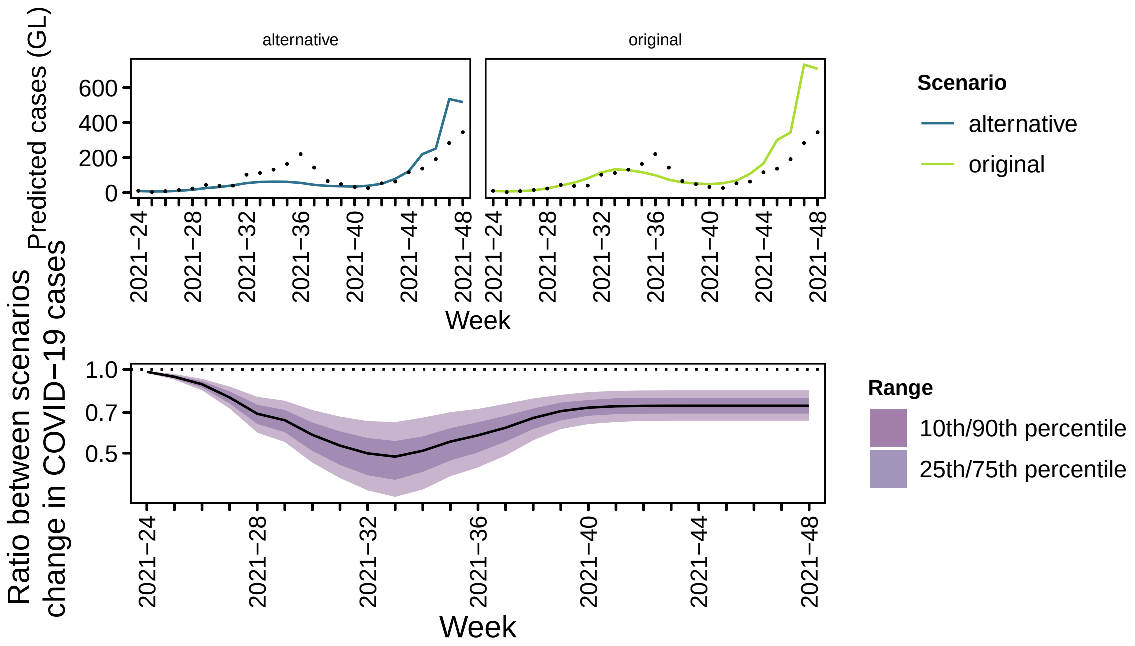
alternative
original
Predicted cases (GL)
600
400
200
0
2021−24
2021−28
2021−32
2021−36
2021−40
2021−44
2021−48
Week
Scenario
alternative
original
Ratio between scenarios
change in COVID−19 cases
1.0
0.7
0.5
Range
10th/90th percentile
25th/75th percentile

123

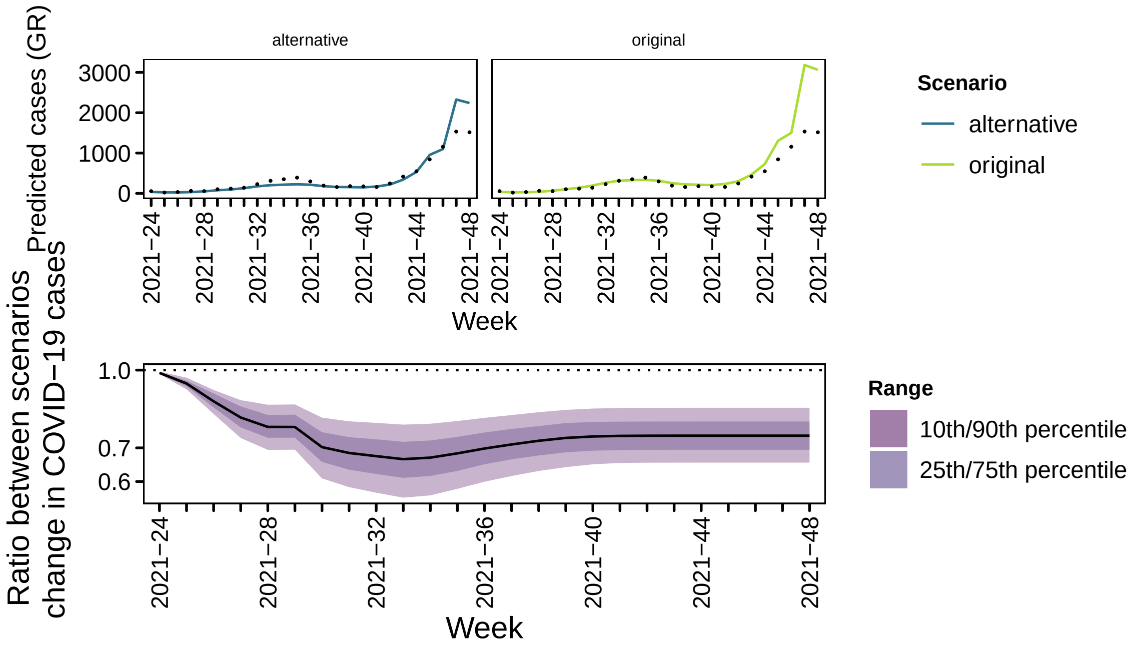
alternative
original
Predicted cases (GR)
3000
2000
1000
0
2021−24
2021−28
2021−32
2021−36
2021−40
2021−44
2021−48
Week
Scenario
alternative
original
Ratio between scenarios
change in COVID−19 cases
1.0
0.7
0.6
Range
10th/90th percentile
25th/75th percentile

124

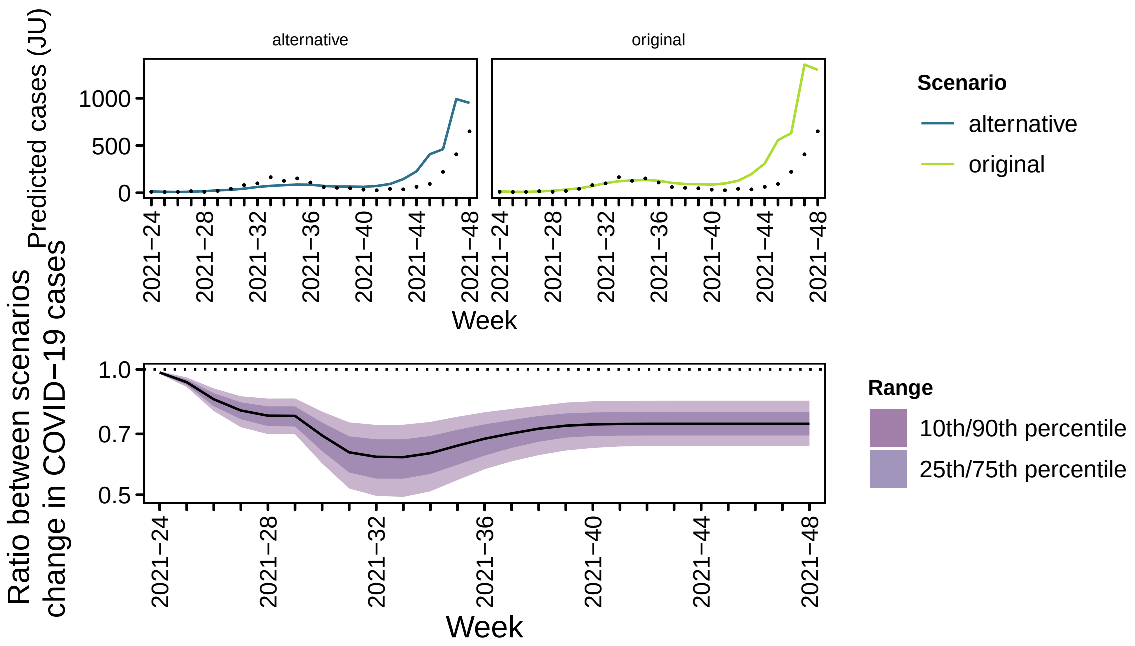
alternative
original
Predicted cases (JU)
1000
500
0
2021−24
2021−28
2021−32
2021−36
2021−40
2021−44
2021−48
Week
Scenario
alternative
original
Ratio between scenarios
change in COVID−19 cases
1.0
0.7
0.5
Range
10th/90th percentile
25th/75th percentile

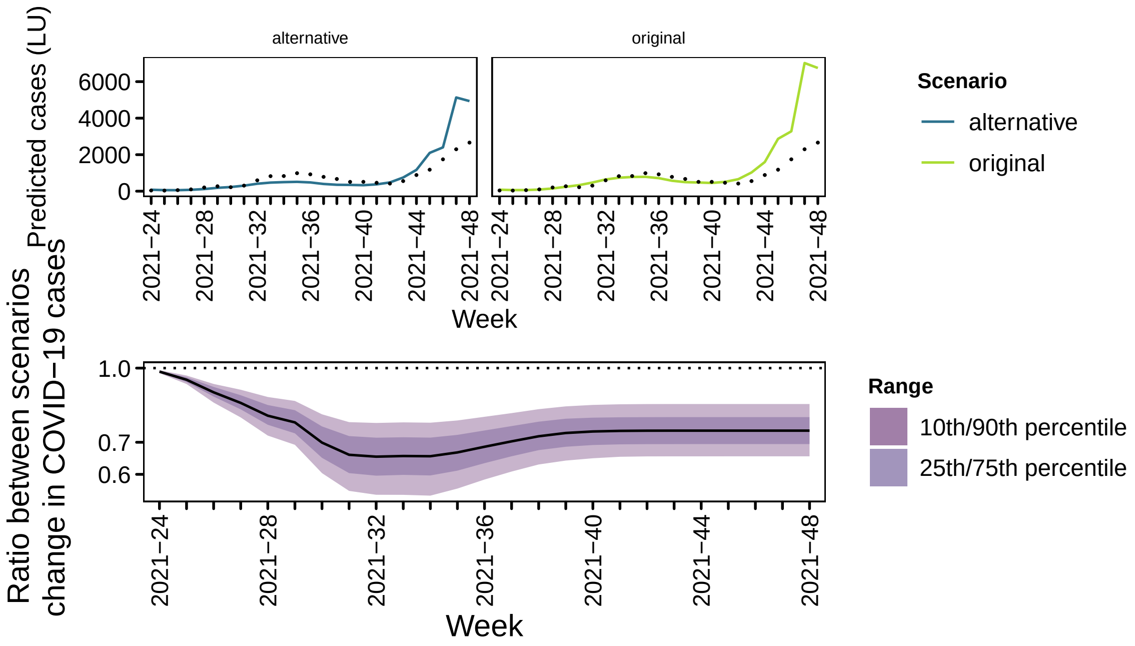
alternative
original
Predicted cases (LU)
6000
4000
2000
0
2021−24
2021−28
2021−32
2021−36
2021−40
2021−44
2021−48
Week
Scenario
alternative
original
Ratio between scenarios
change in COVID−19 cases
1.0
0.7
0.6
Range
10th/90th percentile
25th/75th percentile

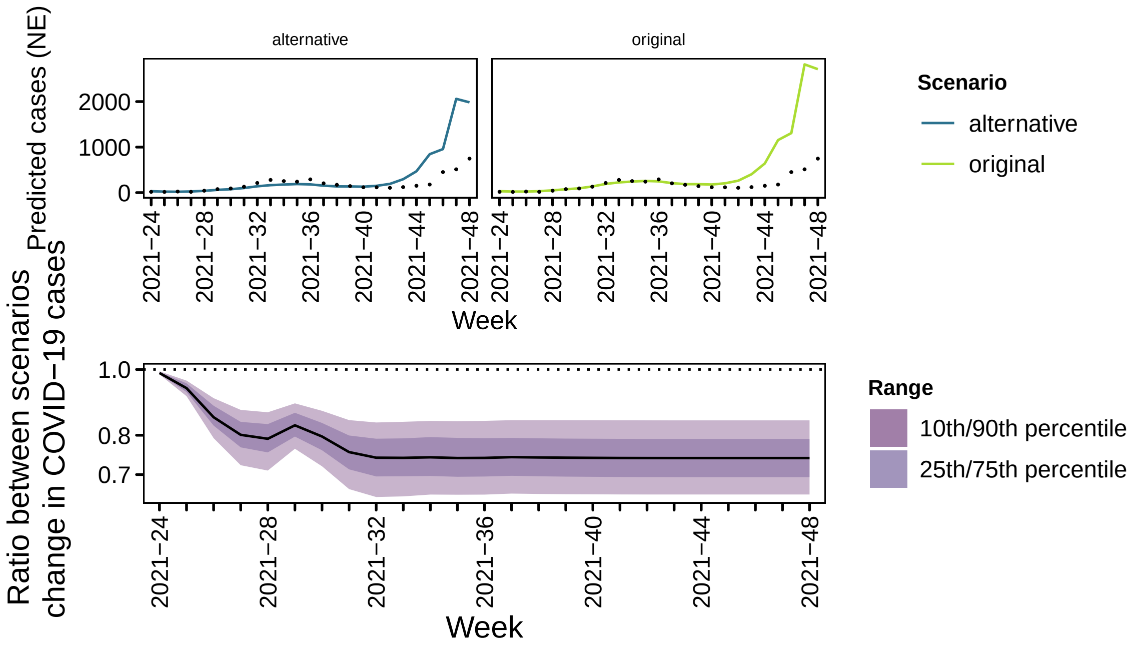
alternative
original
Predicted cases (NE)
2000
1000
0
2021−24
2021−28
2021−32
2021−36
2021−40
2021−44
2021−48
Week
Scenario
alternative
original
Ratio between scenarios
change in COVID−19 cases
1.0
0.8
0.7
Week
Range
10th/90th percentile
25th/75th percentile

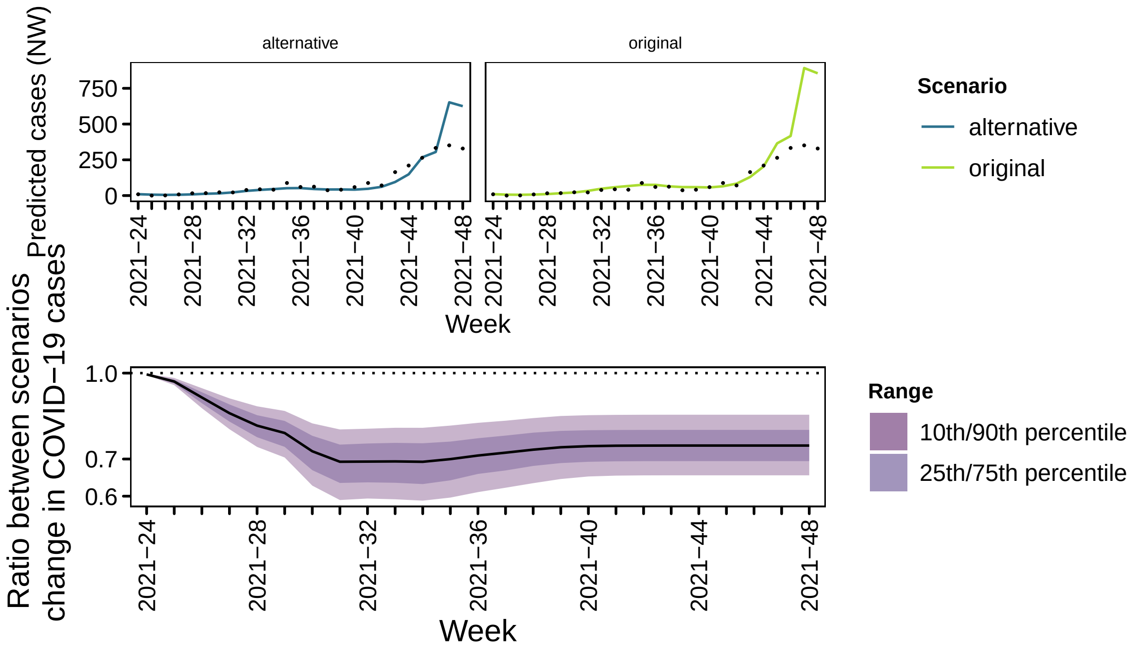
alternative
original
Predicted cases (NW)
750
500
250
0
2021−24
2021−28
2021−32
2021−36
2021−40
2021−44
2021−48
Week
Scenario
alternative
original
Ratio between scenarios
change in COVID−19 cases
1.0
0.7
0.6
Week
Range
10th/90th percentile
25th/75th percentile

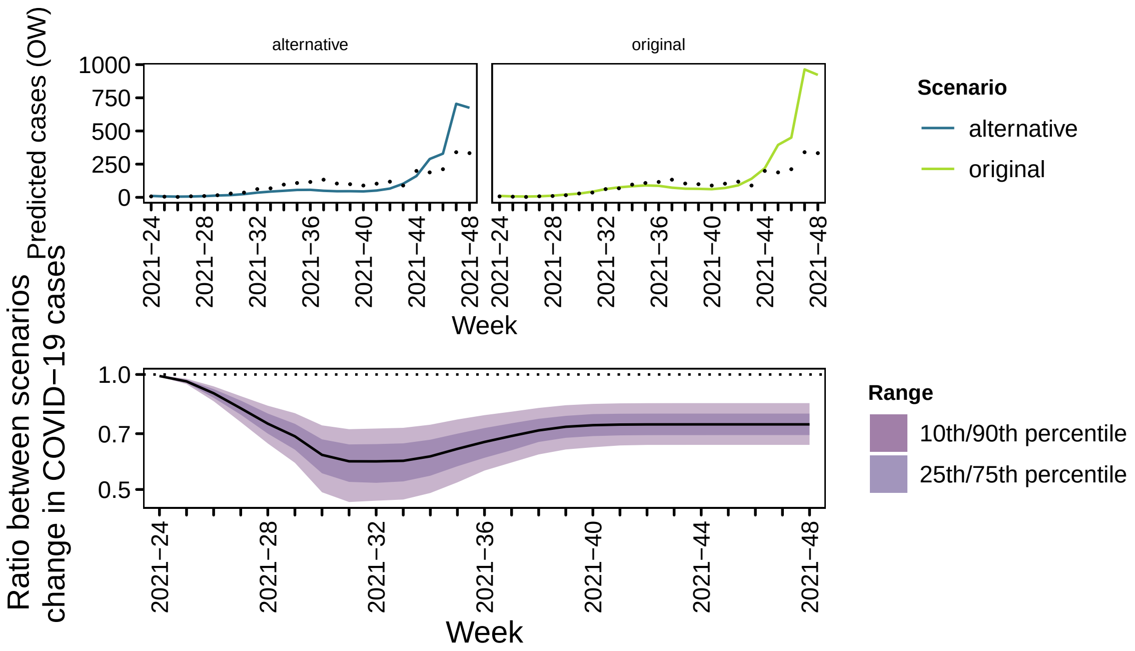
Predicted cases (OW)
alternative
original
Week
Scenario
alternative
original
Ratio between scenarios change in COVID−19 cases
Week
Range
10th/90th percentile
25th/75th percentile

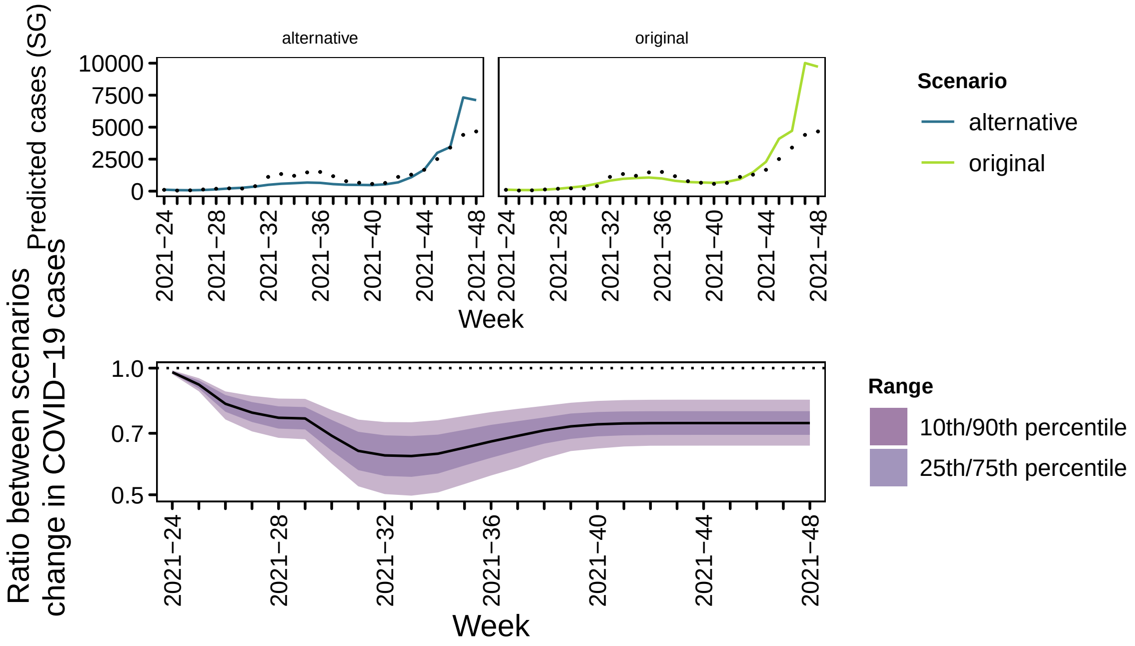
Predicted cases (SG)
alternative
original
Week
Scenario
alternative
original
Ratio between scenarios change in COVID−19 cases
Week
Range
10th/90th percentile
25th/75th percentile

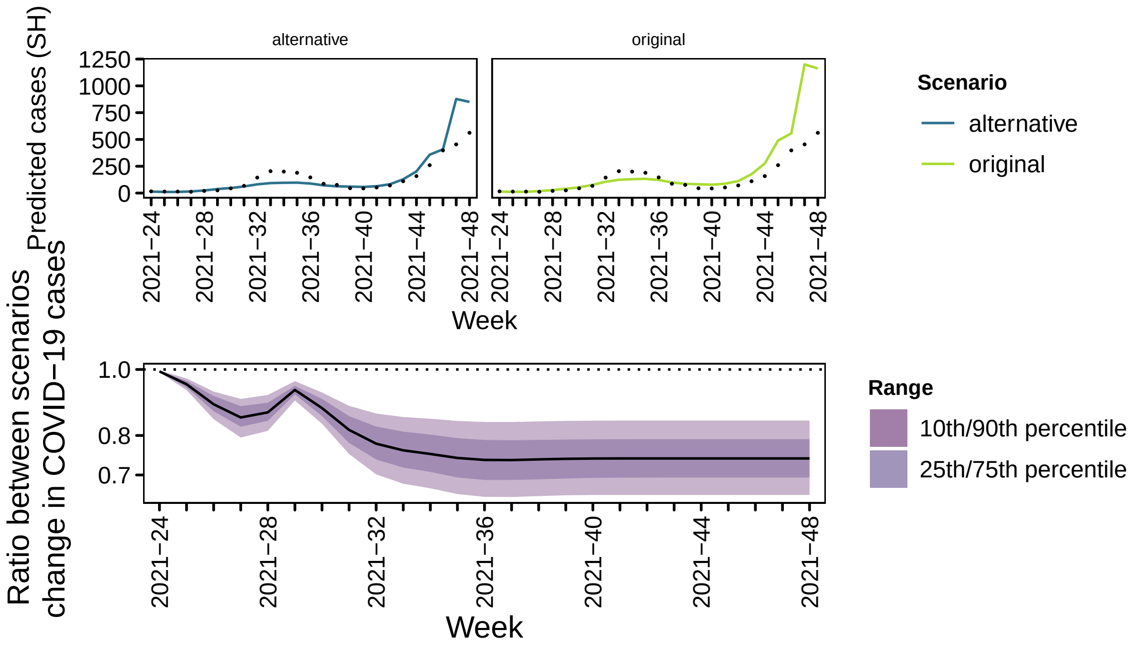
alternative
original
Predicted cases (SH)
1250
1000
750
500
250
0
2021−24
2021−28
2021−32
2021−36
2021−40
2021−44
2021−48
Week
Scenario
alternative
original
Ratio between scenarios
change in COVID−19 cases
1.0
0.8
0.7
Week
Range
10th/90th percentile
25th/75th percentile

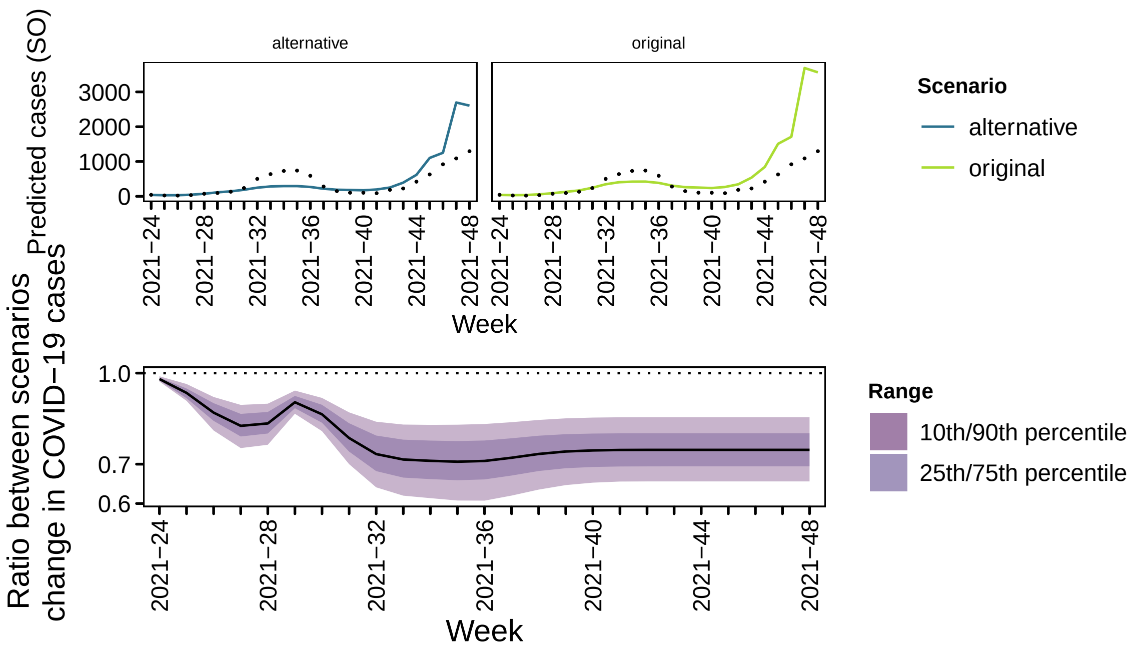
alternative
original
Predicted cases (SO)
3000
2000
1000
0
2021−24
2021−28
2021−32
2021−36
2021−40
2021−44
2021−48
Week
Scenario
alternative
original
Ratio between scenarios
change in COVID−19 cases
1.0
0.7
0.6
Week
Range
10th/90th percentile
25th/75th percentile

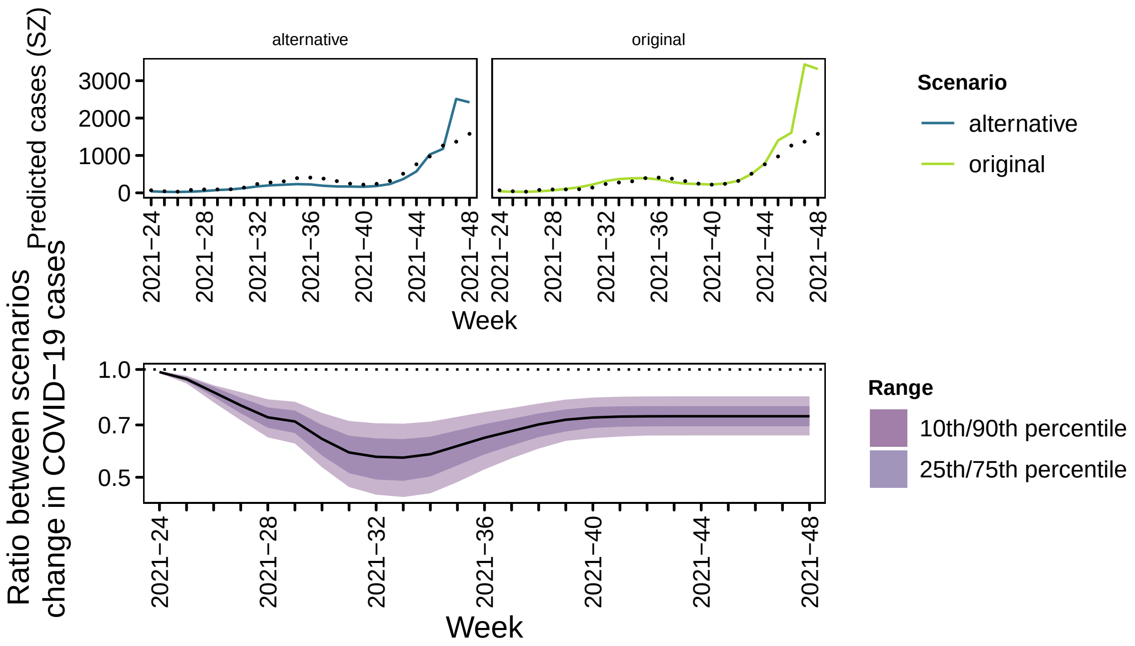
Predicted cases (SZ)
alternative
original
3000
2000
1000
0
2021−24
2021−28
2021−32
2021−36
2021−40
2021−44
2021−48
Week
Scenario
alternative
original
Ratio between scenarios change in COVID−19 cases
1.0
0.7
0.5
Week
Range
10th/90th percentile
25th/75th percentile

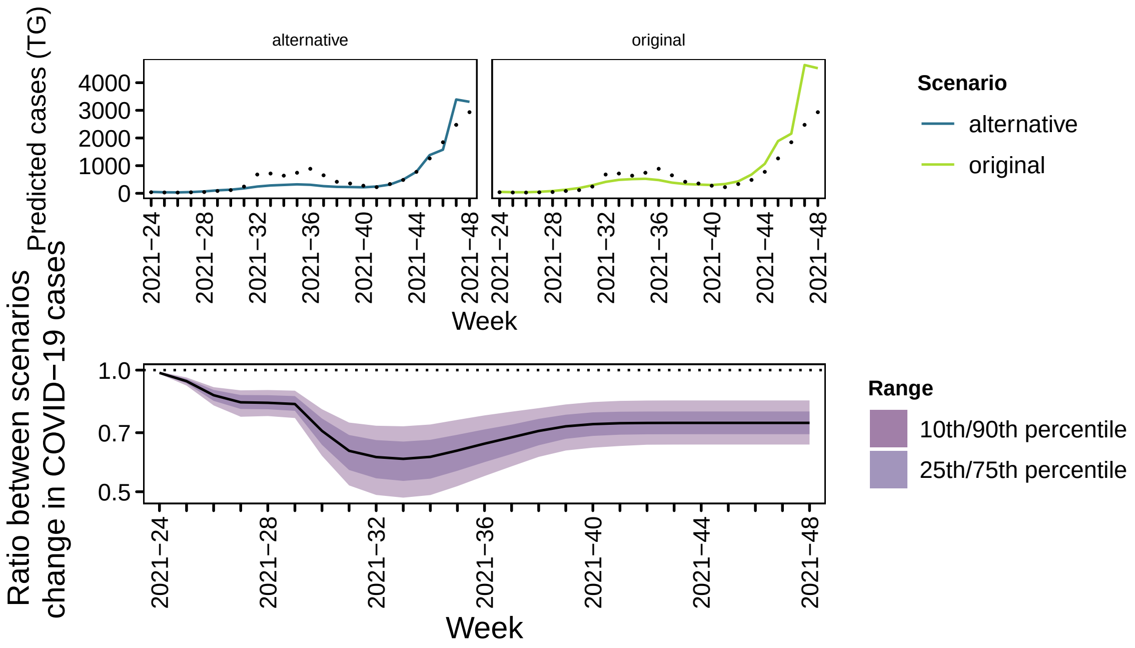
Predicted cases (TG)
alternative
original
4000
3000
2000
1000
0
2021−24
2021−28
2021−32
2021−36
2021−40
2021−44
2021−48
Week
Scenario
alternative
original
Ratio between scenarios change in COVID−19 cases
1.0
0.7
0.5
Week
Range
10th/90th percentile
25th/75th percentile

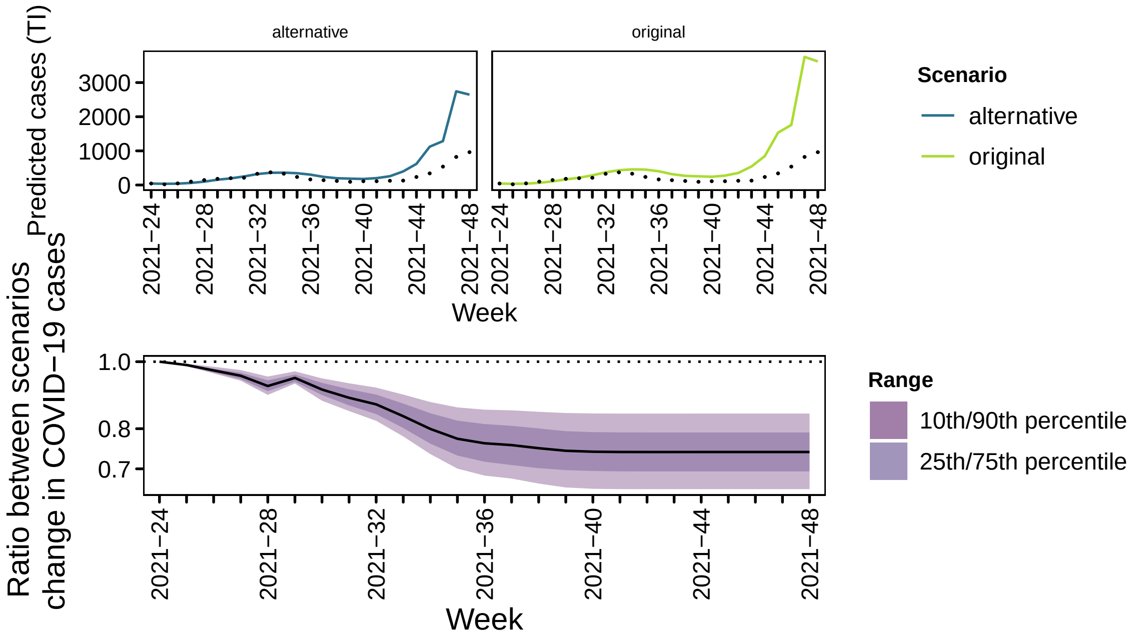
alternative
original
Predicted cases (TI)
3000
2000
1000
0
2021−24
2021−28
2021−32
2021−36
2021−40
2021−44
2021−48
Week
Scenario
alternative
original
Ratio between scenarios
change in COVID−19 cases
1.0
0.8
0.7
Range
10th/90th percentile
25th/75th percentile

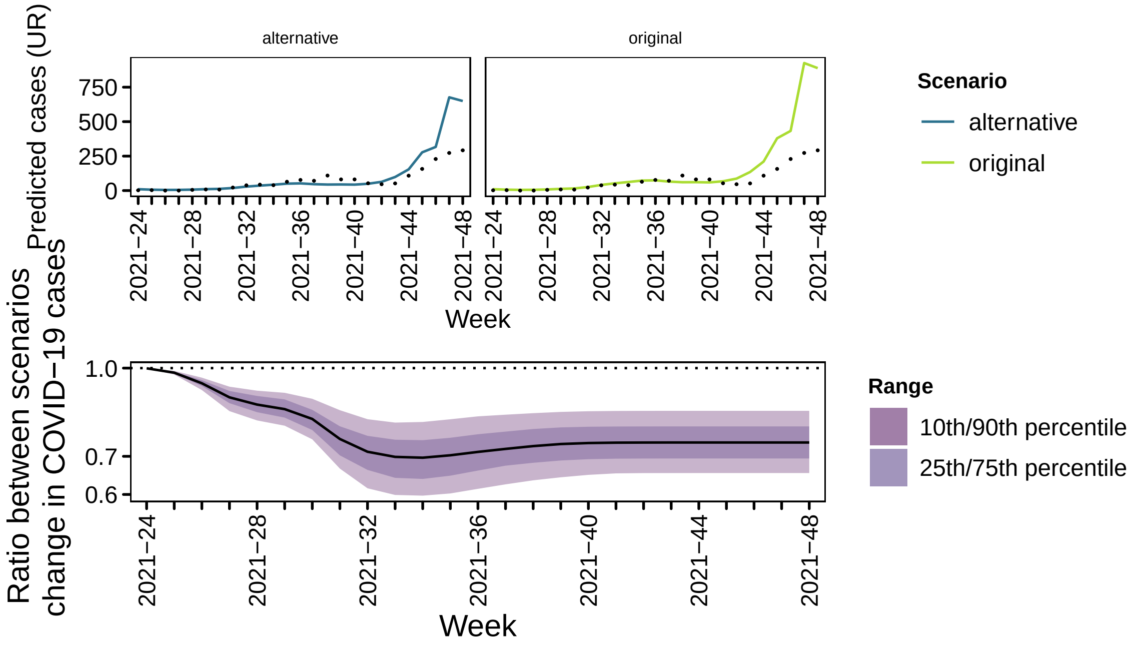
alternative
original
Predicted cases (UR)
750
500
250
0
2021−24
2021−28
2021−32
2021−36
2021−40
2021−44
2021−48
Week
Scenario
alternative
original
Ratio between scenarios
change in COVID−19 cases
1.0
0.7
0.6
Range
10th/90th percentile
25th/75th percentile

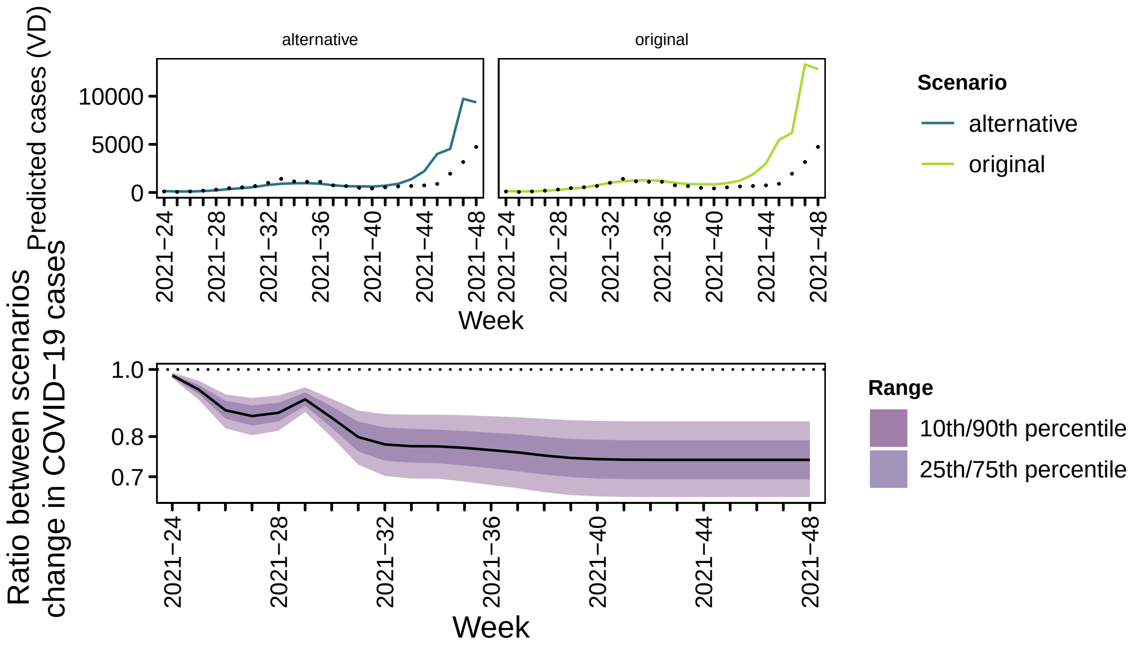
Predicted cases (VD)
alternative
original
10000
5000
0
2021−24
2021−28
2021−32
2021−36
2021−40
2021−44
2021−48
Week
Scenario
alternative
original
Ratio between scenarios change in COVID−19 cases
1.0
0.8
0.7
Week
Range
10th/90th percentile
25th/75th percentile

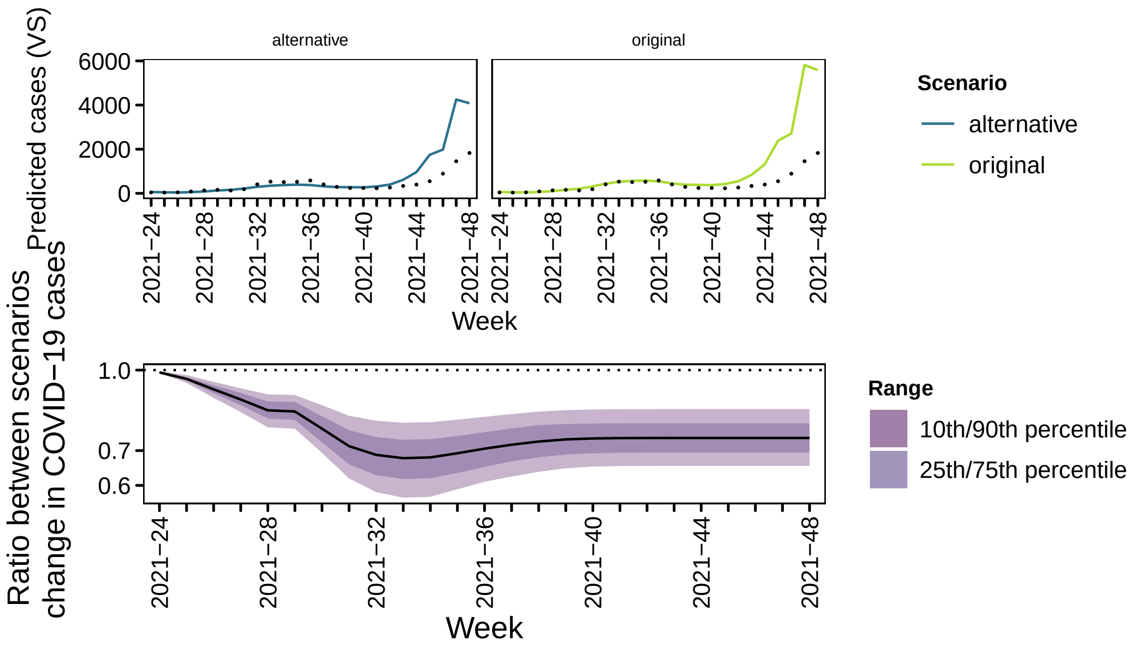
Predicted cases (VS)
alternative
original
6000
4000
2000
0
2021−24
2021−28
2021−32
2021−36
2021−40
2021−44
2021−48
Week
Scenario
alternative
original
Ratio between scenarios change in COVID−19 cases
1.0
0.7
0.6
Week
Range
10th/90th percentile
25th/75th percentile

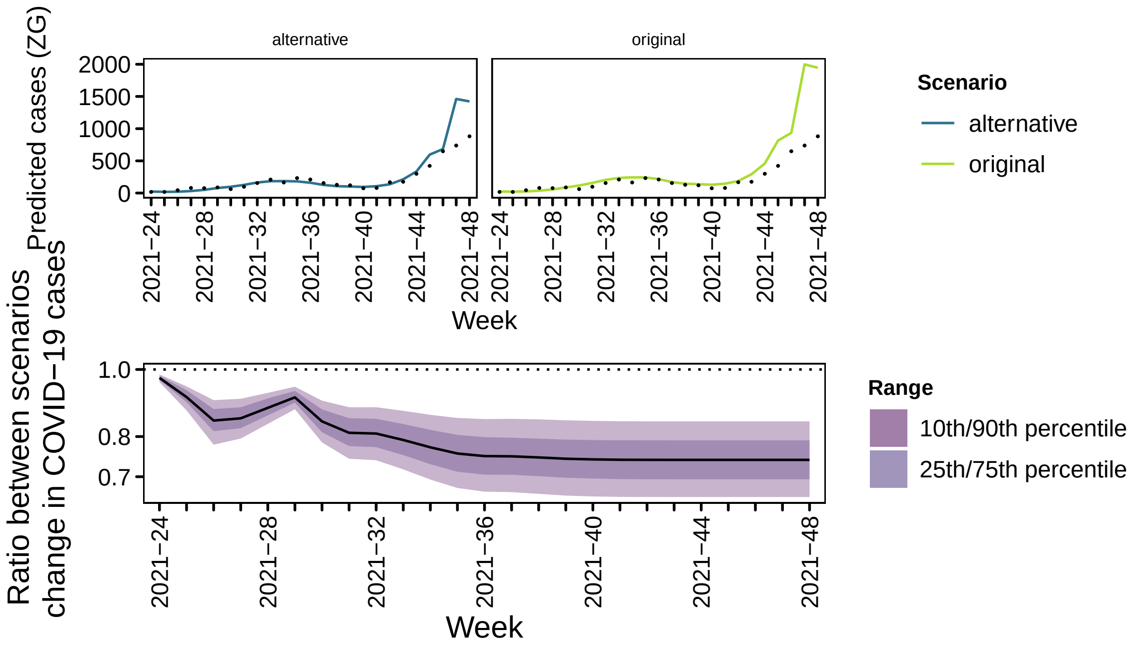
alternative
original
Predicted cases (ZG)
2000
1500
1000
500
0
2021−24
2021−28
2021−32
2021−36
2021−40
2021−44
2021−48
Week
Scenario
alternative
original
Ratio between scenarios change in COVID−19 cases
1.0
0.8
0.7
Range
10th/90th percentile
25th/75th percentile

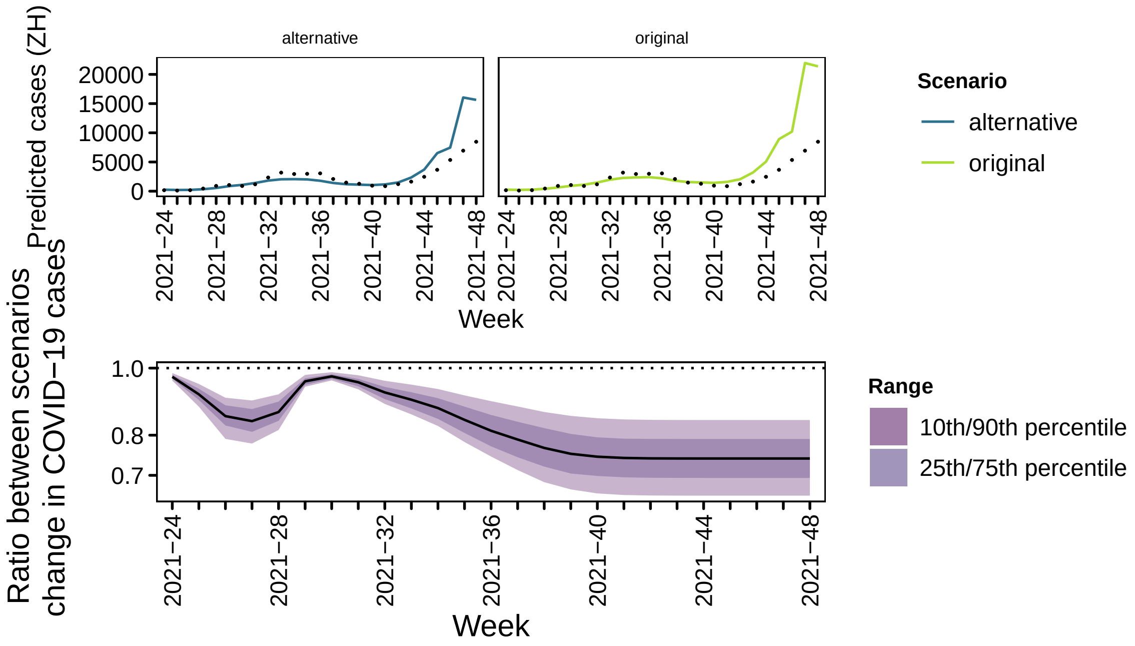
alternative
original
Predicted cases (ZH)
20000
15000
10000
5000
0
2021−24
2021−28
2021−32
2021−36
2021−40
2021−44
2021−48
Week
Scenario
alternative
original
Ratio between scenarios change in COVID−19 cases
1.0
0.8
0.7
Range
10th/90th percentile
25th/75th percentile

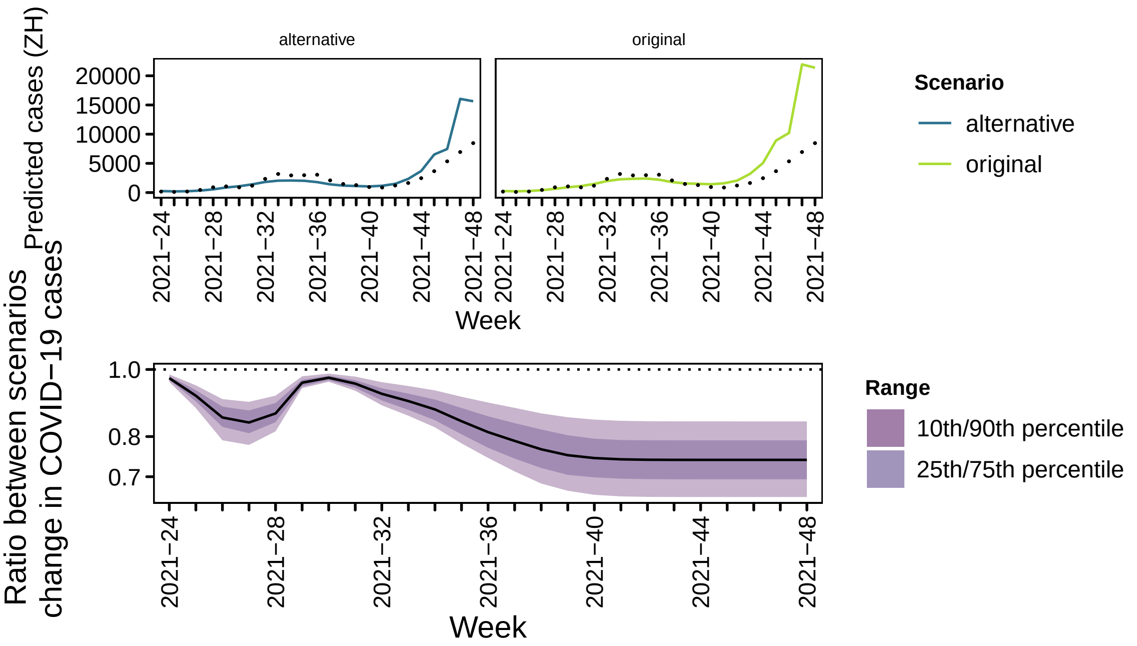

[4] M. Bekker-Nielsen Dunbar, F. Hofmann, S. Meyer, and L. Held (joint last authors). Endemic-epidemic modelling of school closure to prevent spread of COVID-19 in

Switzerland (protocol available at DOI:10.17605/osf.io/qh3gd). *medRxiv*, 2023. DOI:10.1101/2023.03.21.23287519.
