## Supporting information (supp) for "The COVID-19 vaccination campaign in Switzerland and its impact on disease spread"

M. Bekker-Nielsen Dunbar and L. Held
Epidemiology, Biostatistics and Prevention Institute, University of Zurich

For full information on the work this document supports please see the manuscript and the study protocol [1]. This supporting document contains additional plots.

#### Contents

**1 Regions** **2**

**2 Population** **4**

**3 Contacts** **6**

**4 Dispersion** **56**

**5 Vaccination coverage covariate construction** **106**

**6 Cases** **107**

**7 Measles literature** **107**

**8 Predicted cases** **108**

8.1 Summed across prediction window . . . . . . . . . . . . . . . . . . . . . . 109

### 1 Regions

Subnational regions in Switzerland

| Abbreviation | Region |
|---|---|
| AG | Aargau |
| AI | Appenzell Innerrhoden |
| AR | Appenzell Ausserrhoden |
| BE | Bern |
| BL | Basel-Landschaft |
| BS | Basel-Stadt |
| FR | Freiburg |
| GE | Genève |
| GL | Glarus |
| GR | Graubünden |
| JU | Jura |
| LU | Luzern |
| NE | Neuchâtel |
| NW | Nidwalden |
| OW | Obwalden |
| SG | St. Gallen |
| SH | Schaffhausen |
| SO | Solothurn |
| SZ | Schwyz |
| TG | Thurgau |
| TI | Ticino |
| UR | Uri |
| VD | Vaud |
| VS | Valais |
| ZG | Zug |
| ZH | Zürich |

### 2 Population

Population in Switzerland by age and region

| | 10 - 19 | 20 - 29 | 30 - 39 | 40 - 49 | 50 - 59 | 60 - 69 | 70 - 79 | 80+ |
|---|---|---|---|---|---|---|---|---|
| AG | 69526 | 79778 | 100400 | 96825 | 107532 | 83686 | 58426 | 33478 |
| AI | 1666 | 2010 | 2084 | 1930 | 2484 | 2025 | 1433 | 977 |
| AR | 5311 | 5879 | 7181 | 7092 | 8616 | 7372 | 5050 | 3082 |
| BE | 97838 | 115407 | 143365 | 137398 | 155465 | 129560 | 102884 | 63650 |
| BL | 28384 | 30005 | 35909 | 38786 | 45701 | 36436 | 29538 | 20175 |
| BS | 15827 | 23124 | 33455 | 27627 | 26847 | 21416 | 16101 | 13129 |
| FR | 36160 | 42907 | 46547 | 45409 | 48825 | 35229 | 25069 | 13531 |
| GE | 53904 | 64139 | 76283 | 77034 | 72478 | 48922 | 36705 | 26509 |
| GL | 3827 | 4698 | 5693 | 5328 | 6163 | 5302 | 3795 | 2322 |
| GR | 17630 | 22458 | 26120 | 26075 | 31774 | 26969 | 20461 | 12271 |
| JU | 7854 | 8949 | 8826 | 8992 | 10842 | 9320 | 7250 | 4459 |
| LU | 41479 | 52323 | 61154 | 55769 | 62118 | 48108 | 33585 | 22055 |
| NE | 19083 | 21841 | 23159 | 23956 | 25764 | 19209 | 15527 | 10363 |
| NW | 3946 | 4633 | 5642 | 5613 | 7273 | 6047 | 4461 | 2365 |
| OW | 3787 | 4199 | 5035 | 5024 | 6121 | 4946 | 3504 | 1994 |
| SG | 52232 | 64970 | 73653 | 66764 | 76399 | 60389 | 44078 | 26720 |
| SH | 7781 | 9350 | 11206 | 10746 | 12544 | 10864 | 8053 | 5401 |
| SO | 26026 | 31541 | 38552 | 35972 | 43561 | 36105 | 25255 | 15637 |
| SZ | 15362 | 18080 | 22237 | 22731 | 27140 | 20444 | 13712 | 7945 |
| TG | 27865 | 32707 | 40710 | 37213 | 44654 | 35251 | 23637 | 13791 |
| TI | 33880 | 37316 | 39781 | 48290 | 58508 | 43476 | 36178 | 26276 |
| UR | 3752 | 4333 | 4637 | 4684 | 5527 | 4778 | 3595 | 2127 |
| VD | 91874 | 106260 | 120501 | 118412 | 115937 | 80347 | 62222 | 39500 |
| VS | 34634 | 42484 | 47853 | 46165 | 53016 | 42899 | 33457 | 18921 |
| ZG | 12888 | 13188 | 18604 | 19455 | 21071 | 14649 | 10312 | 6309 |
| ZH | 146478 | 185153 | 256370 | 232249 | 223652 | 157468 | 120161 | 79000 |

### 3 Contacts

The time-varying contact matrices are plotted below

```
## [[1]]
```

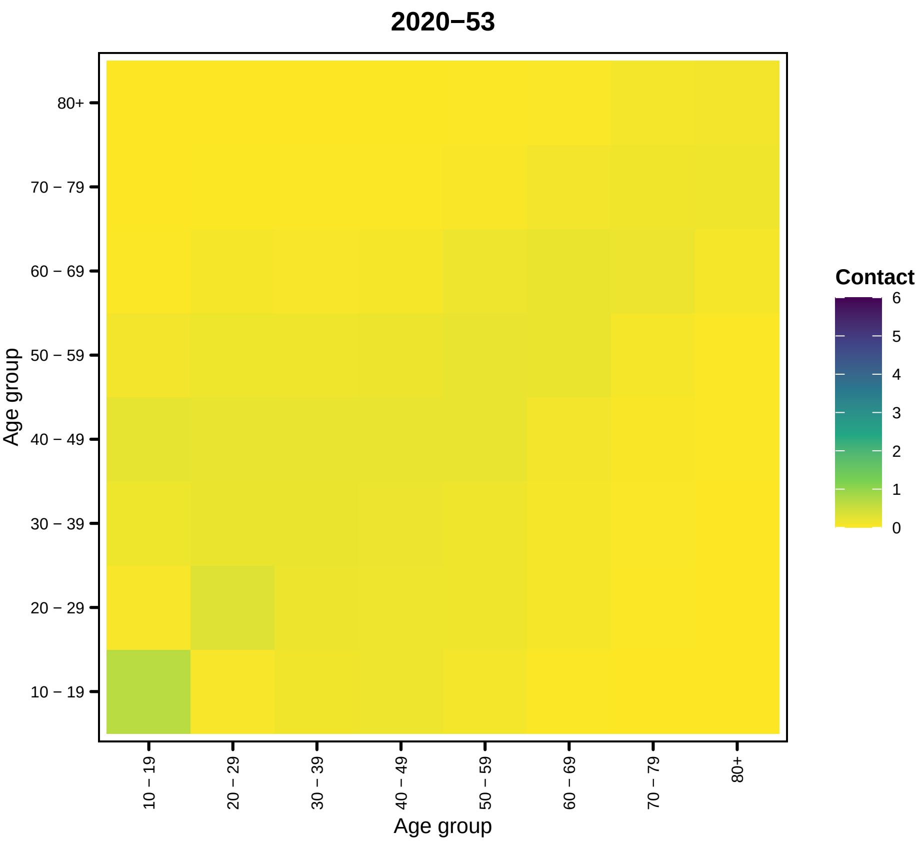


```
##
## [[2]]
```

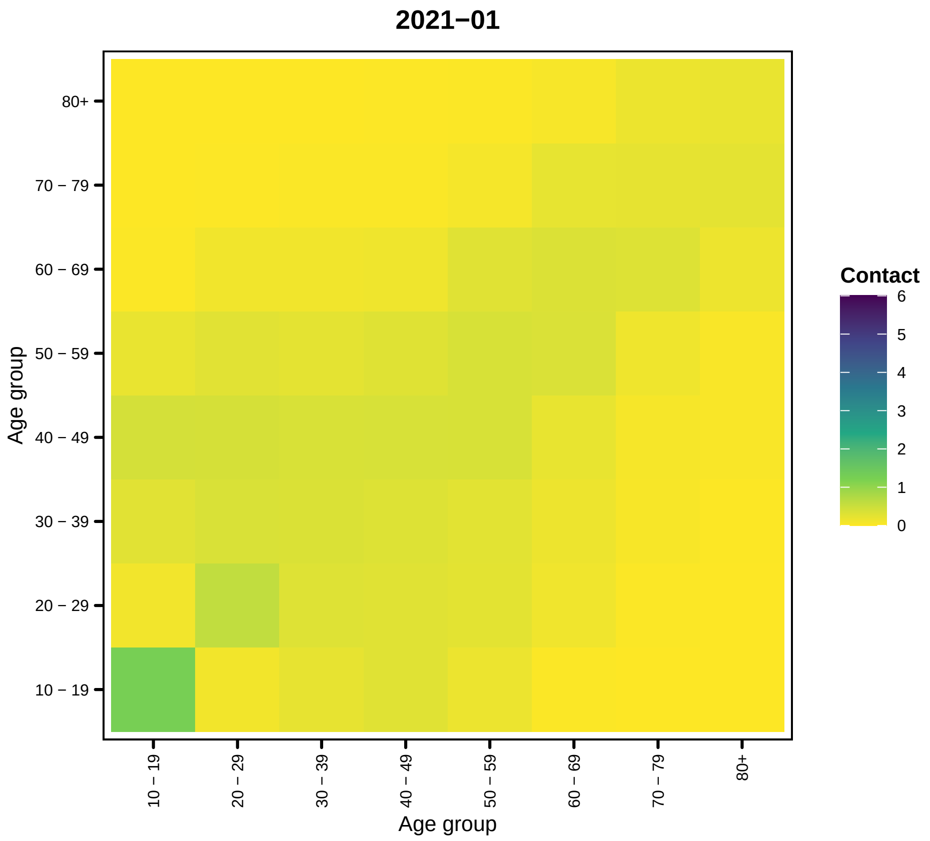
2021−01
Contact
6
5
4
3
2
1
0
80+
70 − 79
60 − 69
50 − 59
40 − 49
30 − 39
20 − 29
10 − 19
Age group
Age group

22

```
##
## [[3]]
```

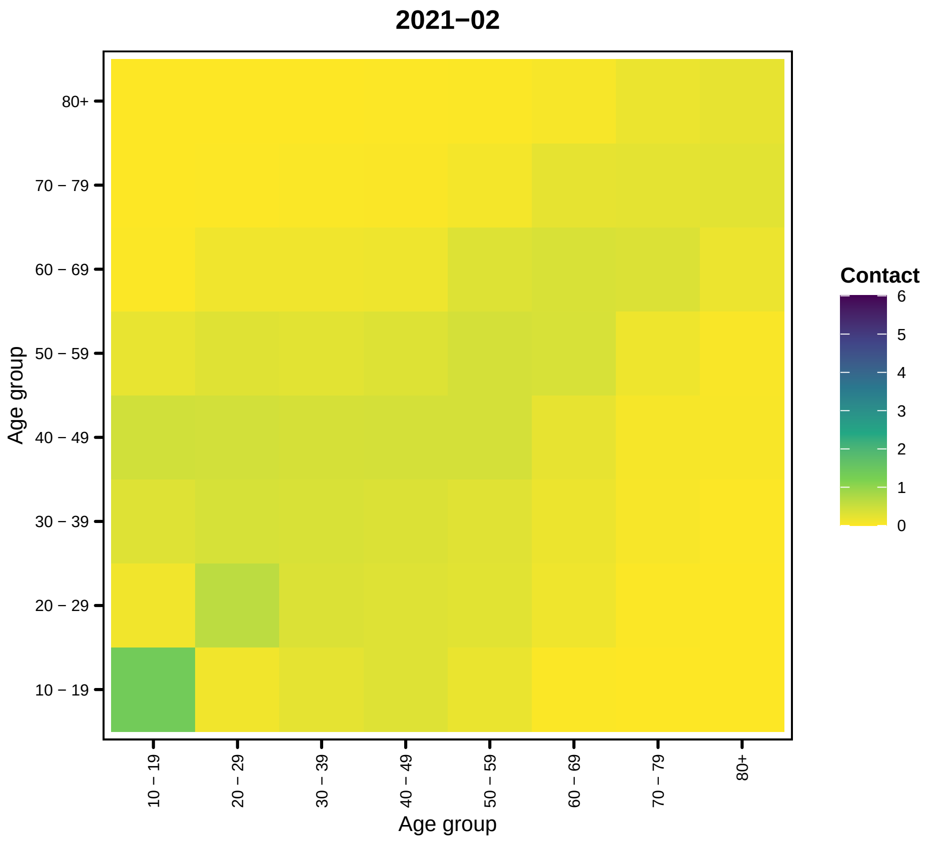

2021−02
80+
70 − 79
60 − 69
50 − 59
40 − 49
30 − 39
20 − 29
10 − 19
Age group
Age group
Contact
6
5
4
3
2
1
0


23

```
##
## [[4]]
```

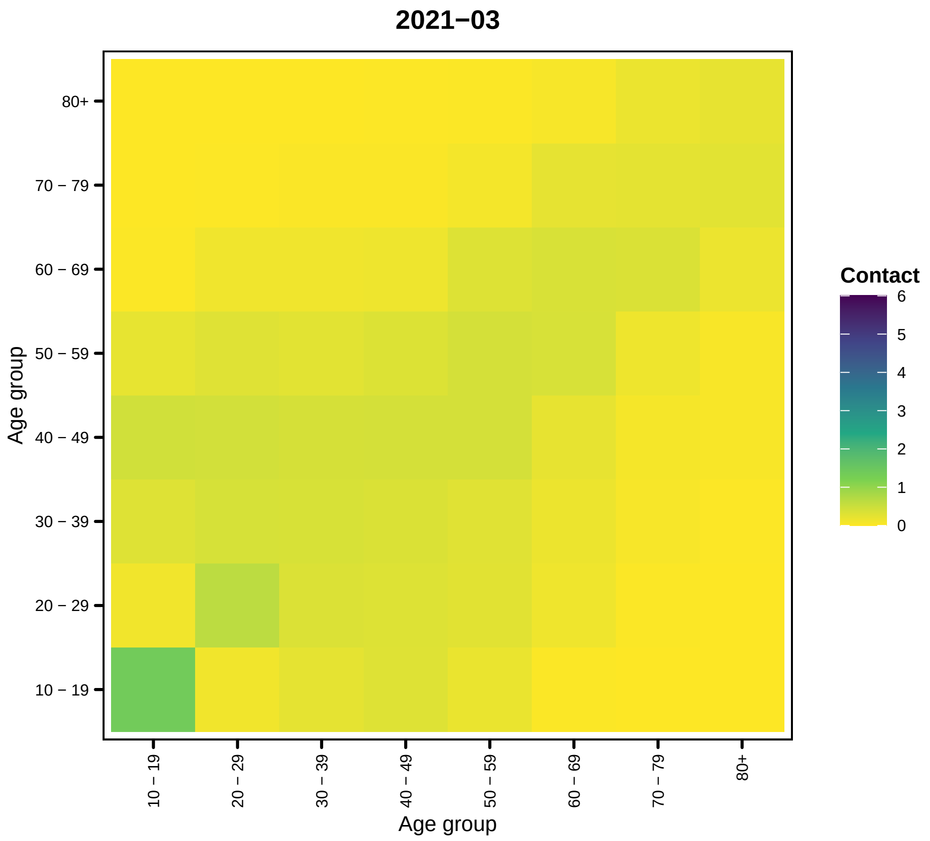

2021−03
80+
70 − 79
60 − 69
50 − 59
40 − 49
30 − 39
20 − 29
10 − 19
Age group
Contact
6
5
4
3
2
1
0
10 − 19
20 − 29
30 − 39
40 − 49
50 − 59
60 − 69
70 − 79
80+
Age group


24

```
##
## [[5]]
```

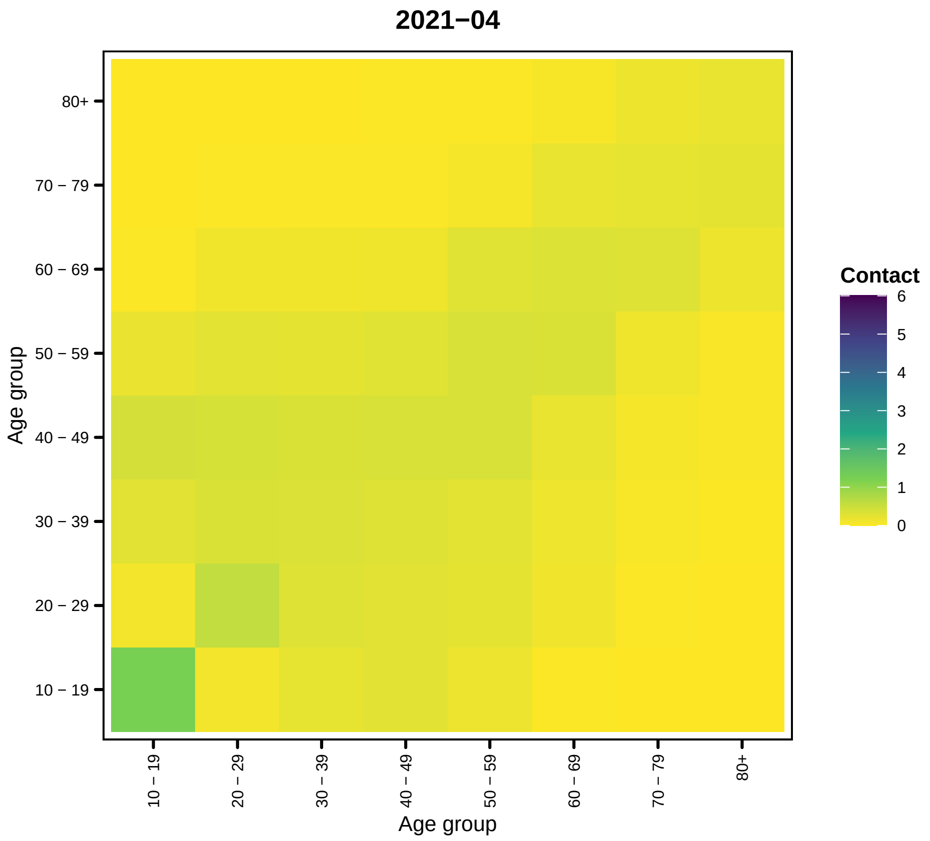
2021−04
Age group
80+
70 − 79
60 − 69
50 − 59
40 − 49
30 − 39
20 − 29
10 − 19
Contact
6
5
4
3
2
1
0

25

```
##
## [[6]]
```

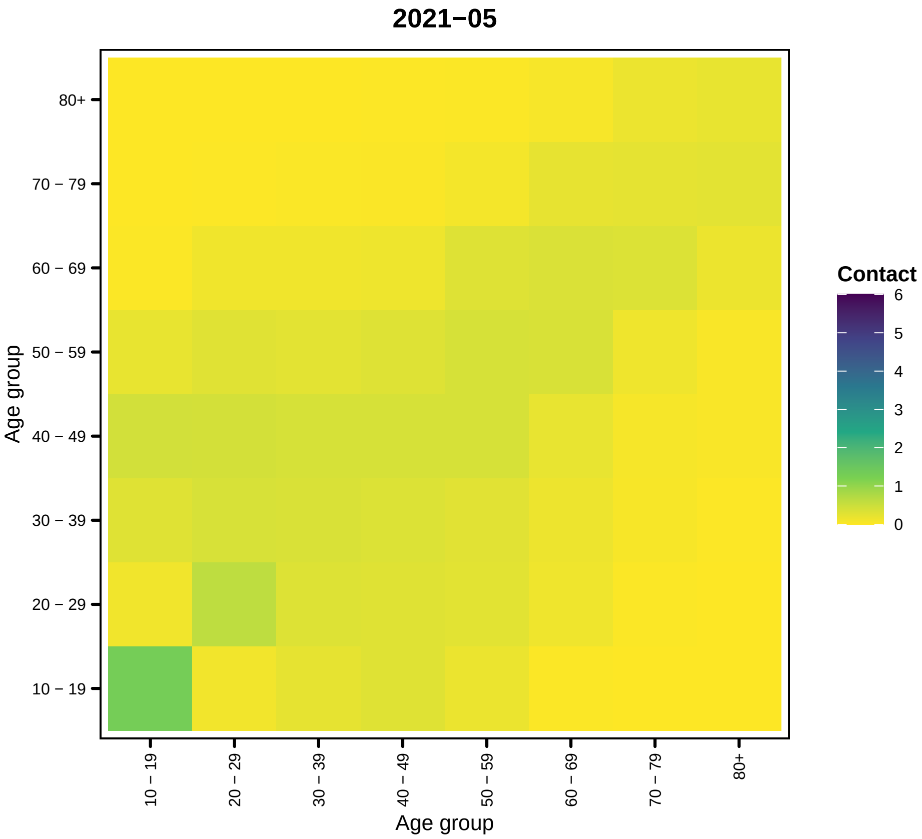

2021−05
80+
70 − 79
60 − 69
50 − 59
40 − 49
30 − 39
20 − 29
10 − 19
Age group
10 − 19
20 − 29
30 − 39
40 − 49
50 − 59
60 − 69
70 − 79
80+
Age group
Contact
6
5
4
3
2
1
0


26

```
##
## [[7]]
```

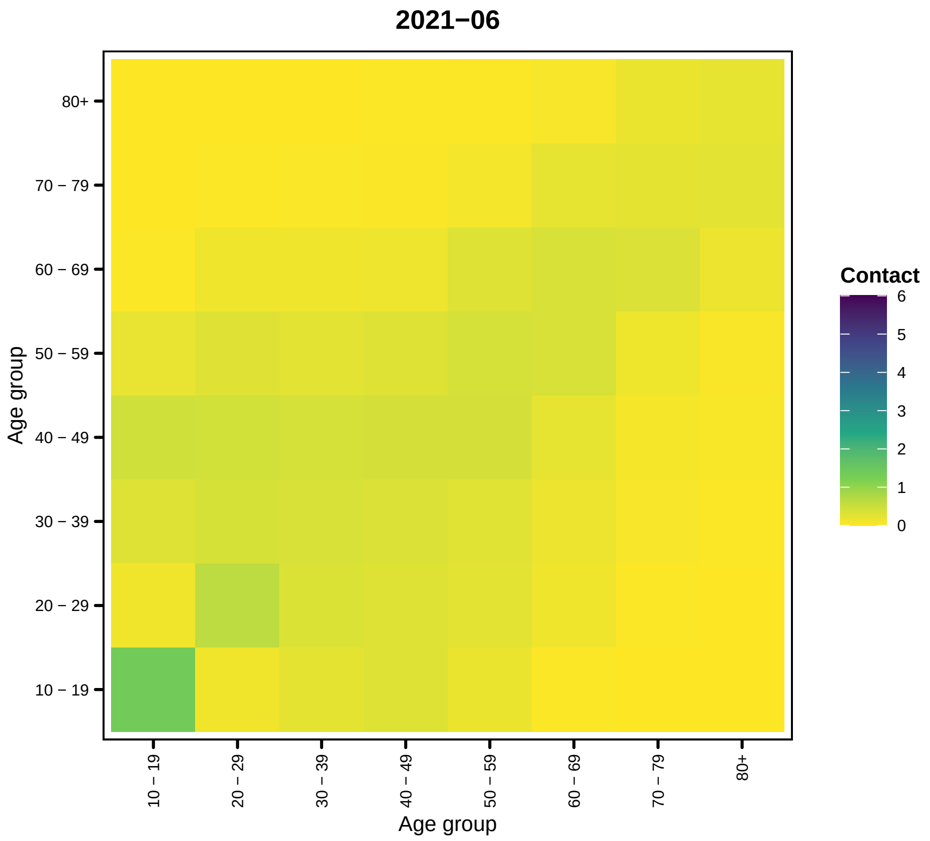
2021−06
Contact
6
5
4
3
2
1
0
80+
70 − 79
60 − 69
50 − 59
40 − 49
30 − 39
20 − 29
10 − 19
Age group
Age group

27

```
##
## [[8]]
```

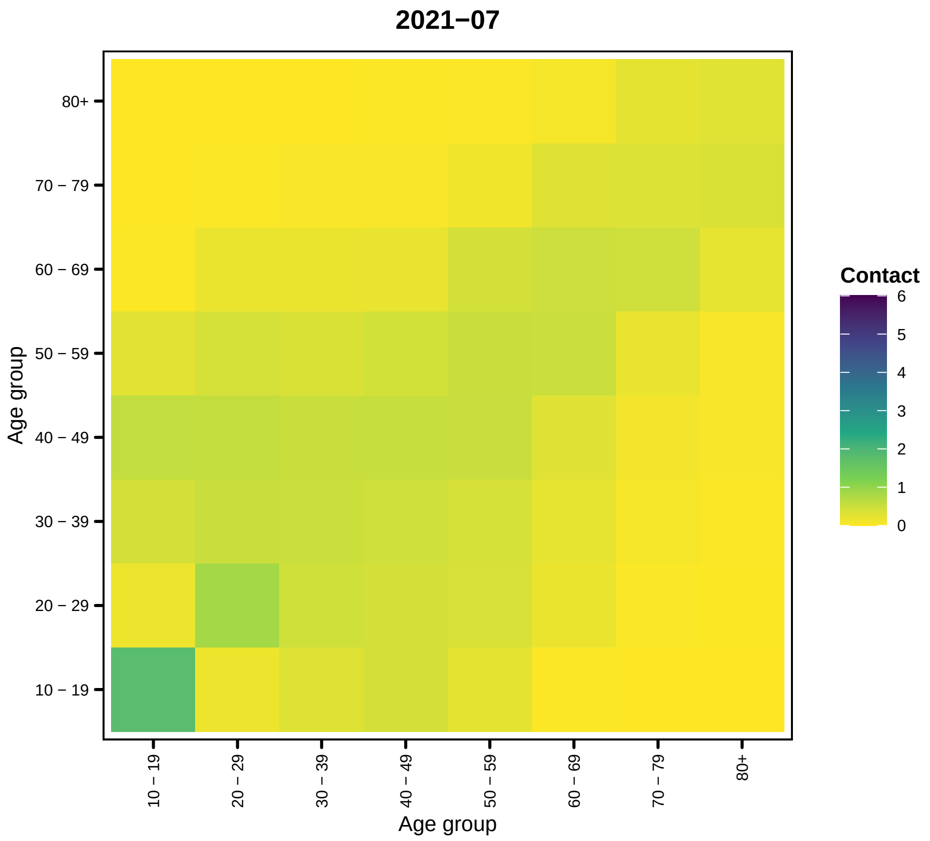
2021−07
80+
70 − 79
60 − 69
50 − 59
40 − 49
30 − 39
20 − 29
10 − 19
Age group
10 − 19
20 − 29
30 − 39
40 − 49
50 − 59
60 − 69
70 − 79
80+
Age group
Contact
6
5
4
3
2
1
0

28

```
##
## [[9]]
```

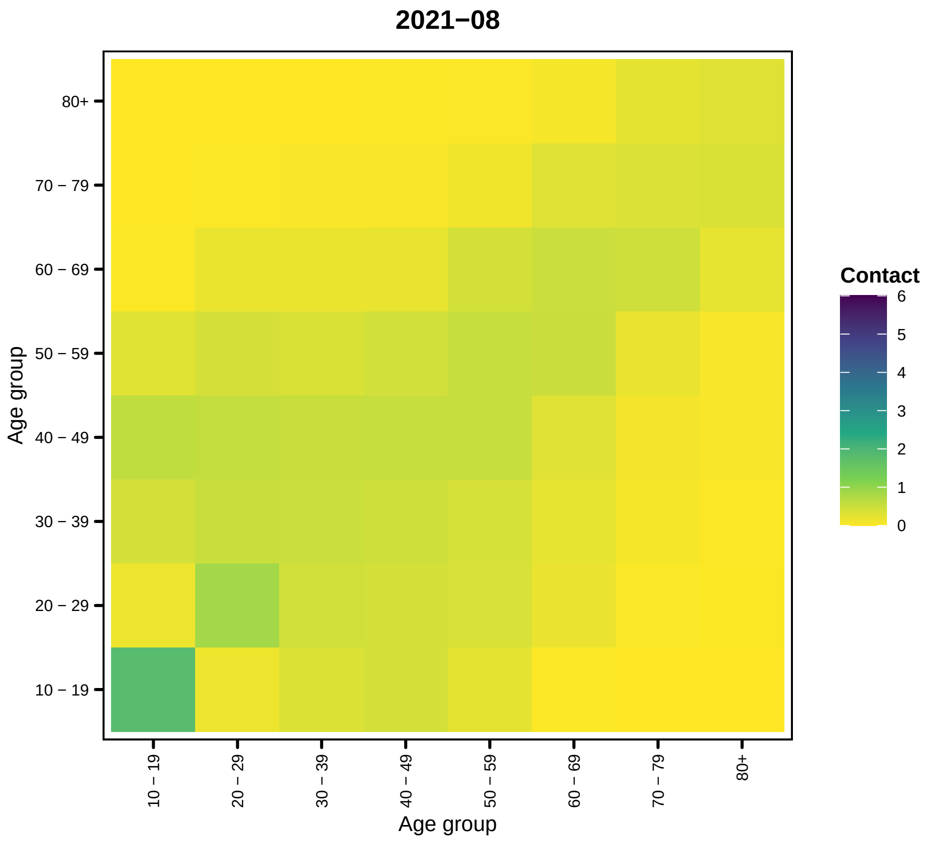

2021−08
80+
70 − 79
60 − 69
50 − 59
40 − 49
30 − 39
20 − 29
10 − 19
Age group
Age group
Contact
6
5
4
3
2
1
0


29

```
##
## [[10]]
```

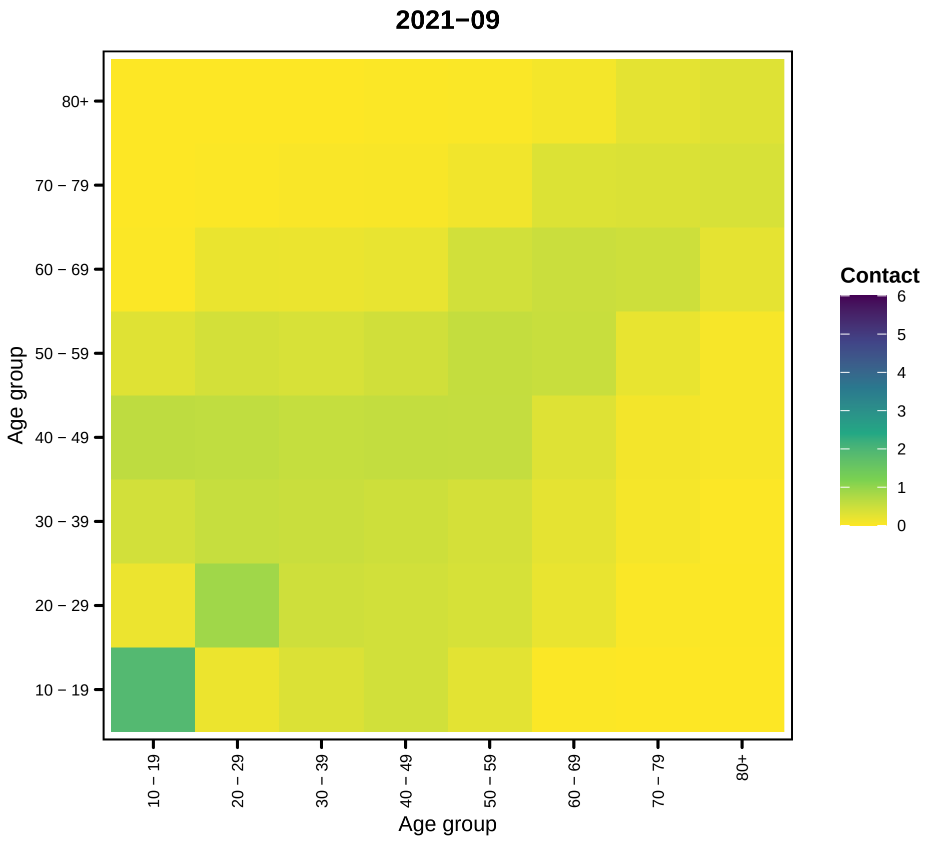
2021−09
Age group
80+
70 − 79
60 − 69
50 − 59
40 − 49
30 − 39
20 − 29
10 − 19
Age group
Contact
6
5
4
3
2
1
0

30

```
##

## [[11]]
```

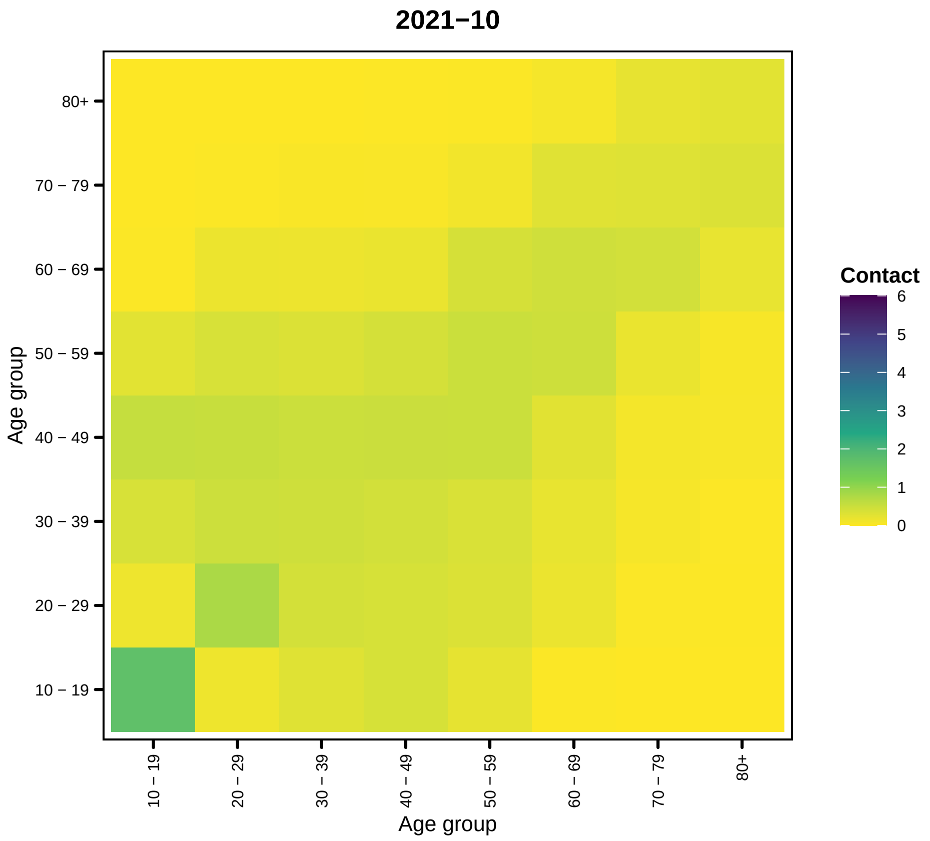
2021−10
80+
70 − 79
60 − 69
50 − 59
40 − 49
30 − 39
20 − 29
10 − 19
Age group
Contact
6
5
4
3
2
1
0
10 − 19
20 − 29
30 − 39
40 − 49
50 − 59
60 − 69
70 − 79
80+
Age group

31

```
##
## [[12]]
```

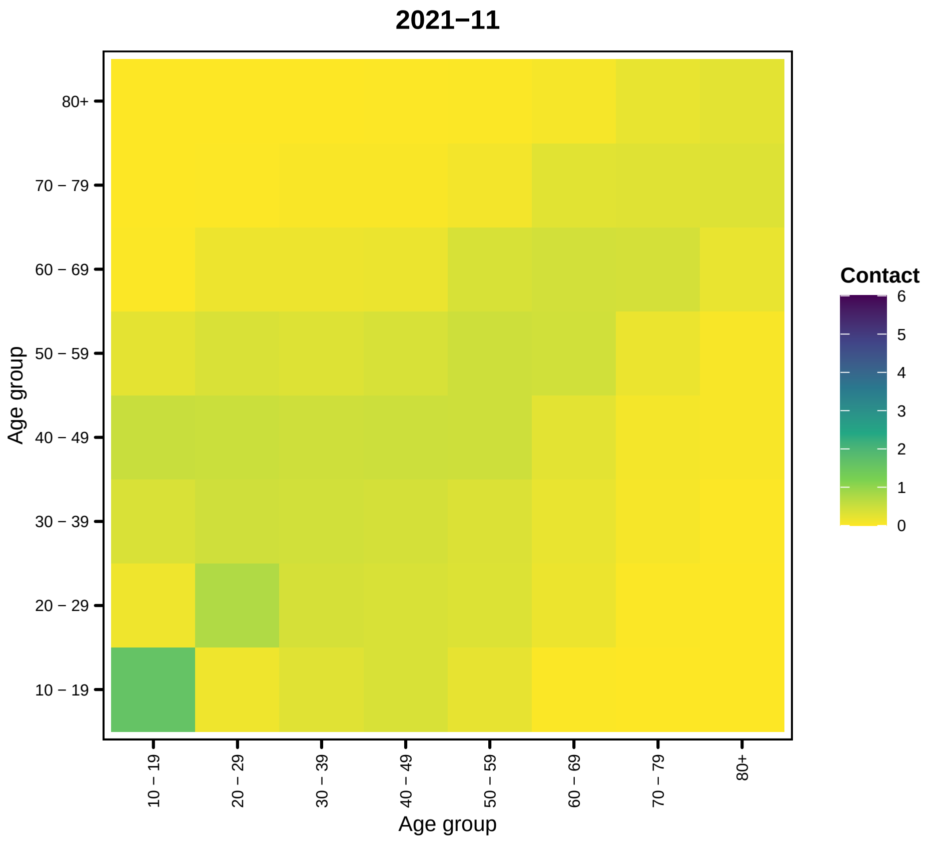
2021−11
Age group
80+
70 − 79
60 − 69
50 − 59
40 − 49
30 − 39
20 − 29
10 − 19
Contact
6
5
4
3
2
1
0
10 − 19
20 − 29
30 − 39
40 − 49
50 − 59
60 − 69
70 − 79
80+
Age group

32

```
##
## [[13]]
```

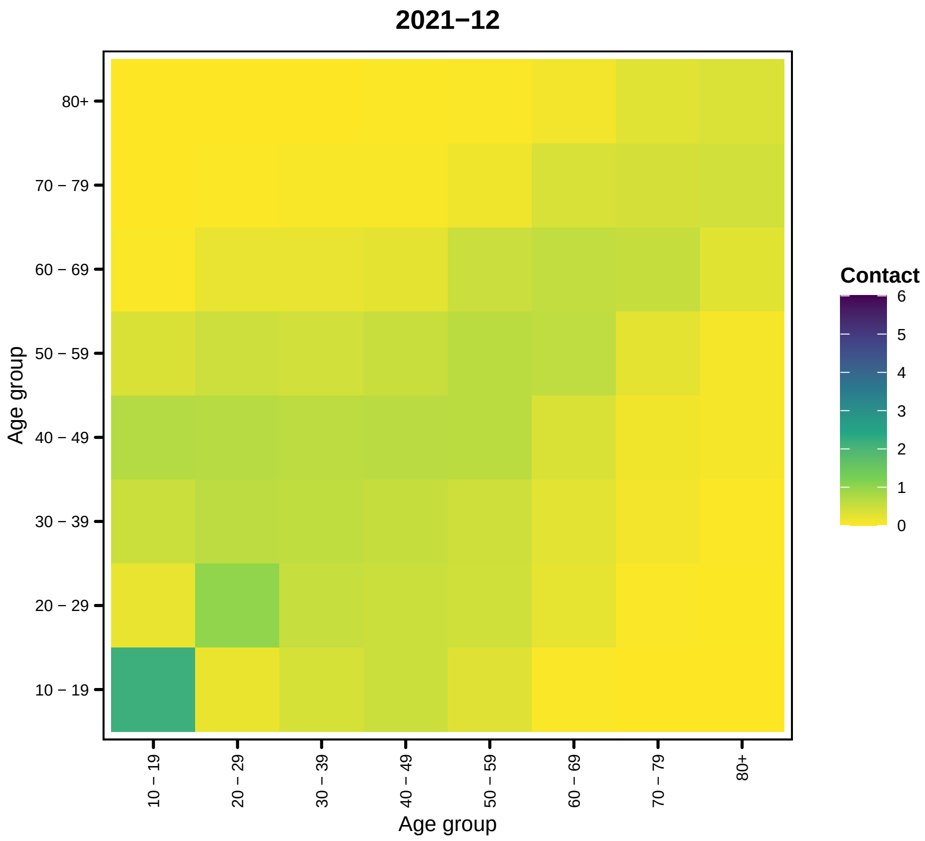

2021−12
80+
70 − 79
60 − 69
50 − 59
40 − 49
30 − 39
20 − 29
10 − 19
Age group
10 − 19
20 − 29
30 − 39
40 − 49
50 − 59
60 − 69
70 − 79
80+
Age group
Contact
6
5
4
3
2
1
0


33

```
##
## [[14]]
```

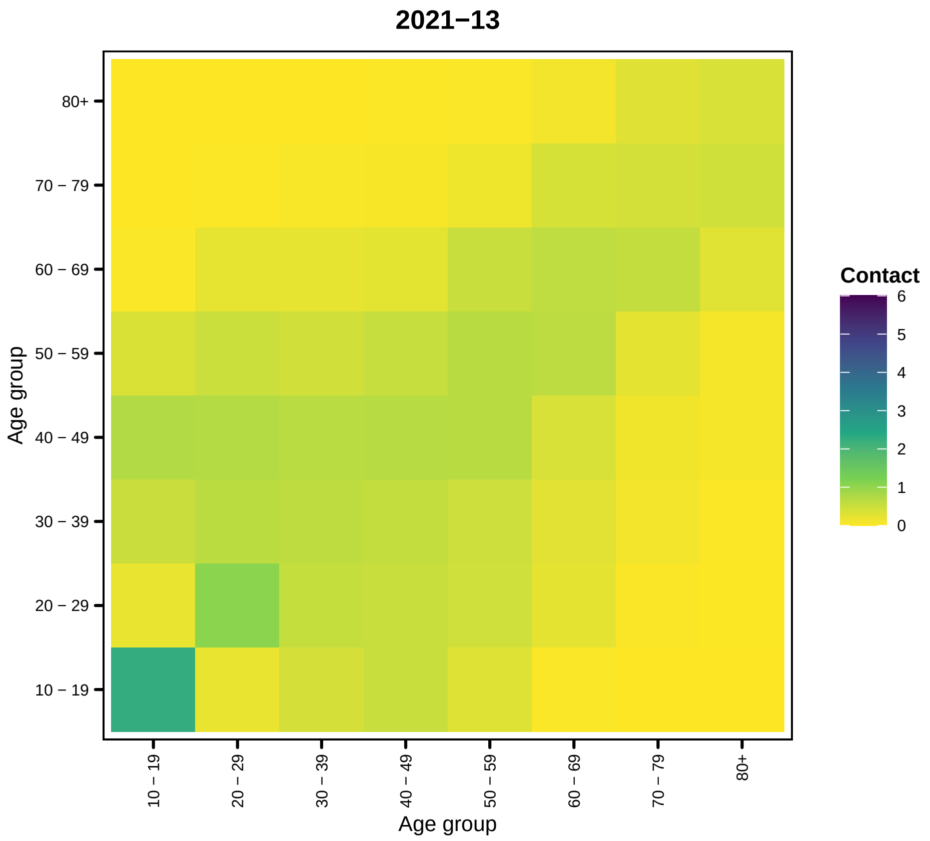
2021−13
Contact
6
5
4
3
2
1
0
80+
70 − 79
60 − 69
50 − 59
40 − 49
30 − 39
20 − 29
10 − 19
Age group
10 − 19
20 − 29
30 − 39
40 − 49
50 − 59
60 − 69
70 − 79
80+
Age group

34

```
##
## [[15]]
```

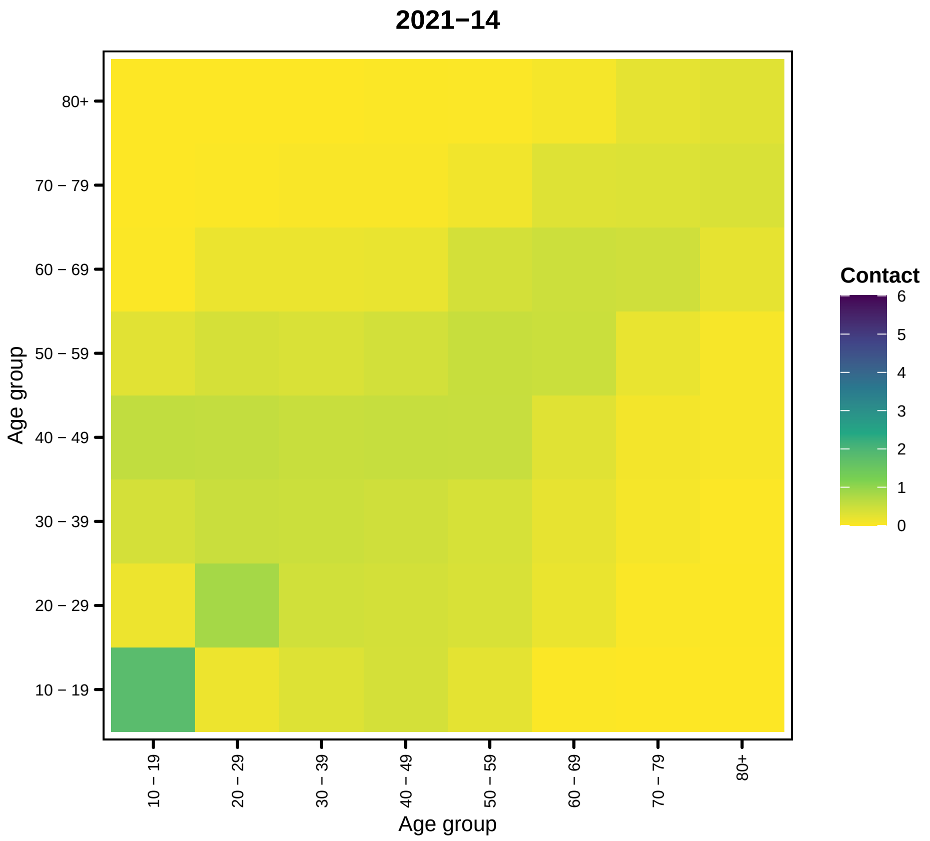
2021−14
Age group
80+
70 − 79
60 − 69
50 − 59
40 − 49
30 − 39
20 − 29
10 − 19
Age group
Contact
6
5
4
3
2
1
0

35

```
##
## [[16]]
```

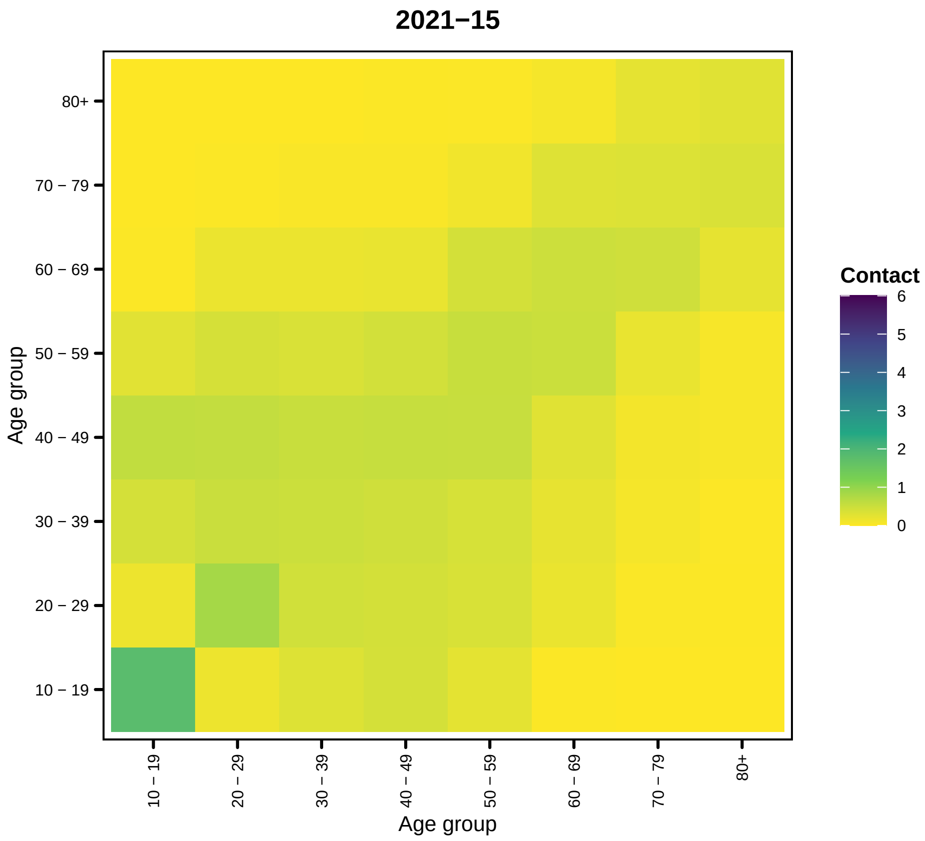
2021−15
80+
70 − 79
60 − 69
50 − 59
40 − 49
30 − 39
20 − 29
10 − 19
Age group
10 − 19
20 − 29
30 − 39
40 − 49
50 − 59
60 − 69
70 − 79
80+
Age group
Contact
6
5
4
3
2
1
0

36

```
##

## [[17]]
```

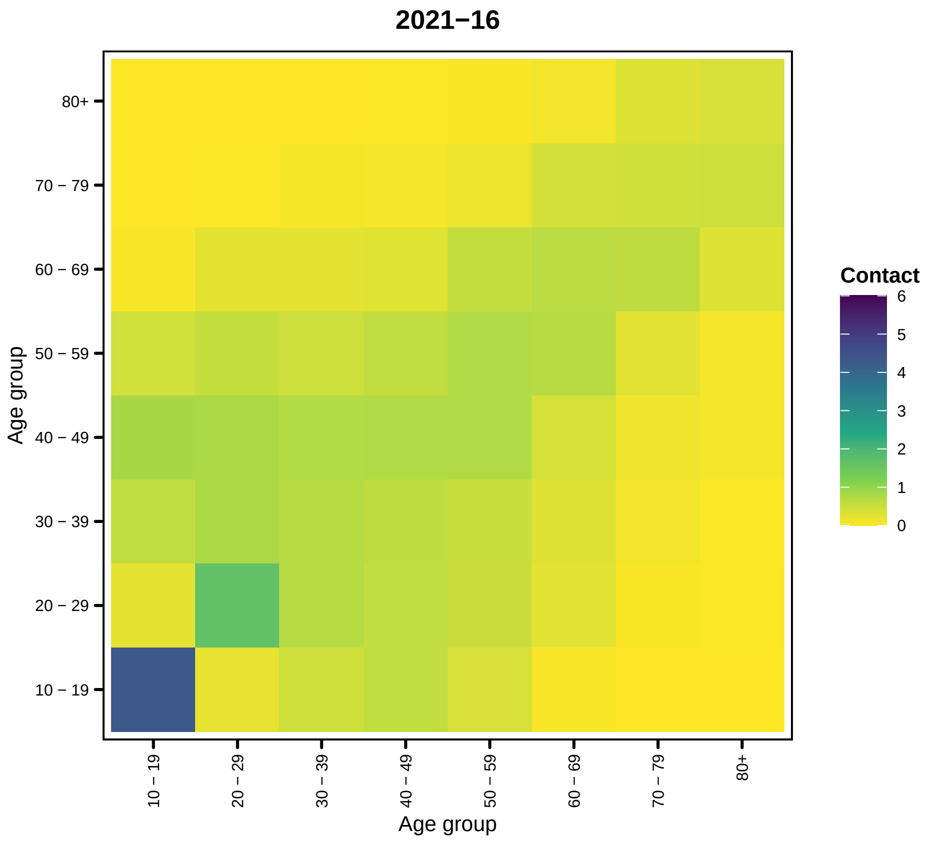
2021−16
Contact
6
5
4
3
2
1
0
80+
70 − 79
60 − 69
50 − 59
40 − 49
30 − 39
20 − 29
10 − 19
Age group
10 − 19
20 − 29
30 − 39
40 − 49
50 − 59
60 − 69
70 − 79
80+
Age group

37

```
##
## [[18]]
```

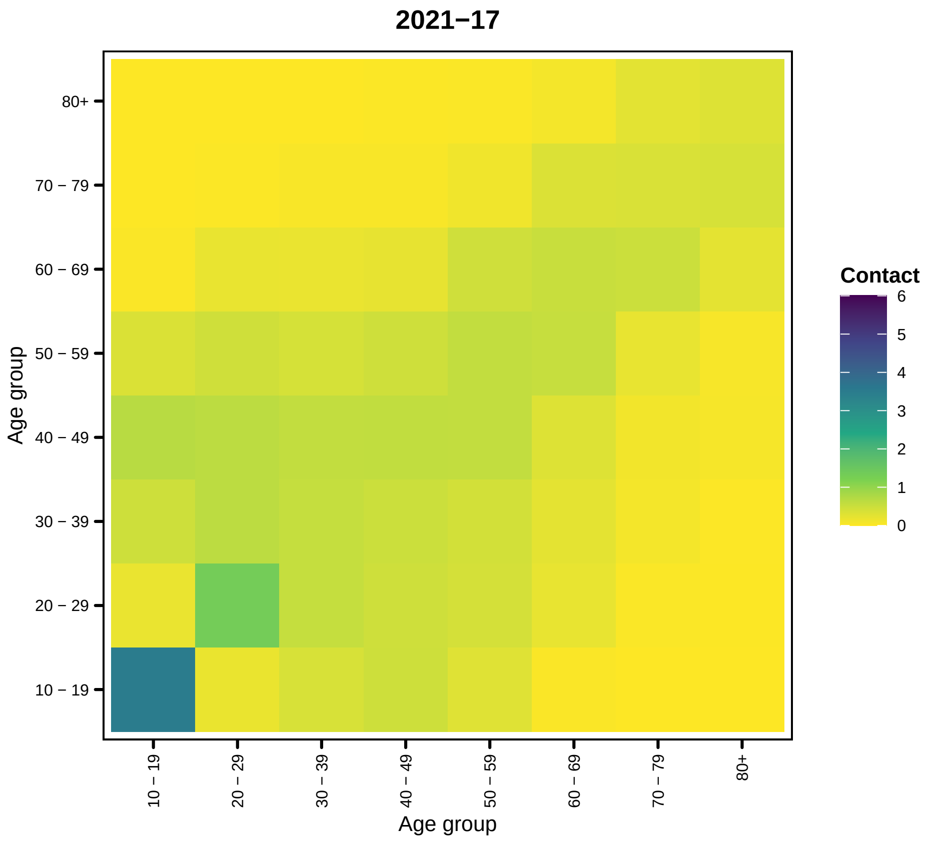
2021−17
Contact
6
5
4
3
2
1
0
Age group
80+
70 − 79
60 − 69
50 − 59
40 − 49
30 − 39
20 − 29
10 − 19
Age group
10 − 19
20 − 29
30 − 39
40 − 49
50 − 59
60 − 69
70 − 79
80+

38

```
##

## [[19]]
```

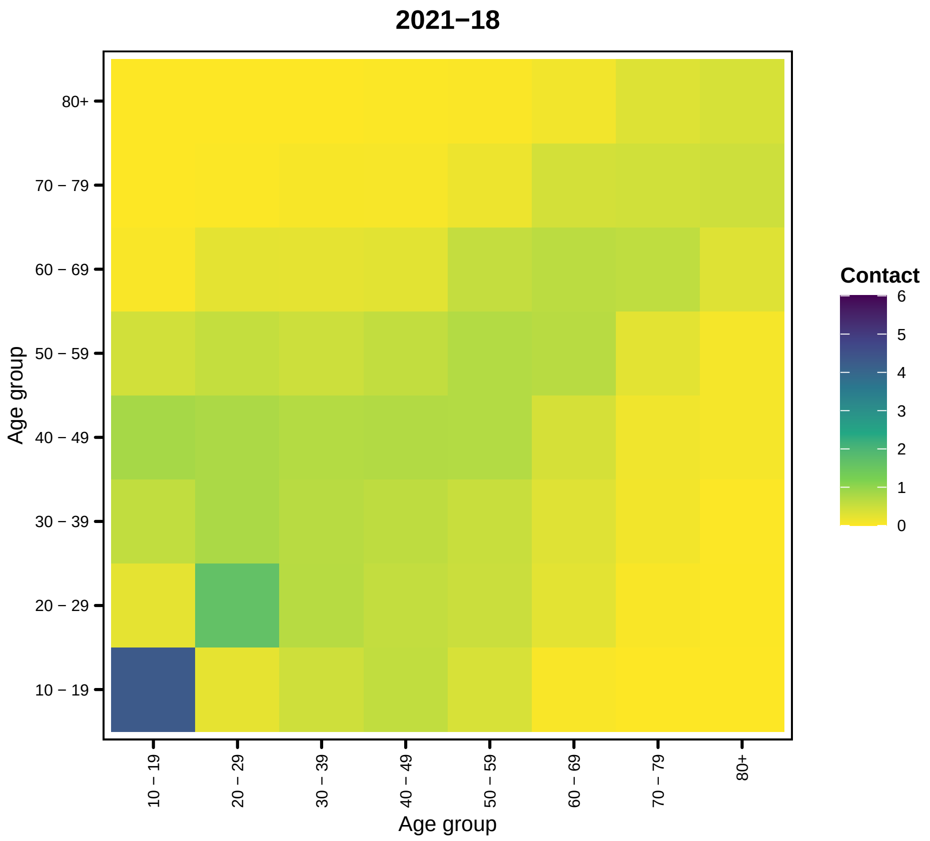

2021−18
80+
70 − 79
60 − 69
50 − 59
40 − 49
30 − 39
20 − 29
10 − 19
Age group
10 − 19
20 − 29
30 − 39
40 − 49
50 − 59
60 − 69
70 − 79
80+
Age group
Contact
6
5
4
3
2
1
0


39

```
##
## [[20]]
```

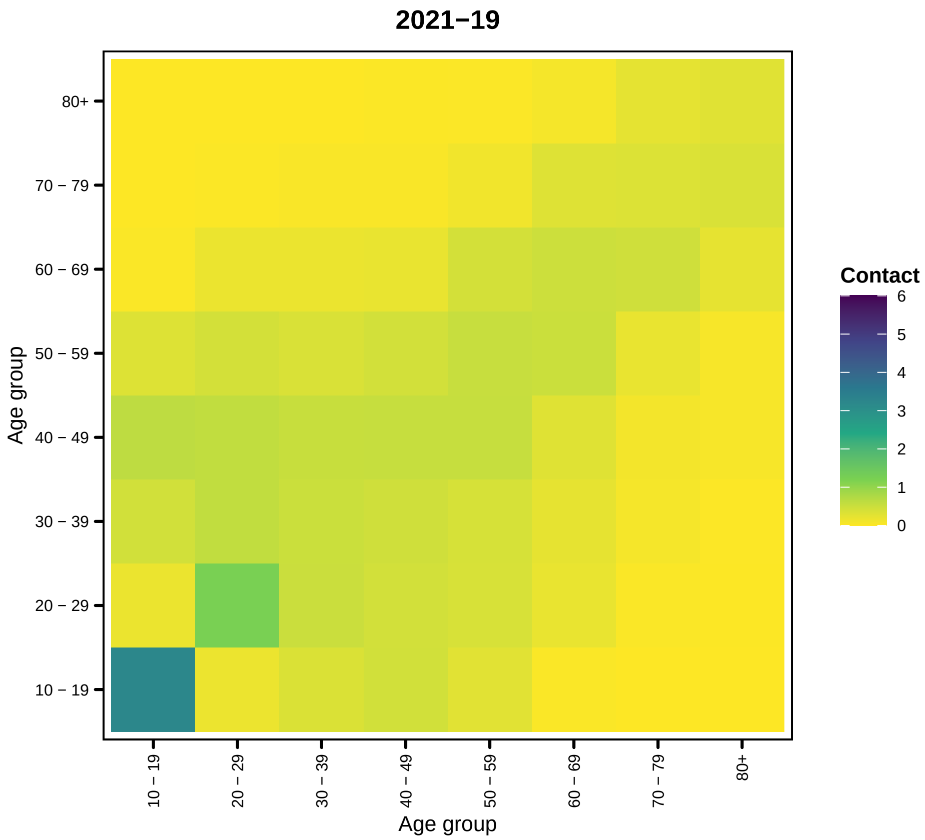
2021−19
80+
70 − 79
60 − 69
50 − 59
40 − 49
30 − 39
20 − 29
10 − 19
Age group
Age group
Contact
6
5
4
3
2
1
0

40

```
##
## [[21]]
```

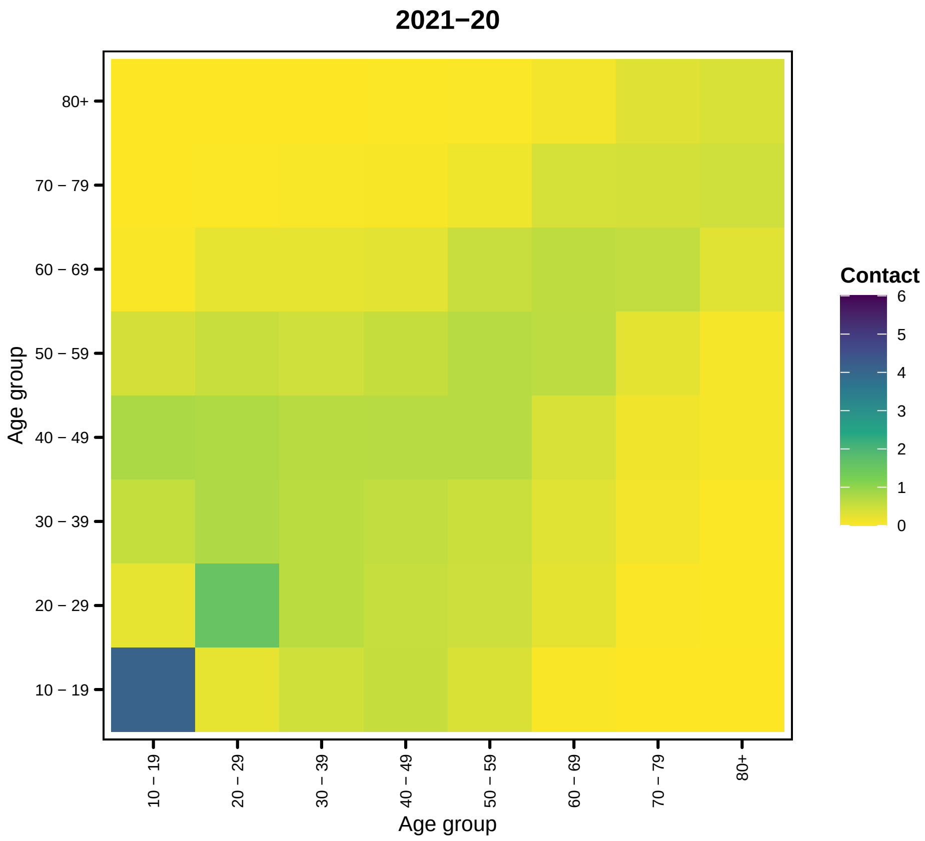

2021−20
80+
70 − 79
60 − 69
50 − 59
40 − 49
30 − 39
20 − 29
10 − 19
Age group
10 − 19
20 − 29
30 − 39
40 − 49
50 − 59
60 − 69
70 − 79
80+
Age group
Contact
6
5
4
3
2
1
0


41

```
##
## [[22]]
```

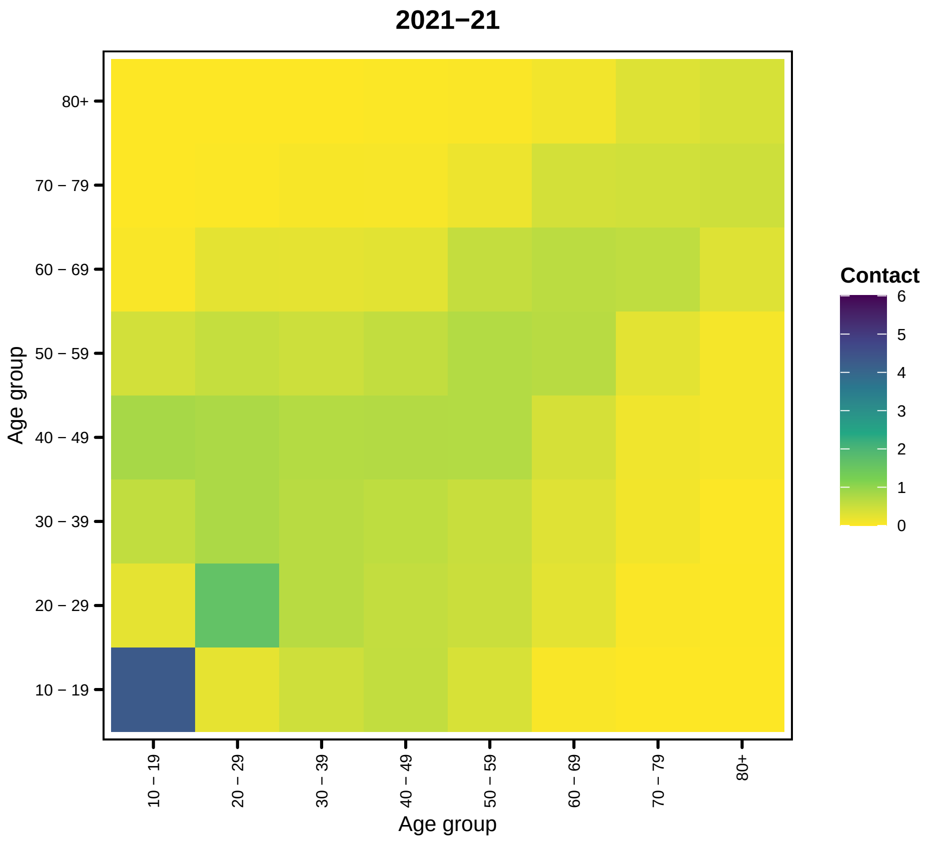
2021−21
80+
70 − 79
60 − 69
50 − 59
40 − 49
30 − 39
20 − 29
10 − 19
Age group
Age group
Contact
6
5
4
3
2
1
0

42

```
##
## [[23]]
```

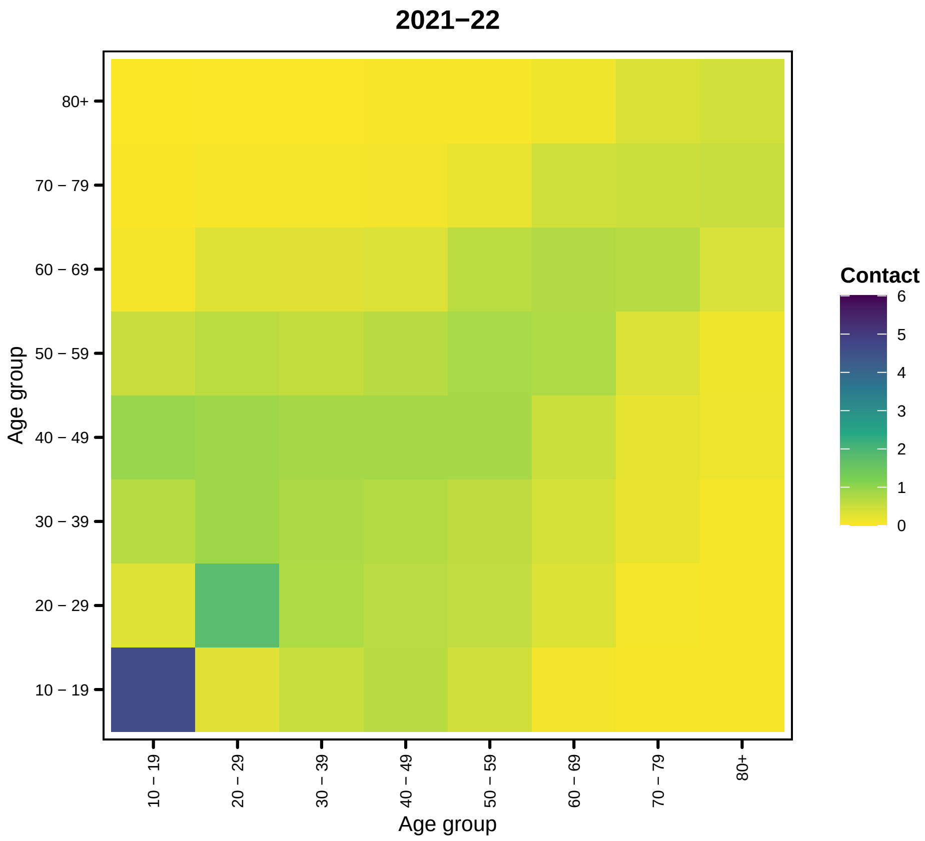
2021−22
80+
70 − 79
60 − 69
50 − 59
40 − 49
30 − 39
20 − 29
10 − 19
Age group
10 − 19
20 − 29
30 − 39
40 − 49
50 − 59
60 − 69
70 − 79
80+
Age group
Contact
6
5
4
3
2
1
0

43

```
##
## [[24]]
```

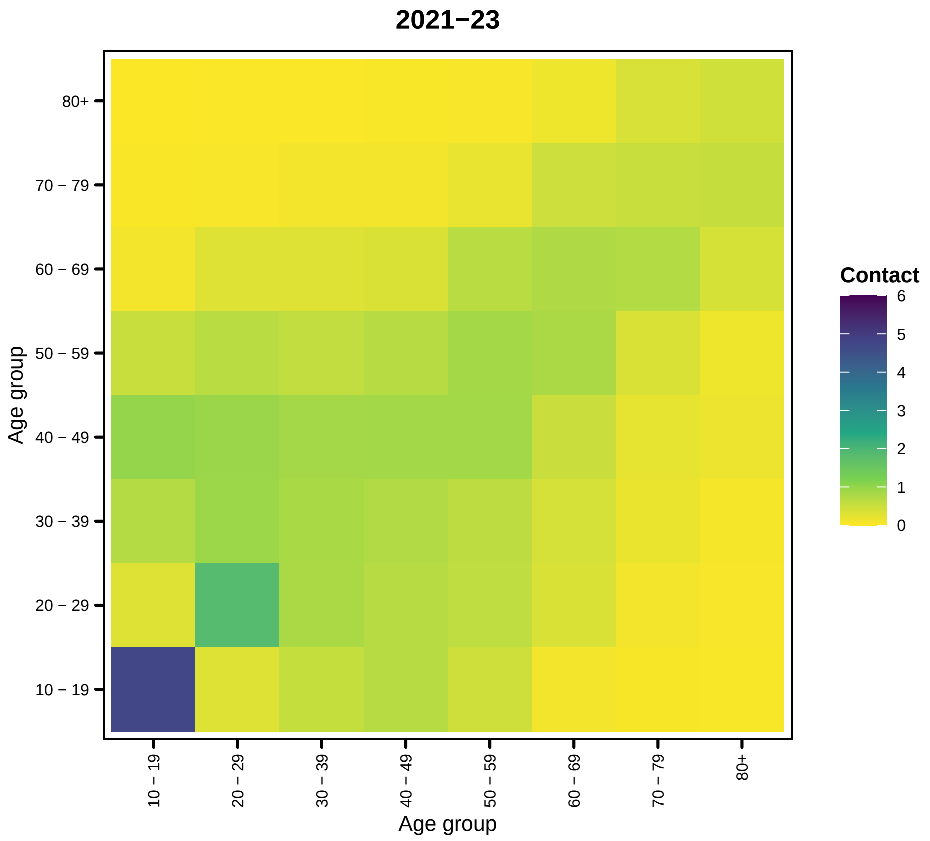

2021−23
80+
70 − 79
60 − 69
50 − 59
40 − 49
30 − 39
20 − 29
10 − 19
Age group
Contact
6
5
4
3
2
1
0


44

```
##
## [[25]]
```

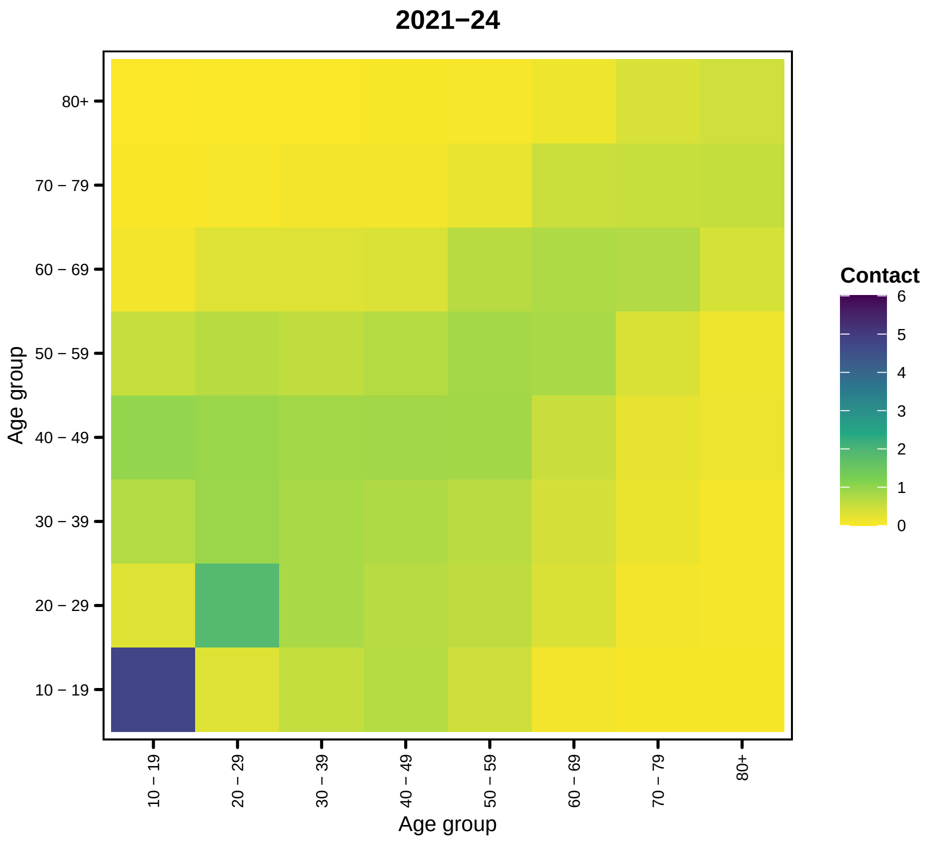

2021−24
80+
70 − 79
60 − 69
50 − 59
40 − 49
30 − 39
20 − 29
10 − 19
Age group
10 − 19
20 − 29
30 − 39
40 − 49
50 − 59
60 − 69
70 − 79
80+
Age group
Contact
6
5
4
3
2
1
0


45

```
##
## [[26]]
```

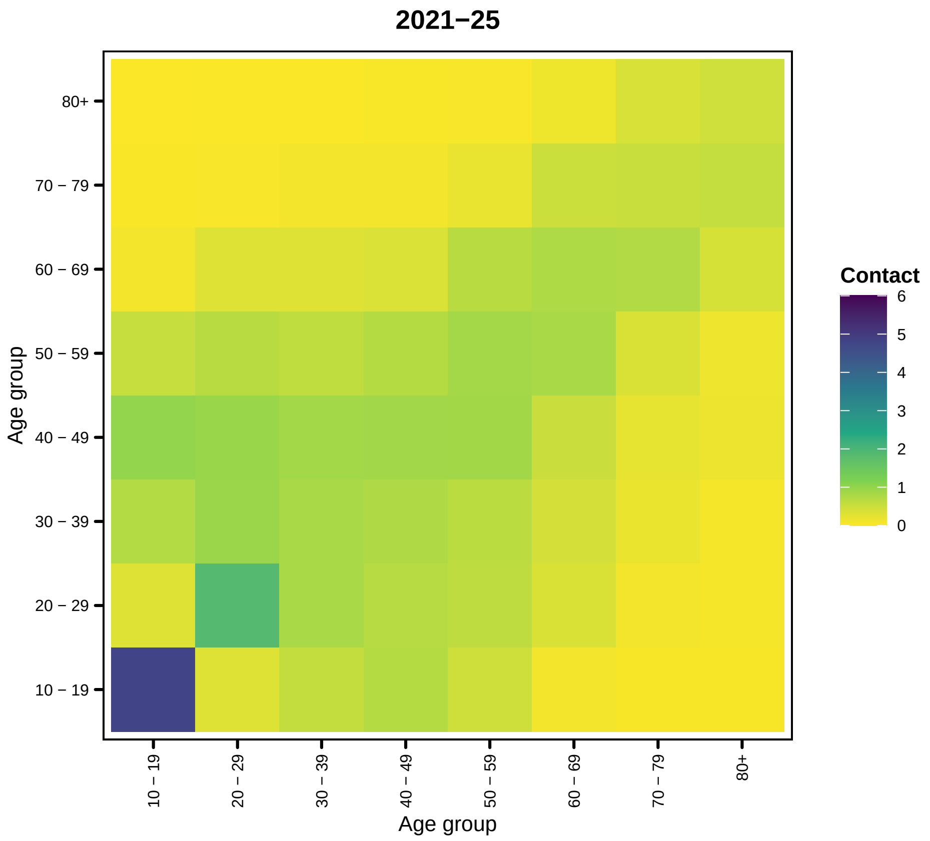

2021−25
80+
70 − 79
60 − 69
50 − 59
40 − 49
30 − 39
20 − 29
10 − 19
Age group
10 − 19
20 − 29
30 − 39
40 − 49
50 − 59
60 − 69
70 − 79
80+
Age group
Contact
6
5
4
3
2
1
0


46

```
##
## [[27]]
```

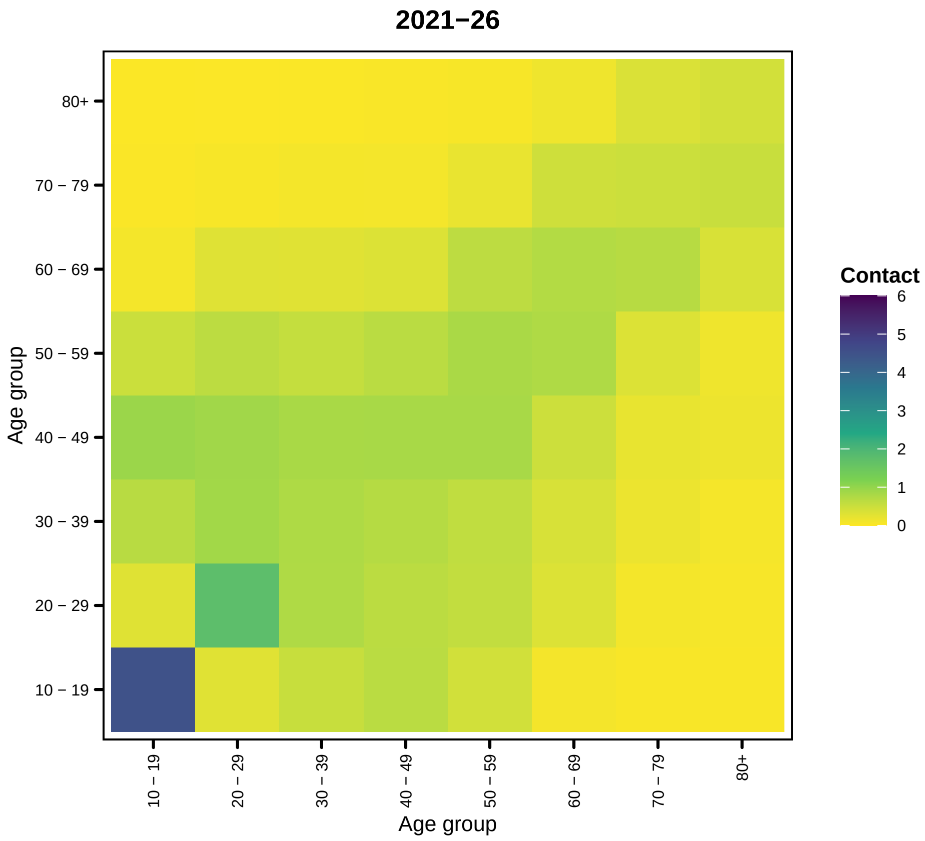
2021−26
80+
70 − 79
60 − 69
50 − 59
40 − 49
30 − 39
20 − 29
10 − 19
Age group
10 − 19
20 − 29
30 − 39
40 − 49
50 − 59
60 − 69
70 − 79
80+
Age group
Contact
6
5
4
3
2
1
0

47

```
##
## [[28]]
```

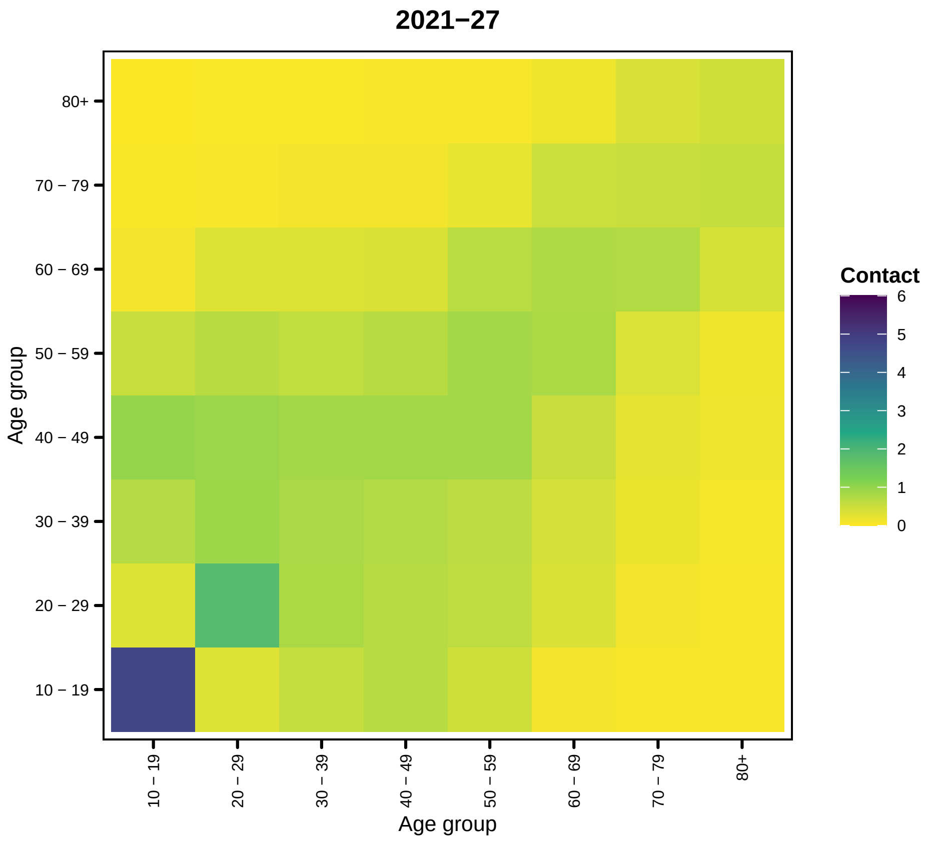
2021−27
Contact
6
5
4
3
2
1
0
Age group
80+
70 − 79
60 − 69
50 − 59
40 − 49
30 − 39
20 − 29
10 − 19
Age group

48

```
##
## [[29]]
```

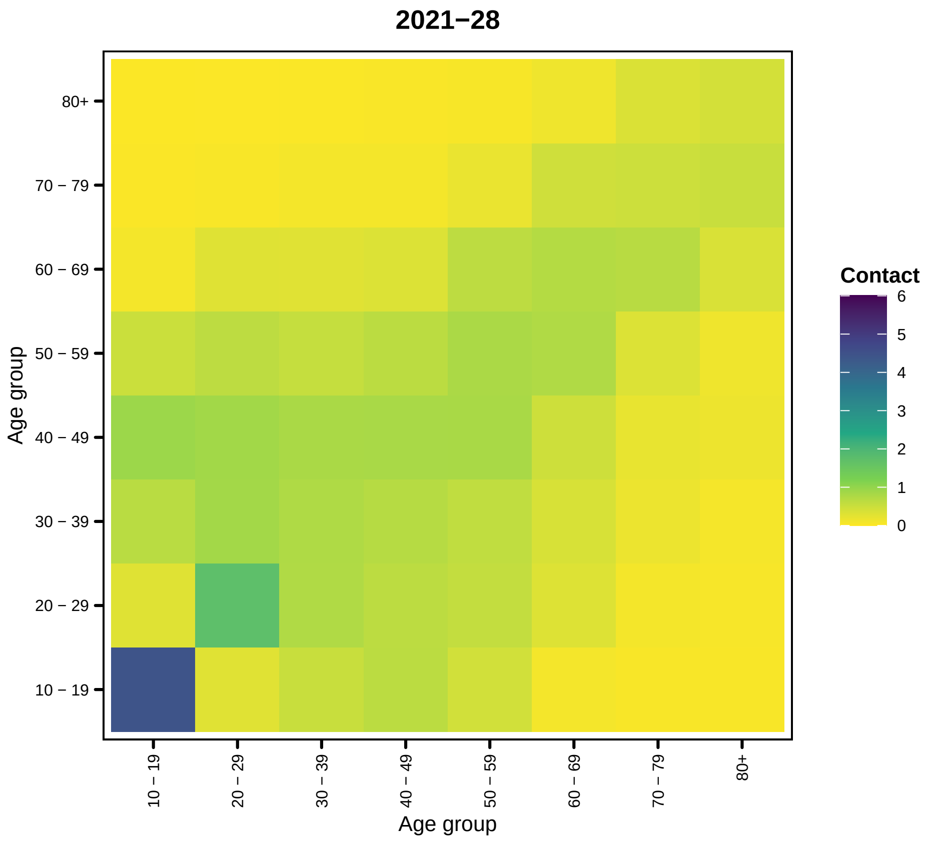

2021−28
80+
70 − 79
60 − 69
50 − 59
40 − 49
30 − 39
20 − 29
10 − 19
Age group
10 − 19
20 − 29
30 − 39
40 − 49
50 − 59
60 − 69
70 − 79
80+
Age group
Contact
6
5
4
3
2
1
0


49

```
##
## [[30]]
```

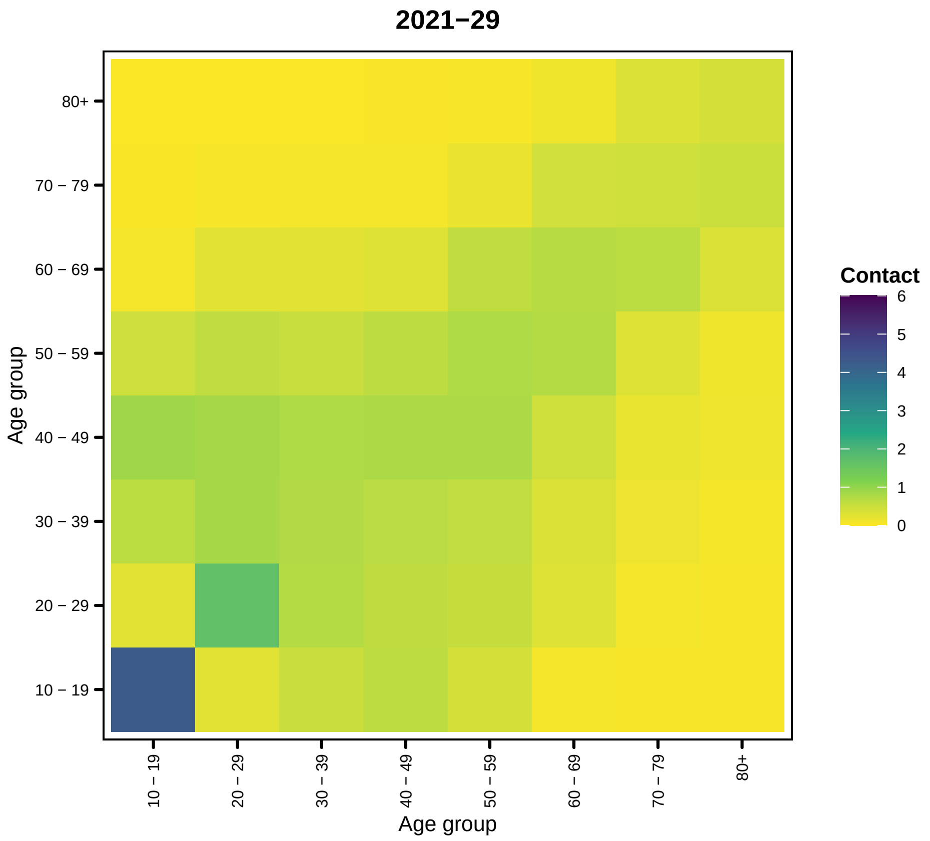
2021−29
Contact
6
5
4
3
2
1
0
80+
70 − 79
60 − 69
50 − 59
40 − 49
30 − 39
20 − 29
10 − 19
Age group
Age group

50

```
##
## [[31]]
```

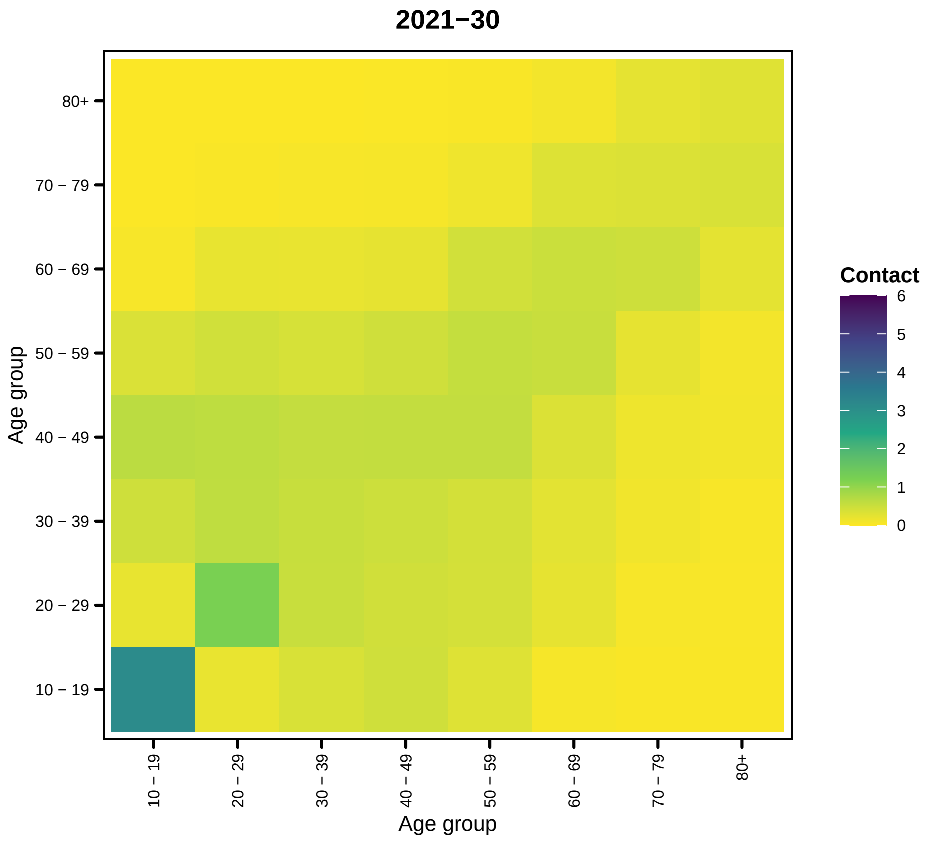
2021−30
80+
70 − 79
60 − 69
50 − 59
40 − 49
30 − 39
20 − 29
10 − 19
Age group
10 − 19
20 − 29
30 − 39
40 − 49
50 − 59
60 − 69
70 − 79
80+
Age group
Contact
6
5
4
3
2
1
0

51

```
##
## [[32]]
```

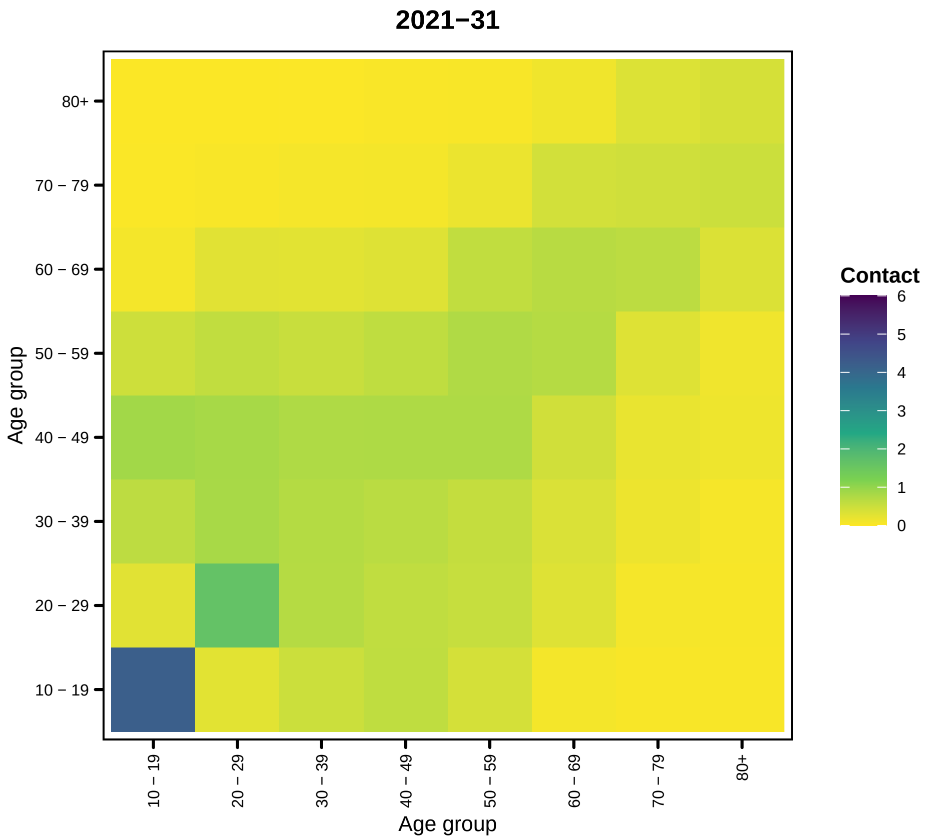

2021−31
80+
70 − 79
60 − 69
50 − 59
40 − 49
30 − 39
20 − 29
10 − 19
Age group
10 − 19
20 − 29
30 − 39
40 − 49
50 − 59
60 − 69
70 − 79
80+
Age group
Contact
6
5
4
3
2
1
0


52

```
##
## [[33]]
```

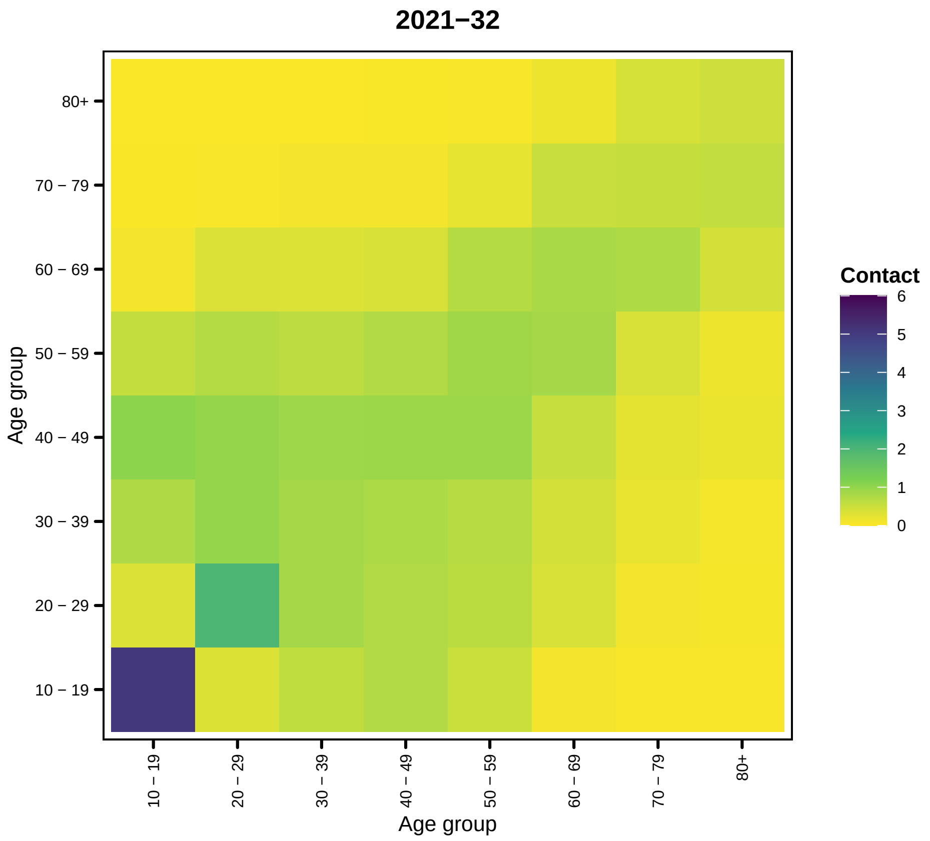
2021−32
Age group
80+
70 − 79
60 − 69
50 − 59
40 − 49
30 − 39
20 − 29
10 − 19
Contact
6
5
4
3
2
1
0

53

```
##
## [[34]]
```

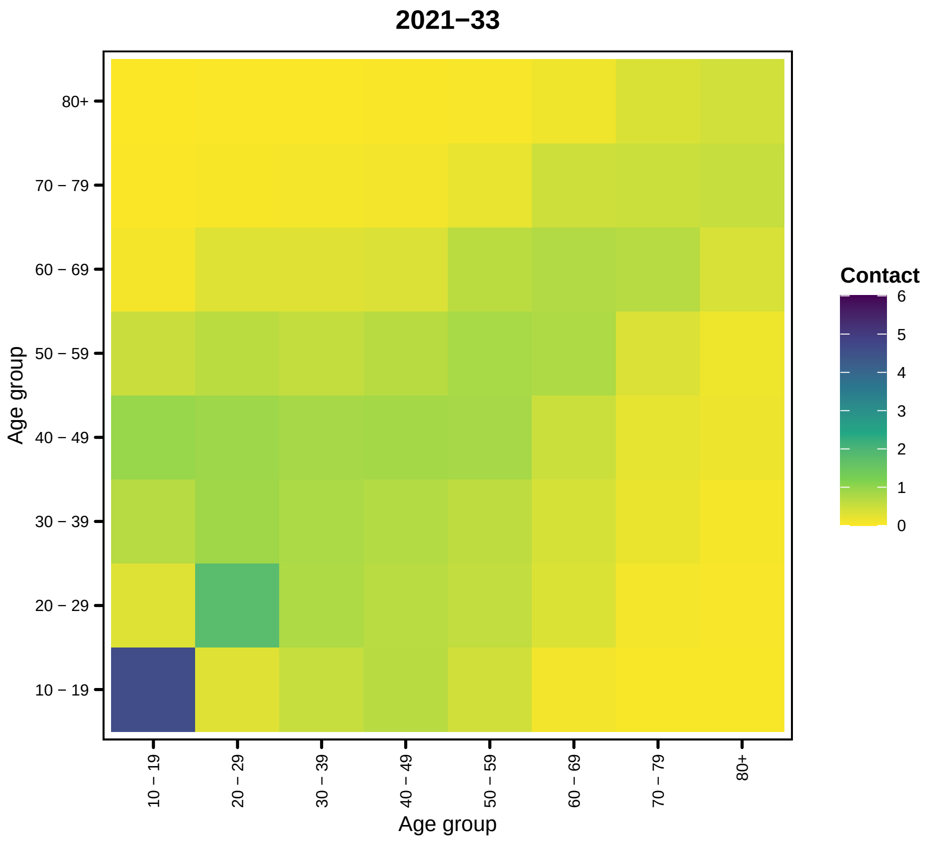
2021−33
80+
70 − 79
60 − 69
50 − 59
40 − 49
30 − 39
20 − 29
10 − 19
Age group
10 − 19
20 − 29
30 − 39
40 − 49
50 − 59
60 − 69
70 − 79
80+
Age group
Contact
6
5
4
3
2
1
0

54

```
##
## [[35]]
```

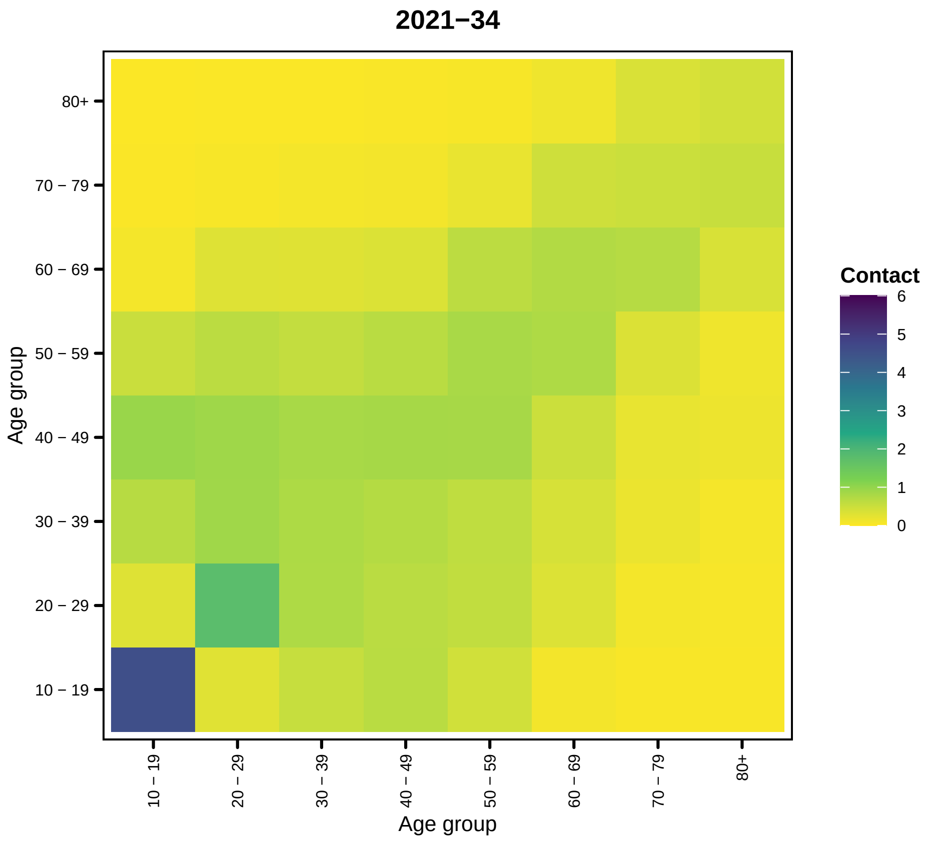
2021−34
Contact
6
5
4
3
2
1
0
80+
70 − 79
60 − 69
50 − 59
40 − 49
30 − 39
20 − 29
10 − 19
Age group
Age group

55

```
##
## [[36]]
```

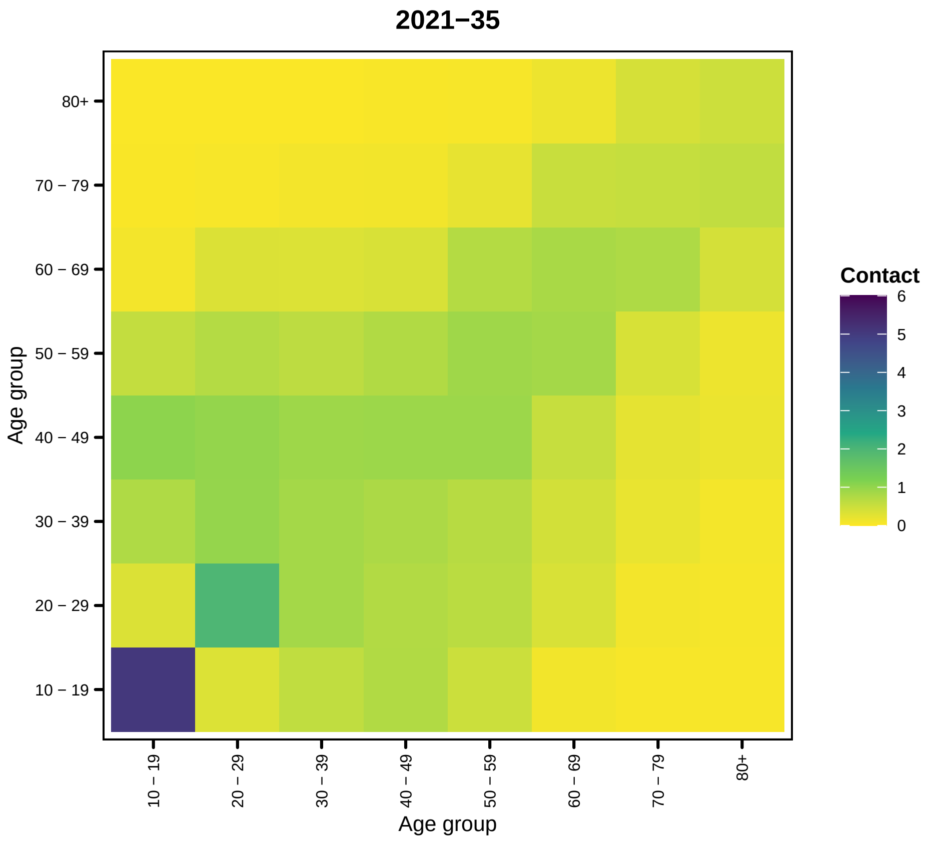
2021−35
Contact
6
5
4
3
2
1
0
80+
70 − 79
60 − 69
50 − 59
40 − 49
30 − 39
20 − 29
10 − 19
Age group
Age group

56

```
##
## [[37]]
```

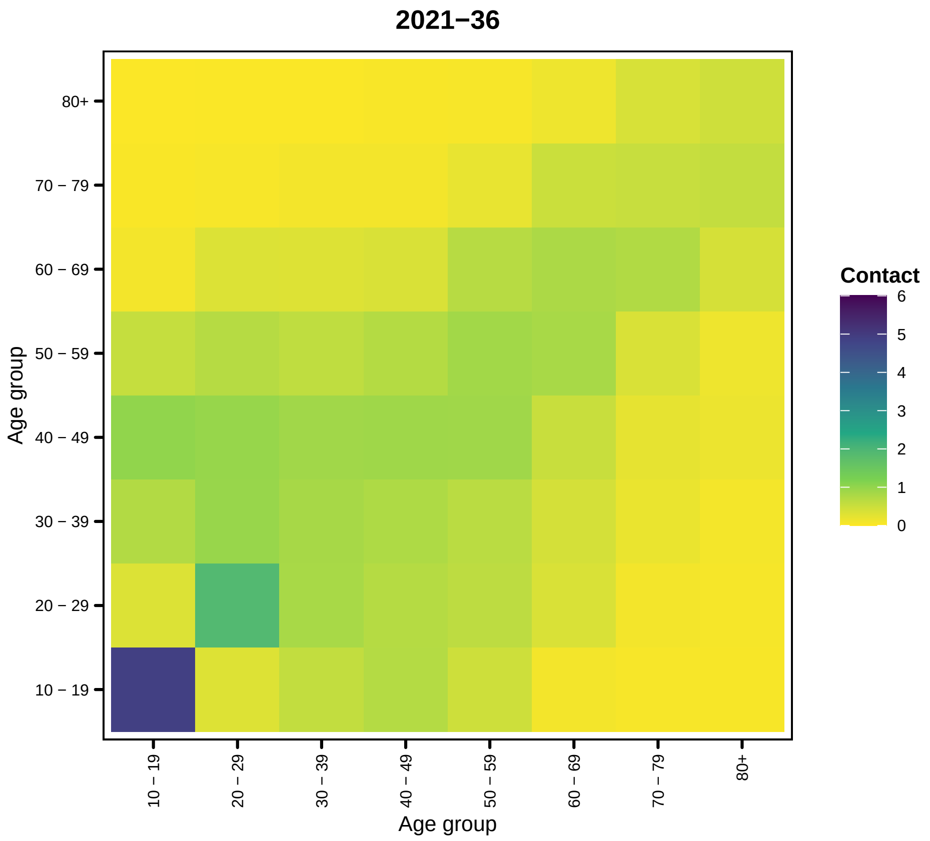
2021−36
80+
70 − 79
60 − 69
50 − 59
40 − 49
30 − 39
20 − 29
10 − 19
Age group
10 − 19
20 − 29
30 − 39
40 − 49
50 − 59
60 − 69
70 − 79
80+
Age group
Contact
6
5
4
3
2
1
0

57

```
##
## [[38]]
```

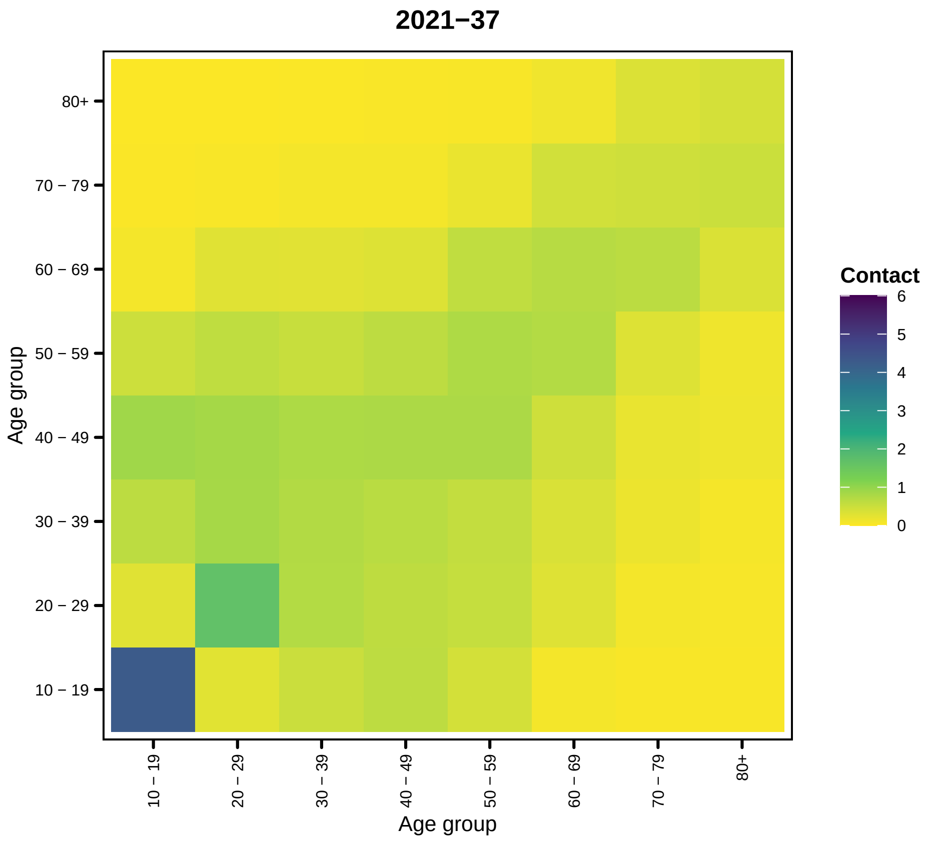
2021−37
Age group
80+
70 − 79
60 − 69
50 − 59
40 − 49
30 − 39
20 − 29
10 − 19
Contact
6
5
4
3
2
1
0

58

```
##
## [[39]]
```

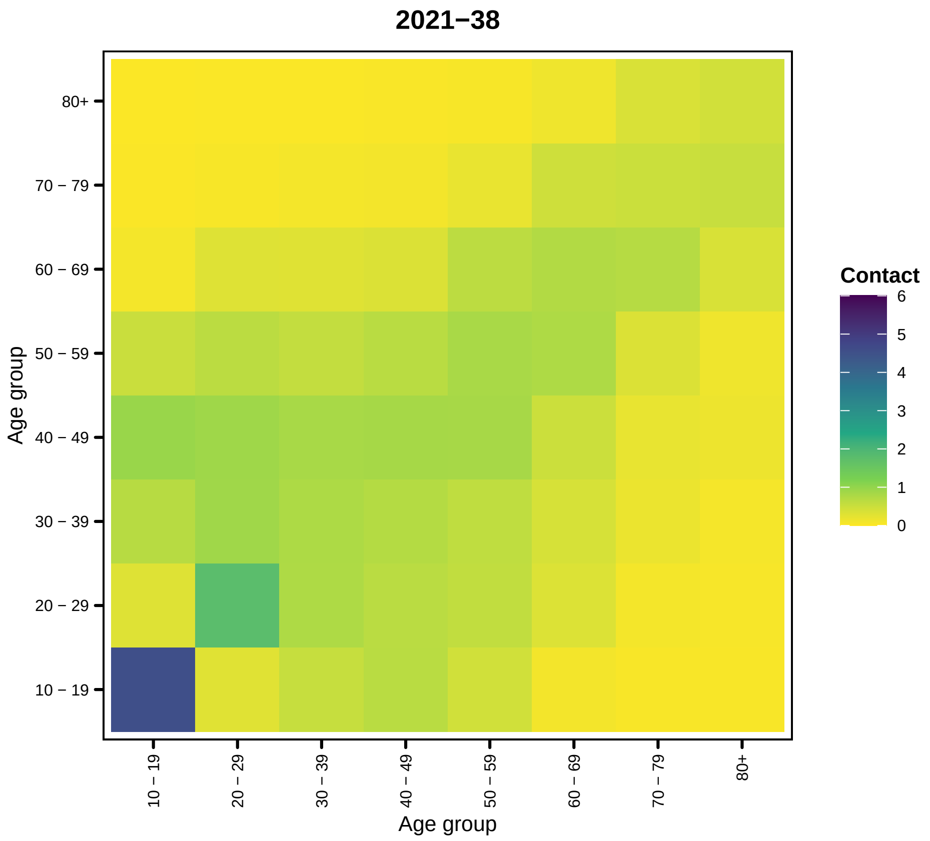
2021−38
80+
70 − 79
60 − 69
50 − 59
40 − 49
30 − 39
20 − 29
10 − 19
Age group
Contact
6
5
4
3
2
1
0
10 − 19
20 − 29
30 − 39
40 − 49
50 − 59
60 − 69
70 − 79
80+
Age group

59

```
##
## [[40]]
```

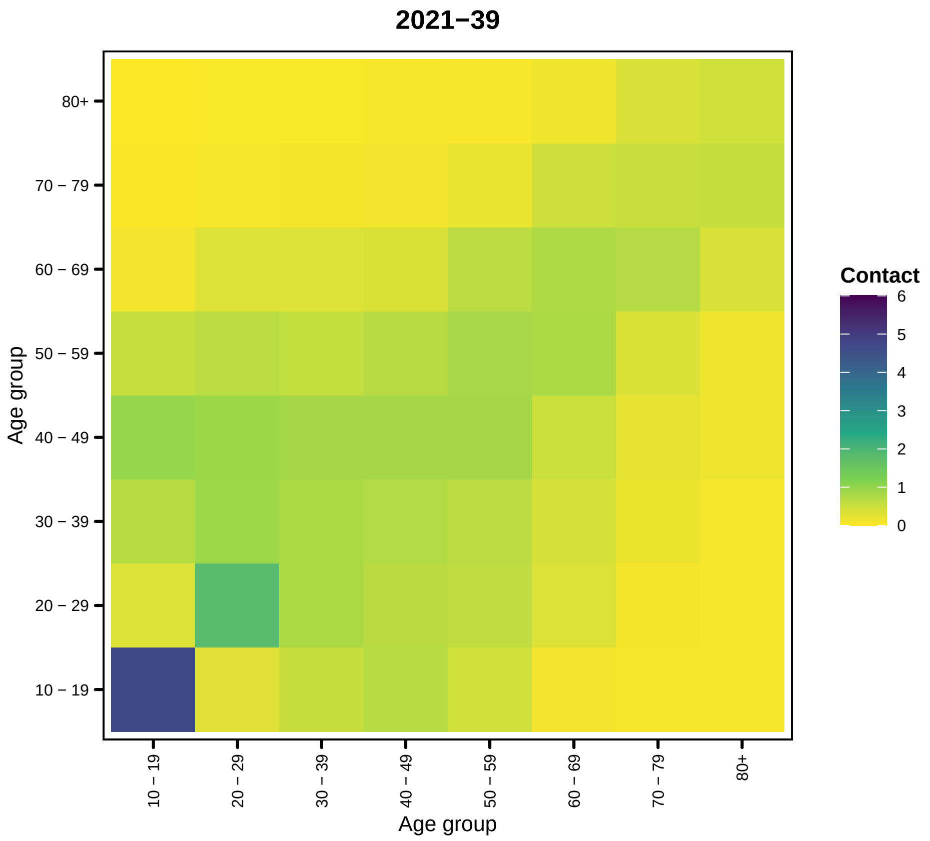
2021−39
Age group
80+
70 − 79
60 − 69
50 − 59
40 − 49
30 − 39
20 − 29
10 − 19
Contact
6
5
4
3
2
1
0

60

```
##
## [[41]]
```

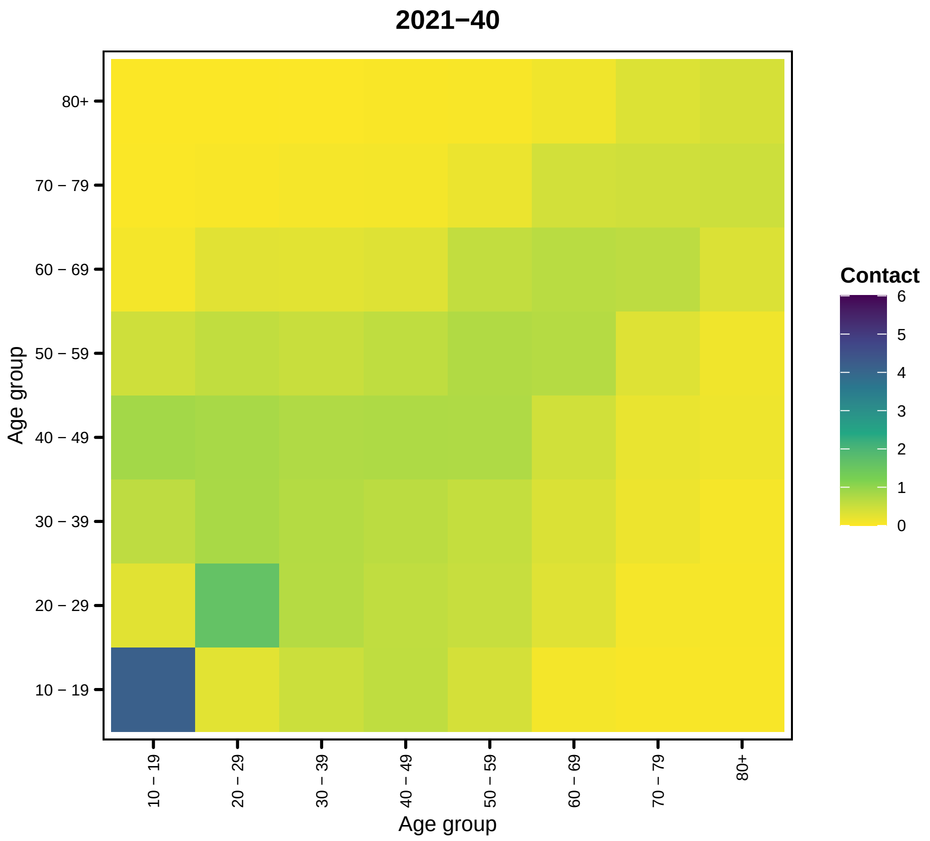

2021−40
80+
70 − 79
60 − 69
50 − 59
40 − 49
30 − 39
20 − 29
10 − 19
Age group
Contact
6
5
4
3
2
1
0


61

```
##
## [[42]]
```

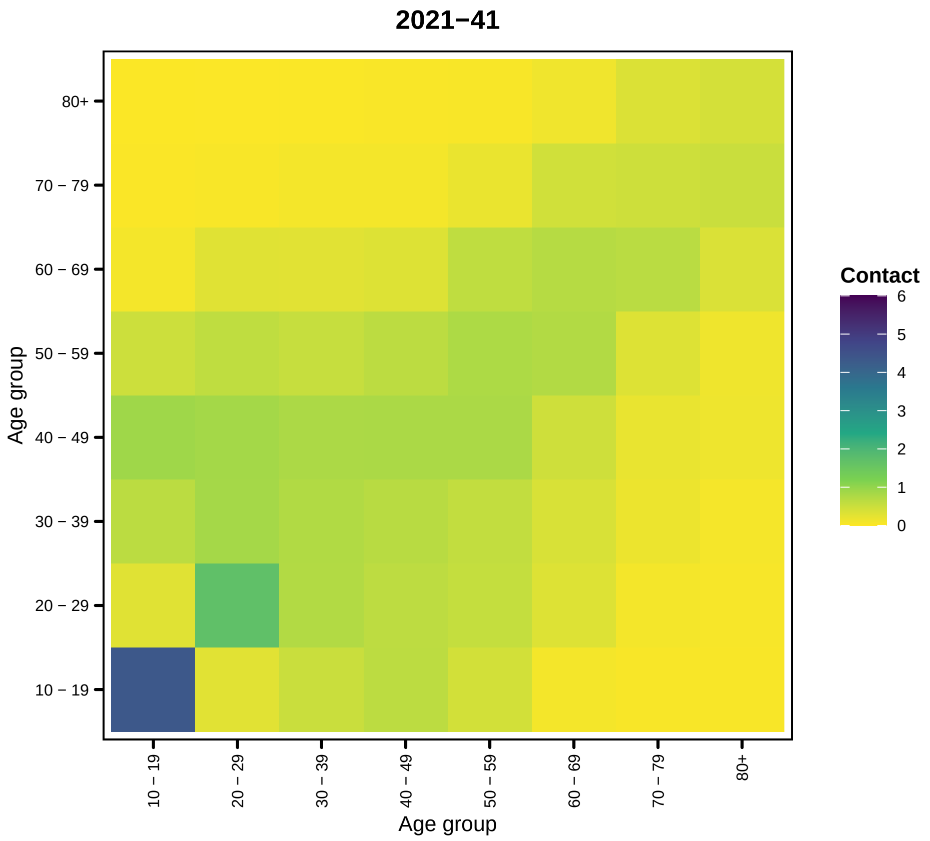
2021−41
Age group
80+
70 − 79
60 − 69
50 − 59
40 − 49
30 − 39
20 − 29
10 − 19
Contact
6
5
4
3
2
1
0

```
##
## [[43]]
```

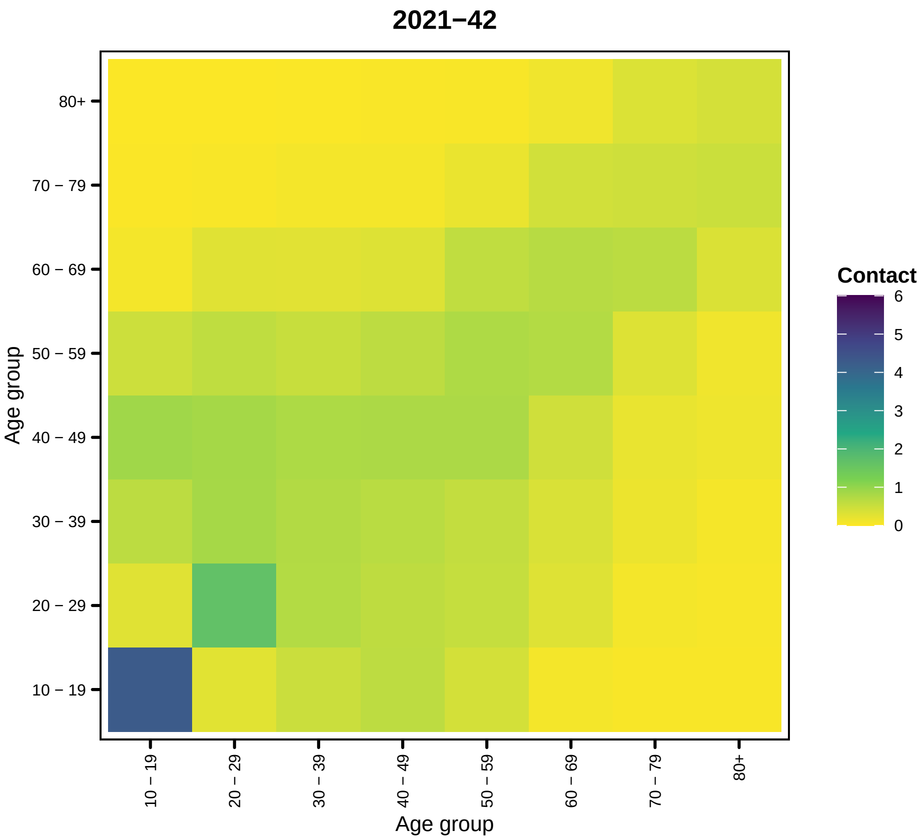
2021−42
Age group
80+
70 − 79
60 − 69
50 − 59
40 − 49
30 − 39
20 − 29
10 − 19
Contact
6
5
4
3
2
1
0

63

```
##
## [[44]]
```

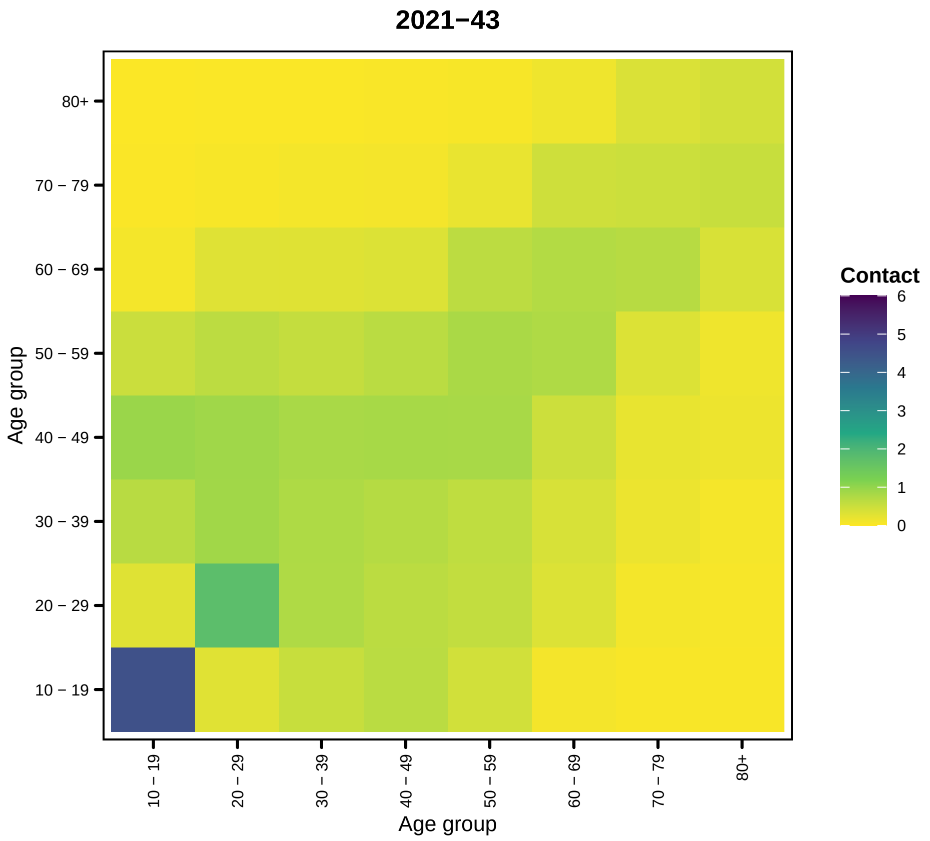
2021−43
80+
70 − 79
60 − 69
50 − 59
40 − 49
30 − 39
20 − 29
10 − 19
Age group
10 − 19
20 − 29
30 − 39
40 − 49
50 − 59
60 − 69
70 − 79
80+
Age group
Contact
6
5
4
3
2
1
0

64

```
##
## [[45]]
```

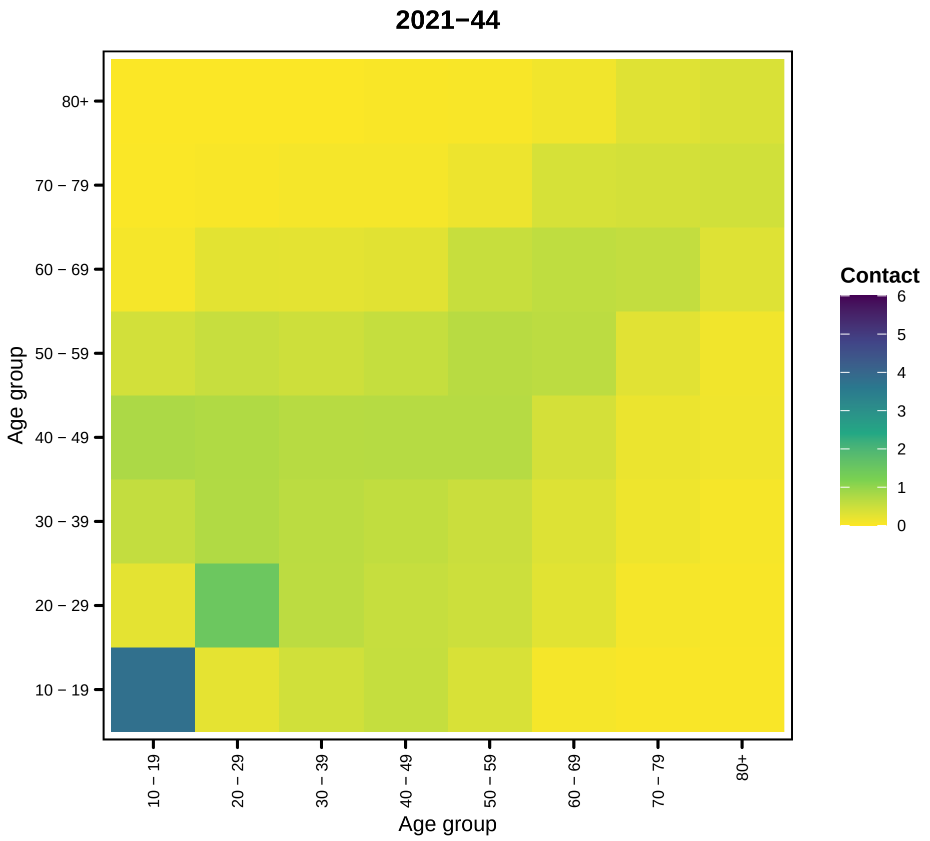
2021−44
80+
70 − 79
60 − 69
50 − 59
40 − 49
30 − 39
20 − 29
10 − 19
Age group
Contact
6
5
4
3
2
1
0
10 − 19
20 − 29
30 − 39
40 − 49
50 − 59
60 − 69
70 − 79
80+
Age group

65

```
##
## [[46]]
```

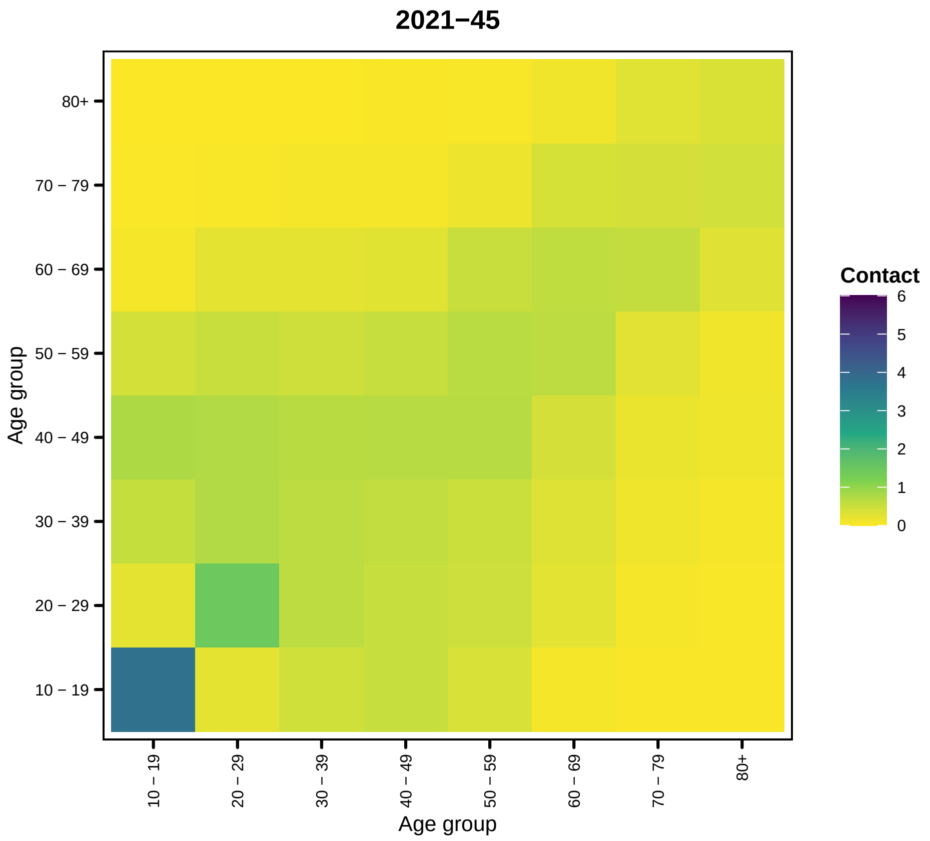

2021−45
80+
70 − 79
60 − 69
50 − 59
40 − 49
30 − 39
20 − 29
10 − 19
Age group
Contact
6
5
4
3
2
1
0
10 − 19
20 − 29
30 − 39
40 − 49
50 − 59
60 − 69
70 − 79
80+
Age group


66

```
##
## [[47]]
```

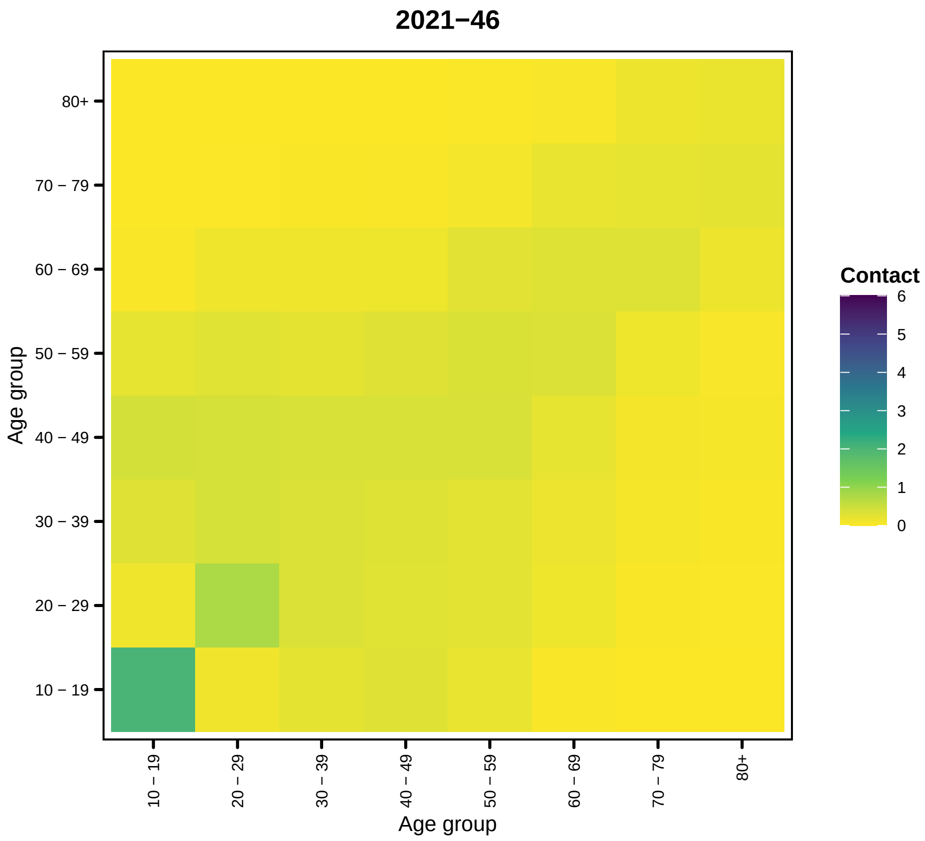
2021−46
80+
70 − 79
60 − 69
50 − 59
40 − 49
30 − 39
20 − 29
10 − 19
Age group
Age group
Contact
6
5
4
3
2
1
0

67

```
##
## [[48]]
```

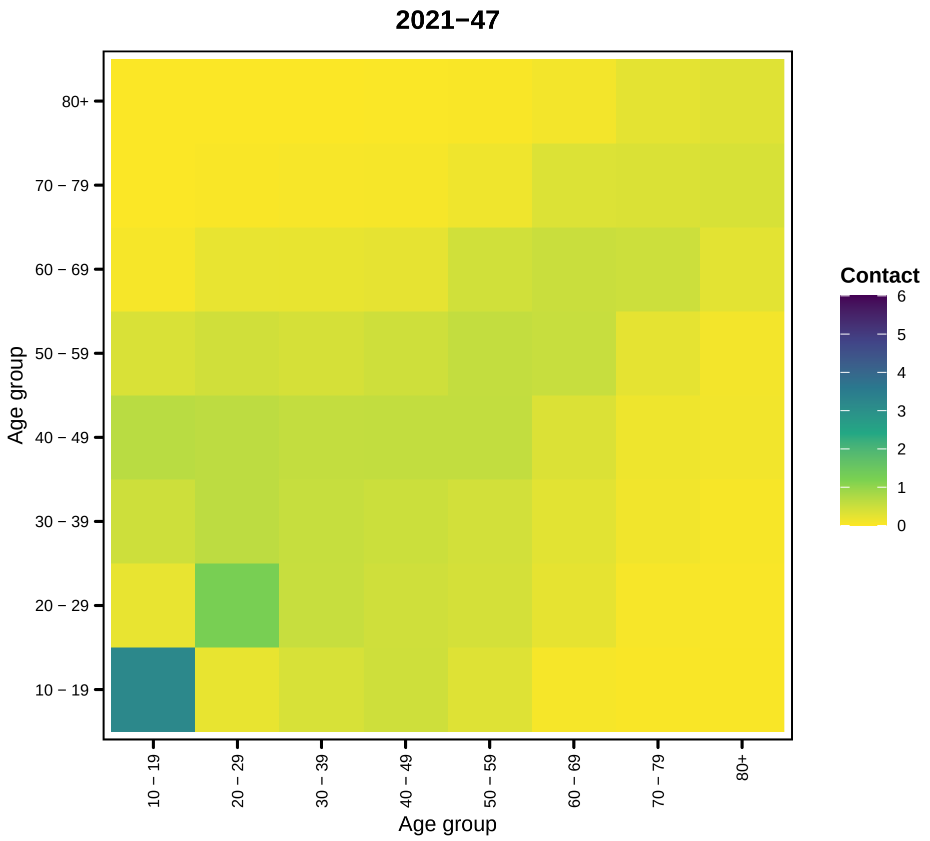
2021−47
80+
70 − 79
60 − 69
50 − 59
40 − 49
30 − 39
20 − 29
10 − 19
Age group
10 − 19
20 − 29
30 − 39
40 − 49
50 − 59
60 − 69
70 − 79
80+
Age group
Contact
6
5
4
3
2
1
0

68

```
##
## [[49]]
```

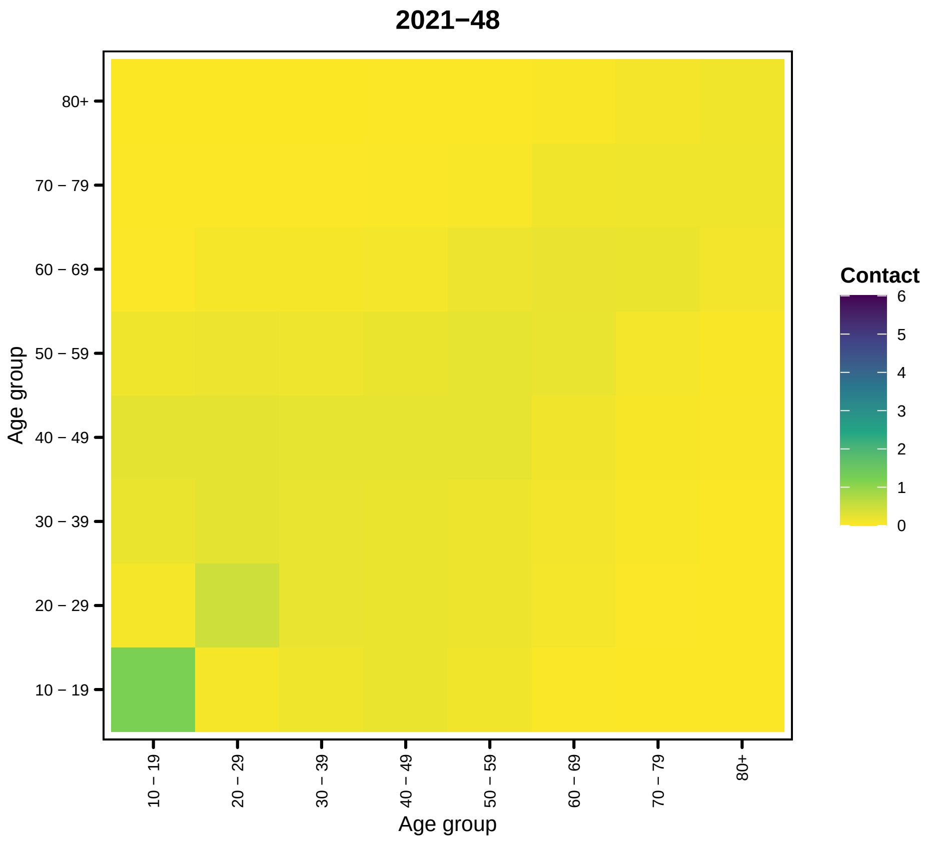
2021−48
80+
70 − 79
60 − 69
50 − 59
40 − 49
30 − 39
20 − 29
10 − 19
Age group
10 − 19
20 − 29
30 − 39
40 − 49
50 − 59
60 − 69
70 − 79
80+
Age group
Contact
6
5
4
3
2
1
0

69

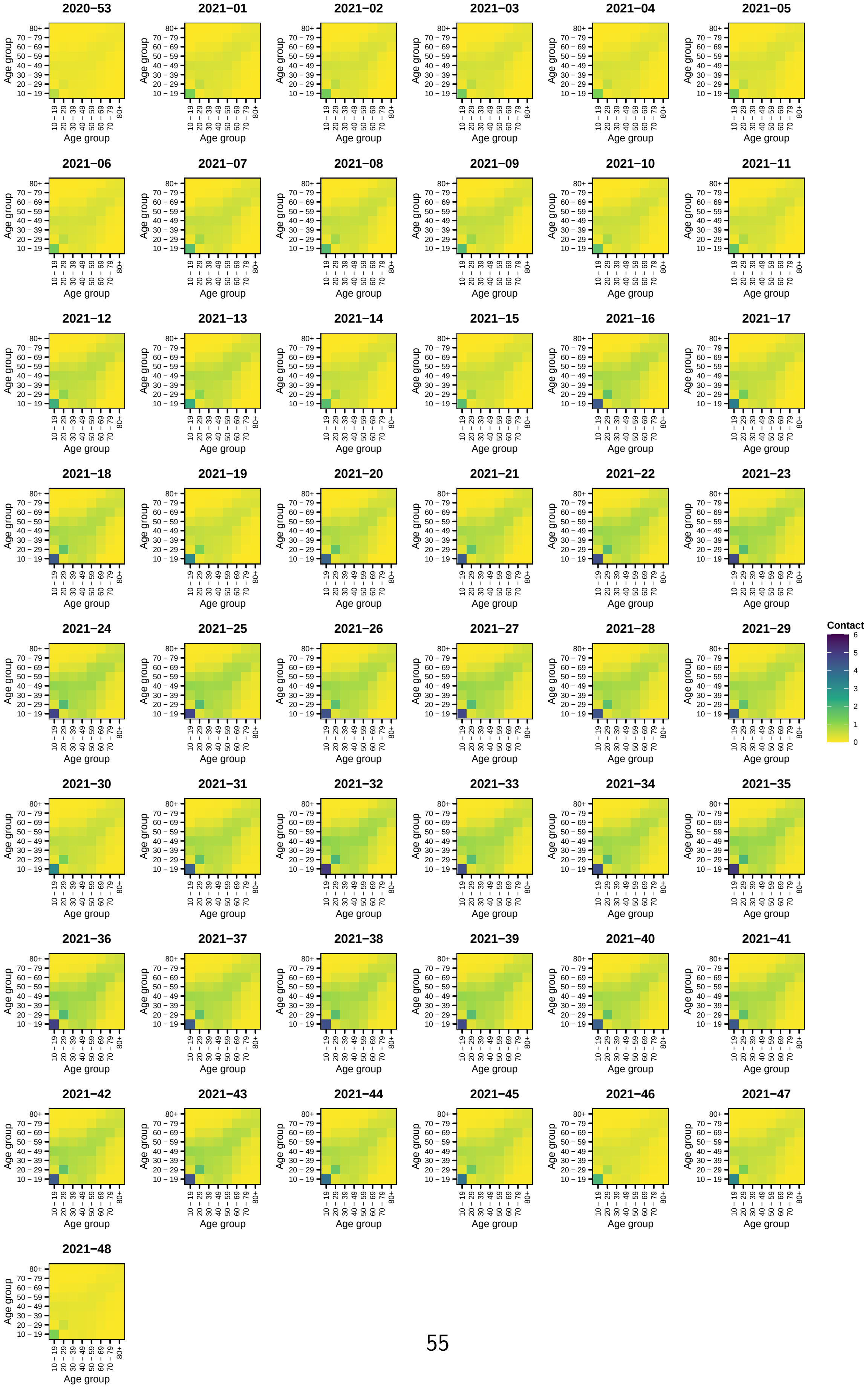
2020–53
2021–01
2021–02
2021–03
2021–04
2021–05
2021–06
2021–07
2021–08
2021–09
2021–10
2021–11
2021–12
2021–13
2021–14
2021–15
2021–16
2021–17
2021–18
2021–19
2021–20
2021–21
2021–22
2021–23
2021–24
2021–25
2021–26
2021–27
2021–28
2021–29
2021–30
2021–31
2021–32
2021–33
2021–34
2021–35
2021–36
2021–37
2021–38
2021–39
2021–40
2021–41
2021–42
2021–43
2021–44
2021–45
2021–46
2021–47
2021–48
Age group
10 – 19
20 – 29
30 – 39
40 – 49
50 – 59
60 – 69
70 – 79
80+
Contact
0
1
2
3
4
5
6

### 4 Dispersion

The time-varying adjacency matrices are plotted below

```
## [[1]]
```

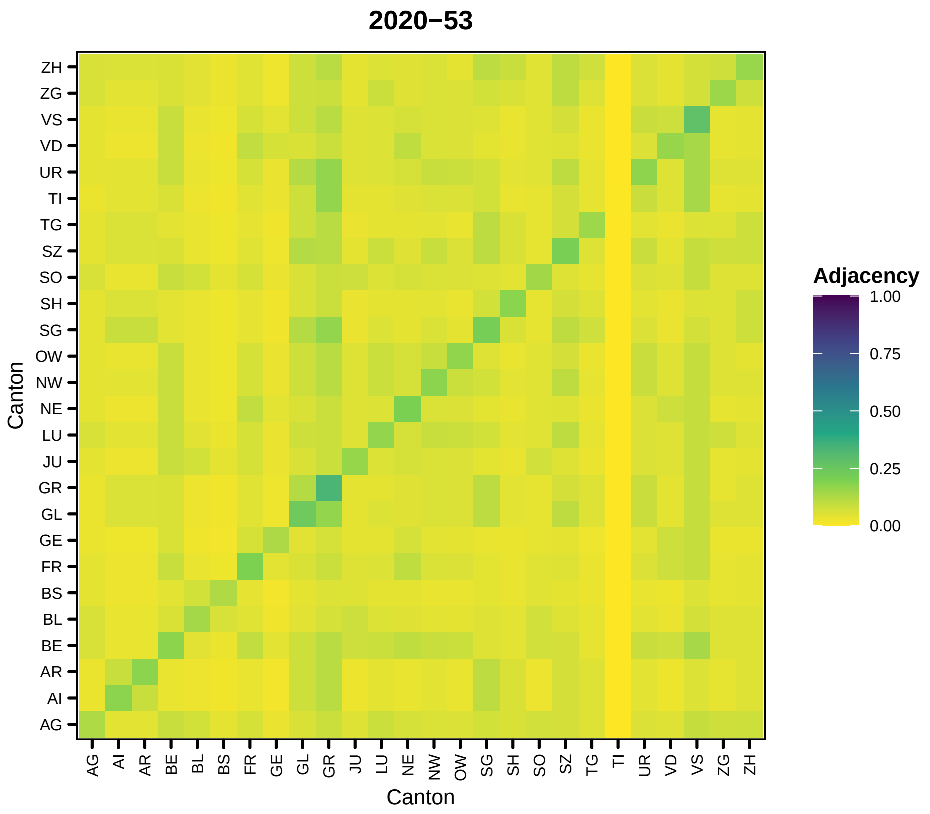


```
##
## [[2]]
```

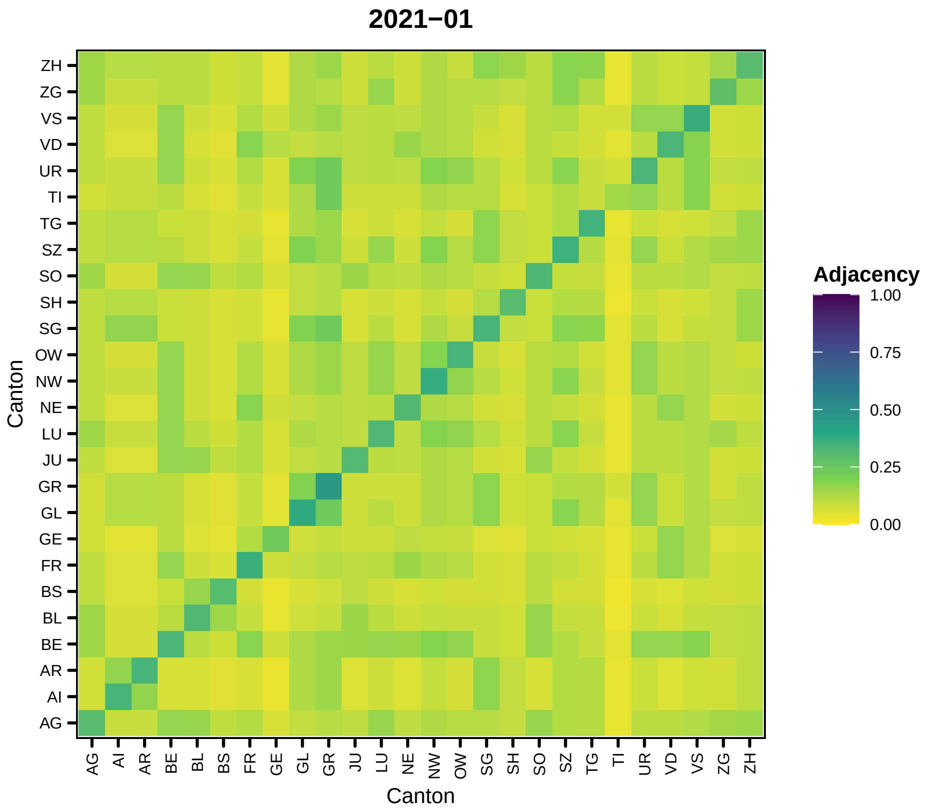
2021−01
Adjacency
1.00
0.75
0.50
0.25
0.00
ZH
ZG
VS
VD
UR
TI
TG
SZ
SO
SH
SG
OW
NW
NE
LU
JU
GR
GL
GE
FR
BS
BL
BE
AR
AI
AG
Canton
AG AI AR BE BL BS FR GE GL GR JU LU NE NW OW SG SH SO SZ TG TI UR VD VS ZG ZH
Canton

74

```
##
## [[3]]
```

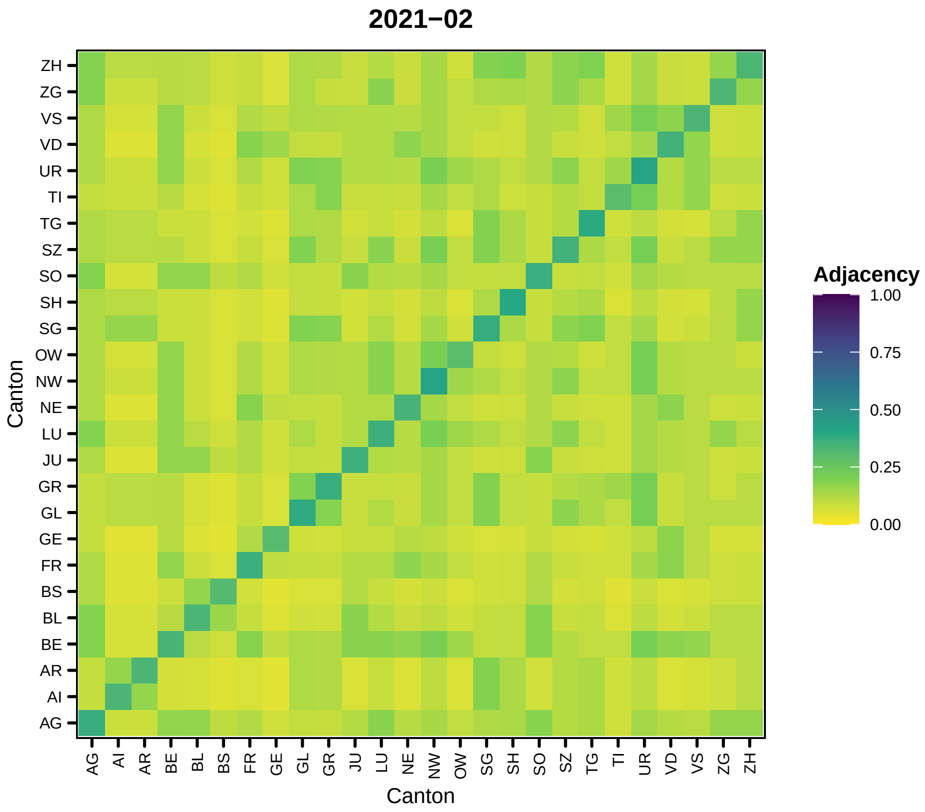
2021−02
Canton
ZH
ZG
VS
VD
UR
TI
TG
SZ
SO
SH
SG
OW
NW
NE
LU
JU
GR
GL
GE
FR
BS
BL
BE
AR
AI
AG
AG
AI
AR
BE
BL
BS
FR
GE
GL
GR
JU
LU
NE
NW
OW
SG
SH
SO
SZ
TG
TI
UR
VD
VS
ZG
ZH
Canton
Adjacency
1.00
0.75
0.50
0.25
0.00

75

```
##
## [[4]]
```

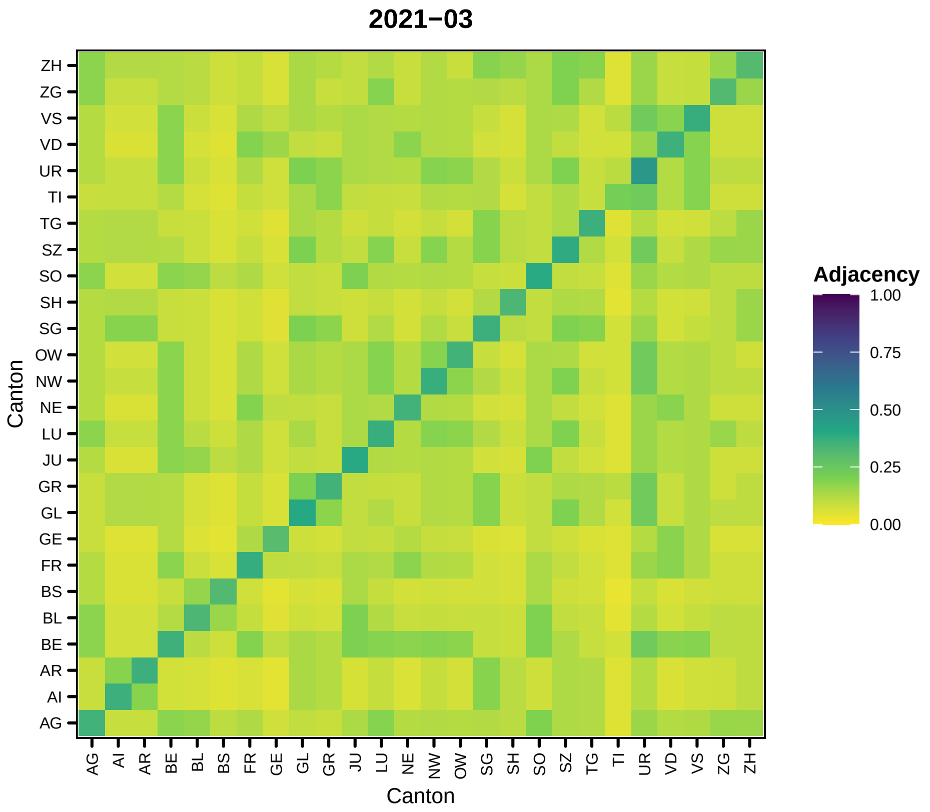

2021−03
Adjacency
1.00
0.75
0.50
0.25
0.00
Canton
Canton
ZH
ZG
VS
VD
UR
TI
TG
SZ
SO
SH
SG
OW
NW
NE
LU
JU
GR
GL
GE
FR
BS
BL
BE
AR
AI
AG


76

```
##
## [[5]]
```

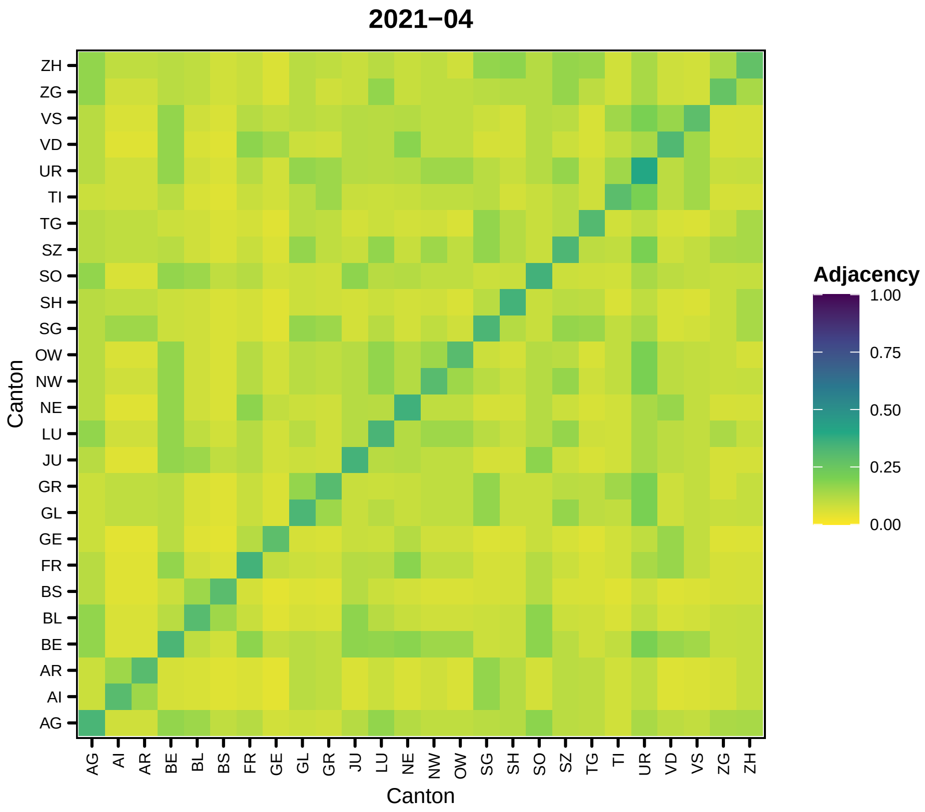
2021−04
Adjacency
1.00
0.75
0.50
0.25
0.00
Canton
Canton
ZH
ZG
VS
VD
UR
TI
TG
SZ
SO
SH
SG
OW
NW
NE
LU
JU
GR
GL
GE
FR
BS
BL
BE
AR
AI
AG

77

```
##
## [[6]]
```

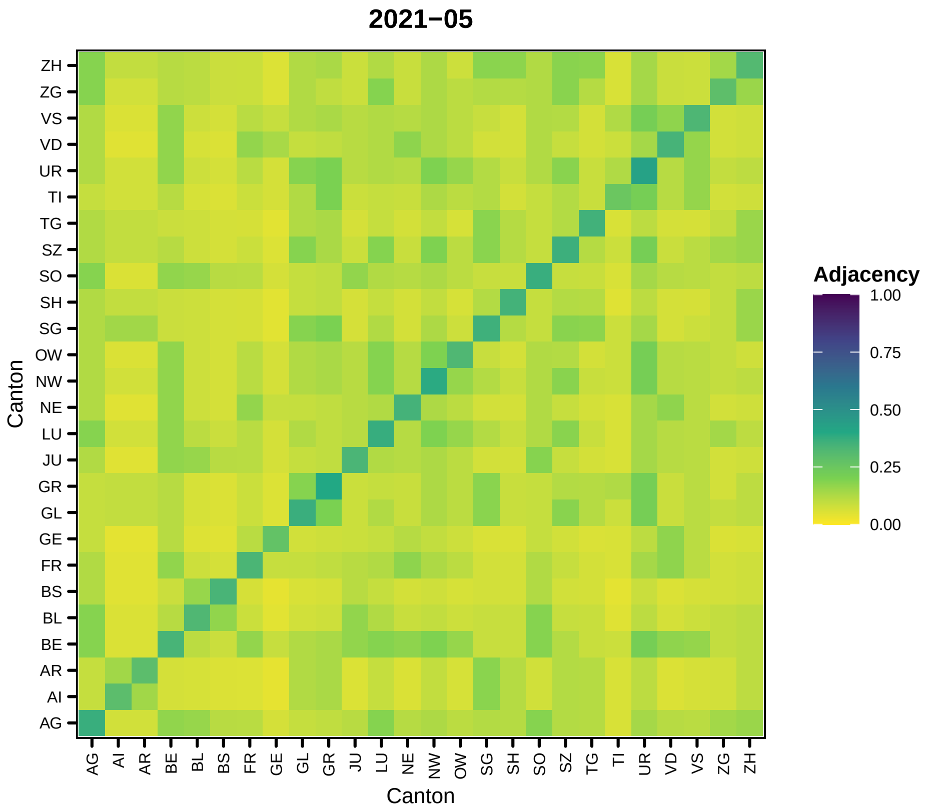
2021−05
Canton
ZH
ZG
VS
VD
UR
TI
TG
SZ
SO
SH
SG
OW
NW
NE
LU
JU
GR
GL
GE
FR
BS
BL
BE
AR
AI
AG
Adjacency
1.00
0.75
0.50
0.25
0.00

78

```
##
## [[7]]
```

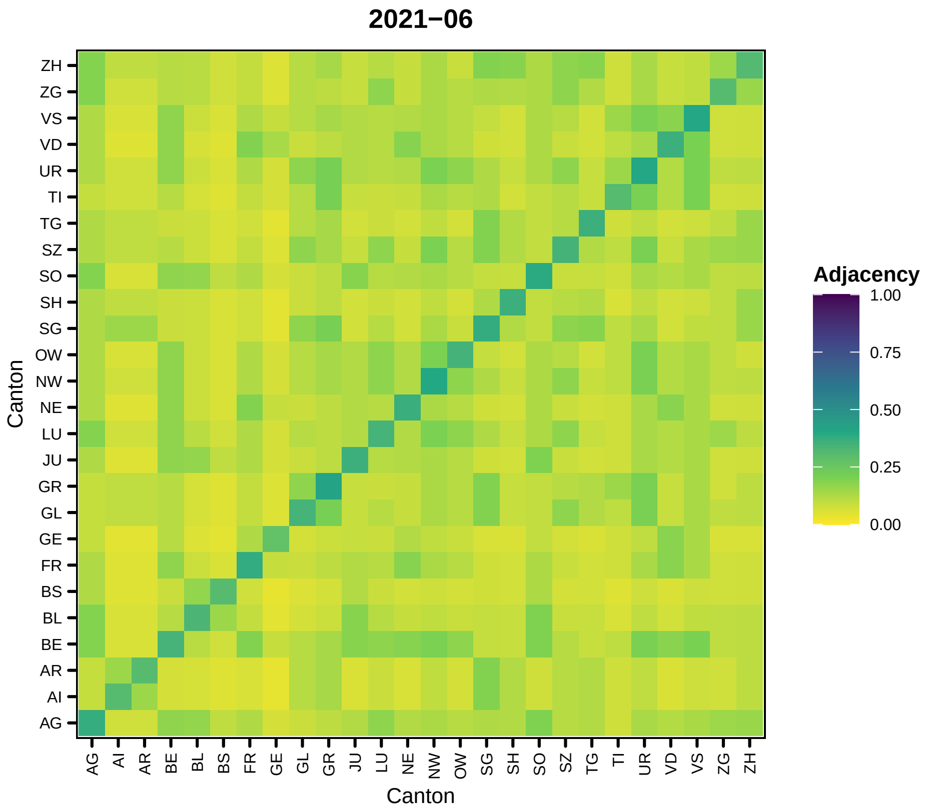
2021−06
Canton
ZH
ZG
VS
VD
UR
TI
TG
SZ
SO
SH
SG
OW
NW
NE
LU
JU
GR
GL
GE
FR
BS
BL
BE
AR
AI
AG
AG AI AR BE BL BS FR GE GL GR JU LU NE NW OW SG SH SO SZ TG TI UR VD VS ZG ZH
Canton
Adjacency
1.00
0.75
0.50
0.25
0.00

79

```
##
## [[8]]
```

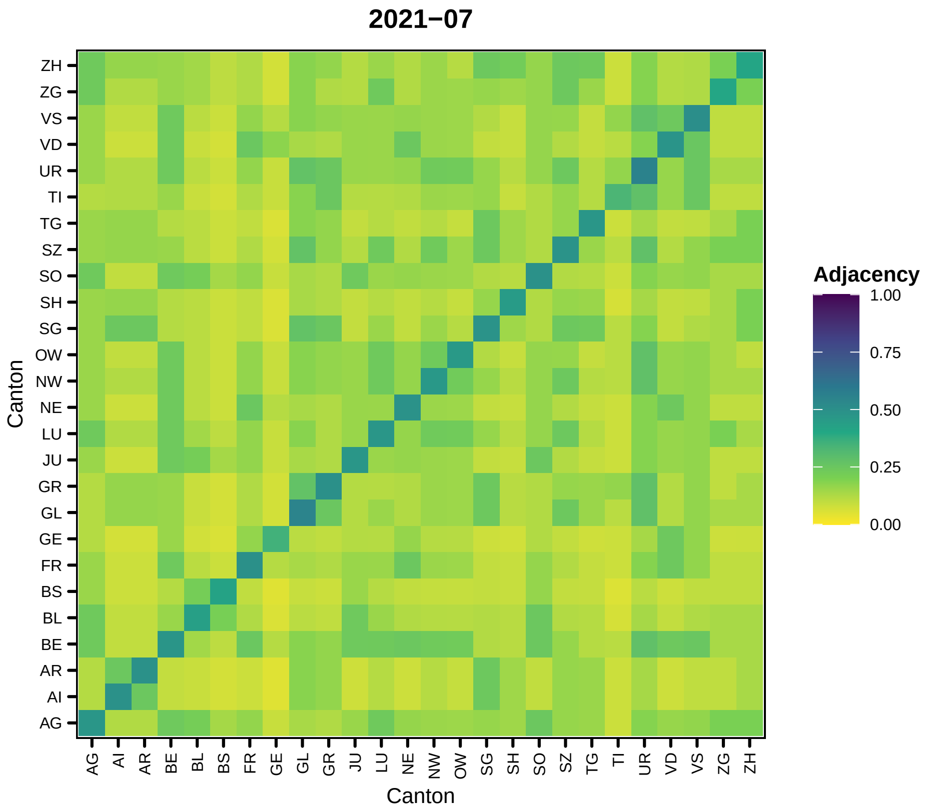
2021−07
Canton
Canton
Adjacency
1.00
0.75
0.50
0.25
0.00
ZH
ZG
VS
VD
UR
TI
TG
SZ
SO
SH
SG
OW
NW
NE
LU
JU
GR
GL
GE
FR
BS
BL
BE
AR
AI
AG

80

```
##

## [[9]]
```

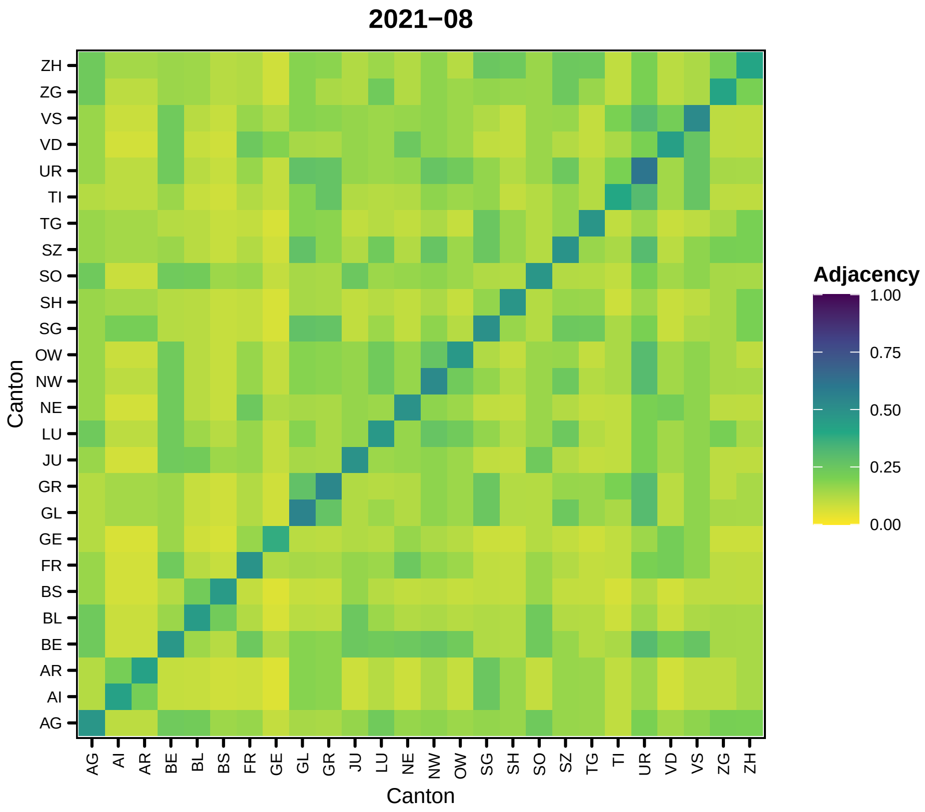
2021−08
Adjacency
1.00
0.75
0.50
0.25
0.00
Canton
Canton
ZH
ZG
VS
VD
UR
TI
TG
SZ
SO
SH
SG
OW
NW
NE
LU
JU
GR
GL
GE
FR
BS
BL
BE
AR
AI
AG

81

```
##
## [[10]]
```

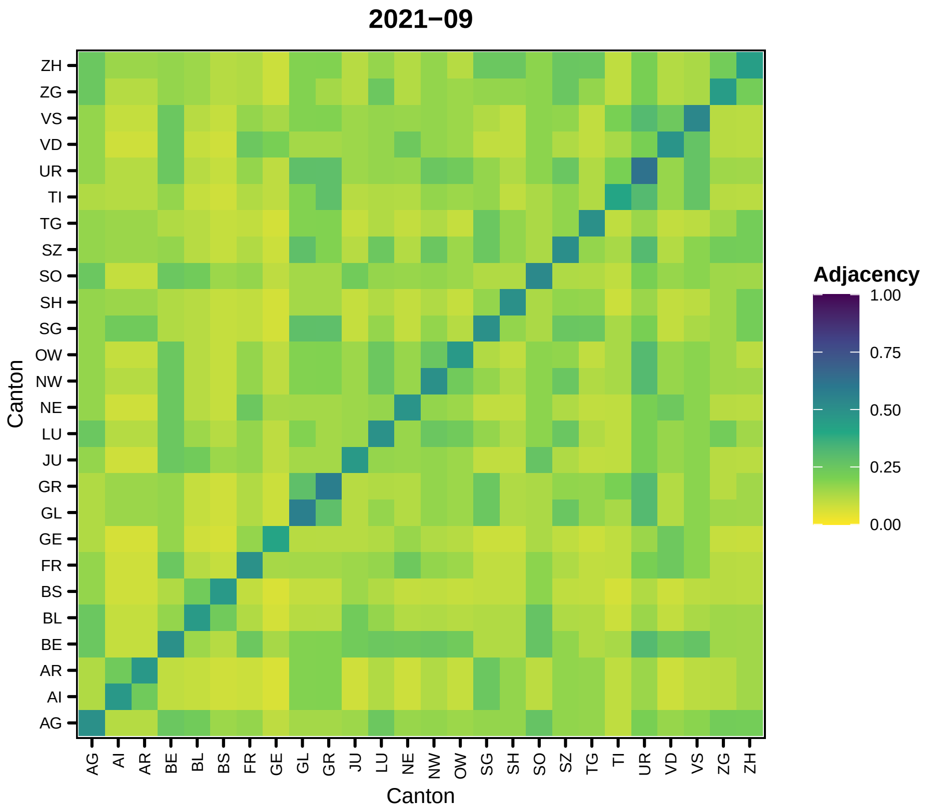
2021−09
Adjacency
1.00
0.75
0.50
0.25
0.00
Canton
Canton
ZH
ZG
VS
VD
UR
TI
TG
SZ
SO
SH
SG
OW
NW
NE
LU
JU
GR
GL
GE
FR
BS
BL
BE
AR
AI
AG

82

```
##
## [[11]]
```

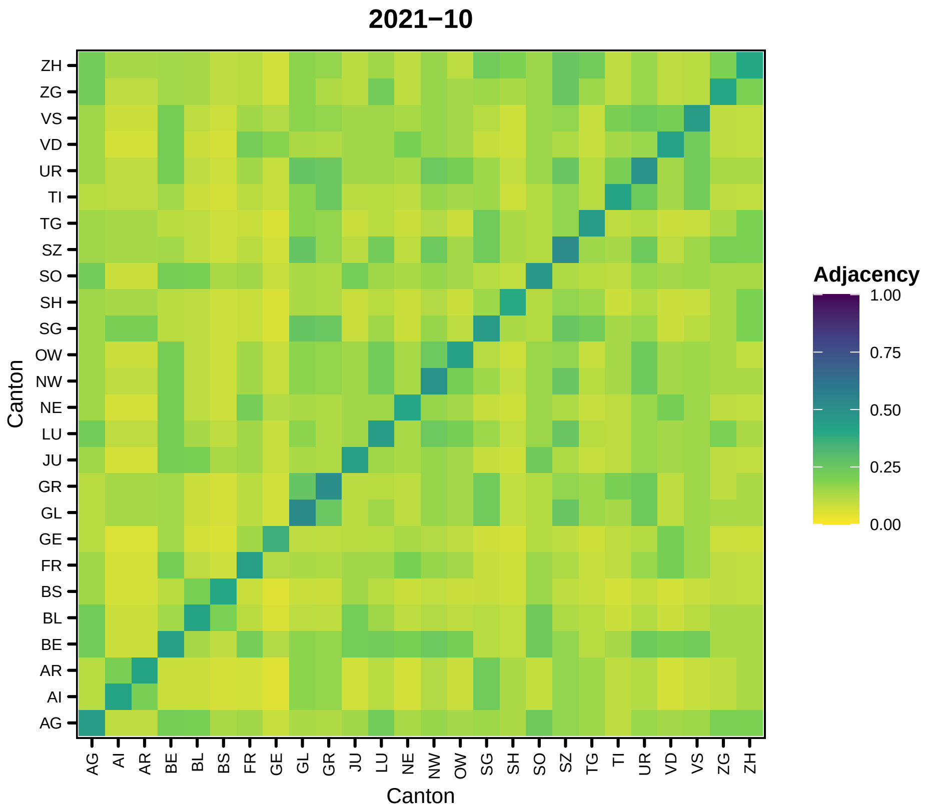

2021−10
Adjacency
1.00
0.75
0.50
0.25
0.00
Canton
Canton
ZH
ZG
VS
VD
UR
TI
TG
SZ
SO
SH
SG
OW
NW
NE
LU
JU
GR
GL
GE
FR
BS
BL
BE
AR
AI
AG


83

```
##
## [[12]]
```

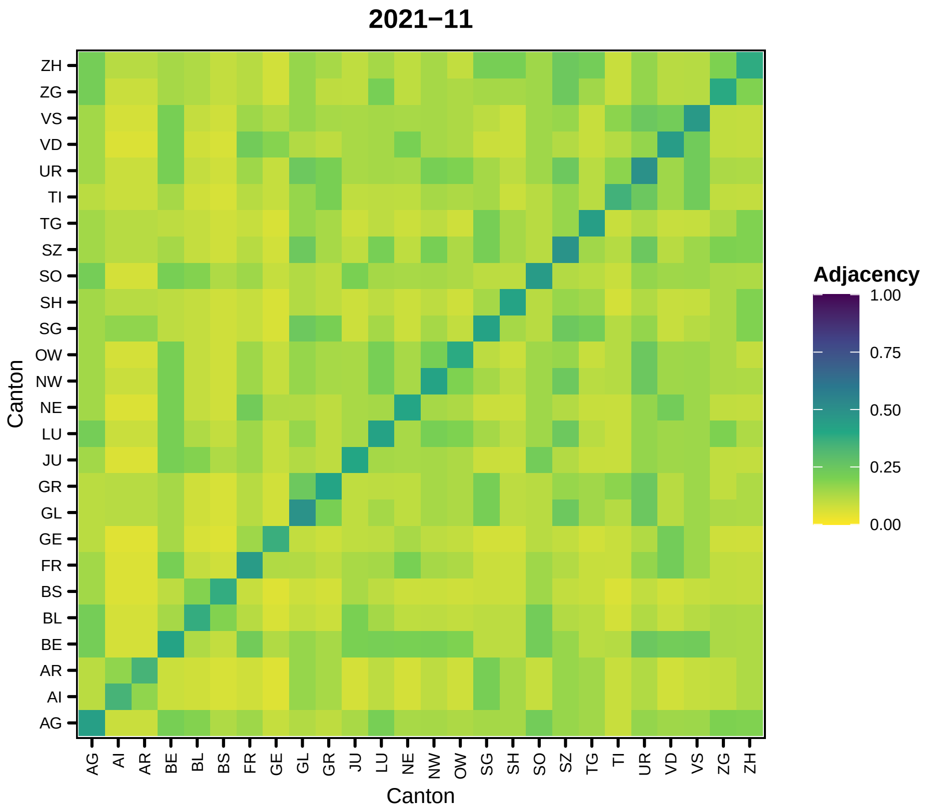
2021−11
Canton
ZH
ZG
VS
VD
UR
TI
TG
SZ
SO
SH
SG
OW
NW
NE
LU
JU
GR
GL
GE
FR
BS
BL
BE
AR
AI
AG
AG AI AR BE BL BS FR GE GL GR JU LU NE NW OW SG SH SO SZ TG TI UR VD VS ZG ZH
Adjacency
1.00
0.75
0.50
0.25
0.00

84

```
##
## [[13]]
```

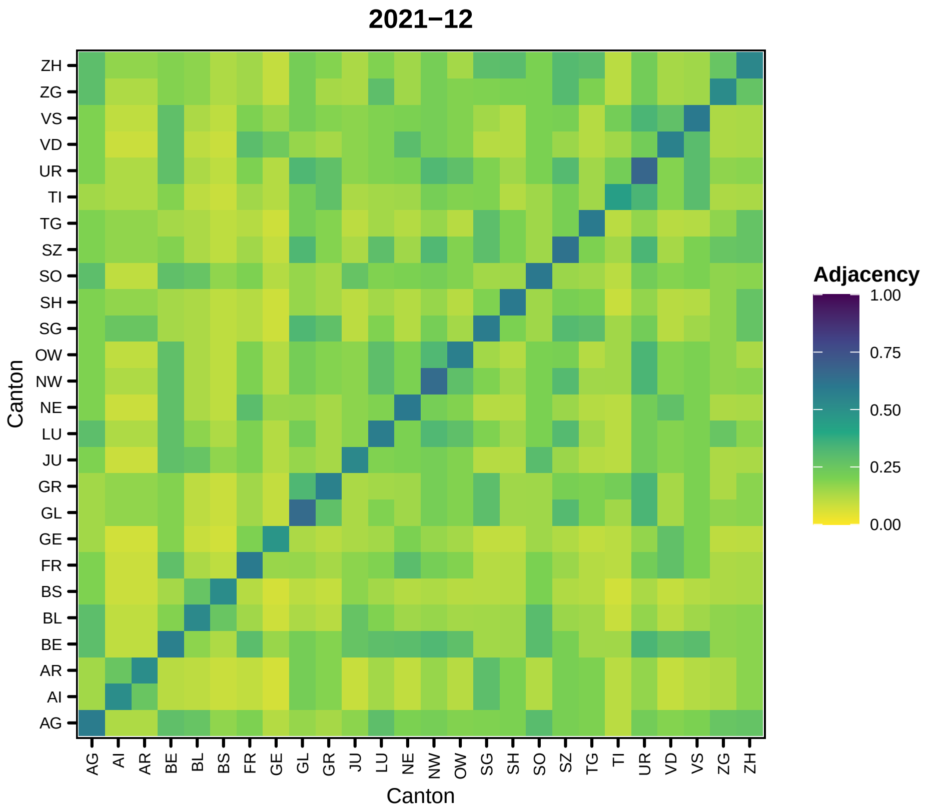
2021−12
Adjacency
1.00
0.75
0.50
0.25
0.00
Canton
Canton
ZH
ZG
VS
VD
UR
TI
TG
SZ
SO
SH
SG
OW
NW
NE
LU
JU
GR
GL
GE
FR
BS
BL
BE
AR
AI
AG

85

```
##
## [[14]]
```

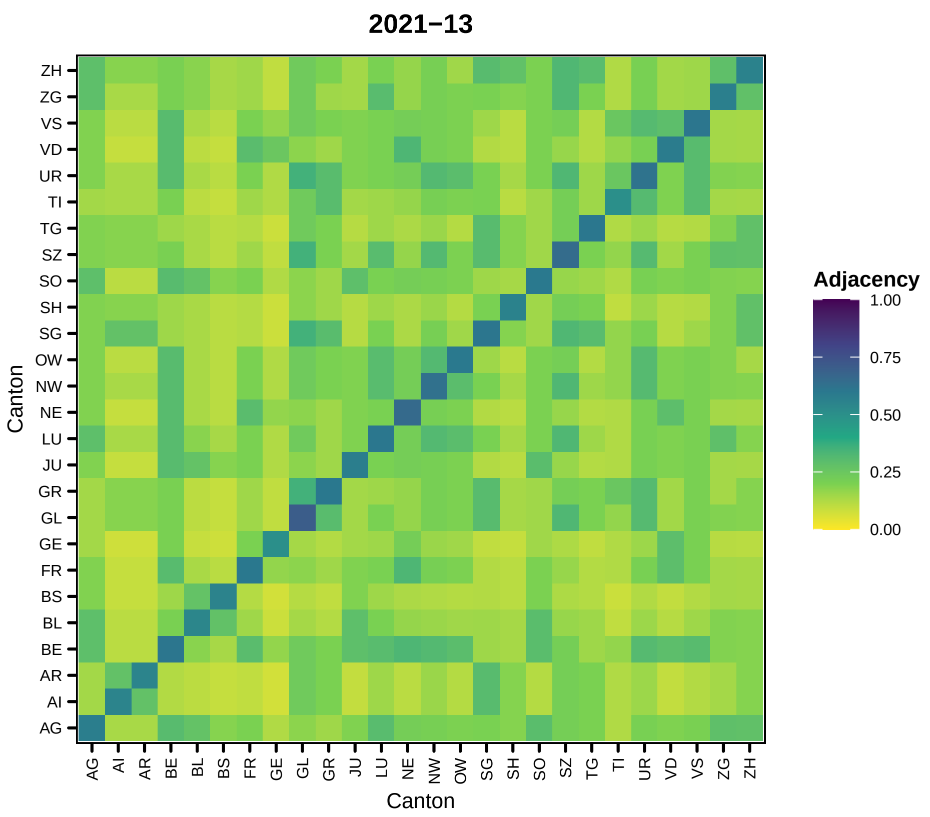
2021−13
Canton
Adjacency
1.00
0.75
0.50
0.25
0.00
ZH
ZG
VS
VD
UR
TI
TG
SZ
SO
SH
SG
OW
NW
NE
LU
JU
GR
GL
GE
FR
BS
BL
BE
AR
AI
AG

86

```
##
## [[15]]
```

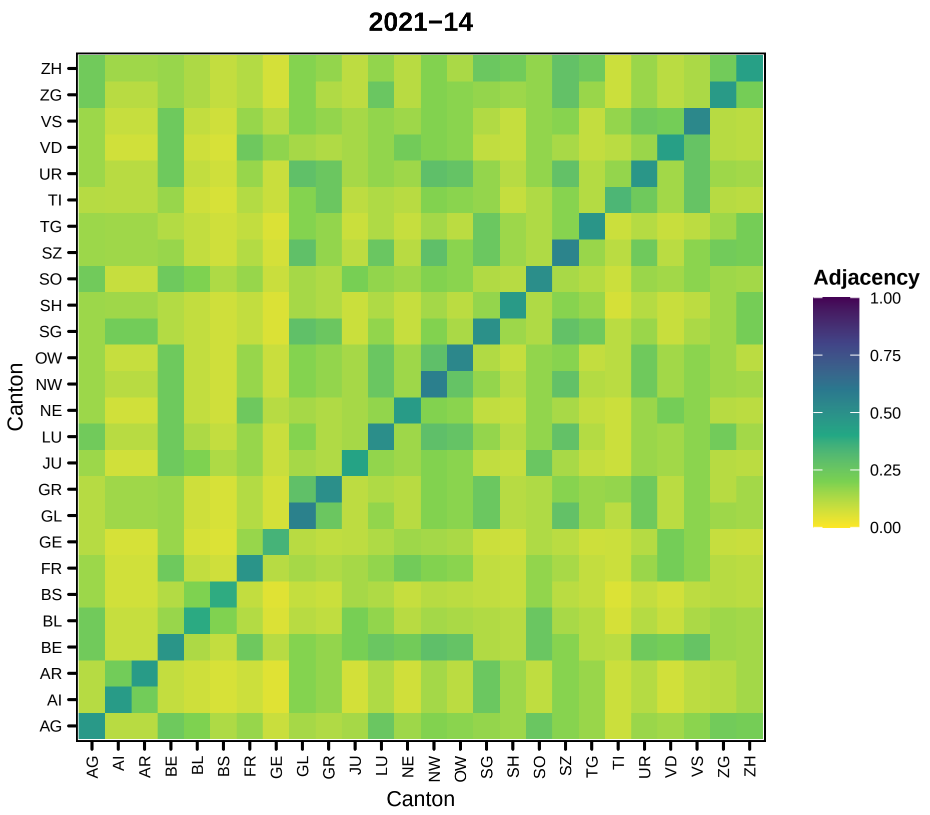
2021−14
Adjacency
1.00
0.75
0.50
0.25
0.00
Canton
ZH
ZG
VS
VD
UR
TI
TG
SZ
SO
SH
SG
OW
NW
NE
LU
JU
GR
GL
GE
FR
BS
BL
BE
AR
AI
AG
AG
AI
AR
BE
BL
BS
FR
GE
GL
GR
JU
LU
NE
NW
OW
SG
SH
SO
SZ
TG
TI
UR
VD
VS
ZG
ZH
Canton

87

```
##

## [[16]]
```

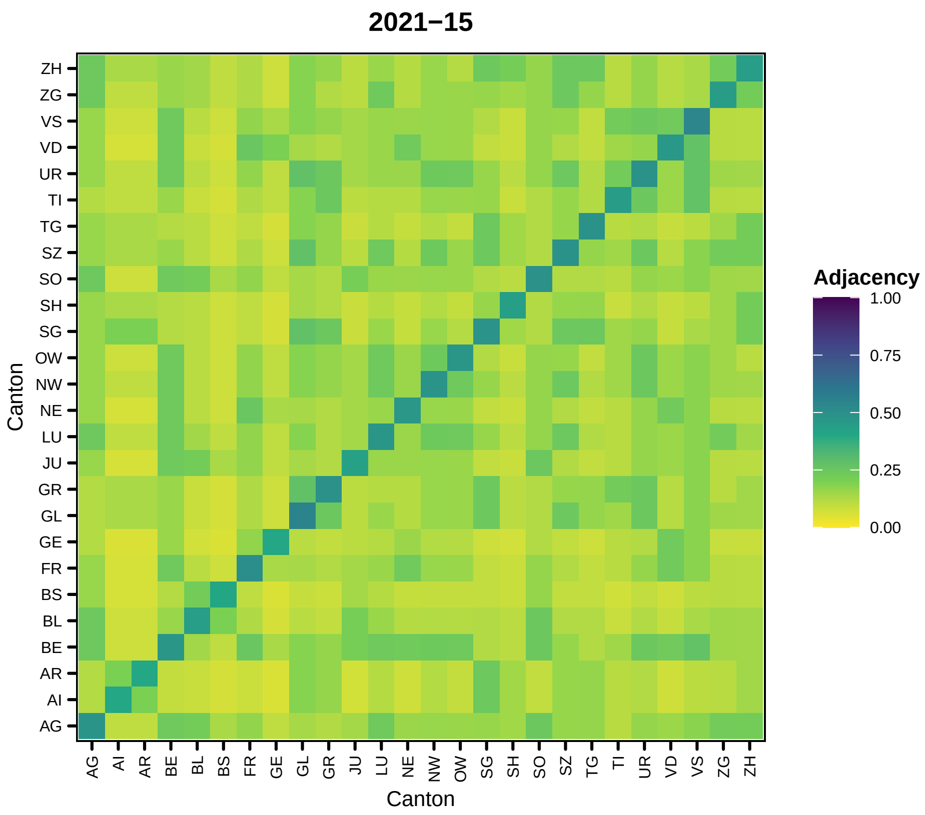
2021−15
Adjacency
1.00
0.75
0.50
0.25
0.00
Canton
ZH
ZG
VS
VD
UR
TI
TG
SZ
SO
SH
SG
OW
NW
NE
LU
JU
GR
GL
GE
FR
BS
BL
BE
AR
AI
AG
Canton
AG AI AR BE BL BS FR GE GL GR JU LU NE NW OW SG SH SO SZ TG TI UR VD VS ZG ZH

88

```
##

## [[17]]
```

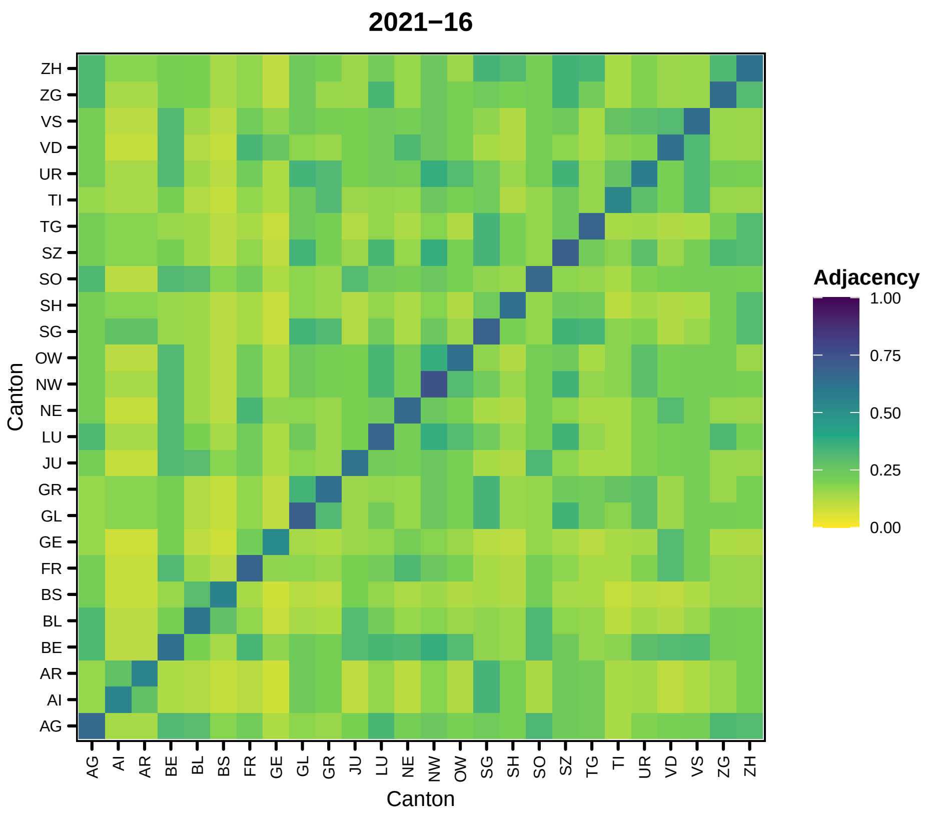
2021−16
Adjacency
1.00
0.75
0.50
0.25
0.00
Canton
ZH
ZG
VS
VD
UR
TI
TG
SZ
SO
SH
SG
OW
NW
NE
LU
JU
GR
GL
GE
FR
BS
BL
BE
AR
AI
AG
Canton

89

```
##
## [[18]]
```

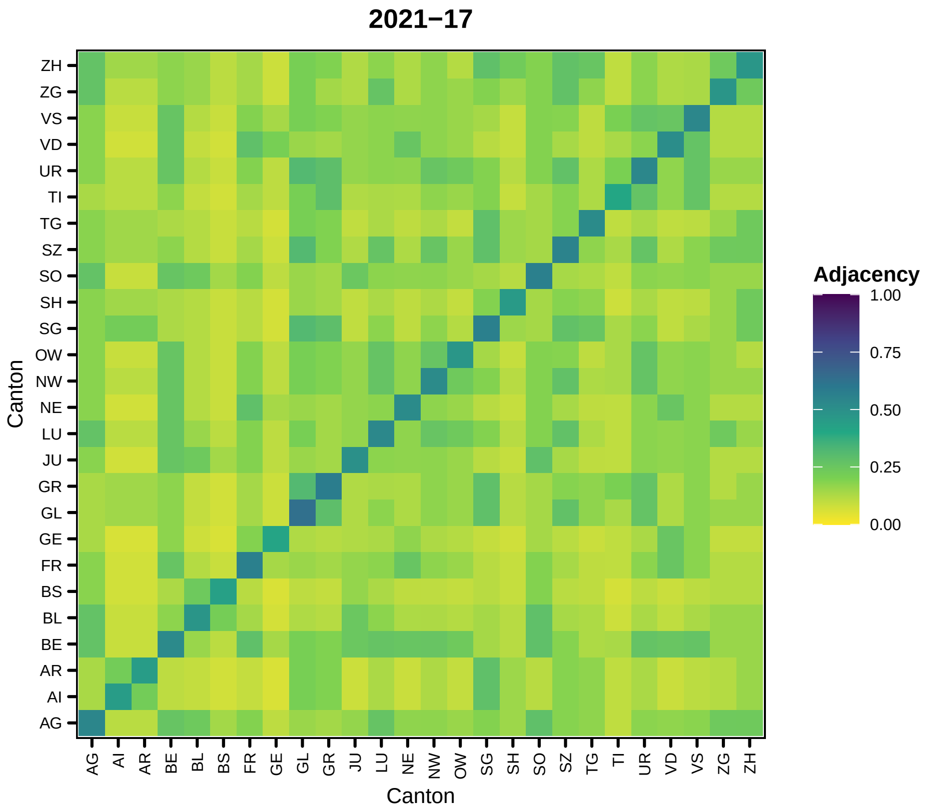
2021−17
Adjacency
1.00
0.75
0.50
0.25
0.00
Canton
Canton
ZH
ZG
VS
VD
UR
TI
TG
SZ
SO
SH
SG
OW
NW
NE
LU
JU
GR
GL
GE
FR
BS
BL
BE
AR
AI
AG

90

```
##

## [[19]]
```

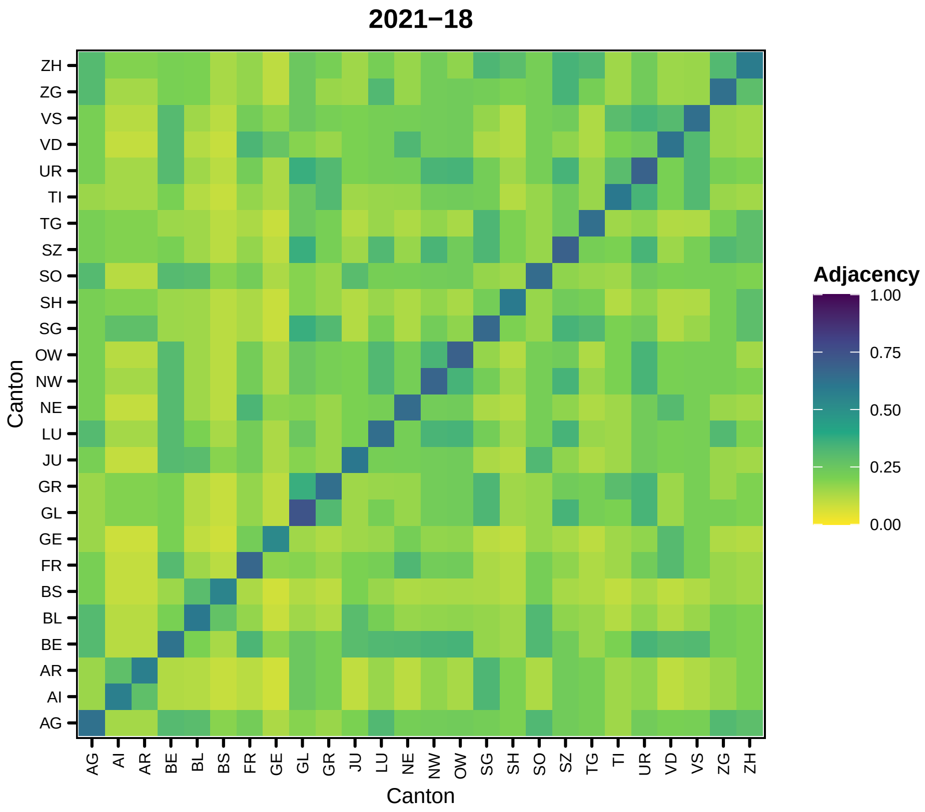
2021−18
Adjacency
1.00
0.75
0.50
0.25
0.00
Canton
Canton
ZH
ZG
VS
VD
UR
TI
TG
SZ
SO
SH
SG
OW
NW
NE
LU
JU
GR
GL
GE
FR
BS
BL
BE
AR
AI
AG

91

```
##
## [[20]]
```

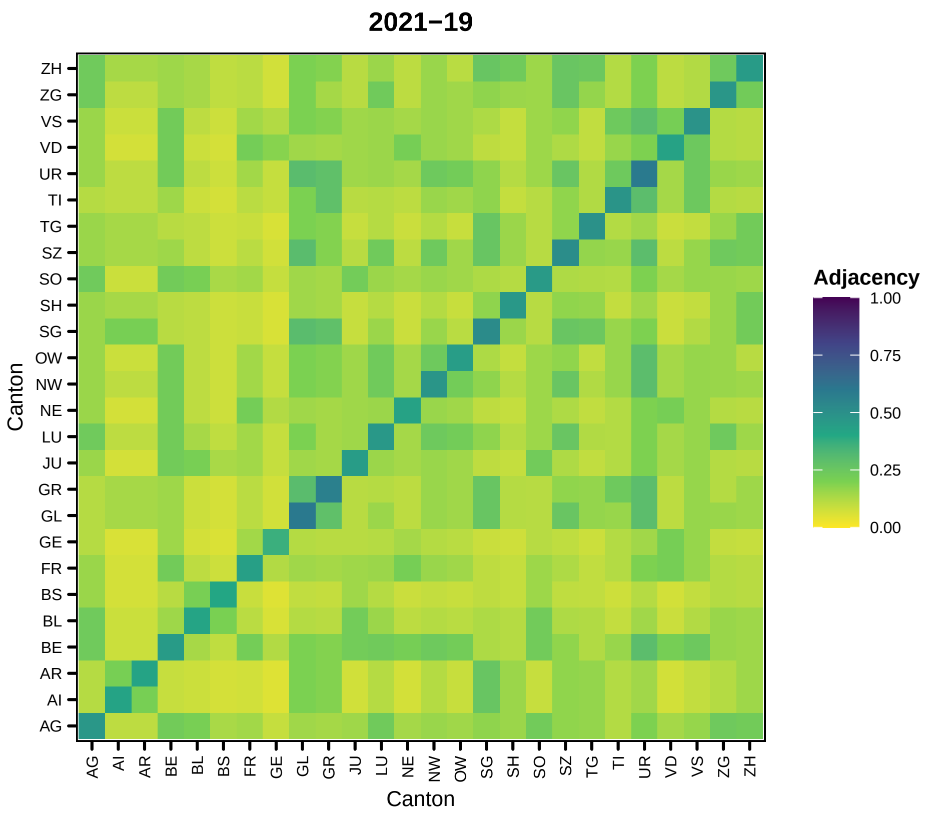

2021−19
Canton
ZH
ZG
VS
VD
UR
TI
TG
SZ
SO
SH
SG
OW
NW
NE
LU
JU
GR
GL
GE
FR
BS
BL
BE
AR
AI
AG
Canton
AG
AI
AR
BE
BL
BS
FR
GE
GL
GR
JU
LU
NE
NW
OW
SG
SH
SO
SZ
TG
TI
UR
VD
VS
ZG
ZH
Adjacency
1.00
0.75
0.50
0.25
0.00


92

```
##
## [[21]]
```

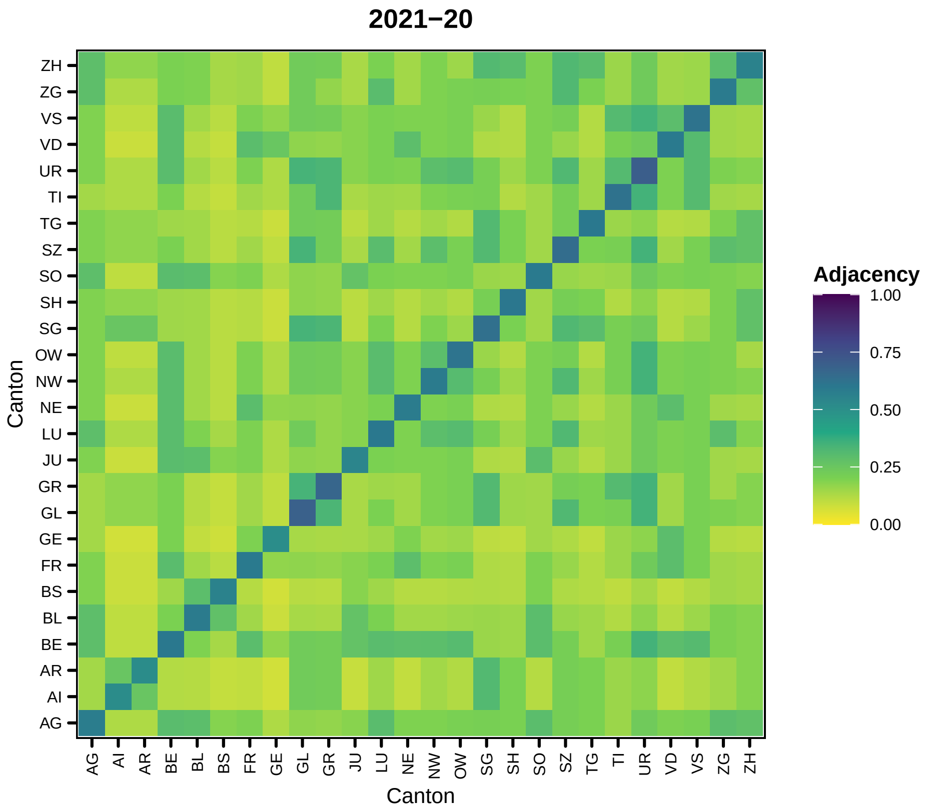
2021−20
Adjacency
1.00
0.75
0.50
0.25
0.00
Canton
Canton
ZH
ZG
VS
VD
UR
TI
TG
SZ
SO
SH
SG
OW
NW
NE
LU
JU
GR
GL
GE
FR
BS
BL
BE
AR
AI
AG

93

```
##
## [[22]]
```

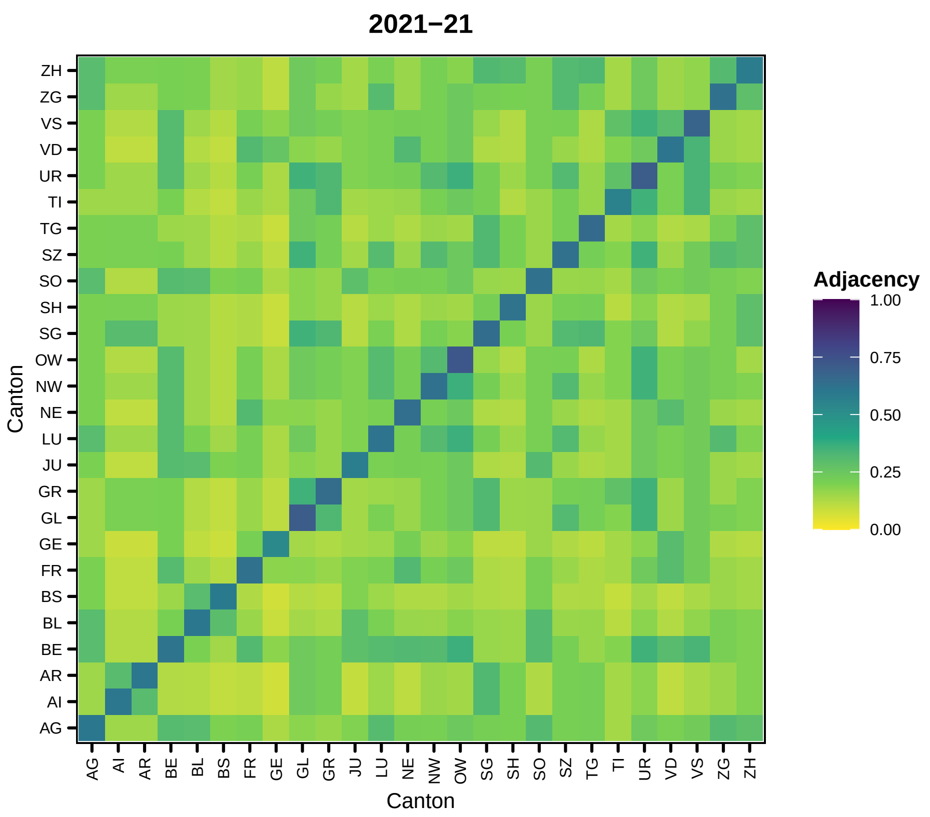

2021−21
Canton
ZH
ZG
VS
VD
UR
TI
TG
SZ
SO
SH
SG
OW
NW
NE
LU
JU
GR
GL
GE
FR
BS
BL
BE
AR
AI
AG
AG AI AR BE BL BS FR GE GL GR JU LU NE NW OW SG SH SO SZ TG TI UR VD VS ZG ZH
Canton
Adjacency
1.00
0.75
0.50
0.25
0.00


94

```
##
## [[23]]
```

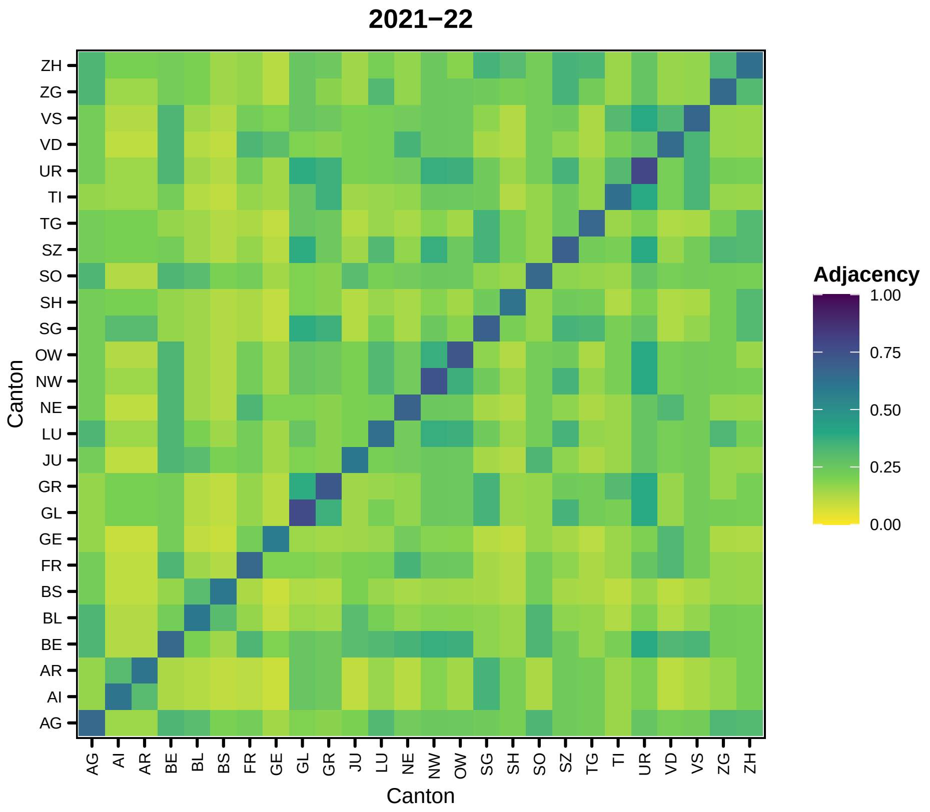

2021−22
Canton
ZH
ZG
VS
VD
UR
TI
TG
SZ
SO
SH
SG
OW
NW
NE
LU
JU
GR
GL
GE
FR
BS
BL
BE
AR
AI
AG
AG
AI
AR
BE
BL
BS
FR
GE
GL
GR
JU
LU
NE
NW
OW
SG
SH
SO
SZ
TG
TI
UR
VD
VS
ZG
ZH
Adjacency
1.00
0.75
0.50
0.25
0.00


95

```
##
## [[24]]
```

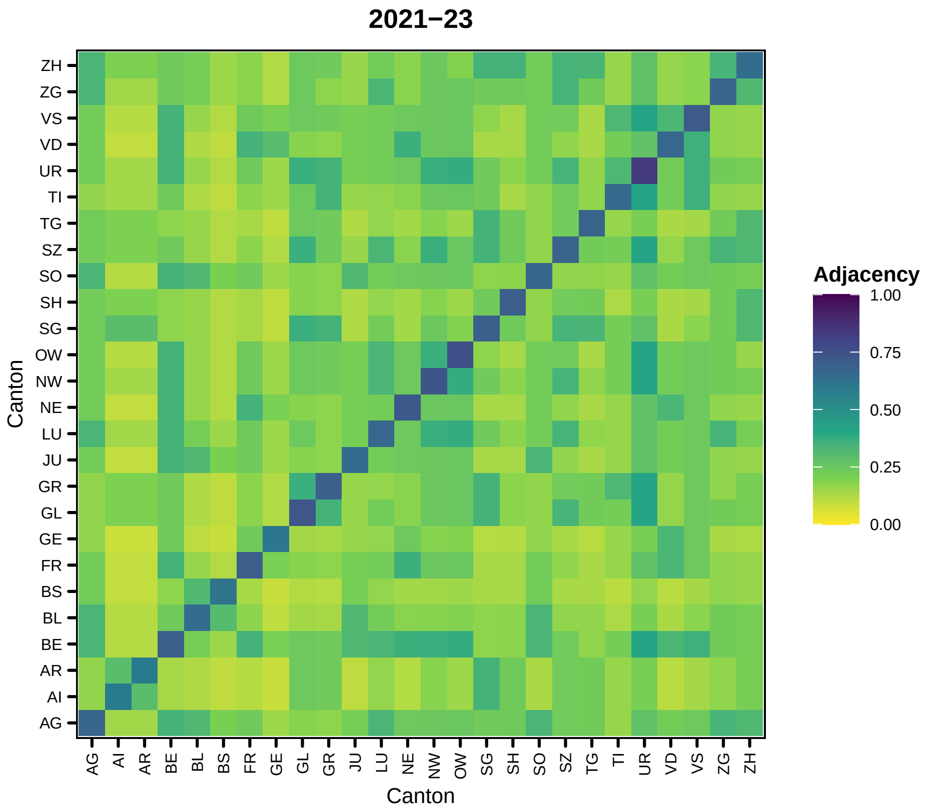
2021−23
Adjacency
1.00
0.75
0.50
0.25
0.00
Canton
ZH
ZG
VS
VD
UR
TI
TG
SZ
SO
SH
SG
OW
NW
NE
LU
JU
GR
GL
GE
FR
BS
BL
BE
AR
AI
AG
Canton
AG AI AR BE BL BS FR GE GL GR JU LU NE NW OW SG SH SO SZ TG TI UR VD VS ZG ZH

96

```
##

## [[25]]
```

97

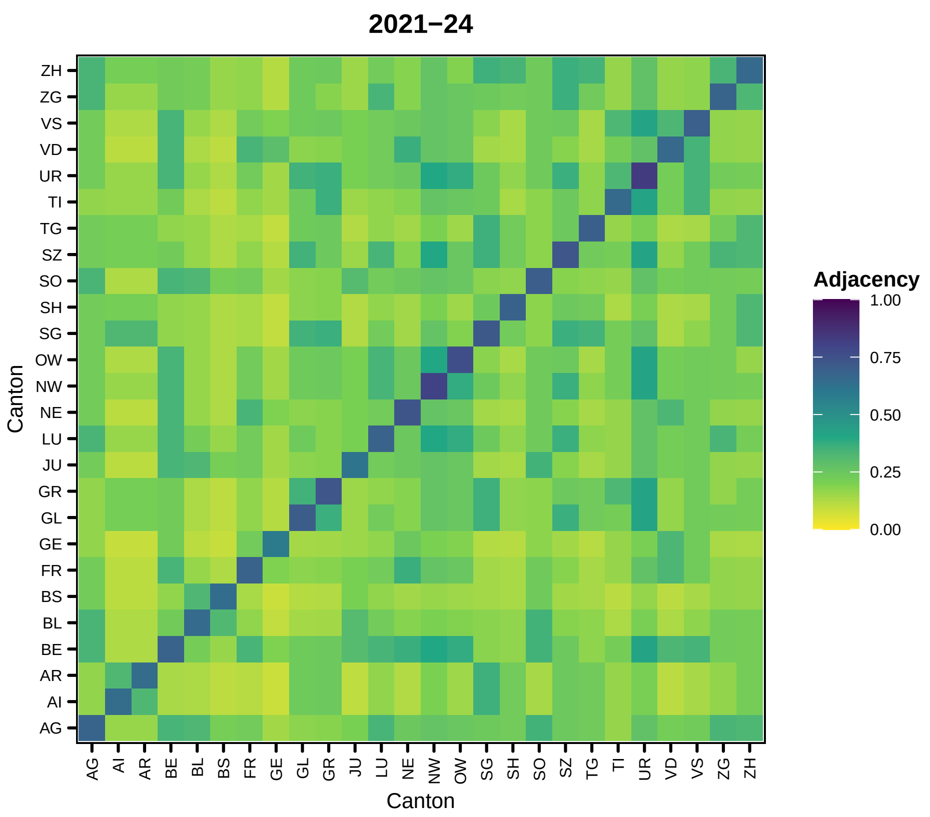

2021−24
Adjacency
1.00
0.75
0.50
0.25
0.00
Canton
Canton
ZH
ZG
VS
VD
UR
TI
TG
SZ
SO
SH
SG
OW
NW
NE
LU
JU
GR
GL
GE
FR
BS
BL
BE
AR
AI
AG


```
##
## [[26]]
```

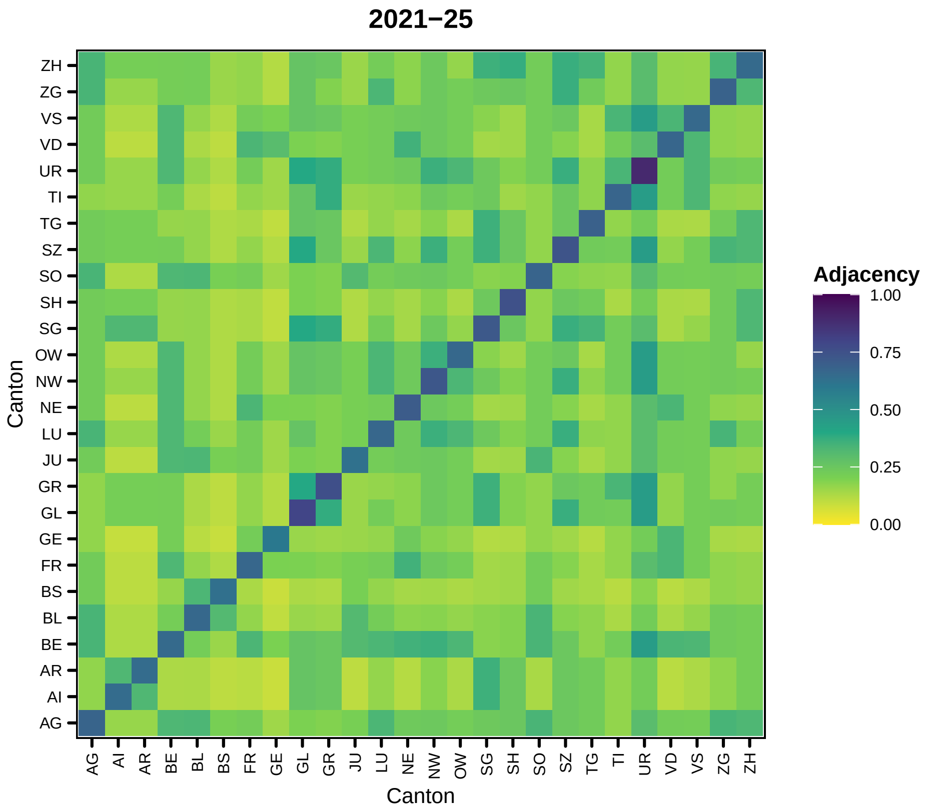
2021−25
ZH
ZG
VS
VD
UR
TI
TG
SZ
SO
SH
SG
OW
NW
NE
LU
JU
GR
GL
GE
FR
BS
BL
BE
AR
AI
AG
Canton
AG AI AR BE BL BS FR GE GL GR JU LU NE NW OW SG SH SO SZ TG TI UR VD VS ZG ZH
Canton
Adjacency
1.00
0.75
0.50
0.25
0.00

98

```
##
## [[27]]
```

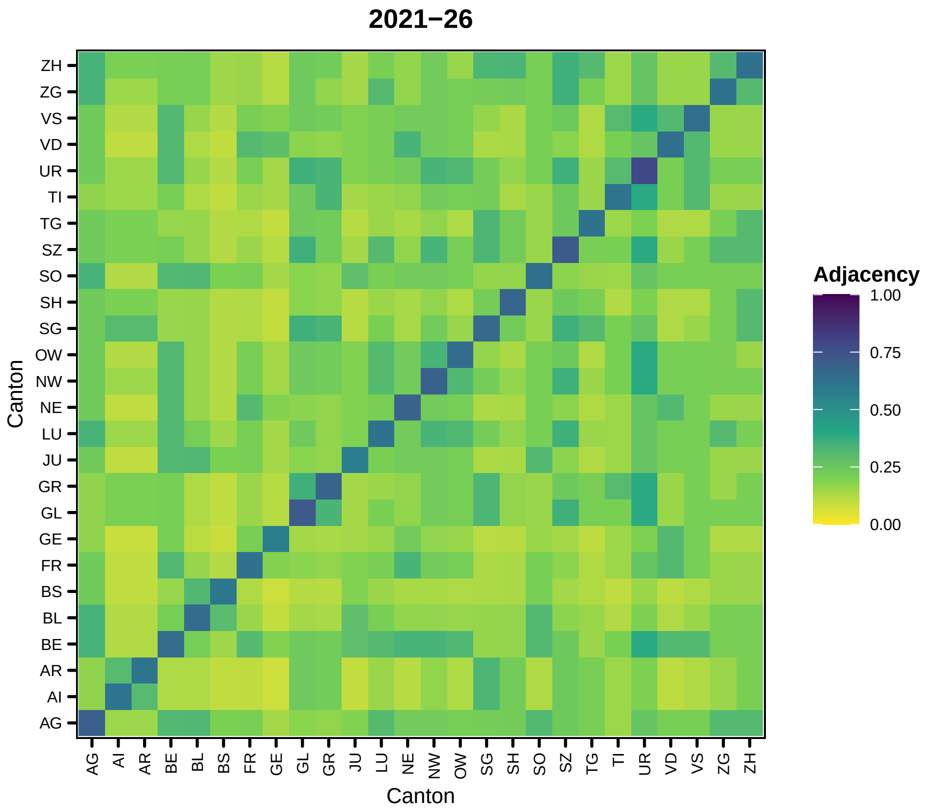
2021−26
Adjacency
1.00
0.75
0.50
0.25
0.00
Canton
Canton
ZH
ZG
VS
VD
UR
TI
TG
SZ
SO
SH
SG
OW
NW
NE
LU
JU
GR
GL
GE
FR
BS
BL
BE
AR
AI
AG

99

```
##

## [[28]]
```

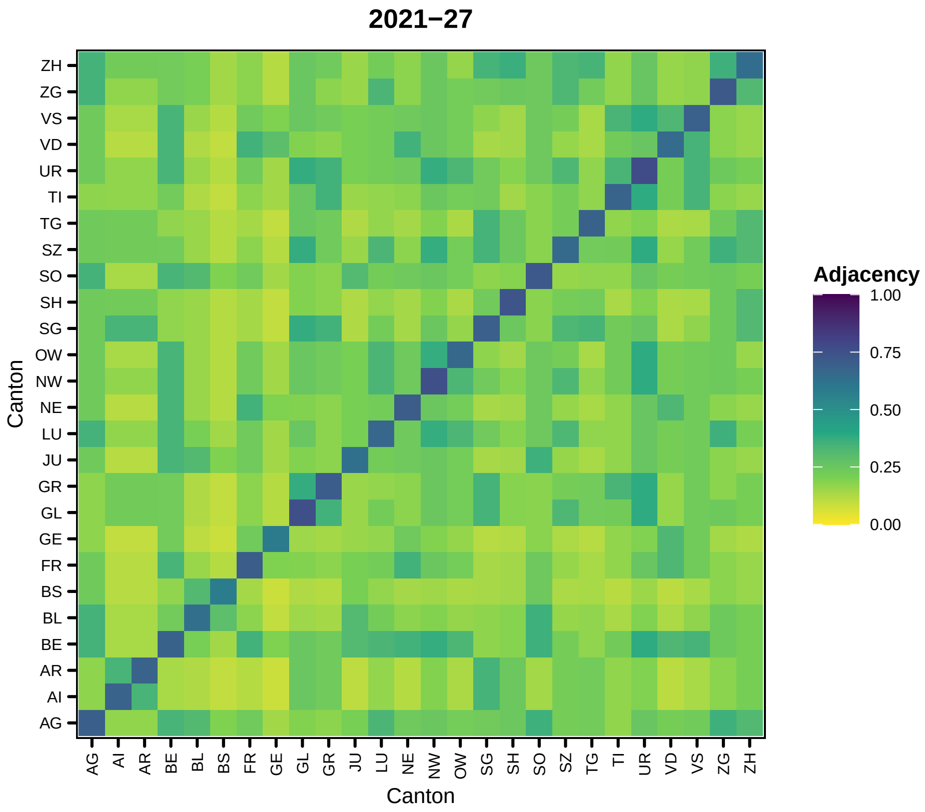
2021−27
Adjacency
1.00
0.75
0.50
0.25
0.00
Canton
Canton
ZH
ZG
VS
VD
UR
TI
TG
SZ
SO
SH
SG
OW
NW
NE
LU
JU
GR
GL
GE
FR
BS
BL
BE
AR
AI
AG

100

```
##

## [[29]]
```

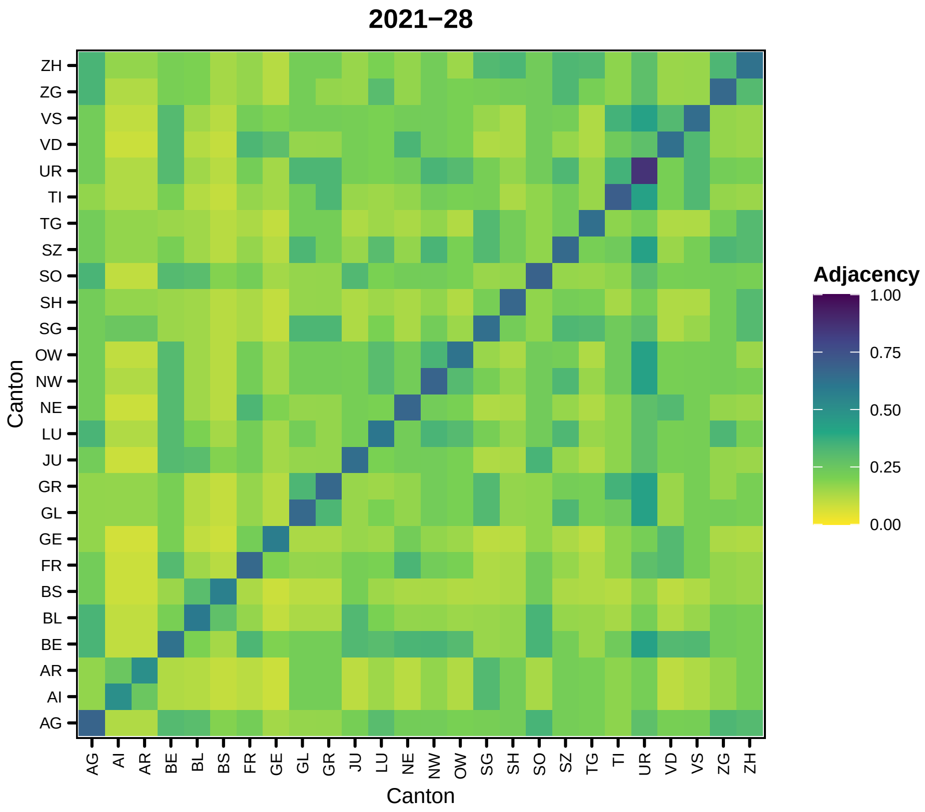
2021−28
Canton
ZH ZG VS VD UR TI TG SZ SO SH SG OW NW NE LU JU GR GL GE FR BS BL BE AR AI AG
AG AI AR BE BL BS FR GE GL GR JU LU NE NW OW SG SH SO SZ TG TI UR VD VS ZG ZH
Canton
Adjacency
1.00
0.75
0.50
0.25
0.00

101

```
##
## [[30]]
```

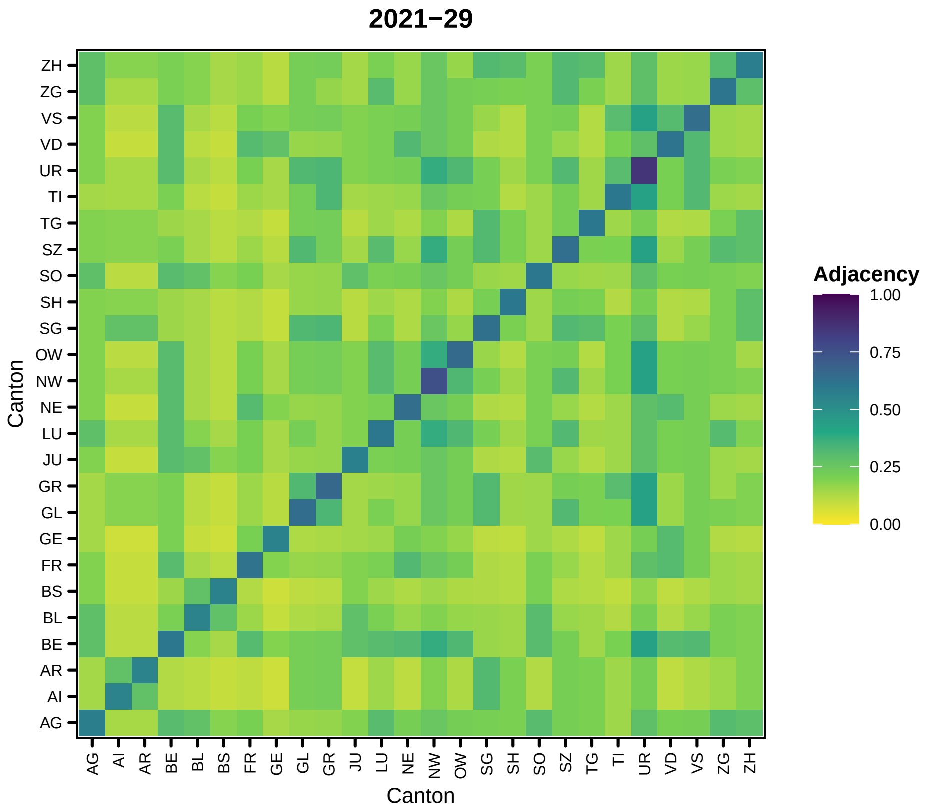
2021−29
Canton
ZH
ZG
VS
VD
UR
TI
TG
SZ
SO
SH
SG
OW
NW
NE
LU
JU
GR
GL
GE
FR
BS
BL
BE
AR
AI
AG
Adjacency
1.00
0.75
0.50
0.25
0.00

102

```
##

## [[31]]
```

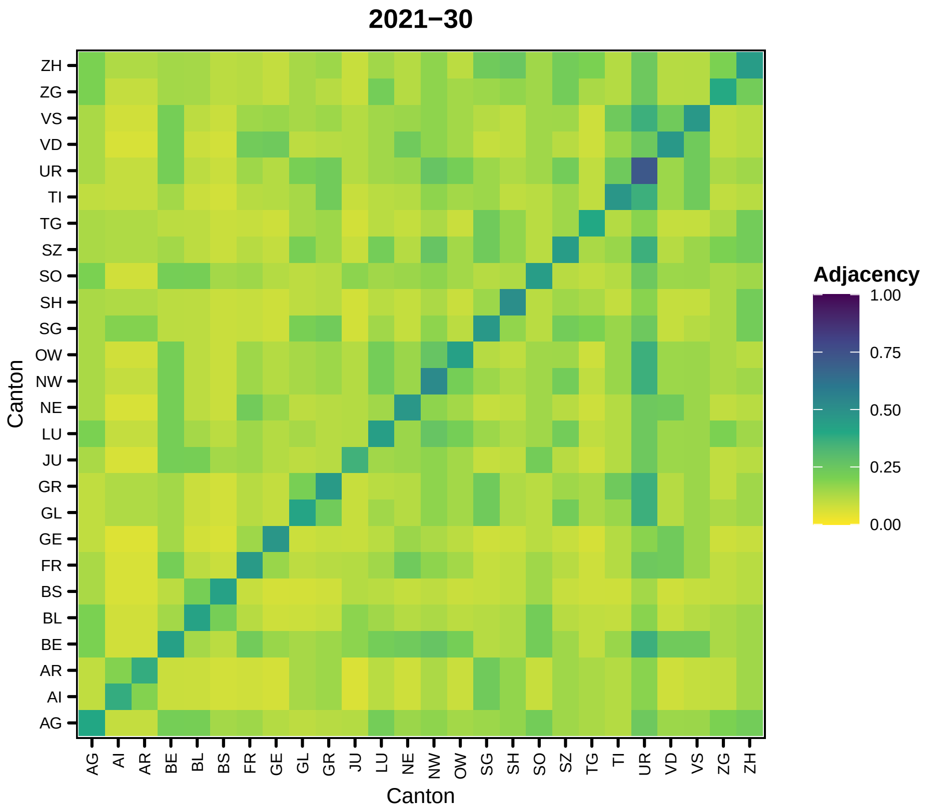
2021−30
Adjacency
1.00
0.75
0.50
0.25
0.00
Canton
Canton
ZH
ZG
VS
VD
UR
TI
TG
SZ
SO
SH
SG
OW
NW
NE
LU
JU
GR
GL
GE
FR
BS
BL
BE
AR
AI
AG

103

```
##
## [[32]]
```

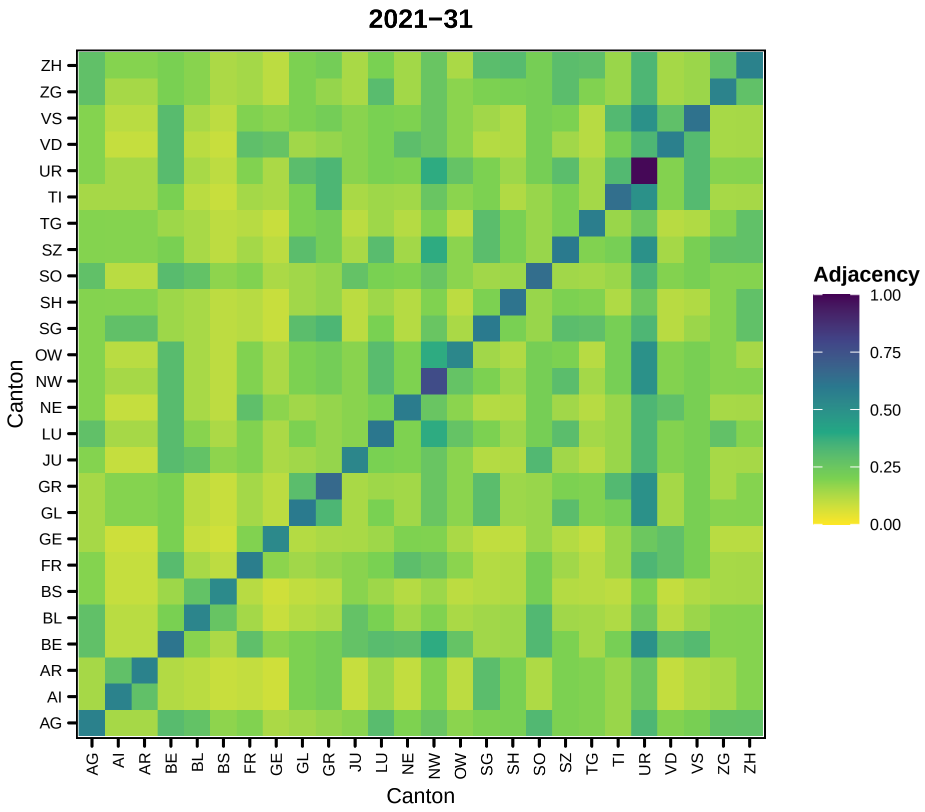
2021−31
Canton
ZH ZG VS VD UR TI TG SZ SO SH SG OW NW NE LU JU GR GL GE FR BS BL BE AR AI AG
AG AI AR BE BL BS FR GE GL GR JU LU NE NW OW SG SH SO SZ TG TI UR VD VS ZG ZH
Canton
Adjacency
1.00
0.75
0.50
0.25
0.00

104

```
##

## [[33]]
```

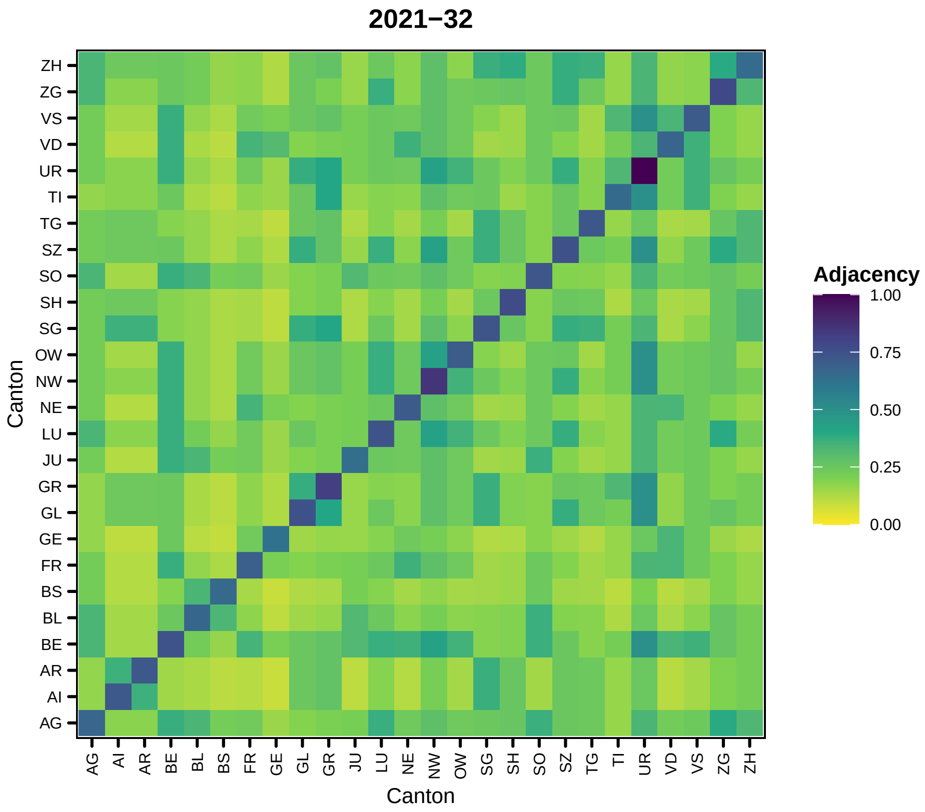
2021−32
Adjacency
1.00
0.75
0.50
0.25
0.00
Canton
Canton
ZH
ZG
VS
VD
UR
TI
TG
SZ
SO
SH
SG
OW
NW
NE
LU
JU
GR
GL
GE
FR
BS
BL
BE
AR
AI
AG

105

```
##
## [[34]]
```

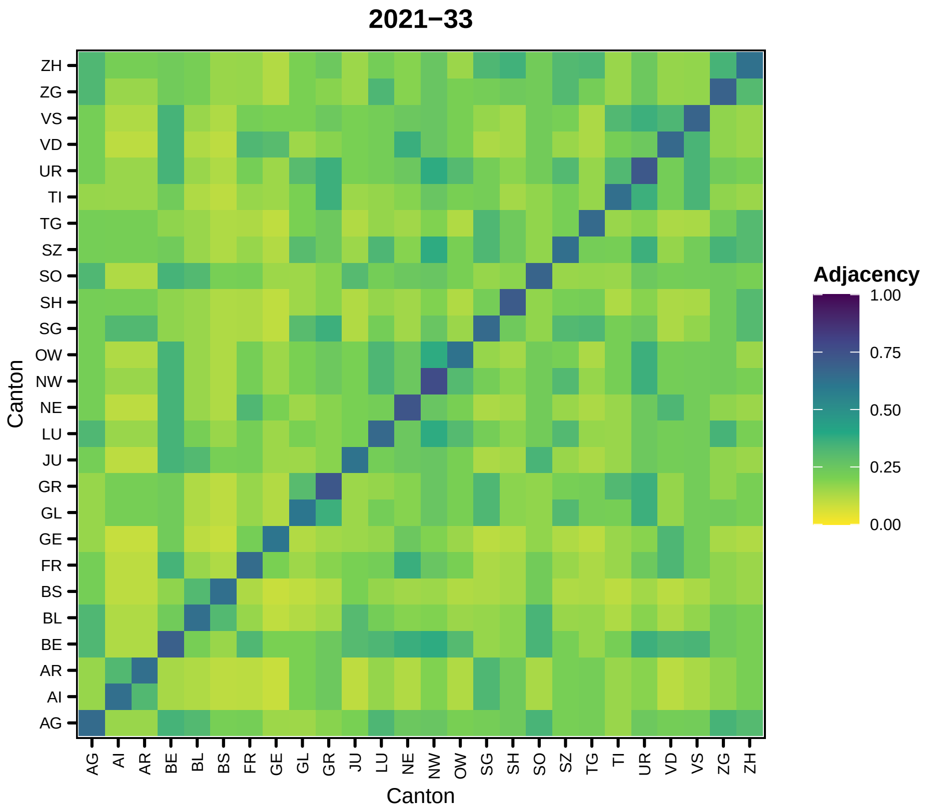
2021−33
Adjacency
1.00
0.75
0.50
0.25
0.00
Canton
Canton
AG AI AR BE BL BS FR GE GL GR JU LU NE NW OW SG SH SO SZ TG TI UR VD VS ZG ZH

106

```
##
## [[35]]
```

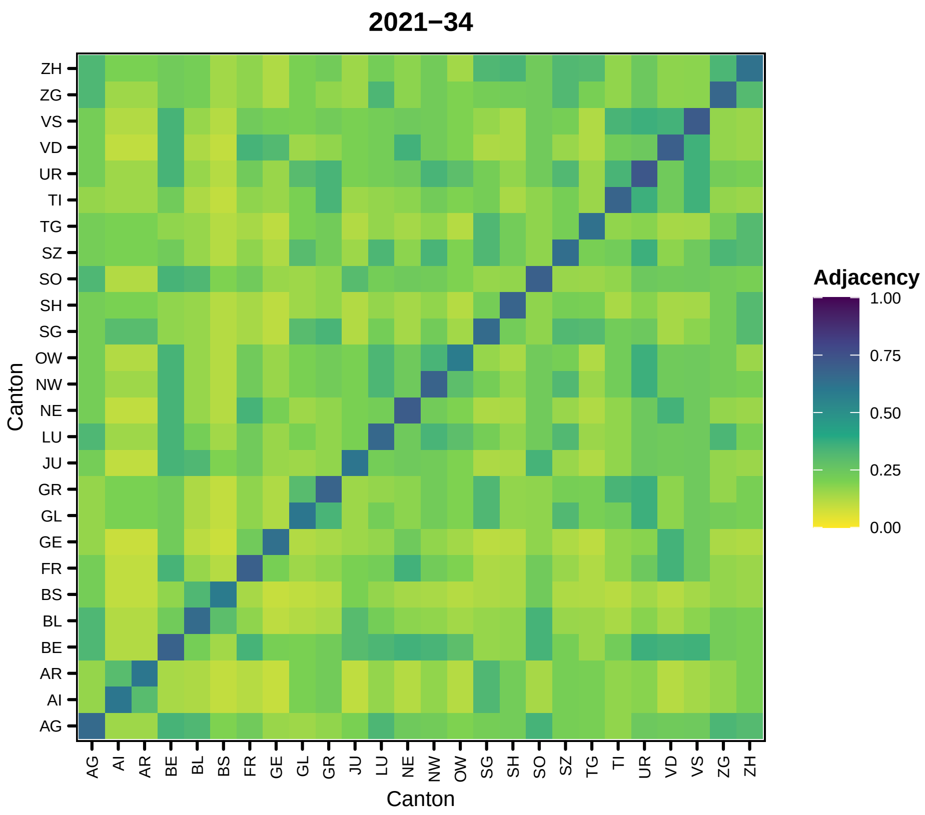
2021−34
Adjacency
1.00
0.75
0.50
0.25
0.00
Canton
ZH
ZG
VS
VD
UR
TI
TG
SZ
SO
SH
SG
OW
NW
NE
LU
JU
GR
GL
GE
FR
BS
BL
BE
AR
AI
AG
Canton
AG
AI
AR
BE
BL
BS
FR
GE
GL
GR
JU
LU
NE
NW
OW
SG
SH
SO
SZ
TG
TI
UR
VD
VS
ZG
ZH

107

```
##
## [[36]]
```

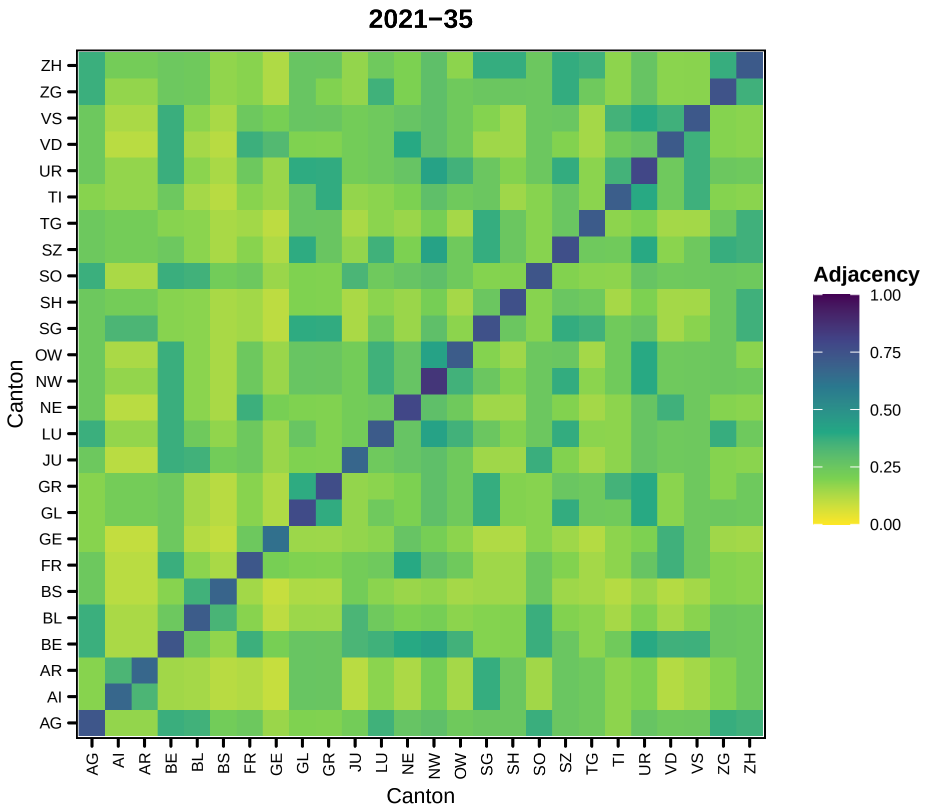
2021−35
Canton
ZH
ZG
VS
VD
UR
TI
TG
SZ
SO
SH
SG
OW
NW
NE
LU
JU
GR
GL
GE
FR
BS
BL
BE
AR
AI
AG
Adjacency
1.00
0.75
0.50
0.25
0.00

108

```
##
## [[37]]
```

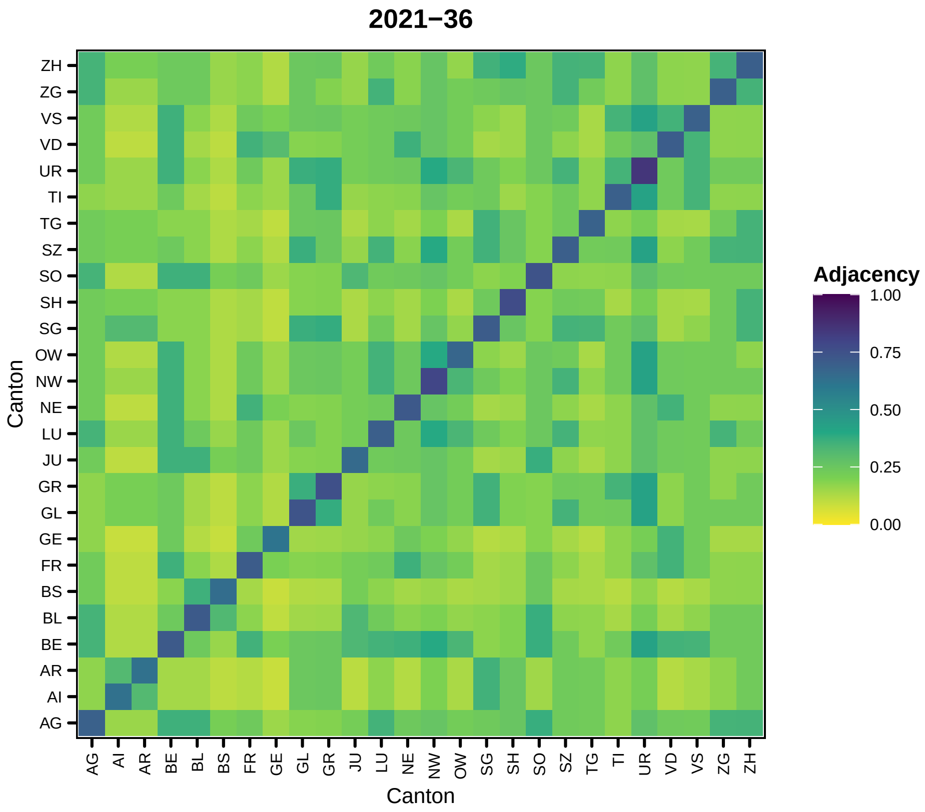
2021−36
Adjacency
1.00
0.75
0.50
0.25
0.00
Canton
ZH
ZG
VS
VD
UR
TI
TG
SZ
SO
SH
SG
OW
NW
NE
LU
JU
GR
GL
GE
FR
BS
BL
BE
AR
AI
AG
Canton
AG
AI
AR
BE
BL
BS
FR
GE
GL
GR
JU
LU
NE
NW
OW
SG
SH
SO
SZ
TG
TI
UR
VD
VS
ZG
ZH

109

```
##
## [[38]]
```

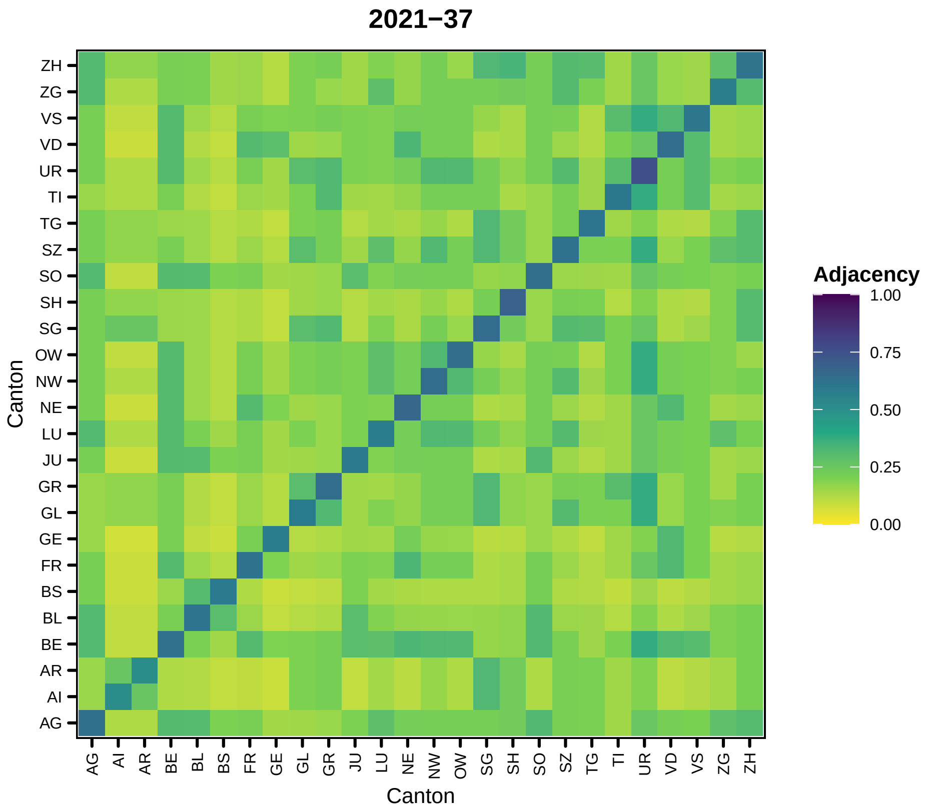

2021−37
Adjacency
1.00
0.75
0.50
0.25
0.00
Canton
Canton
ZH
ZG
VS
VD
UR
TI
TG
SZ
SO
SH
SG
OW
NW
NE
LU
JU
GR
GL
GE
FR
BS
BL
BE
AR
AI
AG


110

```
##
## [[39]]
```

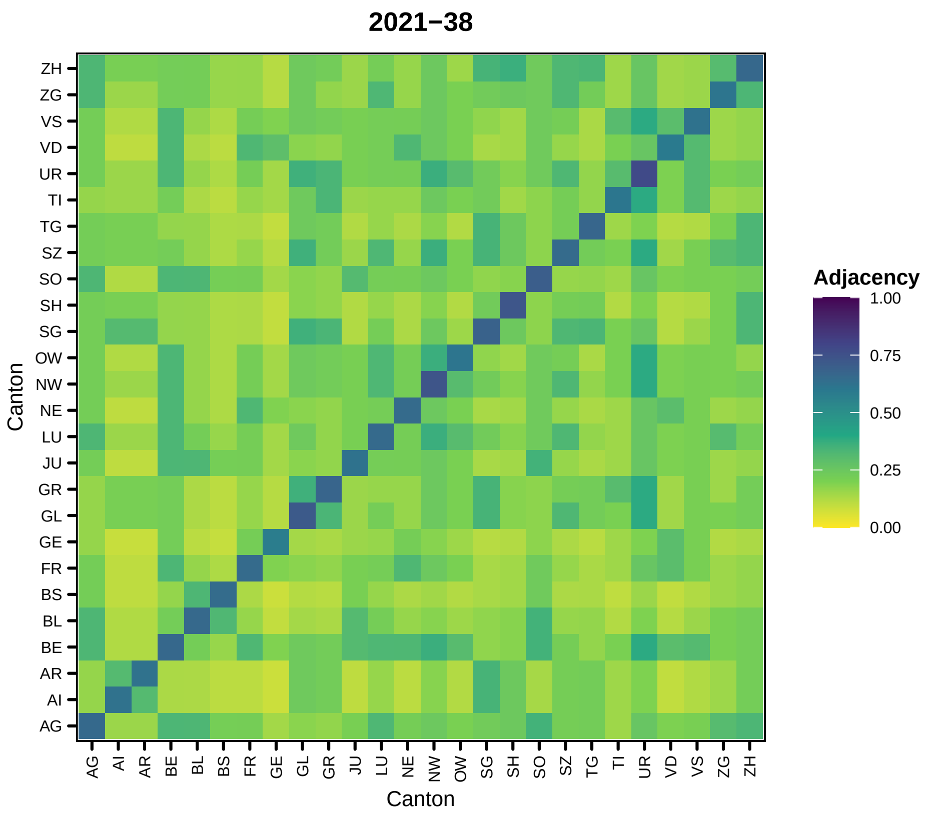
2021−38
Canton
ZH ZG VS VD UR TI TG SZ SO SH SG OW NW NE LU JU GR GL GE FR BS BL BE AR AI AG
AG AI AR BE BL BS FR GE GL GR JU LU NE NW OW SG SH SO SZ TG TI UR VD VS ZG ZH
Canton
Adjacency
1.00
0.75
0.50
0.25
0.00

111

```
##
## [[40]]
```

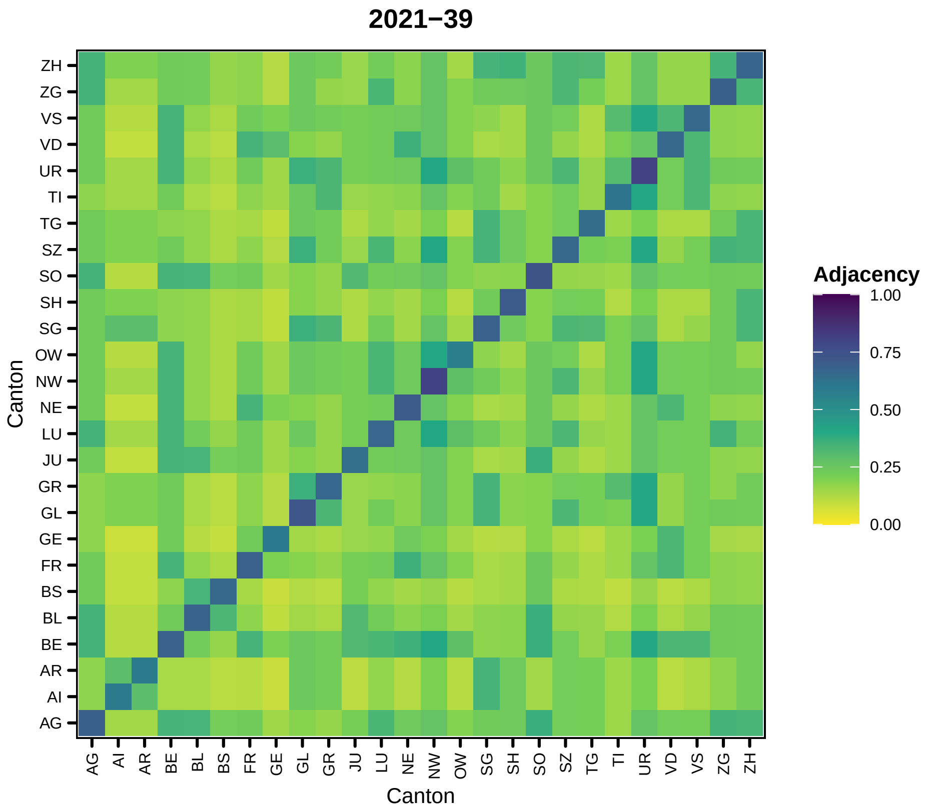
2021−39
Adjacency
1.00
0.75
0.50
0.25
0.00
Canton
Canton
ZH
ZG
VS
VD
UR
TI
TG
SZ
SO
SH
SG
OW
NW
NE
LU
JU
GR
GL
GE
FR
BS
BL
BE
AR
AI
AG

112

```
##
## [[41]]
```

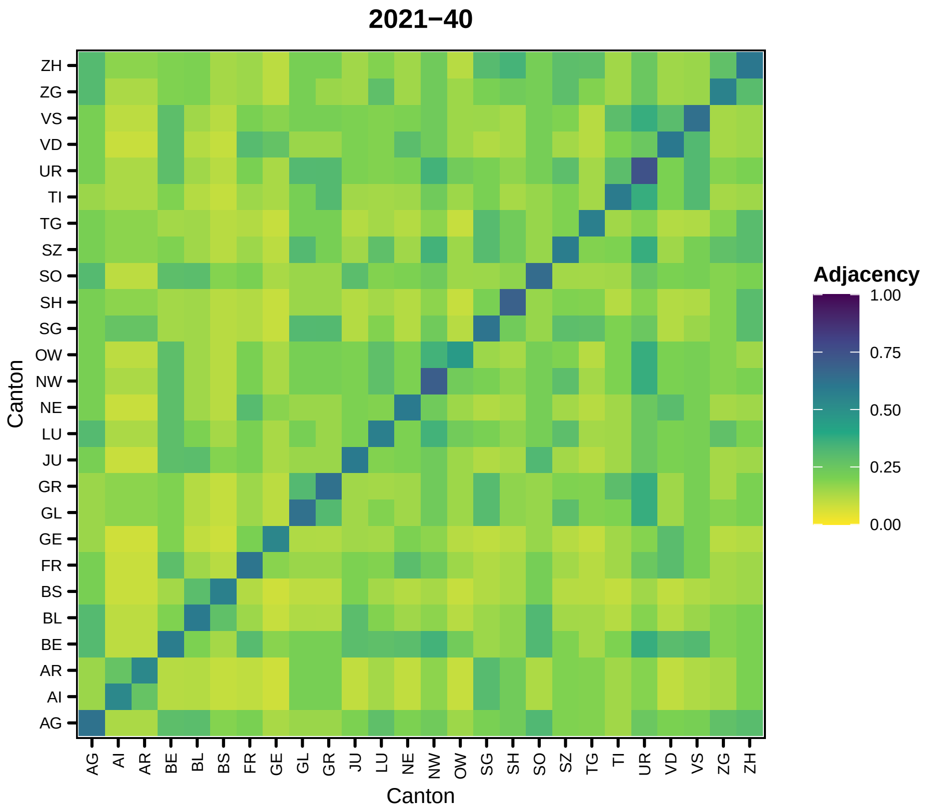
2021−40
Adjacency
1.00
0.75
0.50
0.25
0.00
Canton
Canton
ZH
ZG
VS
VD
UR
TI
TG
SZ
SO
SH
SG
OW
NW
NE
LU
JU
GR
GL
GE
FR
BS
BL
BE
AR
AI
AG

113

```
##
## [[42]]
```

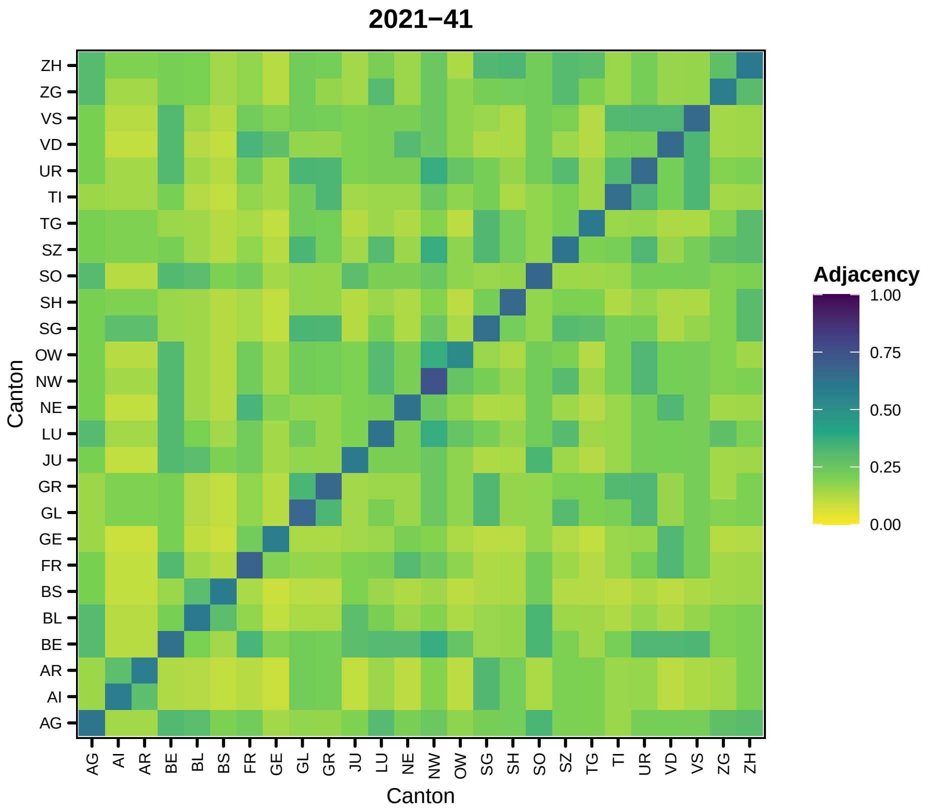
2021−41
Canton
ZH
ZG
VS
VD
UR
TI
TG
SZ
SO
SH
SG
OW
NW
NE
LU
JU
GR
GL
GE
FR
BS
BL
BE
AR
AI
AG
Adjacency
1.00
0.75
0.50
0.25
0.00

114

```
##
## [[43]]
```

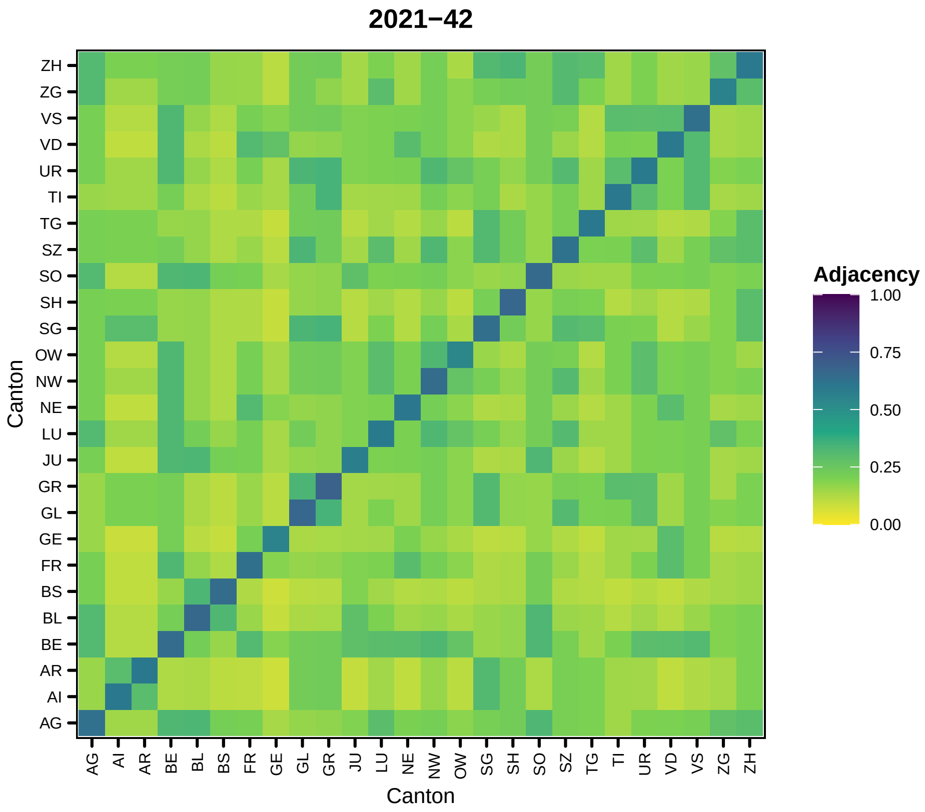
2021−42
Adjacency
1.00
0.75
0.50
0.25
0.00
Canton
ZH
ZG
VS
VD
UR
TI
TG
SZ
SO
SH
SG
OW
NW
NE
LU
JU
GR
GL
GE
FR
BS
BL
BE
AR
AI
AG
Canton
AG
AI
AR
BE
BL
BS
FR
GE
GL
GR
JU
LU
NE
NW
OW
SG
SH
SO
SZ
TG
TI
UR
VD
VS
ZG
ZH

115

```
##
## [[44]]
```

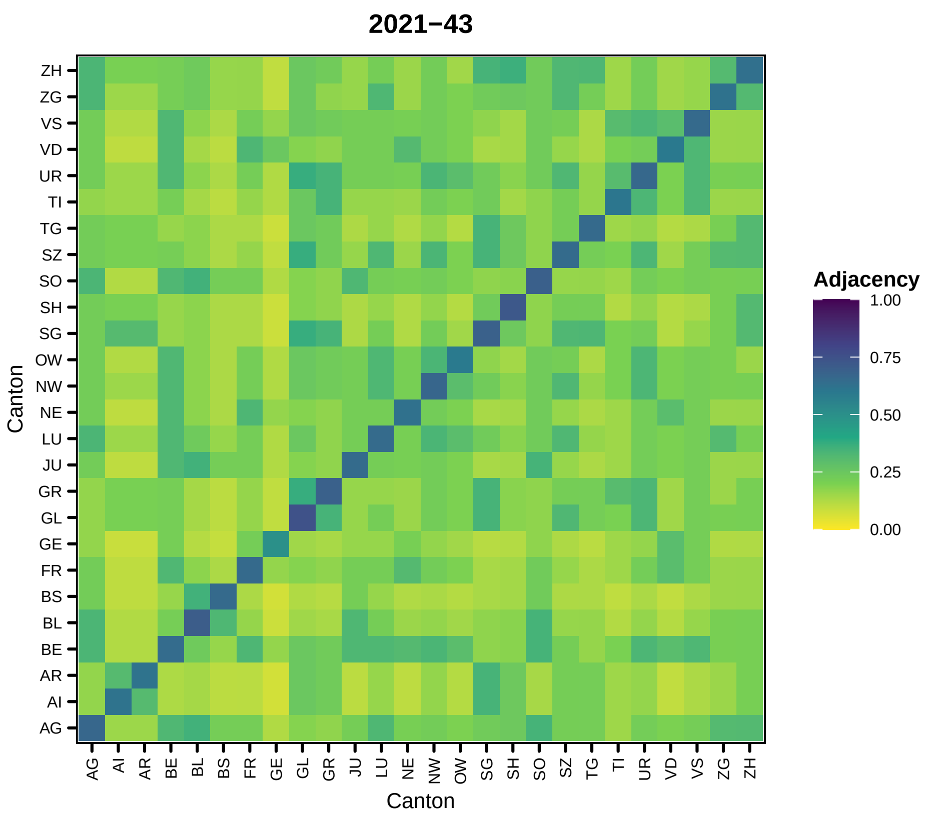
2021−43
Adjacency
1.00
0.75
0.50
0.25
0.00
Canton
ZH
ZG
VS
VD
UR
TI
TG
SZ
SO
SH
SG
OW
NW
NE
LU
JU
GR
GL
GE
FR
BS
BL
BE
AR
AI
AG
Canton
AG
AI
AR
BE
BL
BS
FR
GE
GL
GR
JU
LU
NE
NW
OW
SG
SH
SO
SZ
TG
TI
UR
VD
VS
ZG
ZH

116

```
##

## [[45]]
```

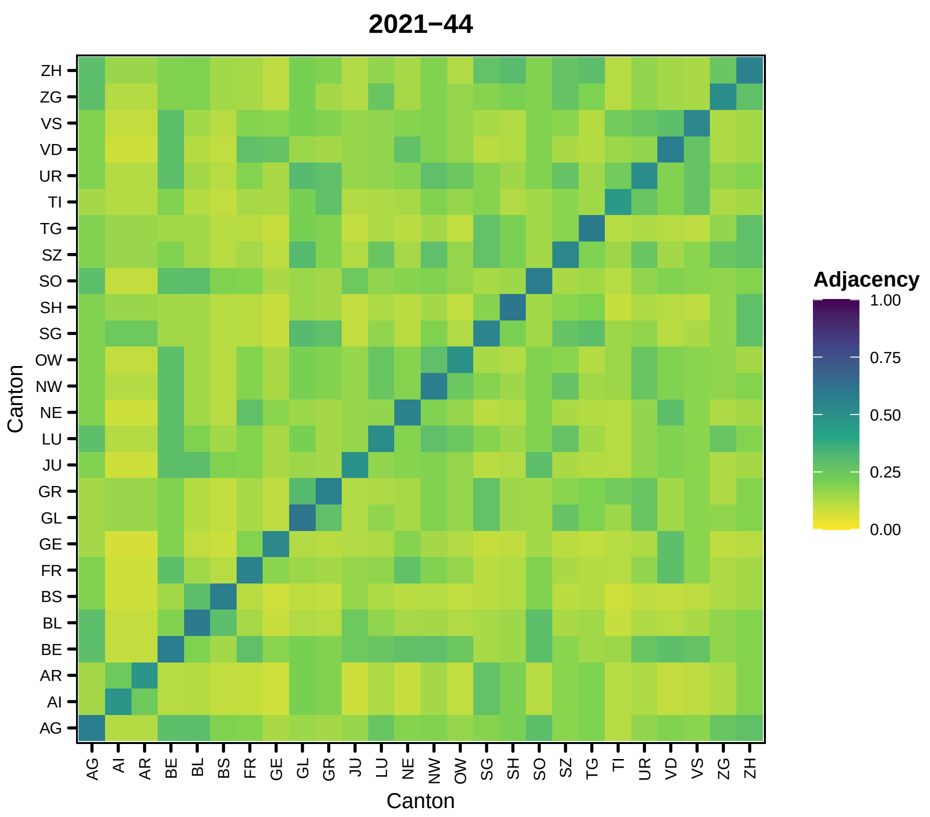

2021−44
Adjacency
1.00
0.75
0.50
0.25
0.00
Canton
ZH
ZG
VS
VD
UR
TI
TG
SZ
SO
SH
SG
OW
NW
NE
LU
JU
GR
GL
GE
FR
BS
BL
BE
AR
AI
AG
Canton
AG
AI
AR
BE
BL
BS
FR
GE
GL
GR
JU
LU
NE
NW
OW
SG
SH
SO
SZ
TG
TI
UR
VD
VS
ZG
ZH


117

```
##
## [[46]]
```

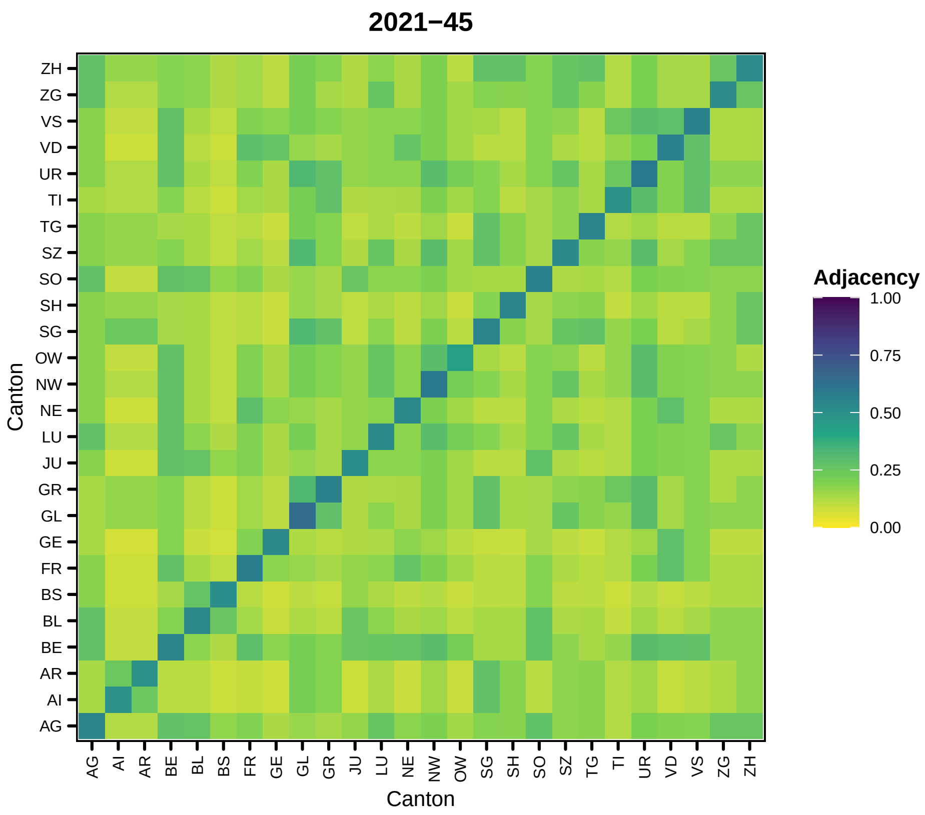
2021−45
Canton
ZH
ZG
VS
VD
UR
TI
TG
SZ
SO
SH
SG
OW
NW
NE
LU
JU
GR
GL
GE
FR
BS
BL
BE
AR
AI
AG
Adjacency
1.00
0.75
0.50
0.25
0.00

118

```
##
## [[47]]
```

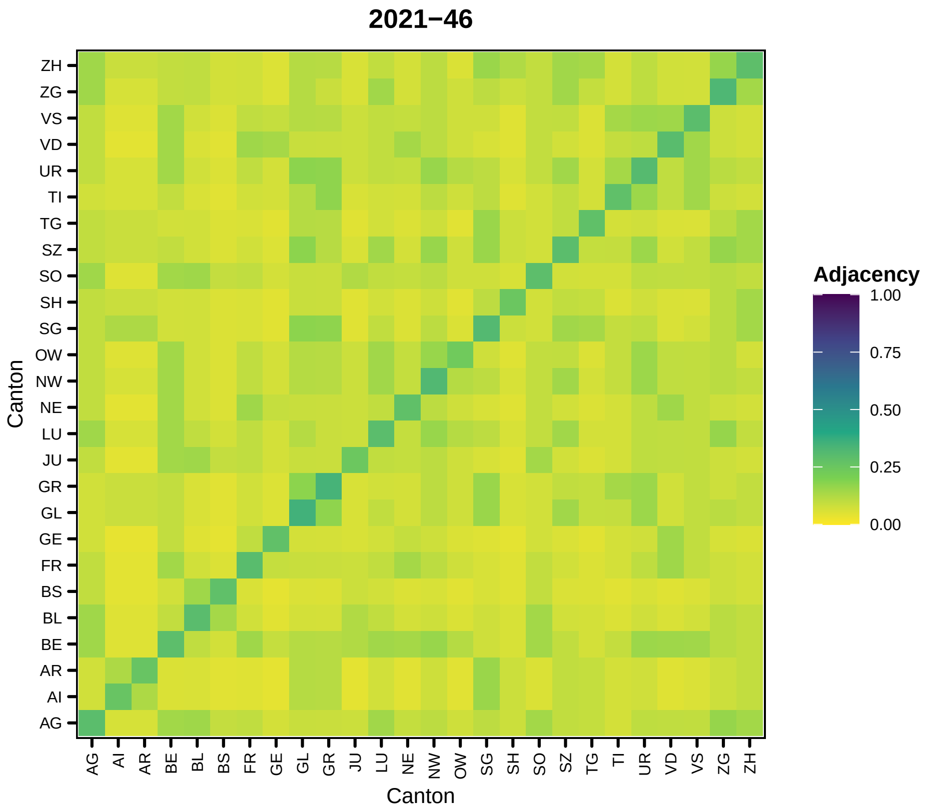
2021−46
Adjacency
1.00
0.75
0.50
0.25
0.00
Canton
ZH
ZG
VS
VD
UR
TI
TG
SZ
SO
SH
SG
OW
NW
NE
LU
JU
GR
GL
GE
FR
BS
BL
BE
AR
AI
AG
Canton
AG
AI
AR
BE
BL
BS
FR
GE
GL
GR
JU
LU
NE
NW
OW
SG
SH
SO
SZ
TG
TI
UR
VD
VS
ZG
ZH

119

```
##
## [[48]]
```

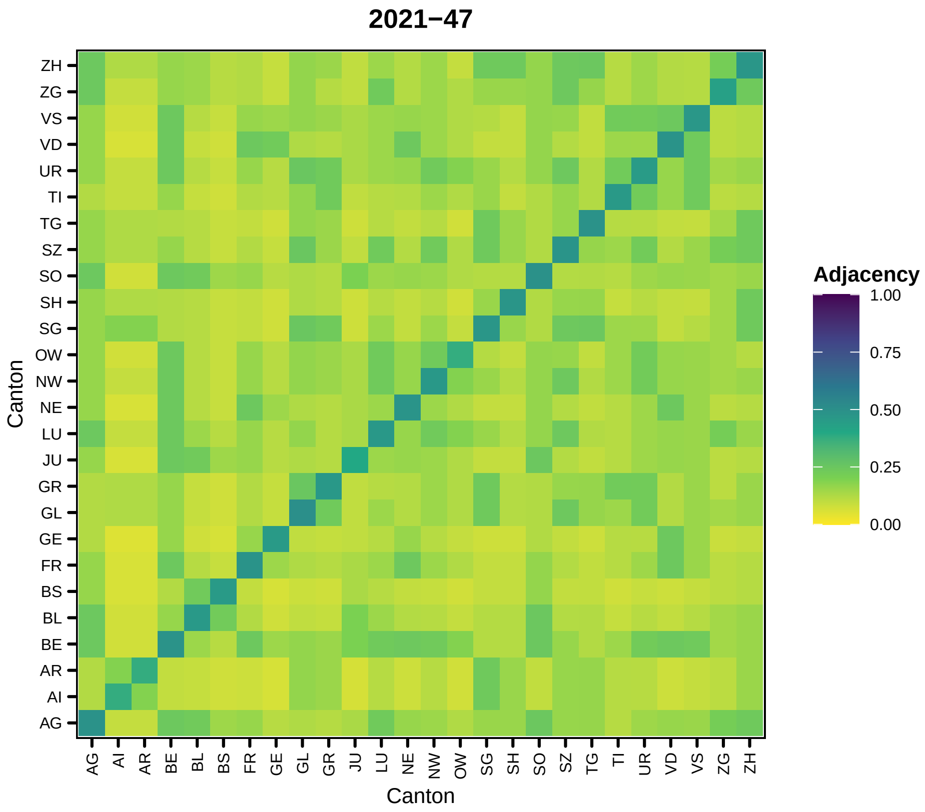

2021−47
Adjacency
1.00
0.75
0.50
0.25
0.00
Canton
Canton
ZH
ZG
VS
VD
UR
TI
TG
SZ
SO
SH
SG
OW
NW
NE
LU
JU
GR
GL
GE
FR
BS
BL
BE
AR
AI
AG


120

```
##
## [[49]]
```

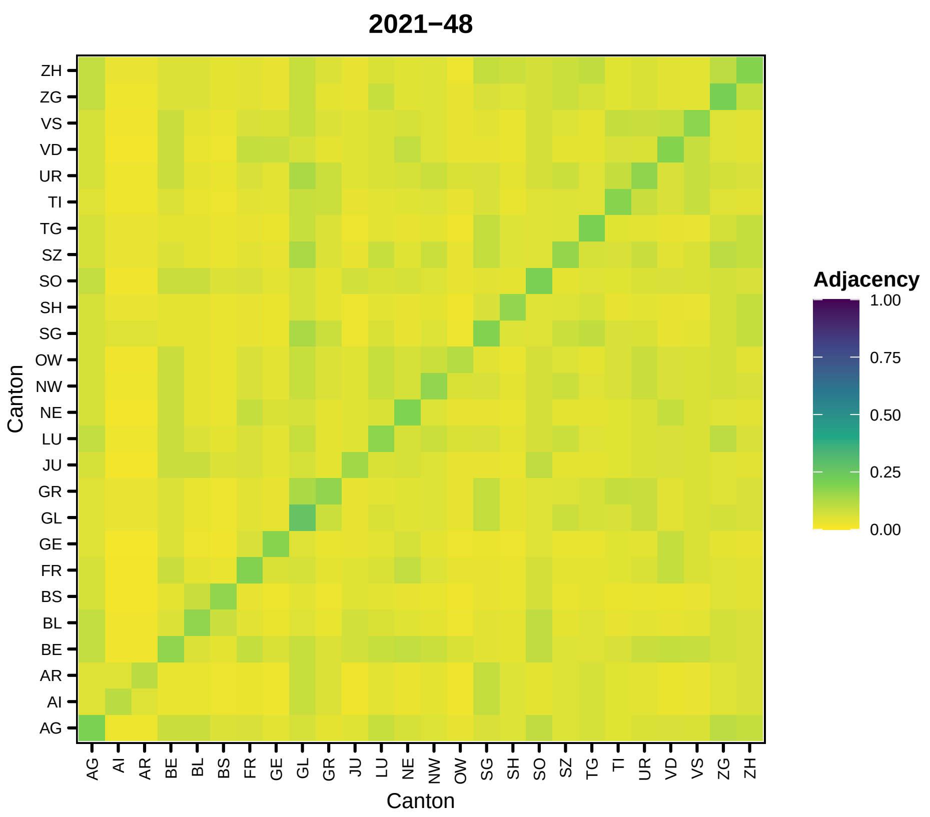
2021−48
Adjacency
1.00
0.75
0.50
0.25
0.00
Canton
Canton
ZH
ZG
VS
VD
UR
TI
TG
SZ
SO
SH
SG
OW
NW
NE
LU
JU
GR
GL
GE
FR
BS
BL
BE
AR
AI
AG

121

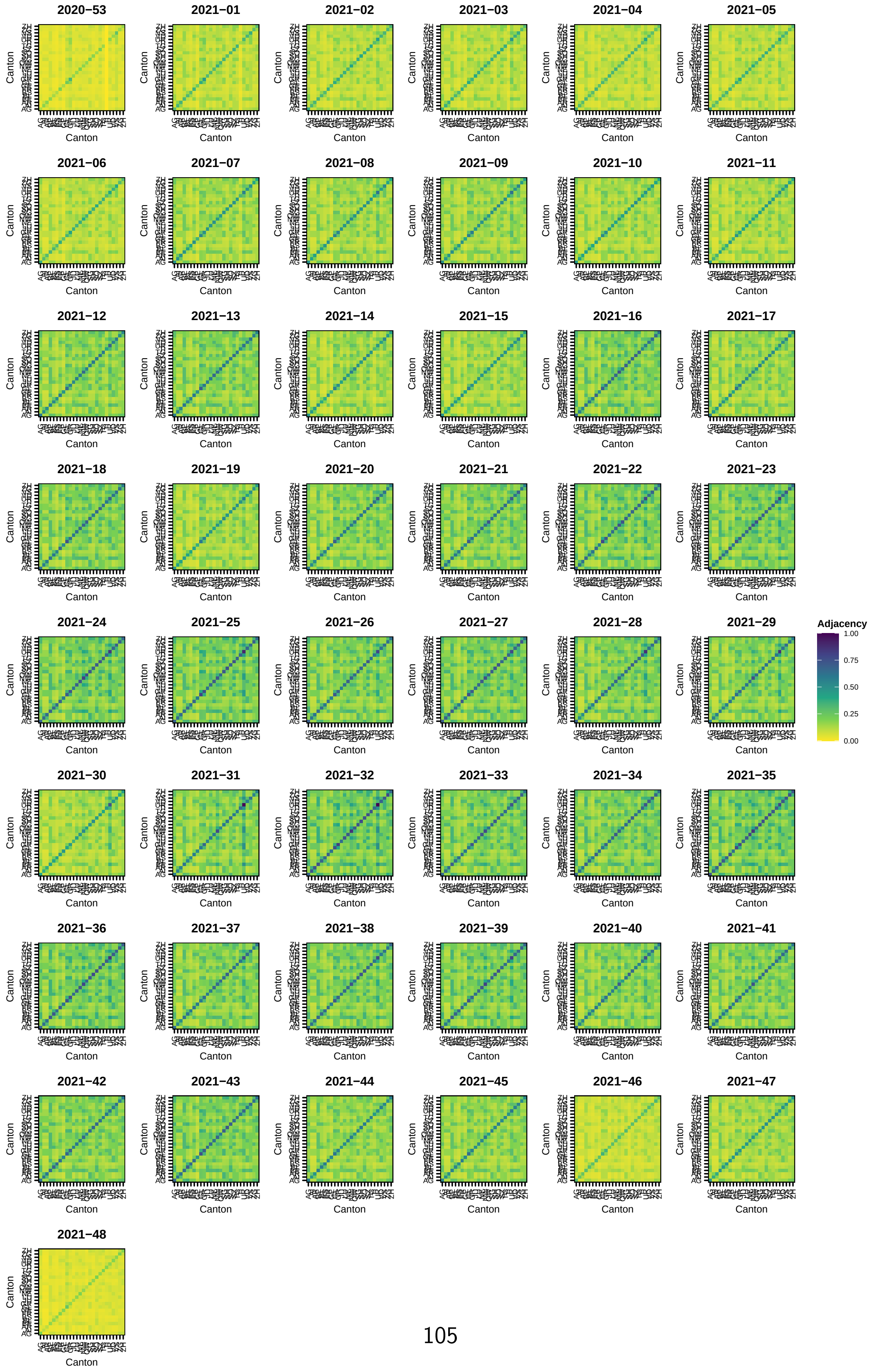
2020−53
2021−01
2021−02
2021−03
2021−04
2021−05
2021−06
2021−07
2021−08
2021−09
2021−10
2021−11
2021−12
2021−13
2021−14
2021−15
2021−16
2021−17
2021−18
2021−19
2021−20
2021−21
2021−22
2021−23
2021−24
2021−25
2021−26
2021−27
2021−28
2021−29
2021−30
2021−31
2021−32
2021−33
2021−34
2021−35
2021−36
2021−37
2021−38
2021−39
2021−40
2021−41
2021−42
2021−43
2021−44
2021−45
2021−46
2021−47
2021−48
Canton
Adjacency
1.00
0.75
0.50
0.25
0.00

#### 5 Vaccination coverage covariate construction

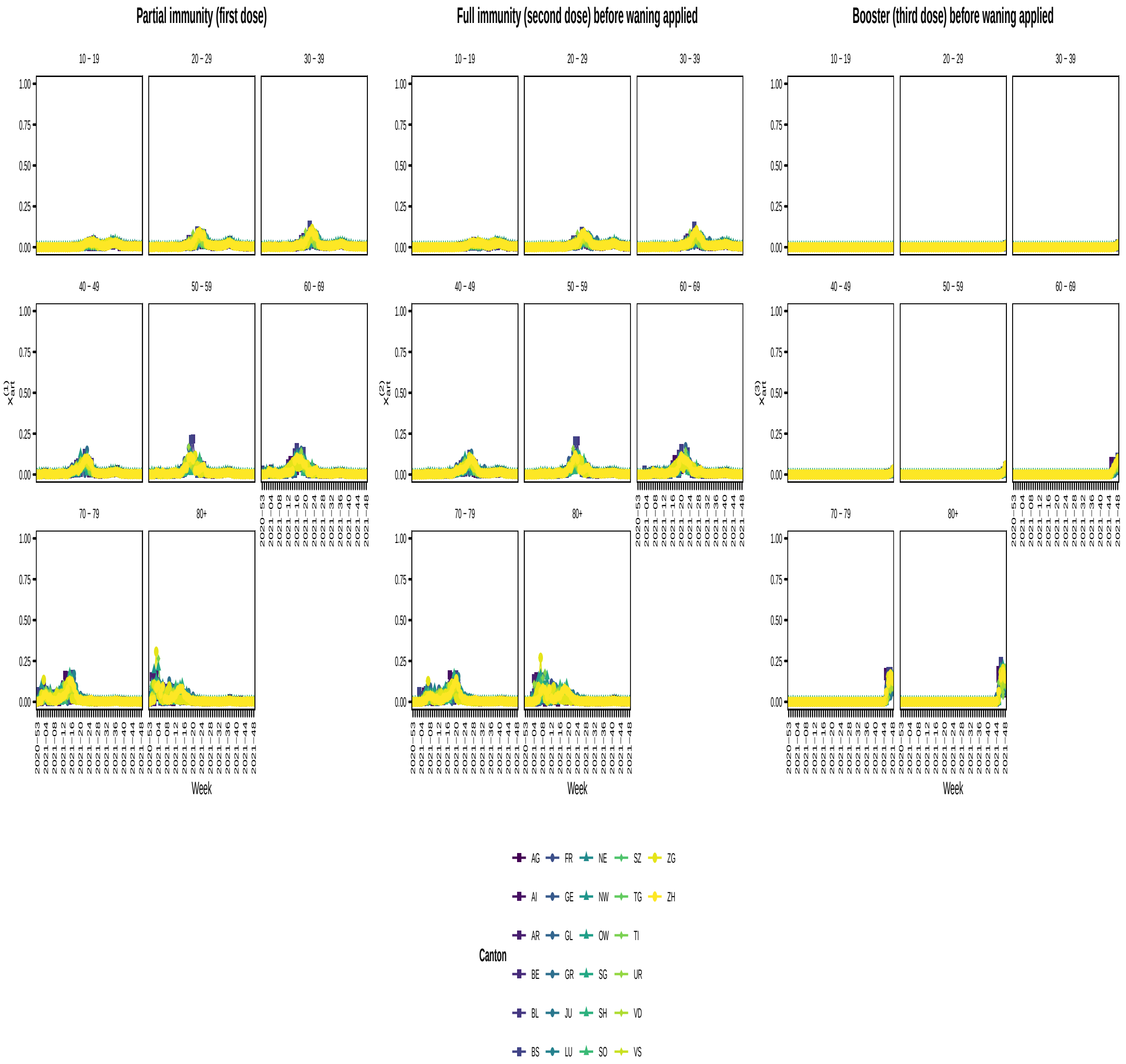


We correct the vaccination coverage for waning as described in Bekker-Nielsen Dunbar and Held [1]. The waning function is multiplied to each time point. This is achieved using a triangle matrix. For each age and canton combination at each day we sum up the entries for that day as well as the waned entries for previous days. We then divide by the population.

### 6 Cases

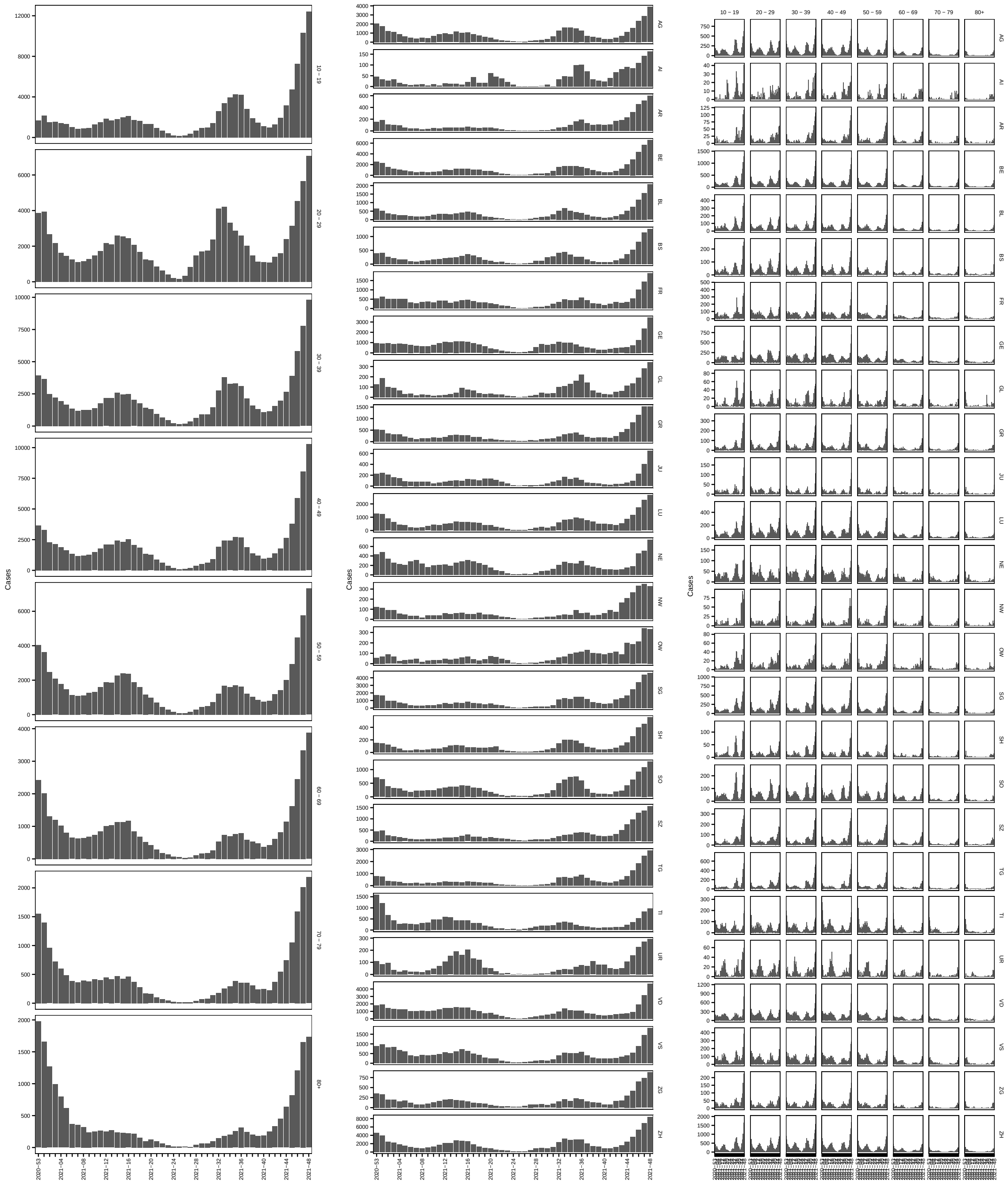


### 7 Measles literature

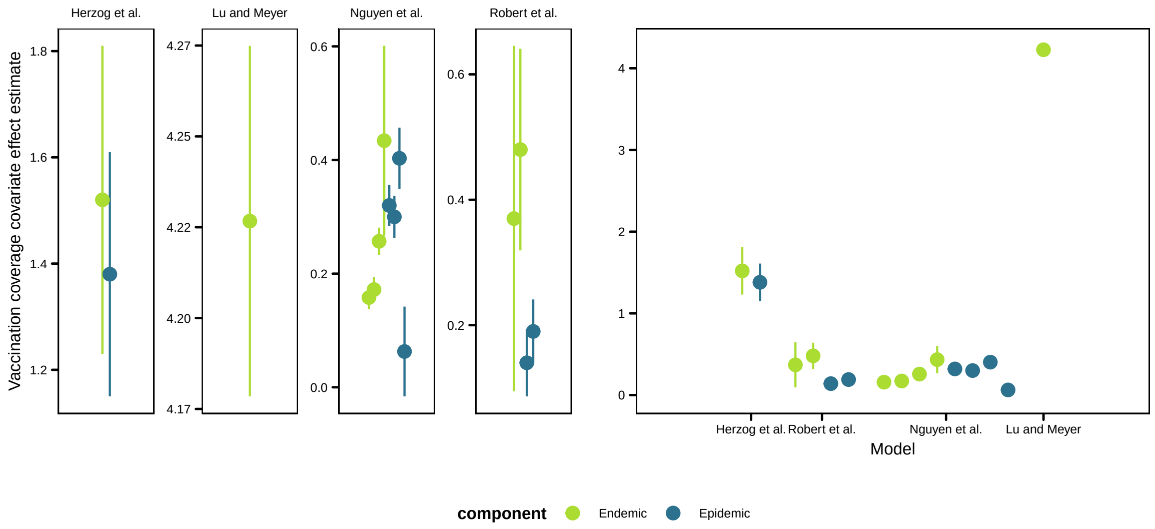


### 8 Predicted cases

Distribution of predicted cases for the model with vaccination coverage in the endemic component under the original and alternative vaccination coverage option for regions (above) and age groups (below)

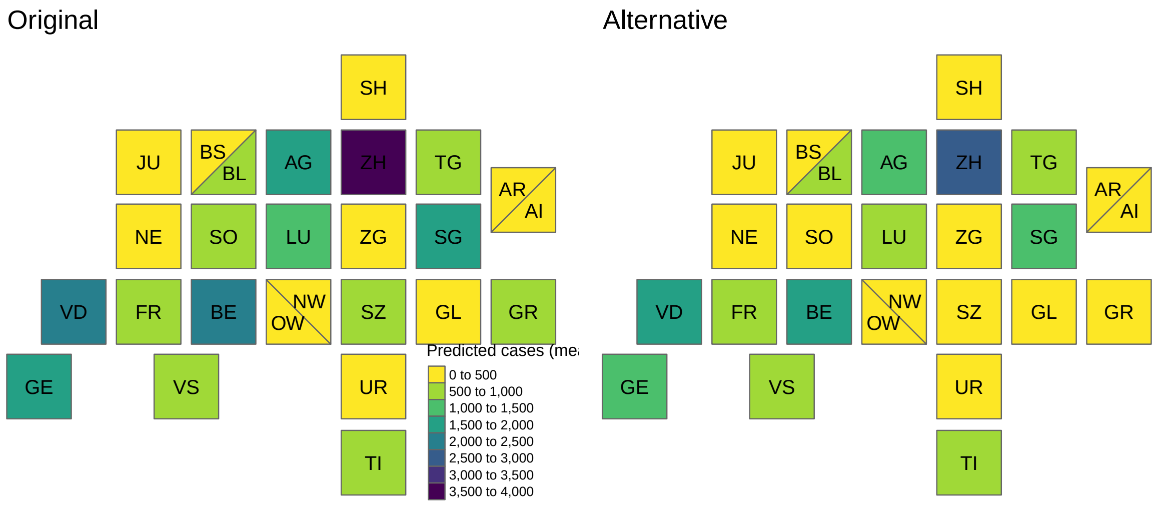

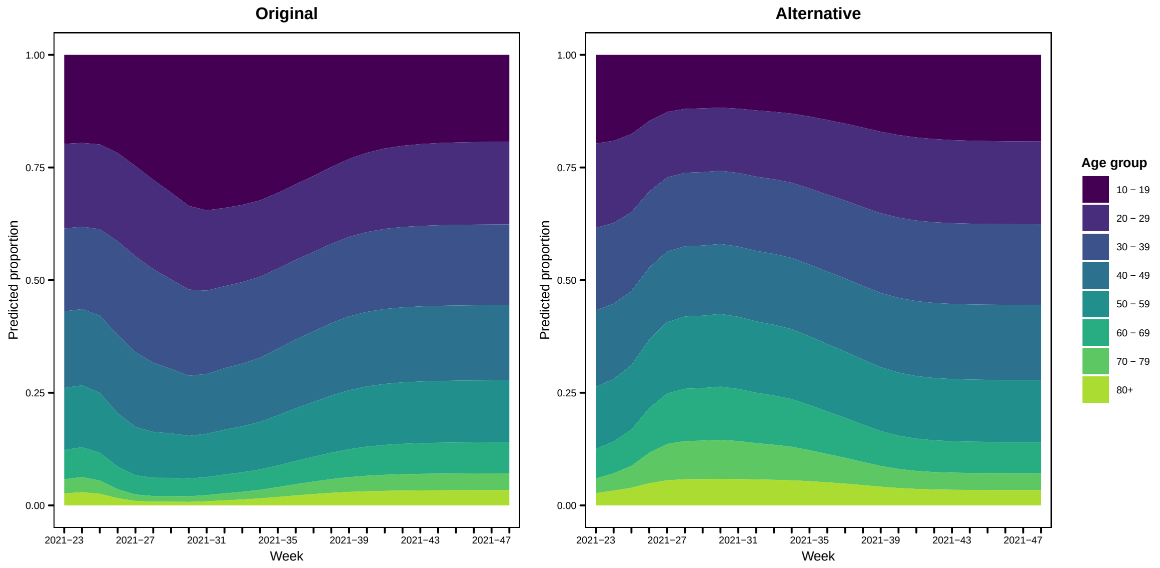


139

140 ## 8.1 Summed across prediction window

141 Final size estimates based on path forecasts (regional model)

| | | Original coverage | | | Alternative coverage | | |
|---|---|---|---|---|---|---|---|
| | True | Endemic | Epidemic | Both | Endemic | Epidemic | Both |
| AG | 24208 | 42919 | 23073 | 22502 | 31254 | 8402 | 6924 |
| AI | 1288 | 1925 | 1339 | 1292 | 1385 | 192 | 154 |
| AR | 3687 | 4795 | 3086 | 2988 | 3481 | 537 | 432 |
| BE | 37976 | 58133 | 28781 | 28026 | 42970 | 11296 | 9738 |
| BL | 10983 | 16992 | 9262 | 8999 | 12751 | 3810 | 3410 |
| BS | 7531 | 11523 | 5482 | 5341 | 8757 | 3007 | 2724 |
| FR | 10182 | 21343 | 9765 | 9495 | 15612 | 3234 | 2659 |
| GE | 19107 | 41614 | 21638 | 21308 | 30515 | 9968 | 8516 |
| GL | 2413 | 3541 | 2433 | 2423 | 2407 | 790 | 589 |
| GR | 9279 | 13867 | 7458 | 7290 | 10029 | 2480 | 2012 |
| JU | 2586 | 5798 | 3493 | 3410 | 4168 | 879 | 695 |
| LU | 18050 | 30844 | 16897 | 16602 | 22215 | 5761 | 4676 |
| NE | 4708 | 11888 | 5779 | 5611 | 8727 | 2022 | 1693 |
| NW | 2377 | 3684 | 2027 | 1969 | 2681 | 469 | 381 |
| OW | 2571 | 4049 | 2491 | 2430 | 2898 | 486 | 376 |
| SG | 30828 | 43552 | 27724 | 27038 | 31215 | 7094 | 5608 |
| SH | 3411 | 5272 | 2966 | 2884 | 3897 | 1087 | 938 |
| SO | 9357 | 16315 | 8919 | 8702 | 11941 | 3390 | 2856 |
| SZ | 10431 | 15106 | 10235 | 10068 | 10690 | 2467 | 1902 |
| TG | 16314 | 20355 | 13565 | 13226 | 14548 | 3399 | 2709 |
| TI | 6105 | 16835 | 8746 | 8485 | 12644 | 3754 | 3341 |
| UR | 1860 | 3762 | 2497 | 2410 | 2747 | 404 | 333 |
| VD | 23841 | 56845 | 25277 | 24551 | 42101 | 10648 | 9175 |
| VS | 10502 | 24863 | 13690 | 13375 | 18077 | 4157 | 3441 |
| ZG | 5337 | 9020 | 4908 | 4759 | 6702 | 2055 | 1775 |
| ZH | 55763 | 97121 | 44679 | 42971 | 73768 | 23259 | 20912 |

#### Final size estimates based on path forecasts (age group model)

| | | Original coverage | | | Alternative coverage | | |
|---|---|---|---|---|---|---|---|
| | True | Endemic | Epidemic | Both | Endemic | Epidemic | Both |
| 10 - 19 | 73195 | 115341 | 497769 | 151064 | 54394 | 54878 | 44820 |
| 20 - 29 | 59099 | 94228 | 282376 | 105667 | 54644 | 74290 | 49647 |
| 30 - 39 | 62337 | 93760 | 265990 | 106912 | 54640 | 63686 | 49436 |
| 40 - 49 | 55949 | 84499 | 226076 | 95705 | 51097 | 57481 | 46829 |
| 50 - 59 | 40373 | 68097 | 171177 | 75906 | 44217 | 49229 | 41602 |
| 60 - 69 | 20807 | 33111 | 78320 | 36738 | 24494 | 26597 | 24109 |
| 70 - 79 | 11779 | 17250 | 49350 | 20853 | 14677 | 15245 | 15032 |
| 80+ | 9342 | 15400 | 59054 | 21075 | 12319 | 13778 | 12827 |
