## Supporting information (sens) for "The COVID-19 vaccination campaign in Switzerland and its impact on disease spread"

### Sensitivity analyses

M. Bekker-Nielsen Dunbar and L. Held
Epidemiology, Biostatistics and Prevention Institute, University of Zurich

---

For full information on the work this document supports please see the manuscript and the study protocol [1]. This supporting document contains sensitivity analyses and knowledge of the analyses from the main manuscript is assumed.

#### 1 Ruminations on vaccination coverage

The vaccination coverage takes values between 0 and 1 by definition and construction.

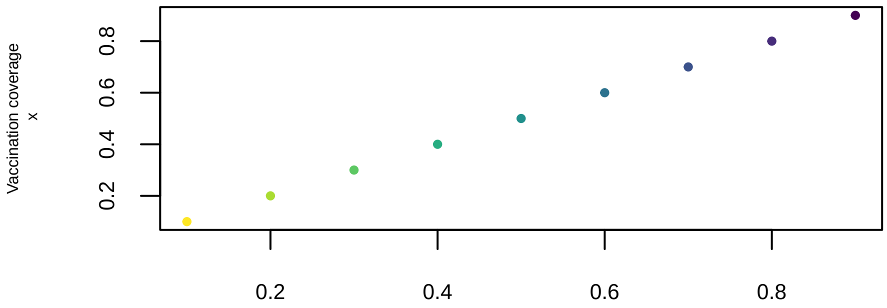


To obtain the proportion of the population that is unvaccinated we transform the vaccination coverage by $f(x) = 1 - x$, meaning this also takes values between 0 and 1.

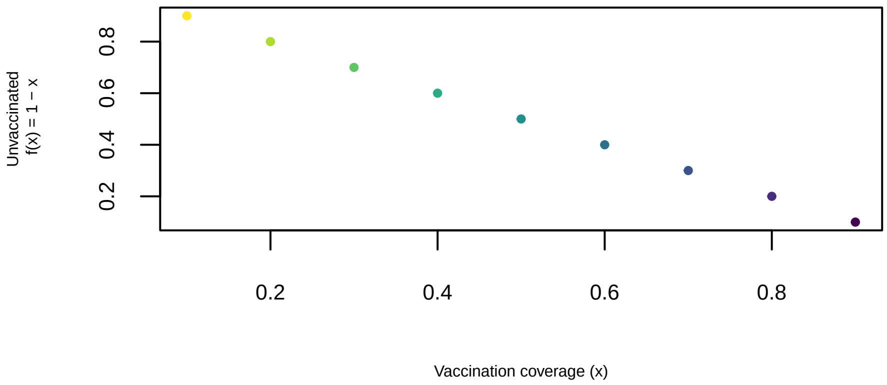


Then log-transforming the unvaccinated via $g(f(x)) = \log(1-x)$, we obtain negative values and the trend goes in the opposite direction.

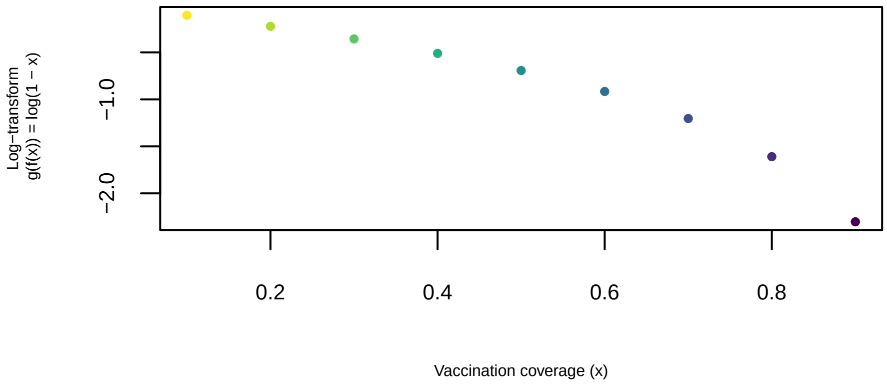


We see that the higher the vaccination coverage is, the more negative the covariate value will be. With a positive effect estimate, a higher proportion of unvaccinated will lead to an increase in cases–as would be expected. All other things equal, an increase in parameters from including the vaccination coverage would lead to an increase in our model comparison measure. However, our exposition and intuition (as outlined in the study protocol) leads us to believe that the model without vaccination coverage ("neither") should not have the lowest value though it has the lowest number of parameters as we believe it to be the most important model effect.

#### 2 Sensitivity analysis of waning functions

We consider the alternative representations of waning outlined in the study protocol (Figure 1). The results we obtain look similar.

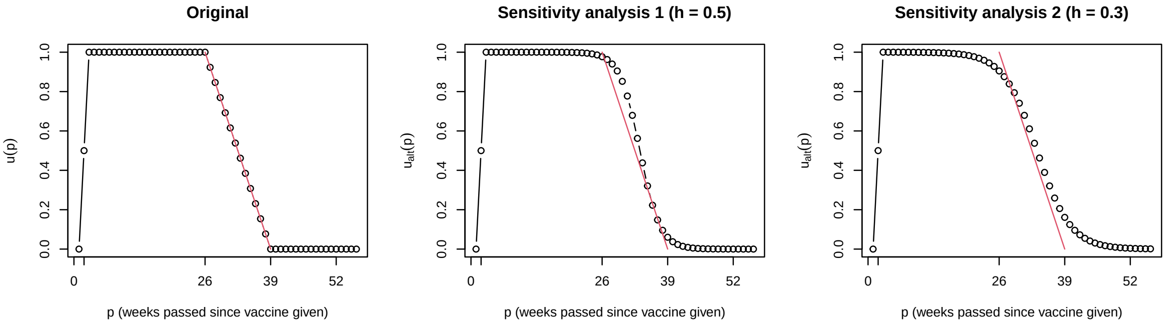


**Figure 1:** Alternative waning functions that could have been used in the construction of the vaccination coverage covariate

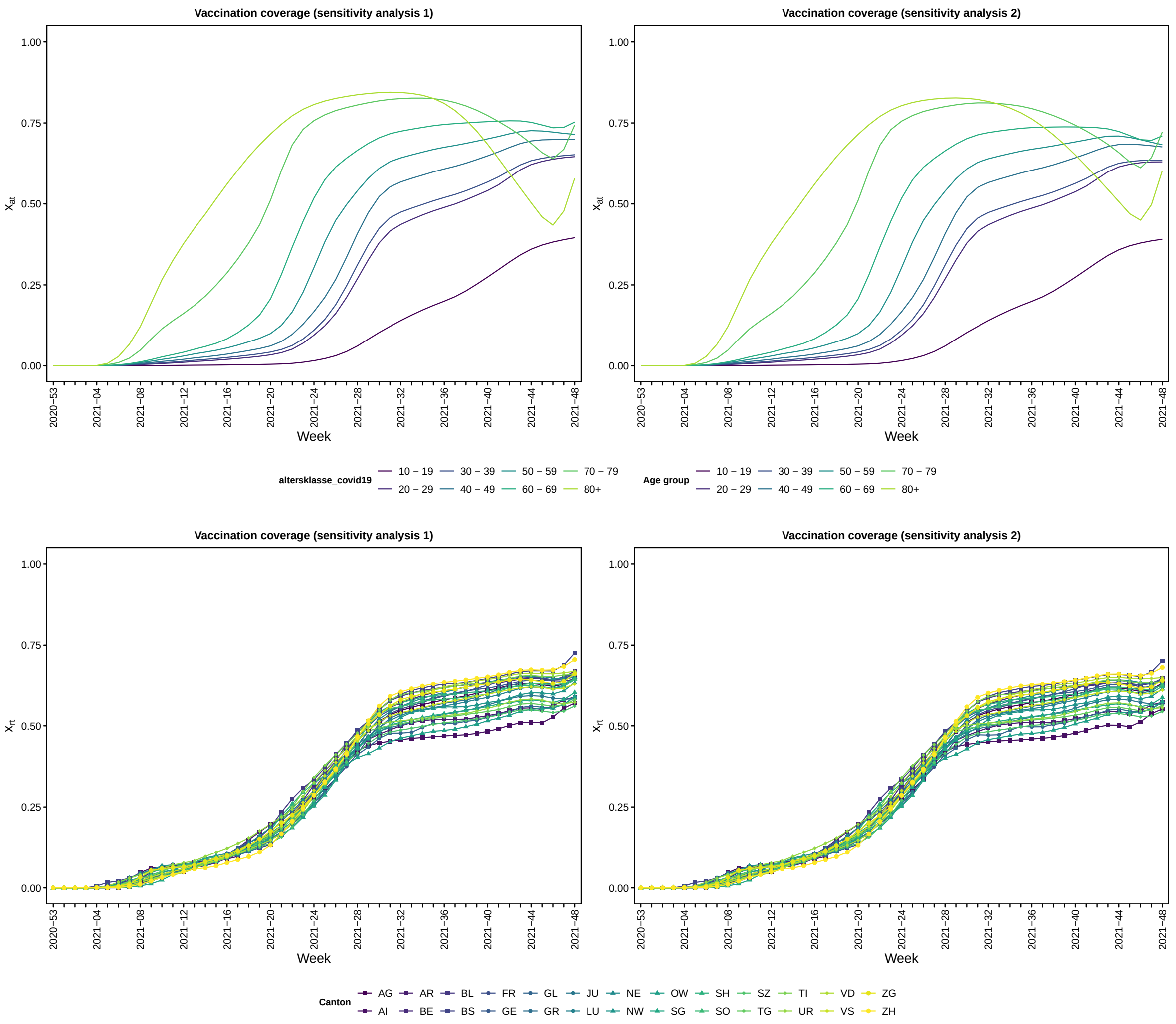


**Figure 2:** Log-transformed vaccination coverage for age groups (above) and regions (below)

**Table 1:** Vaccination coverage covariate estimates $\hat{\beta}$ and standard errors (SE) for alternative waning function 1

| | Weight option | Neither | Endemic | Epidemic | Both | |
|---|---|---|---|---|---|---|
| | | | | | Endemic | Epidemic |
| Region | $w_{r,r'}$ (1R) | – | 4.277 (SE 1.344) | 3.315 (SE 0.212) | 1.538 (SE 1.018) | 3.273 (SE 0.212) |
| | $w_{r,r',t}$ (2R) | – | 3.241 (SE 1.034) | 2.588 (SE 0.198) | 1.698 (SE 0.843) | 2.544 (SE 0.197) |
| Age | $w_{a,a'}$ (1A) | – | 2.042 (SE 0.246) | 0.725 (SE 0.062) | 2.126 (SE 0.143) | 0.42 (SE 0.055) |
| | $w_{a,a',t}$ (2A) | – | 2.227 (SE 0.121) | 0.571 (SE 0.07) | 2.041 (SE 0.117) | 0.226 (SE 0.059) |

**Table 2:** Vaccination coverage covariate estimates $\hat{\beta}$ and standard errors (SE) for alternative waning function 2

| | Weight option | Neither | Endemic | Epidemic | Both | |
|---|---|---|---|---|---|---|
| | | | | | Endemic | Epidemic |
| Region | $w_{r,r'}$ (1R) | – | 4.278 (SE 1.35) | 3.425 (SE 0.223) | 1.521 (SE 1.035) | 3.381 (SE 0.223) |
| | $w_{r,r',t}$ (2R) | – | 3.253 (SE 1.039) | 2.801 (SE 0.208) | 1.654 (SE 0.847) | 2.755 (SE 0.207) |
| Age | $w_{a,a'}$ (1A) | – | 2.044 (SE 0.246) | 0.775 (SE 0.064) | 2.179 (SE 0.134) | 0.475 (SE 0.057) |
| | $w_{a,a',t}$ (2A) | – | 2.307 (SE 0.121) | 0.613 (SE 0.073) | 2.111 (SE 0.115) | 0.268 (SE 0.061) |

We find that the alternative representations of waning outlined in the study protocol (Figure 1) yield similar vaccine coverage effect estimates (Tables 1 and 2) as the main analysis.

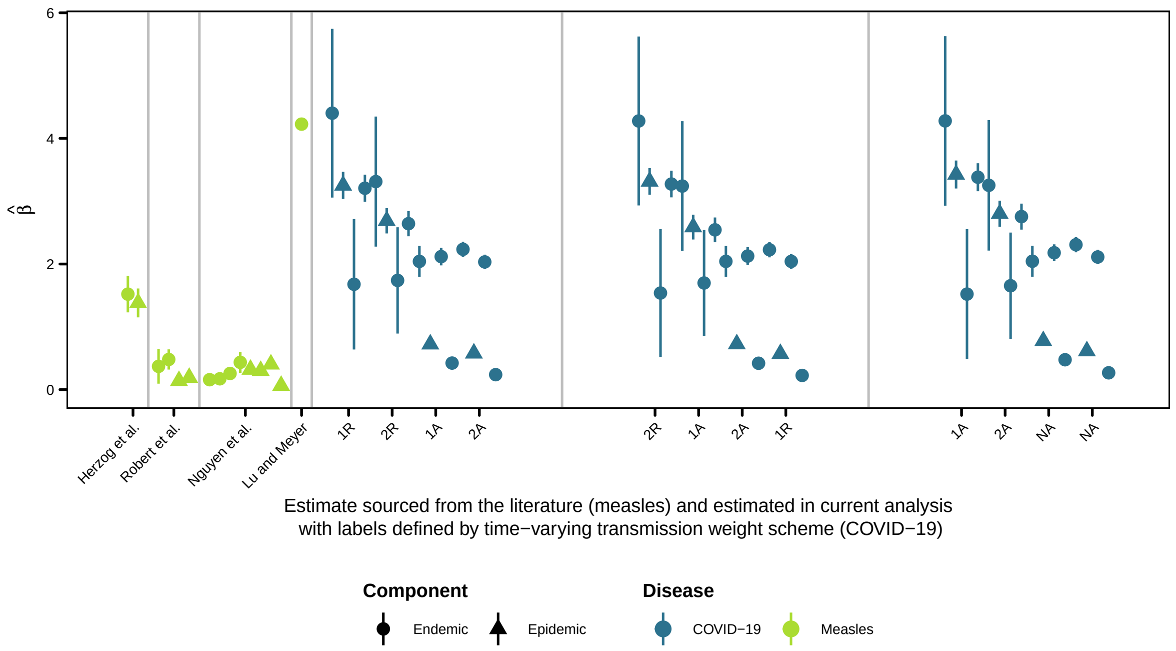


#### 3 Calculation of alternative age-based vaccination coverage

We showcase the result of averaging the vaccination coverage across age groups as outlined in the study protocol to compare with the manual redistribution:

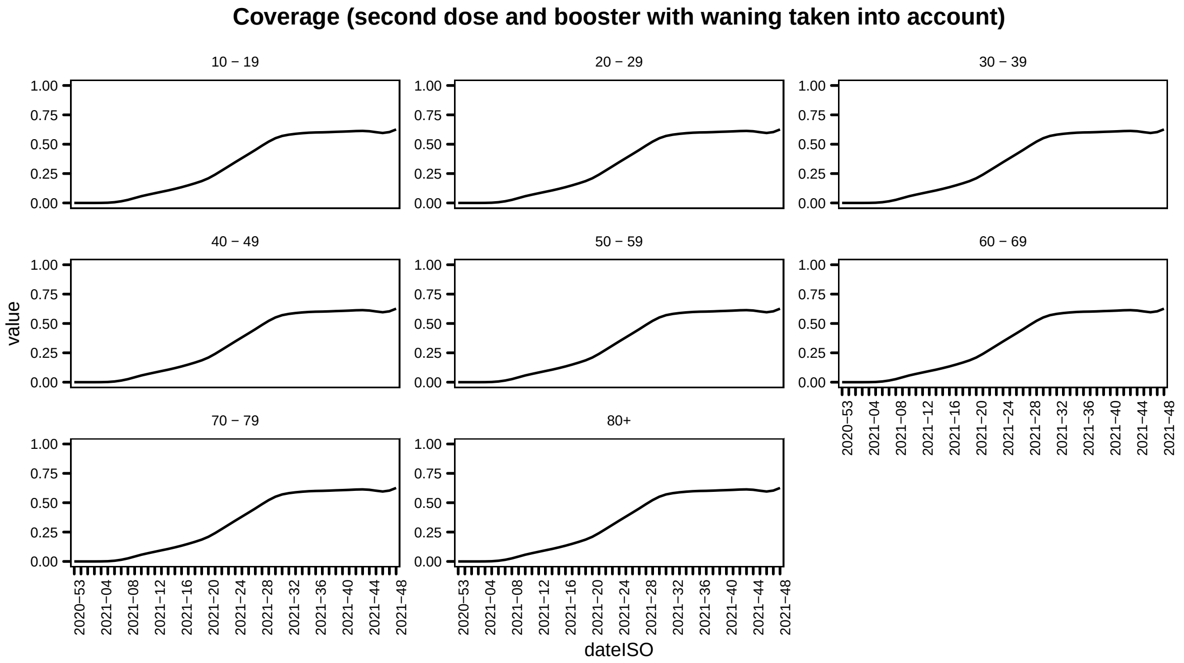

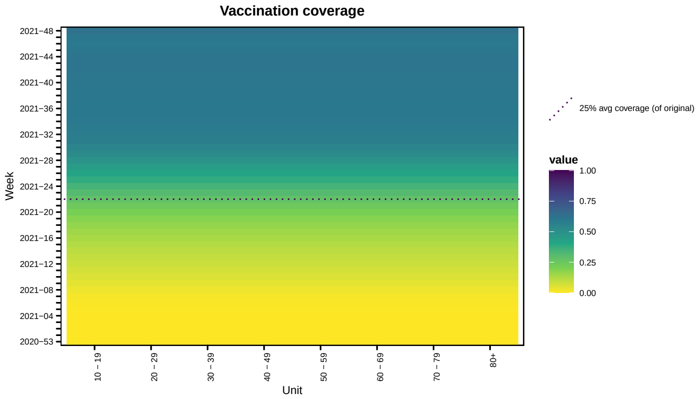


This looks the same as the redistributed one.
